# The Role Of Drug Indication On Incidence Rate Heterogeneity: A Large-Scale, Systematic Evaluation Across An International Network Of Observational Databases

**DOI:** 10.1101/2025.10.22.25338563

**Authors:** Hsin Yi Chen, Christopher Knoll, Elise Boventer, Nicole Pratt, Tara V. Anand, Mui Van Zandt, Hannah Morgan-Cooper, Patrick B. Ryan, George Hripcsak

## Abstract

**Purpose:** Incidence rate estimates are sensitive to a range of factors, including age, sex, and geographical setting (data source). The magnitude of the impact of drug indication on incidence rates remains underexplored.

**Methods:** We conducted an observational cohort study using 13 healthcare databases to estimate the incidence rates of 73 health outcomes across 8 drug classes with multiple indications. We calculated incidence rates for each drug-outcome pair and performed random-effects meta-analyses to pool results across databases. Then, we conducted variance components analysis to find the proportions of variability attributed to database, age, sex, and indication. We reported the median of the variance components across all 73 health outcomes as a measure of the magnitude of differences across indications, age, sex, and database, per drug class.

**Results:** Adjusting for database, age, and sex differences, the drug classes with the highest median VC were trimethoprim (0.49), SGLT-2 inhibitors (0.26), and beta blockers (0.10), while the drug class with the lowest VC was GLP-1 agonists (<0.01). Within each drug class, and adjusting for all other factors, age was frequently the strongest contributor to incidence variation (for 5/8 drug classes, the highest class-wide median VC was the age median VC), followed by database, indication, then biological sex.

**Conclusion:** This study showed that for some drug classes, there exists substantial variation in incidence rates estimates across indications even after accounting for heterogeneity due to age, biological sex, and data source. As many drugs have multiple indications in clinical practice, it may be important to consider drug indication when estimating incidence rates in observational studies for the purpose of patient safety evaluations.

## Introduction

Incidence rate calculation is commonly used in pharmacoepidemiology for a variety of purposes, such as comparative background rates for drug adverse events, assessing the potential public health impact of adverse events, and clinical trial design^1,2^. However, the accuracy of incidence rates reported on drug package inserts and published in the literature is often uncertain. This is because the evidence used to derive the estimates are typically from pre-marketing clinical trials which may not always provide comprehensive data for all adverse events, or for diverse real-world populations and different patient subgroups. Furthermore, drug package inserts rarely provide incidence rates stratified by indication, even for drugs approved to treat multiple conditions. Similarly, published incidence rates found in the broader medical literature, sourced from various study types, including clinical trials, observational cohort studies, and systematic reviews, can exhibit significant heterogeneity. Consequently, the applicability of these incidence rates to a specific patient or a distinct patient population is often complicated by variability in the study populations and methodologies from which the rates were originally derived.

While it may be a common practice to collapse all available incidence rates in the literature into a single estimate (for example, through meta-analysis), this approach is only appropriate when the source studies are sufficiently homogeneous with regard to critical factors such as age distribution, sex, and underlying disease state, particularly when these factors are known to affect the incidence of the outcome under study^3–5^. Other factors that further complicate and increase the uncertainty in meta-analyses include different operational definitions of incidence rates^6^, differing definitions of the adverse event^7^, and the diversity of study designs.

Previously published research that systematically studied the influence of patient demographics on incidence rates^8^ found that they were highly influenced by age and choice of database. However, the paper did not consider the impact of drug indication as a source of heterogeneity when interpreting incidence rates. When a single drug can be used to treat different conditions, the incidence rates of adverse events may depend on the specific condition the medicine is used for, as each indication often represents a distinct target population with different baseline risks. In addition, patterns of drug exposure, such as treatment dosage or frequency, can also differ by indication and have an impact on the potential for adverse events to develop. The relationship between indication and outcome is sometimes direct: for instance, patients prescribed medications for cardiovascular conditions inherently have a higher baseline risk of cardiovascular events (a phenomenon often termed confounding by indication^9–11^). In such cases, stratification by indication is clearly necessary. However, when the drug-indication-outcome relationship is less direct (e.g., beta blockers can be used for both hypertension and acute myocardial infarction, and neither indication have a direct relationship with a health outcome like weight gain), the extent to which indication contributes to incidence rate heterogeneity, beyond factors like age and sex, remains unclear.

This study seeks to quantify the magnitude of heterogeneity in adverse event incidence rates for 73 different health outcomes attributable to drug indication, after accounting for commonly known sources of variability such as patient demographics and database-specific factors. To achieve this, we used real-world data from a large network of observational databases to calculate the incidence rates of the 73 health outcomes across 8 drug classes, stratified by indication. Our primary research question is two-fold: (1) How much does drug indication influence incidence rate estimates for different health outcomes after accounting for variations due to demographics (age and biological sex) and data source? (2) How does the magnitude of this influence compare to the variability attributable to these other factors (age, biological sex, and data source)?

## Methods

### Data Sources

We conducted an observational cohort study using routinely collected health care data from 13 databases mapped to the Observational Medical Outcomes Partnership (OMOP) Common Data Model (CDM)^12^ in October 2023. The data sources varied in provenance (administrative claims, electronic health records (EHR)), origin (e.g., US, Belgium, Italy, Australia, France), and patient populations (e.g., commercially insured patients in Merative Commercial Claims (CCAE), versus patients with limited income in Merative Multi-state Medicaid (MDCD)). Database source descriptions are included in the Supplement (Table S1).

### Target Cohorts

We selected target cohorts of patients observed in the database at any time between January 1, 2012, and December 31, 2022, with at least 365 days of prior observation. Patients were selected based on the first occurrence of an exposure to a drug class in their longitudinal record.

We selected 8 drug classes of interest that are commonly prescribed and known to have multiple indications: 1) beta blockers, 2) cephalosporins, 3) fluoroquinolones, 4) GLP-1 agonists, 5) JAK inhibitors, 6) SGLT2 inhibitors, 7) trimethoprim, and 8) TNF-alpha inhibitors. These drug classes were defined using RxNorm^13^ terms for marketed drug ingredients, and the included ingredients for each of these drug classes is described further in the Supplement (Table S2). We further defined subgroups for each drug class based on the indication(s) for which the drug was prescribed, described in Table 1.

**Table 1.** Overview of the 8 drug classes and their indications, and the median variance components (VC)) of the log incidence rates across 73 outcomes for indications, age, sex, and biological sex, by drug class.

| Drug Class | Indications | Median VC (IQR) attributed to Indications | Median VC (IQR) attributed to Age | Median VC (IQR) attributed to Biological sex | Median VC (IQR) attributed to Database |
| --- | --- | --- | --- | --- | --- |
| **Beta Blocker** | 1) Essential Hypertension, 2) Left Heart Failure, 3) Acute Myocardial Infarction | 0.1013 (0.1628) | 0.3102 (0.5385) | 0.0204 (0.0487) | 0.1537 (0.1288) |
| **SGLT-2 Inhibitor** | 1) Type 2 Diabetes Mellitus, 2) Left Heart Failure | 0.2642 (0.4585) | 0.2779 (0.4016) | 0.0155 (0.0828) | 0.3170 (0.6178) |
| **GLP-1 Agonist** | 1) Type 2 Diabetes Mellitus, 2) Obesity | <0.001<br>(0.0017) | 0.3678<br>(0.7298) | 0.0289<br>(0.0831) | 0.6117<br>(1.1611) |
| **Trimethoprim** | 1) Urinary tract infection, 2) Acute Typical Pneumonia | 0.4887<br>(0.6656) | 1.5228<br>(2.6802) | 0.1219<br>(0.1614) | 0.2772<br>(0.7984) |
| **Cephalosporin** | 1) Urinary Tract Infection, 2) Acute Typical Pneumonia | 0.0397<br>(0.0942) | 1.4631<br>(2.5357) | 0.0515<br>(0.0857) | 0.7230<br>(1.1065) |
| **Fluoroquinolone** | 1) Urinary Tract Infection, 2) Acute Typical Pneumonia | 0.0983<br>(0.1766) | 0.8573<br>(1.2045) | 0.0696<br>(0.1039) | 0.9940<br>(1.6399) |
| **JAK Inhibitor** | 1) Rheumatoid Arthritis, 2) Ulcerative Colitis | 0.0383*<br>(0.1681) | 0.2055*<br>(0.6144) | 0.0937*<br>(0.2735) | 0.1792*<br>(0.5046) |
| **TNFα inhibitor** | 1) Rheumatoid Arthritis, 2) Ulcerative Colitis, 3) Plaque Psoriasis, 4) Psoriatic Arthritis, 5) Crohn's Disease | 0.0332<br>(0.0662) | 0.1815<br>(1.0491) | 0.0221<br>(0.0673) | 0.1675<br>(0.3101) |

\* For JAK inhibitors, the VC attributed to indications was only adjusted by databases (and not age or sex) because of low counts in the age and sex subgroups in some databases when we simultaneously adjust for both age and sex, or sex and database. The JAK inhibitor $VC_{age}$ , $VC_{sex}$ , and $VC_{DB}$ was only adjusted by indications for the same reason.

### Health Outcomes of Interest

The study examined 73 different health outcomes, chosen due to their relevance in public health surveillance and drug/vaccine safety. This list was curated based on (1) the protocol published by the FDA center for Biologics Evaluation and Research^14^, (2) Designated Medical Events^15^, and (3) outcomes previously published in the Observational Health Data Sciences and Informatics (OHDSI) studies such as the LEGEND-Hypertension and Type 2 Diabetes studies^16,17^. The full list of outcomes is available in the Supplement (Table S3).

All outcome cohorts were constructed using previously validated phenotype algorithms, typically involving one or more diagnosis codes recorded in inpatient or outpatient settings. For each outcome, patients with a history of the event prior to drug initiation were excluded. Patient time-at-risk was defined using an intention-to-treat approach, following patients for 365 days after the index drug exposure date. Reproducible cohort definitions can be found in the OHDSI Phenotype Library^18^.

### Statistical Analysis

We calculated incidence rates (IR) for each combination of target drug class, indication, and outcome (Equation 1). To ensure stable IR estimates, estimates were excluded if the number of persons at risk for any single IR calculation was less than 1000.

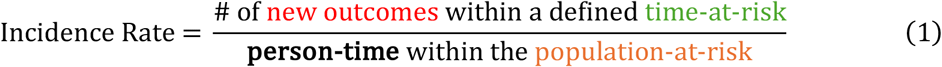

For all calculated IRs, we conducted random-effects meta-analyses using the R package metafor (version 4.6)^19^ to generate pooled log incidence rates and 95% prediction intervals across data sources for each drug-indication-outcome triad. We conducted variance components analysis (VCA)^20^ to quantify the magnitude of log incidence rate heterogeneity attributable to each factor: age (categorized into deciles), biological sex, database, and indication.

VCA partitions the total variance for each log IR into components associated with different sources (e.g., database, age, biological sex, and drug indication). We estimated these variance components (VC) for each drug class-outcome pair (resulting in 73 sets of variance component estimates for each drug class). We reported the median VC for each factor (indication, age, sex, database) across the 73 outcomes for each drug class (Figure 1), while adjusting for all other factors. A higher variance component for a given factor indicates a greater contribution of that factor to the overall heterogeneity. For example, a high variance component due to indication, after adjusting for age, sex, and database, would suggest that indication has a large contribution to the observed variation in IRs.

**Figure 1.**
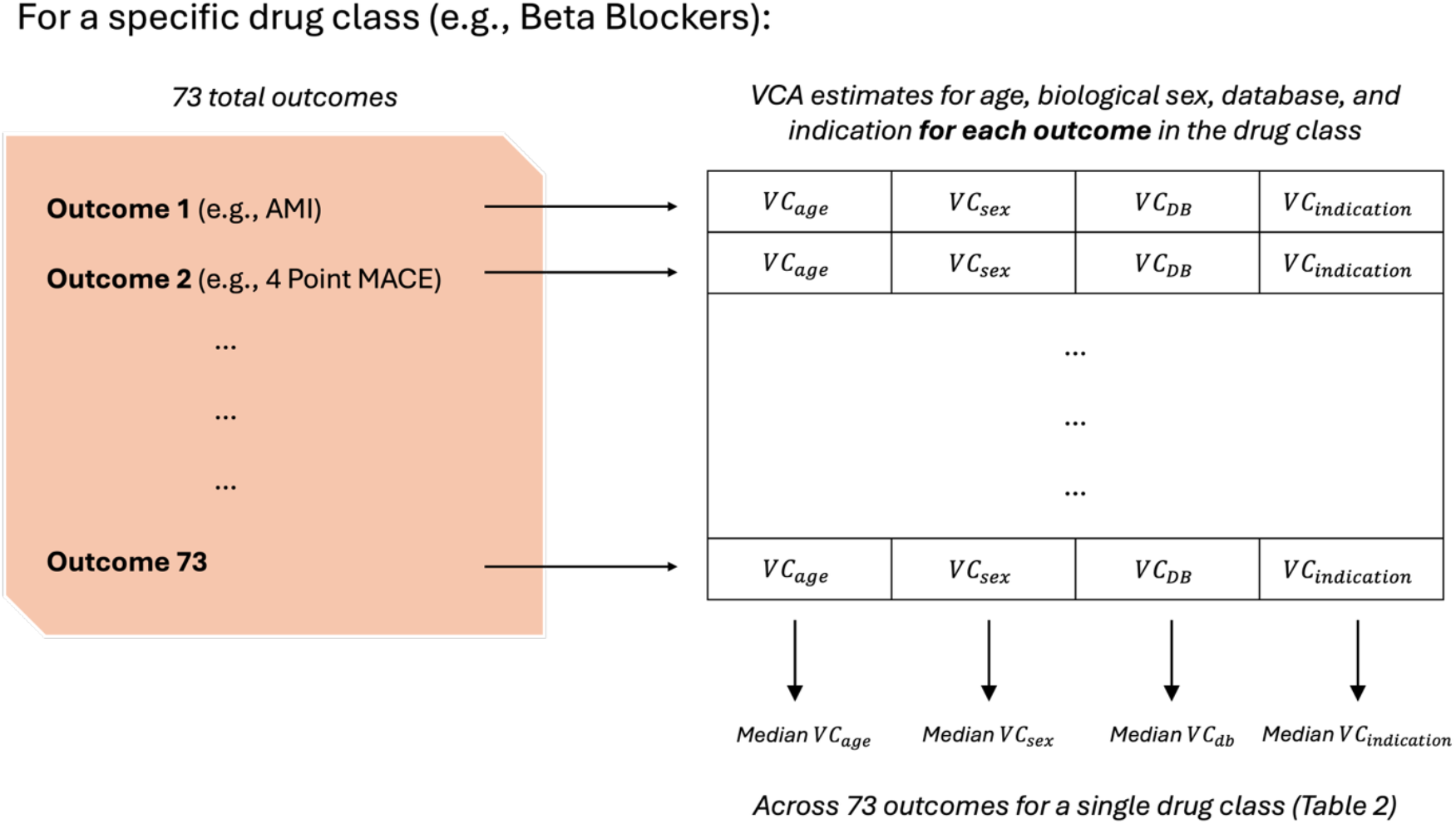
Analysis procedure for VCA for *each drug class* so that we get median variance components (across all outcomes) *per drug class*.

### Data Availability Statement

Detailed descriptions of the analysis protocol and results can be found on Github (https://github.com/ohdsi-studies/HowOften) and https://results.ohdsi.org.

### Results

We calculated a total of 77,631 incidence rates across all target drug-indication-outcome-database combinations.

The VC estimates attributed to indications (*VC*_*ind*_), after adjusting for age, sex, and database, were calculated for *each* drug-outcome pair for each class (resulting in 73 VC estimates per drug class, because there are 73 different outcomes). The class-wide median *VC*_*ind*_ and interquartile ranges are reported in Table 1. It was the highest for trimethoprim which had 2 indications, Urinary tract infection and Acute Typical Pneumonia, (*VC*_*ind*_ = 0.49, IQR = 0.67), and lowest for GLP-1 agonists, which had two indications, Type 2 Diabetes Mellitus and Obesity (*VC*_*ind*_ = <0.01, IQR <0.01).

We repeated the class-wide VC calculations for age, biological sex, and database (Table 1). Comparing within classes, for beta blockers, SGLT-2 inhibitors, fluoroquinolones, and TNF-alpha inhibitors, had *VC*_*ind*_ values greater than *VC*_*sex*_, but not *VC*_*DB*_ or *VC*_*age*_. For trimethoprim, *VC*_*ind*_ was greater than both *VC*_*sex*_ and *VC*_*DB*_, but not *VC*_*age*_, while for JAK inhibitors, GLP-1 Agonists, and Cephalosporins, *VC*_*ind*_ was the smallest of all the median VCs.

#### Distribution of Incidence Rate Variability Across Indications

We plotted histograms of Incidence Rate Ratios (IRRs) to illustrate the distribution of incidence rate differences across indications (Figure 2). IRRs are used to quantify the difference in incidence rates between two specific indications of a given drug class (e.g., indication A versus indication B for beta blockers) for each drug-indication-outcome triad. The incidence rates used to calculate these IRRs were pooled across the remaining stratifying factors not being directly compared.

**Figure 2.**
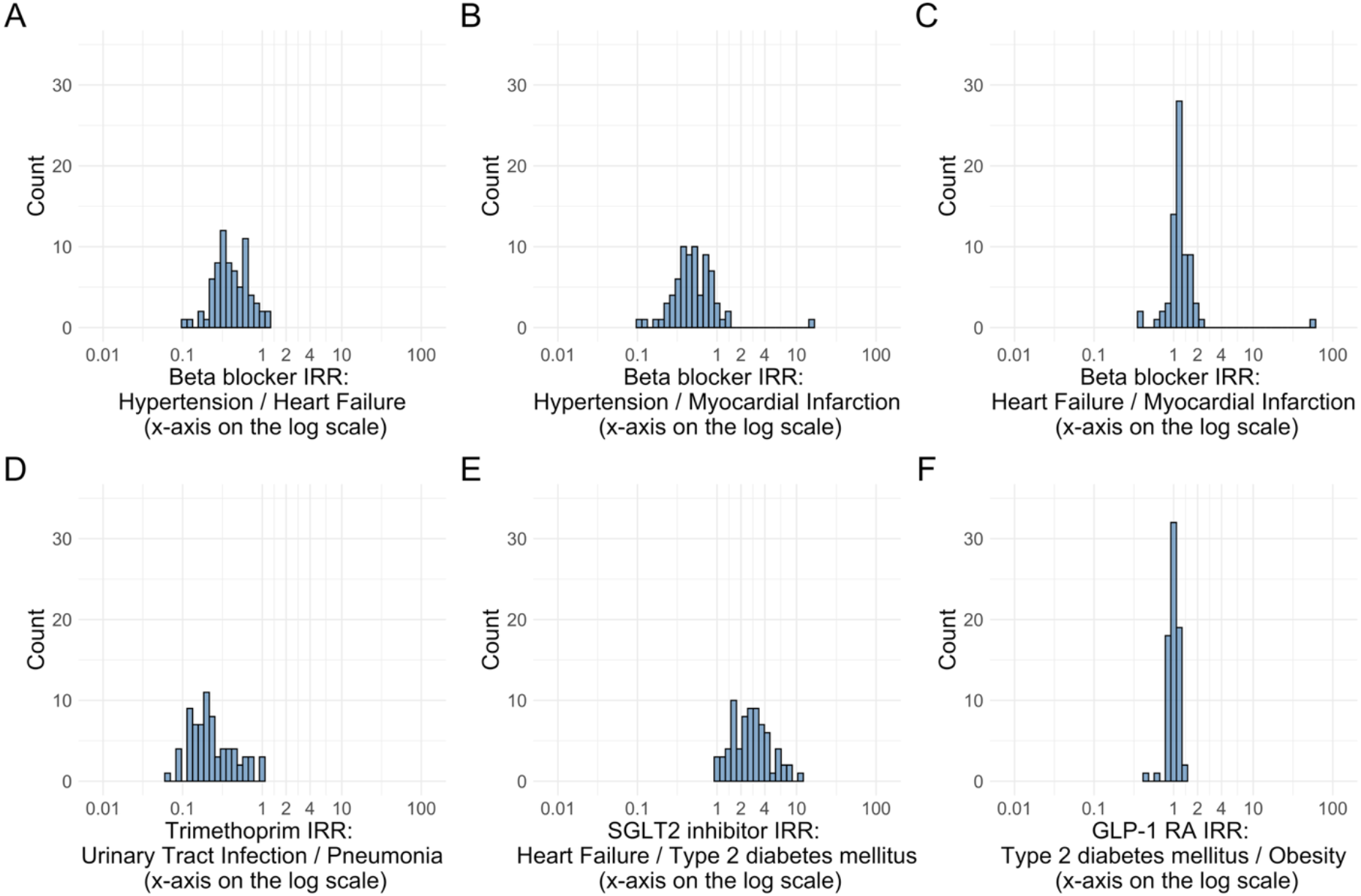
Histograms of IRRs comparing different indications within selected drug classes (Panel A-C: Beta blockers, Panel D: Trimethoprim, Panel E: SGLT-2 inhibitors, Panel F: GLP-1 agonists).

The results from the histograms were congruent with the median *VC*_*ind*_ results from Table 1. For example, a higher median VC attributed to indication, such as trimethoprim (*VC*_*ind*_ = 0.49) generally exhibited a wider spread in the IRR histogram. In contrast, drug classes with a lower median VC for indication, like GLP-1 agonists (*VC*_*ind*_ < 0.01) showed a narrower spread in the IRR histograms. Because we had 8 different drug classes, we present only the 3 drug classes with the highest median *VC*_*ind*_ (trimethoprim, SGLT-2 inhibitors, and beta blockers), and the drug class with the lowest median *VC*_*ind*_ (GLP-1) in Figure 2. Table 2 shows the most extreme IRRs observed across these indication comparisons. The remaining class-wide plots are available in the supplement (Supplementary Material S4, Figures S4.01-S4.19).

**Table 2.** IRRs with the greatest differences between drug-classes. Note: Some of the extreme IRRs may reflect an obvious, direct relationship between the indication and outcome (ex: SGLT-2, left heart failure (indication), and Hospitalization with heart failure events (outcome)). Instead of removing them, we opt to report them for completeness in this table.

| Drug Class | Indication comparison | Outcome | IRR |
| --- | --- | --- | --- |
| Trimethoprim | Urinary Tract Infection vs Acute Typical Pneumonia | Persons with vertigo | 1.073 |
|  |  | Narcolepsy | 1.020 |
|  |  | Bladder Cancer | 0.9597 |
|  |  | Febrile Neutropenia or Neutropenic Fever | 0.0652 |
|  |  | Hospitalization with heart failure events | 0.0910 |
|  |  | Cardiovascular-related mortality | 0.0924 |
| SGLT-2 Inhibitors | Left Heart Failure vs. Type 2 Diabetes | Hospitalization with heart failure events | 10.85 |
|  |  | Total cardiovascular disease events (ischemic stroke, hemorrhagic stroke, heart failure, acute myocardial infarction or sudden cardiac death) | 7.807 |
|  |  | 4-point MACE | 7.708 |
|  |  | Thyroid tumor | 0.9570 |
|  |  | All events of Autoimmune hepatitis, with a washout period of 365 days | 1.032 |
|  |  | Bells Palsy | 1.051 |
| Beta Blockers | Essential Hypertension vs. Left Heart Failure | Persons with vertigo | 1.091 |
|  |  | Thyroid tumor | 1.087 |
|  |  | Headache events | 1.066 |
|  |  | Hospitalization with heart failure events | 0.1099 |
|  |  | Cardiovascular-related mortality | 0.1247 |
|  |  | Total cardiovascular disease events (ischemic stroke, hemorrhagic stroke, heart failure, acute myocardial infarction or sudden cardiac death) | 0.1678 |
| Beta Blockers | Essential Hypertension vs. Acute Myocardial Infarction | Thrombosis with Thrombocytopenia (TWT) | 15.68 |
|  |  | Thyroid tumor | 1.332 |
|  |  | Narcolepsy | 1.274 |
|  |  | Cardiovascular-related mortality | 0.1029 |
|  |  | Acute Myocardial Infarction including its complications | 0.1217 |
|  |  | Coronary Vessel Revascularization | 0.1812 |
| Beta Blockers | Left Heart Failure vs. Acute Myocardial Infarction | Thrombosis with Thrombocytopenia (TWT) | 58.70 |
|  |  | Hospitalization with heart failure events | 2.149 |
|  |  | Total cardiovascular disease events (ischemic stroke, hemorrhagic stroke, heart failure, acute myocardial infarction or sudden cardiac death) | 2.014 |
|  |  | Coronary Vessel Revascularization | 0.3689 |
|  |  | Acute Myocardial Infarction including its complications | 0.4108 |
|  |  | Myocarditis Pericarditis | 0.6019 |
| GLP-1 | Type 2 Diabetes vs. Obesity | Coronary Vessel Revascularization | 1.359 |
|  |  | Guillain Barre Syndrome | 1.279 |
|  |  | Lower extremity amputation | 1.256 |
|  |  | Abnormal weight gain events | 0.4178 |
|  |  | Headache events | 0.6466 |
|  |  | Nausea events | 0.7877 |

#### Distribution of Incidence Rate Variability Across Demographic Factors and Database

To understand the distribution of incidence rate differences across the remaining factors (age, sex, and database), we similarly constructed IRR histograms (Figures 3-5). Each IRR quantifies the difference in incidence rates between two specific subgroups of a given factor (e.g., females versus males for gender; database X vs. database Y) for each drug-indication-outcome triad.

**Figure 3.**
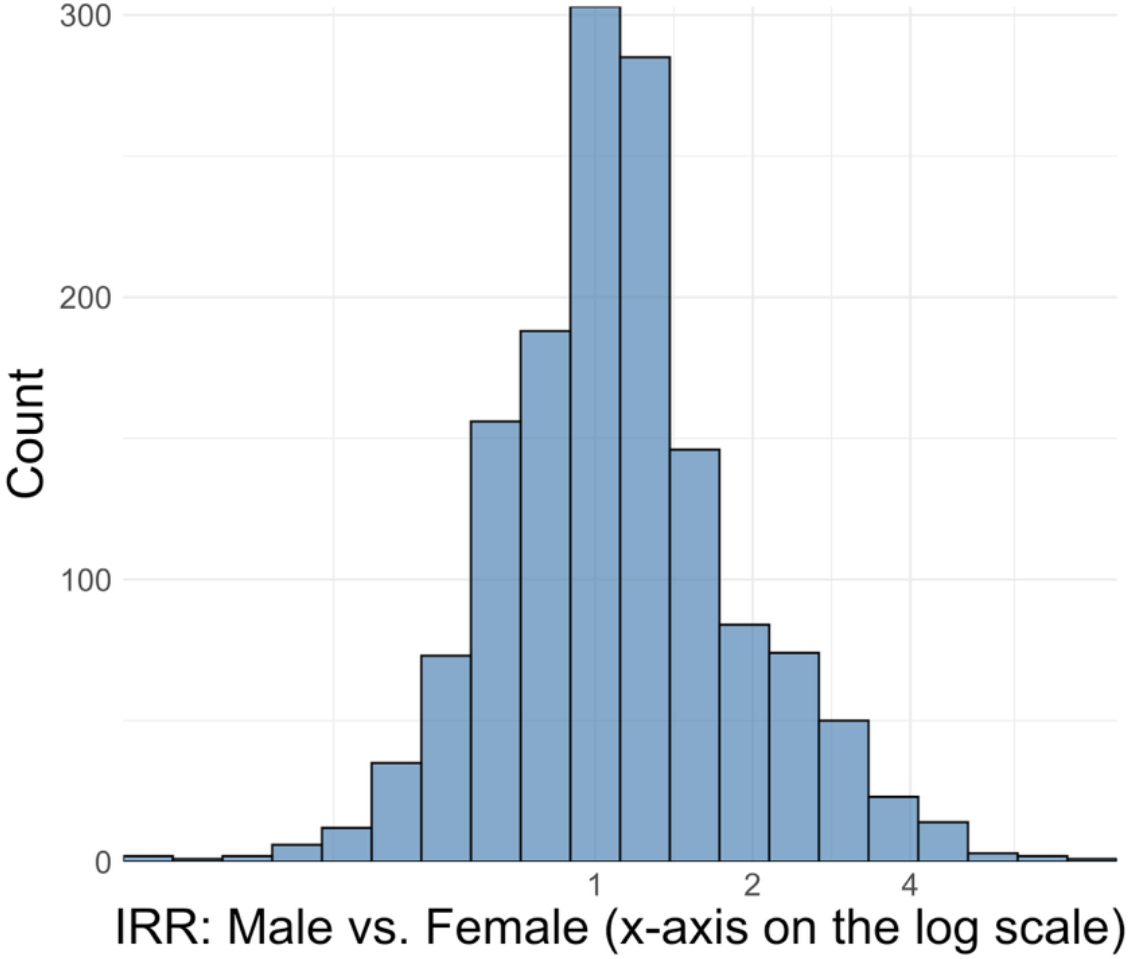
Histogram of IRRs comparing male vs. female, across all drug-indication-outcome pairs. Log scale is used for the X-axis as to better accommodate visualization of IRRs between 0 and 1.

**Figure 4.**
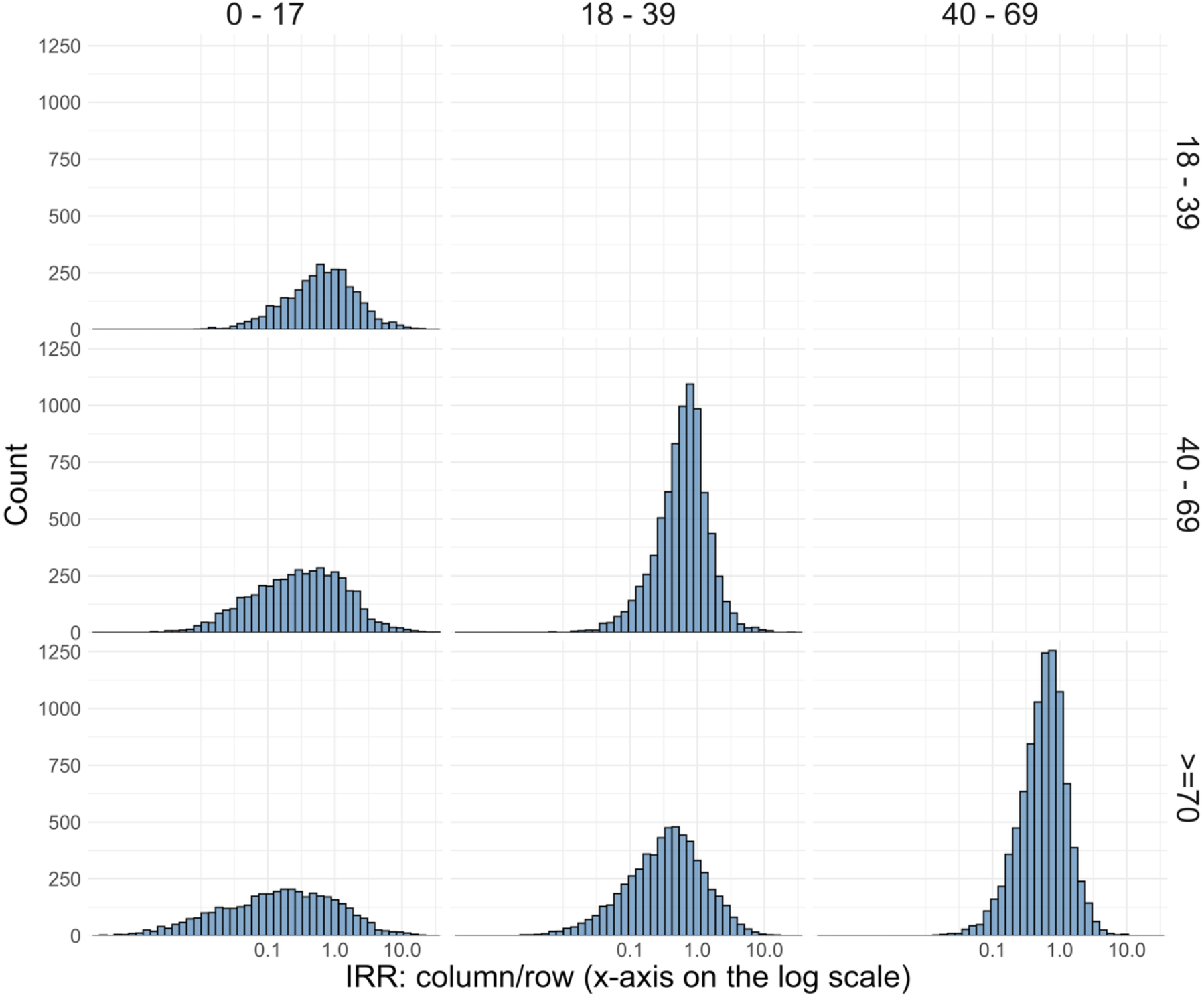
Histogram of IRRs comparing different age groups. Log scale is used for the X-axis as to better accommodate visualization of IRRs between 0 and 1.

**Figure 5.**
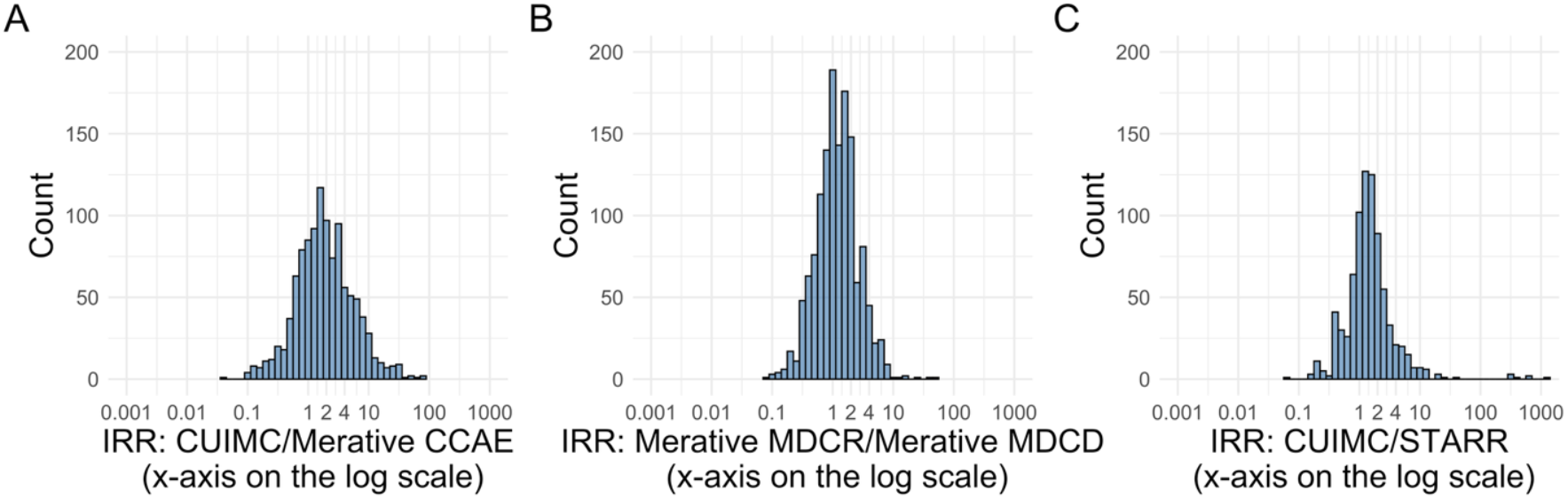
Histograms of IRRs comparing different database sources. Log scale is used for the X-axis as to better accommodate visualization of IRRs between 0 and 1. (Panel A: EHR vs. claims, Panel B: EHR vs. EHR, Panel C: Claims vs. Claims).

Figure 3 shows IRRs comparing male versus female IRs across all drug-indication-outcome pairs, and Table 3 lists the most extreme IRRs. For example, TNF-alpha inhibitors used for plaque psoriasis showed a much lower IR in males compared to females for the health outcome, Severe Cutaneous Adverse Reactions (SCAR) (IRR=0.13).

**Table 3.** IRRs with the greatest sex-based differences in drug-indication-outcome IRs (female vs. male).

| Drug | Indication | Outcome | IRR (Male/Female) |
| --- | --- | --- | --- |
| Trimethoprim | Urinary Tract Infection | Bladder Cancer | 8.529 |
| Tumor Necrosis Factor alpha (TNFa) inhibitors | Psoriatic Arthritis | Bladder Cancer | 6.474 |
| Cephalosporin | Urinary Tract Infection | Bladder Cancer | 6.438 |
| Tumor Necrosis Factor alpha (TNFa) inhibitors | Plaque psoriasis | Severe Cutaneous Adverse Reaction (SCAR = SJS+TEN+DRESS) | 0.1337 |
| Tumor Necrosis Factor alpha (TNFa) inhibitors | Psoriatic Arthritis | Anaphylaxis | 0.1348 |
| Tumor Necrosis Factor alpha (TNFa) inhibitors | Plaque psoriasis | Thyroid tumor | 0.1789 |

To facilitate visualization, we grouped the age deciles into 4 total age groups (0-17, 18-39, 40-69, and ≥70) and pooled the incidence rates, thus reducing the number of pairwise comparisons from 45 to 6, before plotting their IRRs (Figure 4). The most extreme IRRs in each age group comparisons are shown in Table 4.

**Table 4.** IRRs with the greatest age-based differences in drug-indication-outcome IRs.

| Drug | Indication | Outcome | IRR | Age group comparison |
| --- | --- | --- | --- | --- |
| GLP 1-agonists | Obesity | Polymorphic Ventricular Tachycardia or Torsades de Pointes | 40.55 | 0-17 vs. 40-69 |
| GLP 1-agonists | Obesity | All events Drug Rash with Eosinophilia and Systemic Symptoms (DRESS) | 38.43 | 0-17 vs. 40-69 |
| GLP 1-agonists | Obesity | Progressive multifocal leukoencephalopathy | 35.92 | 0-17 vs. 40-69 |
| Trimethoprim | Urinary Tract Infection | Acute Myocardial Infarction including its complications | 0.0002 | 0-17 vs. ≥70 |
| Trimethoprim | Urinary Tract Infection | Persons with heart failure | 0.0003 | 0-17 vs. ≥70 |
| Trimethoprim | Urinary Tract Infection | Acute Myocardial Infarction including its complications | 0.0003 | 0-17 vs. ≥70 |

Finally, we present IRR histograms (Figure 5) and a table of most extreme IRRs (Table 5) for a subset of the 13 database comparisons. These comparisons are chosen to reflect comparisons between a EHR database (CUMC) and a claims database (Merative CCAE), between two different EHR databases (CUMC and STARR), and between two different claims databases that cover different populations (Merative MDCR (older individuals) vs MDCD (individuals with limited income)). The comparison between the two EHR databases (CUMC vs STARR) was the most extreme of all the IRR histograms presented here; with the highest IRR exceeding 1000.

**Table 5.** IRRs with the greatest differences between database sources shown in Figure 4 (CUIMC vs. CCAE, MDCR vs. MDCD, and CUIMC vs. STARR).

| Drug | Indication | Outcome | IRR | Database comparison |
| --- | --- | --- | --- | --- |
| Fluoroquinolone | Acute Typical Pneumonia | Febrile Neutropenia or Neutropenic Fever | 596.64 | CUMC vs STARR |
| Beta Blockers | Essential Hypertension | Febrile Neutropenia or Neutropenic Fever | 631.43 | MDCR vs STARR |
| Beta Blockers | Essential Hypertension | Febrile Neutropenia or Neutropenic Fever | 1202.73 | CUMC vs STARR |
| Tumor Necrosis Factor alpha (TNFa) inhibitors | Crohns Disease | Abnormal weight gain events | 0.0256 | CUMC vs MDCR |
| Tumor Necrosis Factor alpha (TNFa) inhibitors | Rheumatoid Arthritis | Abnormal weight gain events | 0.0323 | CUMC vs MDCR |
| Tumor Necrosis Factor alpha (TNFa) inhibitors | Rheumatoid Arthritis | All events of Acute Liver Injury | 0.0359 | CUMC vs MDCR |

The full set of database comparisons is provided in the supplement (Supplementary Material S5, Figures and Tables S5.01-S5.156).

Finally, we quantify the relative heterogeneity between different stratification factors (age, biological sex, database, and indication) by summarizing the IRRs (Table 6). Notably, database IRRs had the largest IQR, followed by age, indications, then sex.

**Table 6.** Summary of the distributions of all sex, age, and database source IRRs. The last column summarizes the percentage of IRRs that are between 0.5 and 2 to provide a relative measure of how often a factor has less than a doubling effect on the incidence rate.

|  | Median | 1<sup>st</sup> Quartile | 3<sup>rd</sup> Quartile | IQR | Range | % IRR between 0.5-2 |
| --- | --- | --- | --- | --- | --- | --- |
| Sex IRR | 0.91 | 0.70 | 1.24 | 0.54 | [0.11, 7.47] | 81.71% |
| Age IRR | 0.51 | 0.21 | 0.96 | 0.75 | [0, 40.55] | 43.50% |
| Database Source IRR | 1 | 0.4 | 2.2 | 1.76 | [0, 581913] | 43.13% |
| Indications IRR | 0.86 | 0.54 | 1.19 | 0.65 | [0.06, 58.69] | 71.52% |

## Discussion

In this study, we investigated the extent to which drug indication influences incidence rate estimates across multiple drug classes and outcomes, while adjusting for other factors. Our findings showed that in general, drug indication contributes more to incidence rate heterogeneity than sex, but less so than age and database. However, the contribution of indication to incidence variability differs substantially across drug classes.

### Indication differences

Trimethoprim, SGLT-2 inhibitors, and beta blockers exhibit substantial incidence rate variation due to indications. This difference in indication may be due to the distinct clinical profiles for patients being treated for different indications.

Patients treated with a beta blocker for Essential Hypertension, versus those treated for Left Heart Failure or Acute Myocardial Infarction, may differ significantly in baseline cardiovascular risk, acuity, and polypharmacy. Similarly, the baseline risk profiles for patients treated with an SGLT-2 inhibitor for heart failure may be different from those treated with an SGLT-2 inhibitor for heart failure. This may explain the observed heterogeneity that persisted even after adjusting for age and sex.

Trimethoprim is another drug class where high indication-driven differences were observed. This may be because trimethoprim is not a typical treatment for pneumonia ^21,22^. Thus, patients on trimethoprim for pneumonia may have been on that drug due to their medical history (ex, allergies), risk factors, or local resistance patterns. Furthermore, polypharmacy was not accounted for in our phenotyping algorithm, so it is possible that these patients are additionally on a macrolide, the current first-line recommendation for community-acquired pneumonia, and the trimethoprim was added on for another reason.

While pneumonia is an atypical indication for trimethoprim, we chose the two indications (UTI and pneumonia) in order to contrast it with the other antibiotics in the study (cephalosporins and fluoroquinolones). Trimethoprim had the largest *VC*_*ind*_ among the antibiotic classes studied here. This could stem from the fact that patient populations receiving the other antibiotics have a more similar baseline risk profile across both UTI and pneumonia (i.e. if used for more consistently severe or complex cases in both indications; or used consistently for non-complex cases across both indications), leading to more uniformity across the side effect profiles of these drug classes across indications.

In contrast to beta blockers, SGLT-2 inhibitors, and trimethoprim, GLP-1 agonists exhibited minimal incidence rate differences across indications. This pattern may reflect the fact that the two indications studied, Type 2 Diabetes Mellitus and Obesity, have overlapping patient characteristics and risk profiles for common health outcomes. This result suggests that pooling across indications for GLP-1 agonists might be reasonable.

### Age, sex, and database differences

Beyond indication, we assessed the contribution of age, sex, and database to incidence rate variability. Age was frequently the strongest contributor to incidence variation, with 5/8 of our drug classes having their largest median VC attributed to age, but database came at a close second, with 3/8 of our drug classes having their largest median VC attributed to database. Biological sex, on the other hand, was the weakest contributor to incidence rate variation among our 8 drug classes, with 5/8 of drug classes having the smallest median VC attributed to sex. This indicates that incidence rates for the studied outcomes are relatively similar between males and females, especially after adjusting for database, age, and indication.

The results from the IRR plots were congruent with the findings from the VC calculations: for example, the IQR of IRRs for database sources were the most extreme, followed by age, indications then sex. We also found that most IRR histograms were unimodal, but we still observed extreme IRRs. Some potential explanations for these outliers include biological differences (e.g., women might be more susceptible to auto-immune related health outcomes for certain drug-indication pairs) or prescribing biases due to age, database-specific practices, or patient populations.

It is important to note that the variability attributed to “database” encapsulates a multitude of underlying factors beyond simple data recording differences. These can include variations in geographic location, local healthcare system structures, specific clinical practices, coding behaviors, patient population characteristics not fully captured by age and sex (e.g., underlying severity of illness, comorbidities common in that region/system), and even genomic or environmental factors prevalent in the populations covered by each database.

### Limitations

A potential limitation in this study is its observational nature, which are subject to unmeasured confounding that could influence the observed incidence rates despite our adjustments for age, sex, and database. However, the objective of this study was *not* to establish causal relationships, but to assess the sensitivity of incidence rate estimates to drug indication. As such, while unmeasured factors like healthcare utilization, patient clinical presentation, or health state awareness may influence the observed rates, they do not invalidate the descriptive comparisons that form the basis of our findings. Our phenotype algorithms used to define drug exposures, indications, and health outcomes may be another limitation as they could be subject to measurement error. This could lead to inaccuracies in the calculated incidence rates. In order to mitigate this risk, we used previously validated phenotype algorithms when possible.

Future work may include considering additional stratification factors such as disease severity, comorbidities, off-label use, and additional formulations (ex, topical beta-blockers, as opposed to only oral beta-blockers in this study). Because our findings were limited to the 8 drug classes and 73 outcomes analyzed, it may be beneficial to do a large-scale study of all drugs and all side effects and see whether different patterns emerge.

## Conclusion

This study examined the impact of drug indication on incidence rate estimation across a diverse set of drug classes and health outcomes. Stratifying across age, sex, and database is common practice in observational studies, but the results from this study suggests that indication should also be explicitly considered as a stratification factor, especially for drug classes where different indications correspond to markedly different patient populations in terms of baseline severity.

## Data Availability

https://results.ohdsi.org

