## Supplements S1-3 for "The Role Of Drug Indication On Incidence Rate Heterogeneity: A Large-Scale, Systematic Evaluation Across An International Network Of Observational Databases"

Supplement Table S1: Data Sources

| Data source | Description |
|---|---|
| Merative MarketScan Commercial Claims and Encounters Database (CCAE) | Merative MarketScan Commercial Claims and Encounters Database (CCAE) is a US employer-based private-payer administrative claims database. The data include adjudicated health insurance claims (e.g. inpatient, outpatient, and outpatient pharmacy) as well as enrollment data from large employers and health plans who provide private healthcare coverage to employees, their spouses, and dependents. Additionally, it captures laboratory tests for a subset of the covered lives. This administrative claims database includes a variety of fee-for-service, preferred provider organizations, and capitated health plans. |
| Columbia University Irving Medical Center (CUIMC) | The clinical data warehouse of NewYork-Presbyterian Hospital/Columbia University Irving Medical Center, New York, NY, based on its current and previous electronic health record systems, with data spanning over 30 years and including over 6 million patients |
| Merative MarketScan Multi-State Medicaid Database (MDCD) | Merative MarketScan Multi-State Medicaid Database (MDCD) contains adjudicated US health insurance claims for Medicaid enrollees from multiple states and includes hospital discharge diagnoses, outpatient diagnoses and procedures, and outpatient pharmacy claims as well as ethnicity and Medicare eligibility. Members maintain their same identifier even if they leave the system for a brief period; however the dataset lacks lab data. |
| Merative MarketScan Medicare Supplemental and Coordination of Benefits Database (MDCR) | Merative MarketScan Medicare Supplemental and Coordination of Benefits Database (MDCR) represents health services of retirees in the United States with primary or Medicare supplemental coverage through privately insured fee-for-service, point-of-service, or capitated health plans. These data include adjudicated health insurance claims (e.g. inpatient, outpatient, and outpatient pharmacy).Additionally, it captures laboratory tests for a subset of the covered lives. |
| PharMetrics Plus | PharMetrics Plus is a US-based, de-identified, longitudinal patient-level database containing adjudicated medical and pharmacy claims. It includes data on inpatient and outpatient services, prescription drugs (retail and mail-order), office-administered drugs, procedures, diagnoses (ICD-9/10 coded), costs, and enrollment information across diverse sites of care for over 318 million individuals. |
| Optum® De-Identified Clinformatics® Data Mart Database – Socio-Economic Status (Optum SES) | Optum© De-Identified Clinformatics® Data Mart Database (Optum Insight, Eden Prairie, MN) is an adjudicated administrative health claims database for members with private health insurance, who are fully insured in commercial plans or in administrative services only (ASOs), Legacy Medicare Choice Lives (prior to January 2006), and Medicare Advantage (Medicare Advantage Prescription Drug coverage starting January 2006). The population is primarily representative of US commercial claims patients (0-65 years old) with some Medicare (65+ years old) however ages are capped at 90 years. It includes data captured from administrative claims processed from inpatient and outpatient medical services and prescriptions as dispensed, as well as results for outpatient lab tests processed by large national lab vendors who participate in data exchange with Optum. Optum SES provides socio-economic status for members with both medical and pharmacy coverage and location information for patients at the US Census Division level. |
| Optum© de-identified Electronic Health Record Dataset (Optum EHR) | Optum© de-identified Electronic Health Record Dataset is derived from dozens of healthcare provider organizations in the United States (that include more than 700 hospitals and 7,000 Clinics treating more than 103 million patients) receiving care in the United States. The medical record data includes clinical information, inclusive of prescriptions as prescribed and administered, lab results, vital signs, body |

| | |
|---|---|
| | measurements, diagnoses, procedures, and information derived from clinical Notes using Natural Language Processing (NLP). |
| IQVIA Ambulatory EMR | The IQVIA Ambulatory EMR database is comprised of approximately 80 million patients with a face-to-face physician interaction since 2006 and are sourced from an “opt-in” provider research network of non-hospital (ambulatory) practices. The aggregated database comprises records collected by over 100,000 physicians who are affiliated with over 800 practices (medium to large medical offices, outpatient clinics, and physician networks) from all 50 US states. |
| IQVIA Australia Electronic Medical Records (Australia EMR) | Anonymized patient records of more than 6 million patients in Australia collected from Patient Management software used by GPs during an office visit to document patients’ clinical records. |
| STARR-OMOP (Stanford Healthcare) | STARR-OMOP is the clinical data warehouse of Stanford Medicine, containing de-identified patient data from Stanford Health Care and Stanford Children's Health electronic health record (EHR) systems. It contains data for more than 3.7 million patients. |
| IQVIA Longitudinal Patient Data France (LPD France) | Anonymized patient records of 7.8 million patients in France collected from Patient Management software used by GPs and select specialists during an office visit to document patients’ clinical records |
| Longitudinal Patient Database (LPD) in Italy | Anonymized patient records of 2.5 million patients in Italy collected from Patient Management software used by GPs and select specialists during an office visit to document patients’ clinical records |
| Longitudinal Patient Database (LPD) in Belgium | Anonymized patient records of 1.1 million patients in Belgium collected from Patient Management software used by GPs and select specialists during an office visit to document patients’ clinical records |

Supplement Table S2: Drug Class Definitions, Ingredients, and Selected Indications

| Drug class | Ingredients | Indications |
|---|---|---|
| **Beta Blockers** | metoprolol, atenolol, carvedilol, propranolol, labetalol, bisoprolol, nebivolol, nadolol, betaxolol, acebutolol, pindolol, penbutolol | 1) Essential Hypertension, 2) Left Heart Failure, 3) Acute Myocardial Infarction |
| **SGLT2 Inhibitors** | empagliflozin, dapagliflozin, canagliflozin, ertugliflozin | 1) Type 2 Diabetes Mellitus, 2) Left Heart Failure |
| **GLP-1 agonists** | dulaglutide, liraglutide, semaglutide, exenatide, lixisenatide, albiglutide | 1) Type 2 Diabetes Mellitus, 2) Obesity |
| **Trimethoprim** | trimethoprim | 1) Urinary tract infection, 2) Acute Typical Pneumonia |
| **Cephalosporins** | cephalexin, ceftriaxone, cefazolin, cefdinir, cefuroxime, cefprozil, cefadroxil, cefoxitin, cefditoren, cefpodoxime, cefaclor, ceftazidime, cefixime, cefotetan, cephradine | 1) Urinary Tract Infection, 2) Acute Typical Pneumonia |
| **Fluoroquinolones** | ciprofloxacin, levofloxacin, ofloxacin, moxifloxacin, gatifloxacin, nadifloxacin, besifloxacin, norfloxacin, lomefloxacin, gemifloxacin | 1) Urinary Tract Infection, 2) Acute Typical Pneumonia |
| **JAK inhibitors** | tofacitinib, ruxolitinib, baricitinib | 1) Rheumatoid Arthritis, 2) Ulcerative Colitis |
| **TNF-alpha inhibitors** | adalimumab, infliximab, etanercept, certolizumab pegol, golimumab | 1) Rheumatoid Arthritis, 2) Ulcerative Colitis, 3) Plaque Psoriasis, 4) Psoriatic Arthritis, 5) Crohn’s Disease |

Supplement Table S3: Phenotype Definitions for Outcomes

| ID | Phenotype name | Clean Period | Link to package SQL |
|---|---|---|---|
| 207 | Acquired Pure Red Cell Aplasia | 9999 | https://github.com/OHDSI/PhenotypeLibrary/blob/main/inst/sql/sql_server/207.sql |
| 208 | Febrile Neutropenia or Neutropenic Fever | 30 | https://github.com/OHDSI/PhenotypeLibrary/blob/main/inst/sql/sql_server/208.sql |
| 211 | Pancytopenia, Acquired | 365 | https://github.com/OHDSI/PhenotypeLibrary/blob/main/inst/sql/sql_server/211.sql |
| 213 | Neutropenia or unspecified leukopenia | 365 | https://github.com/OHDSI/PhenotypeLibrary/blob/main/inst/sql/sql_server/213.sql |
| 218 | Rhabdomyolysis | 180 | https://github.com/OHDSI/PhenotypeLibrary/blob/main/inst/sql/sql_server/218.sql |
| 219 | Sudden Cardiac arrest or cardiac death | 9999 | https://github.com/OHDSI/PhenotypeLibrary/blob/main/inst/sql/sql_server/219.sql |
| 222 | Stevens-Johnson syndrome, toxic epidermal necrolysis spectrum | 9999 | https://github.com/OHDSI/PhenotypeLibrary/blob/main/inst/sql/sql_server/222.sql |
| 229 | Progressive multifocal leukoencephalopathy | 9999 | https://github.com/OHDSI/PhenotypeLibrary/blob/main/inst/sql/sql_server/229.sql |
| 275 | Polymorphic Ventricular Tachycardia or Torsades de Pointes | 9999 | https://github.com/OHDSI/PhenotypeLibrary/blob/main/inst/sql/sql_server/275.sql |
| 276 | Sudden Vision Loss | 365 | https://github.com/OHDSI/PhenotypeLibrary/blob/main/inst/sql/sql_server/276.sql |
| 362 | Acute Kidney Injury AKI | 30 | https://github.com/OHDSI/PhenotypeLibrary/blob/main/inst/sql/sql_server/362.sql |
| 720 | Aplastic Anemia | 9999 | https://github.com/OHDSI/PhenotypeLibrary/blob/main/inst/sql/sql_server/720.sql |
| 723 | Earliest event of Acute Hepatic Failure | 9999 | https://github.com/OHDSI/PhenotypeLibrary/blob/main/inst/sql/sql_server/723.sql |
| 724 | Earliest event of Acute Hepatic Failure, NO viral hepatitis or alcoholic hepatic failure | 9999 | https://github.com/OHDSI/PhenotypeLibrary/blob/main/inst/sql/sql_server/724.sql |
| 727 | Angioedema | 180 | https://github.com/OHDSI/PhenotypeLibrary/blob/main/inst/sql/sql_server/727.sql |
| 728 | Autoimmune hemolytic anemia | 90 | https://github.com/OHDSI/PhenotypeLibrary/blob/main/inst/sql/sql_server/728.sql |
| 729 | All events of Autoimmune hepatitis, with a washout period of 365 days | 365 | https://github.com/OHDSI/PhenotypeLibrary/blob/main/inst/sql/sql_server/729.sql |
| 731 | All events of Sudden Hearing Loss, No congenital anomaly or middle or inner ear conditions | 180 | https://github.com/OHDSI/PhenotypeLibrary/blob/main/inst/sql/sql_server/731.sql |
| 732 | All events of Severe Cutaneous Adverse Reaction (SCAR = SJS+TEN+DRESS) | 365 | https://github.com/OHDSI/PhenotypeLibrary/blob/main/inst/sql/sql_server/732.sql |
| 734 | All events Drug Rash with Eosinophilia and Systemic Symptoms (DRESS) | 365 | https://github.com/OHDSI/PhenotypeLibrary/blob/main/inst/sql/sql_server/734.sql |
| 735 | All events of Acute Liver Injury | 90 | https://github.com/OHDSI/PhenotypeLibrary/blob/main/inst/sql/sql_server/735.sql |
| 736 | All events of Acute Liver Injury, NO viral hepatitis or alcoholic hepatic failure | 90 | https://github.com/OHDSI/PhenotypeLibrary/blob/main/inst/sql/sql_server/736.sql |

| | | | |
|---|---|---|---|
| 739 | Isolated Immune Thrombocytopenia (ITP), with a washout period of 365 days | 365 | https://github.com/OHDSI/PhenotypeLibrary/blob/main/inst/sql/sql_server/739.sql |
| 741 | Earliest event of Thrombotic microangiopathy (TMA) or Microangiopathic hemolytic anemia (MAHA) | 9999 | https://github.com/OHDSI/PhenotypeLibrary/blob/main/inst/sql/sql_server/741.sql |
| 884 | Diarrhea events | 30 | https://github.com/OHDSI/PhenotypeLibrary/blob/main/inst/sql/sql_server/884.sql |
| 888 | Gastrointestinal bleeding events | 30 | https://github.com/OHDSI/PhenotypeLibrary/blob/main/inst/sql/sql_server/888.sql |
| 889 | Hyponatremia events | 90 | https://github.com/OHDSI/PhenotypeLibrary/blob/main/inst/sql/sql_server/889.sql |
| 890 | Hypotension events | 90 | https://github.com/OHDSI/PhenotypeLibrary/blob/main/inst/sql/sql_server/890.sql |
| 891 | Nausea events | 30 | https://github.com/OHDSI/PhenotypeLibrary/blob/main/inst/sql/sql_server/891.sql |
| 892 | Stroke (ischemic or hemorrhagic) events | 180 | https://github.com/OHDSI/PhenotypeLibrary/blob/main/inst/sql/sql_server/892.sql |
| 893 | Persons with vertigo | 9999 | https://github.com/OHDSI/PhenotypeLibrary/blob/main/inst/sql/sql_server/893.sql |
| 894 | Abdominal pain events | 90 | https://github.com/OHDSI/PhenotypeLibrary/blob/main/inst/sql/sql_server/894.sql |
| 895 | Abnormal weight gain events | 90 | https://github.com/OHDSI/PhenotypeLibrary/blob/main/inst/sql/sql_server/895.sql |
| 896 | Abnormal weight loss events | 90 | https://github.com/OHDSI/PhenotypeLibrary/blob/main/inst/sql/sql_server/896.sql |
| 920 | Total cardiovascular disease events (ischemic stroke, hemorrhagic stroke, heart failure, acute myocardial infarction or sudden cardiac death) | 180 | https://github.com/OHDSI/PhenotypeLibrary/blob/main/inst/sql/sql_server/920.sql |
| 921 | Cardiovascular-related mortality | 9999 | https://github.com/OHDSI/PhenotypeLibrary/blob/main/inst/sql/sql_server/921.sql |
| 923 | Persons with chronic kidney disease | 9999 | https://github.com/OHDSI/PhenotypeLibrary/blob/main/inst/sql/sql_server/923.sql |
| 925 | Cough events | 90 | https://github.com/OHDSI/PhenotypeLibrary/blob/main/inst/sql/sql_server/925.sql |
| 929 | Edema events | 180 | https://github.com/OHDSI/PhenotypeLibrary/blob/main/inst/sql/sql_server/929.sql |
| 930 | Persons with end stage renal disease | 9999 | https://github.com/OHDSI/PhenotypeLibrary/blob/main/inst/sql/sql_server/930.sql |
| 931 | Fall events | 180 | https://github.com/OHDSI/PhenotypeLibrary/blob/main/inst/sql/sql_server/931.sql |
| 932 | Persons with gout | 9999 | https://github.com/OHDSI/PhenotypeLibrary/blob/main/inst/sql/sql_server/932.sql |
| 933 | Headache events | 30 | https://github.com/OHDSI/PhenotypeLibrary/blob/main/inst/sql/sql_server/933.sql |
| 934 | Persons with heart failure | 9999 | https://github.com/OHDSI/PhenotypeLibrary/blob/main/inst/sql/sql_server/934.sql |
| 936 | Persons with hepatic failure | 9999 | https://github.com/OHDSI/PhenotypeLibrary/blob/main/inst/sql/sql_server/936.sql |
| 938 | Hospitalization with heart failure events | 30 | https://github.com/OHDSI/PhenotypeLibrary/blob/main/inst/sql/sql_server/938.sql |
| 940 | Hyperkalemia events | 90 | https://github.com/OHDSI/PhenotypeLibrary/blob/main/inst/sql/sql_server/940.sql |

| | | | |
|---|---|---|---|
| 941 | Hypokalemia events | 90 | https://github.com/OHDSI/PhenotypeLibrary/blob/main/inst/sql/sql_server/941.sql |
| 942 | Hypomagnesemia events | 90 | https://github.com/OHDSI/PhenotypeLibrary/blob/main/inst/sql/sql_server/942.sql |
| 956 | Transient ischemic attack events | 30 | https://github.com/OHDSI/PhenotypeLibrary/blob/main/inst/sql/sql_server/956.sql |
| 965 | 3-point MACE | 180 | https://github.com/OHDSI/PhenotypeLibrary/blob/main/inst/sql/sql_server/965.sql |
| 967 | 4-point MACE | 180 | https://github.com/OHDSI/PhenotypeLibrary/blob/main/inst/sql/sql_server/967.sql |
| 980 | Coronary Vessel Revascularization | 30 | https://github.com/OHDSI/PhenotypeLibrary/blob/main/inst/sql/sql_server/980.sql |
| 989 | Bladder Cancer | 9999 | https://github.com/OHDSI/PhenotypeLibrary/blob/main/inst/sql/sql_server/989.sql |
| 990 | Bone Fracture | 90 | https://github.com/OHDSI/PhenotypeLibrary/blob/main/inst/sql/sql_server/990.sql |
| 996 | Hypoglycemia | 30 | https://github.com/OHDSI/PhenotypeLibrary/blob/main/inst/sql/sql_server/996.sql |
| 999 | Lower extremity amputation | 9999 | https://github.com/OHDSI/PhenotypeLibrary/blob/main/inst/sql/sql_server/999.sql |
| 1003 | Renal Cancer | 9999 | https://github.com/OHDSI/PhenotypeLibrary/blob/main/inst/sql/sql_server/1003.sql |
| 1004 | Thyroid tumor | 9999 | https://github.com/OHDSI/PhenotypeLibrary/blob/main/inst/sql/sql_server/1004.sql |
| 1075 | Narcolepsy | 365 | https://github.com/OHDSI/PhenotypeLibrary/blob/main/inst/sql/sql_server/1075.sql |
| 1076 | Anaphylaxis | 30 | https://github.com/OHDSI/PhenotypeLibrary/blob/main/inst/sql/sql_server/1076.sql |
| 1078 | Bells Palsy | 183 | https://github.com/OHDSI/PhenotypeLibrary/blob/main/inst/sql/sql_server/1078.sql |
| 1079 | Encephalomyelitis | 183 | https://github.com/OHDSI/PhenotypeLibrary/blob/main/inst/sql/sql_server/1079.sql |
| 1080 | Guillain Barre Syndrome | 365 | https://github.com/OHDSI/PhenotypeLibrary/blob/main/inst/sql/sql_server/1080.sql |
| 1081 | Acute Myocardial Infarction including its complications | 365 | https://github.com/OHDSI/PhenotypeLibrary/blob/main/inst/sql/sql_server/1081.sql |
| 1082 | Myocarditis Pericarditis | 365 | https://github.com/OHDSI/PhenotypeLibrary/blob/main/inst/sql/sql_server/1082.sql |
| 1083 | Immune Thrombocytopenia (ITP) | 365 | https://github.com/OHDSI/PhenotypeLibrary/blob/main/inst/sql/sql_server/1083.sql |
| 1084 | Disseminated Intravascular Coagulation | 365 | https://github.com/OHDSI/PhenotypeLibrary/blob/main/inst/sql/sql_server/1084.sql |
| 1087 | Hemorrhagic Stroke | 365 | https://github.com/OHDSI/PhenotypeLibrary/blob/main/inst/sql/sql_server/1087.sql |
| 1088 | Deep Vein Thrombosis (DVT) | 365 | https://github.com/OHDSI/PhenotypeLibrary/blob/main/inst/sql/sql_server/1088.sql |
| 1089 | Non-hemorrhagic Stroke | 365 | https://github.com/OHDSI/PhenotypeLibrary/blob/main/inst/sql/sql_server/1089.sql |
| 1090 | Pulmonary Embolism | 365 | https://github.com/OHDSI/PhenotypeLibrary/blob/main/inst/sql/sql_server/1090.sql |
| 1091 | Thrombosis with Thrombocytopenia (TWT) | 365 | https://github.com/OHDSI/PhenotypeLibrary/blob/main/inst/sql/sql_server/1091.sql |
