## Supplement S4 for "The Role Of Drug Indication On Incidence Rate Heterogeneity: A Large-Scale, Systematic Evaluation Across An International Network Of Observational Databases"

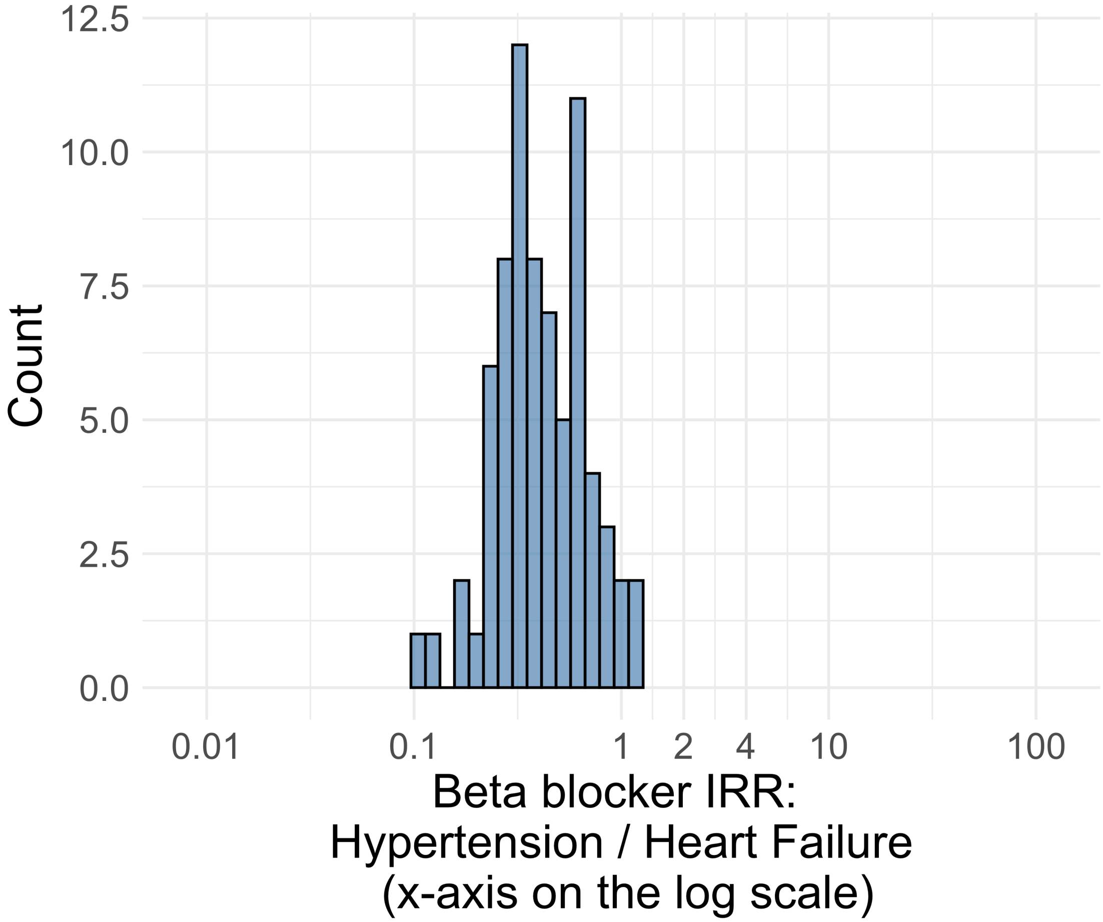


Figure S4.01. IRR distribution comparing Hypertension and Heart_Failure in drug class: Betablocker.

*Table S4.01. Most extreme IRRs for Hypertension vs Heart_Failure (rounded to 0.001) in drug class: Betablocker.*

| Outcome | Indication 1 | Indication 2 | IRR |
|---|---|---|---|
| Persons with vertigo | Hypertension | Heart Failure | 1.1 |
| Thyroid tumor | Hypertension | Heart Failure | 1.1 |
| Headache events | Hypertension | Heart Failure | 1.1 |
| Hospitalization with heart failure events | Hypertension | Heart Failure | 0.1 |
| Cardiovascular-related mortality | Hypertension | Heart Failure | 0.1 |
| Total cardiovascular disease events (ischemic stroke, hemorrhagic stroke, heart failure, acute myocardial infarction or sudden cardiac death) | Hypertension | Heart Failure | 0.2 |

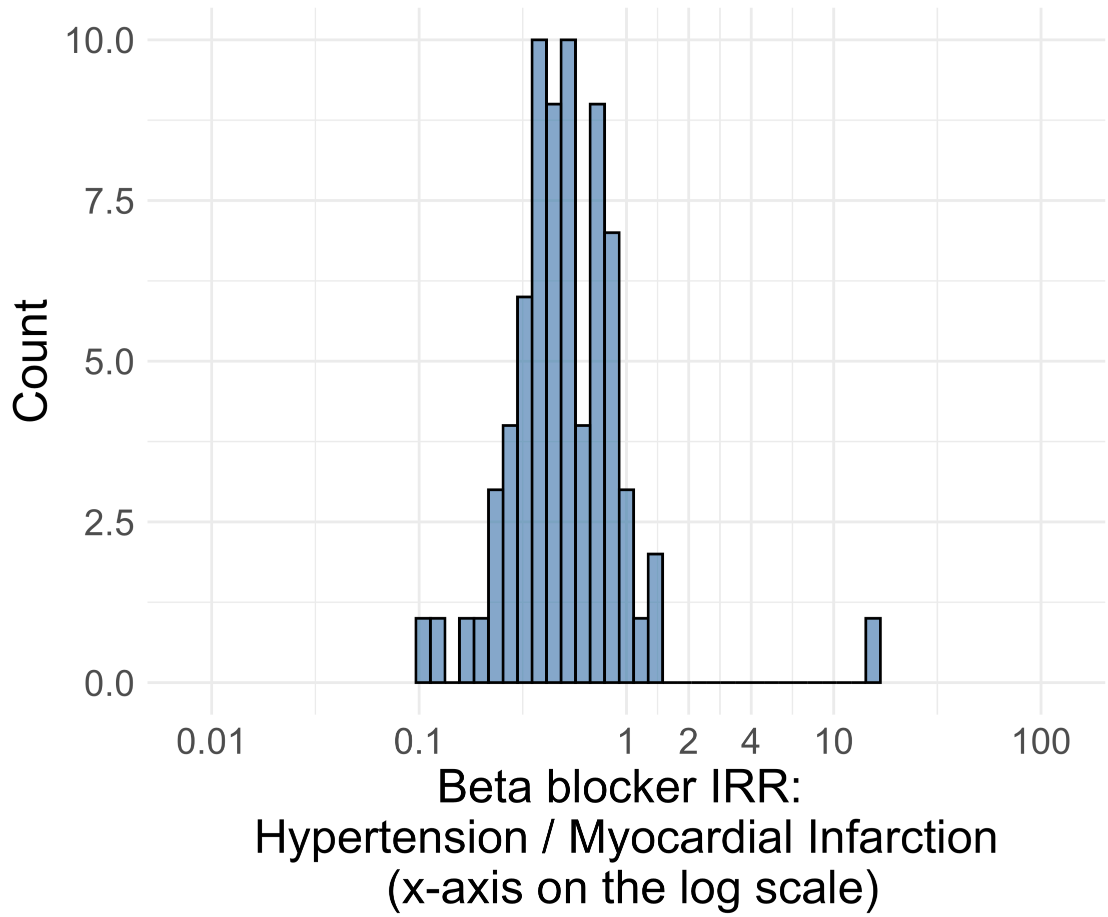


Figure S4.02. IRR distribution comparing Hypertension and Myocardial_Infarction in drug class: Betablocker.

*Table S4.02. Most extreme IRRs for Hypertension vs Myocardial_Infarction (rounded to 0.001) in drug class: Betablocker.*

| Outcome | Indication 1 | Indication 2 | IRR |
|---|---|---|---|
| Thrombosis with Thrombocytopenia (TWT) | Hypertension | Myocardial Infarction | 15.7 |
| Thyroid tumor | Hypertension | Myocardial Infarction | 1.3 |
| Narcolepsy | Hypertension | Myocardial Infarction | 1.3 |
| Cardiovascular-related mortality | Hypertension | Myocardial Infarction | 0.1 |
| Acute Myocardial Infarction including its complications | Hypertension | Myocardial Infarction | 0.1 |
| Coronary Vessel Revascularization | Hypertension | Myocardial Infarction | 0.2 |

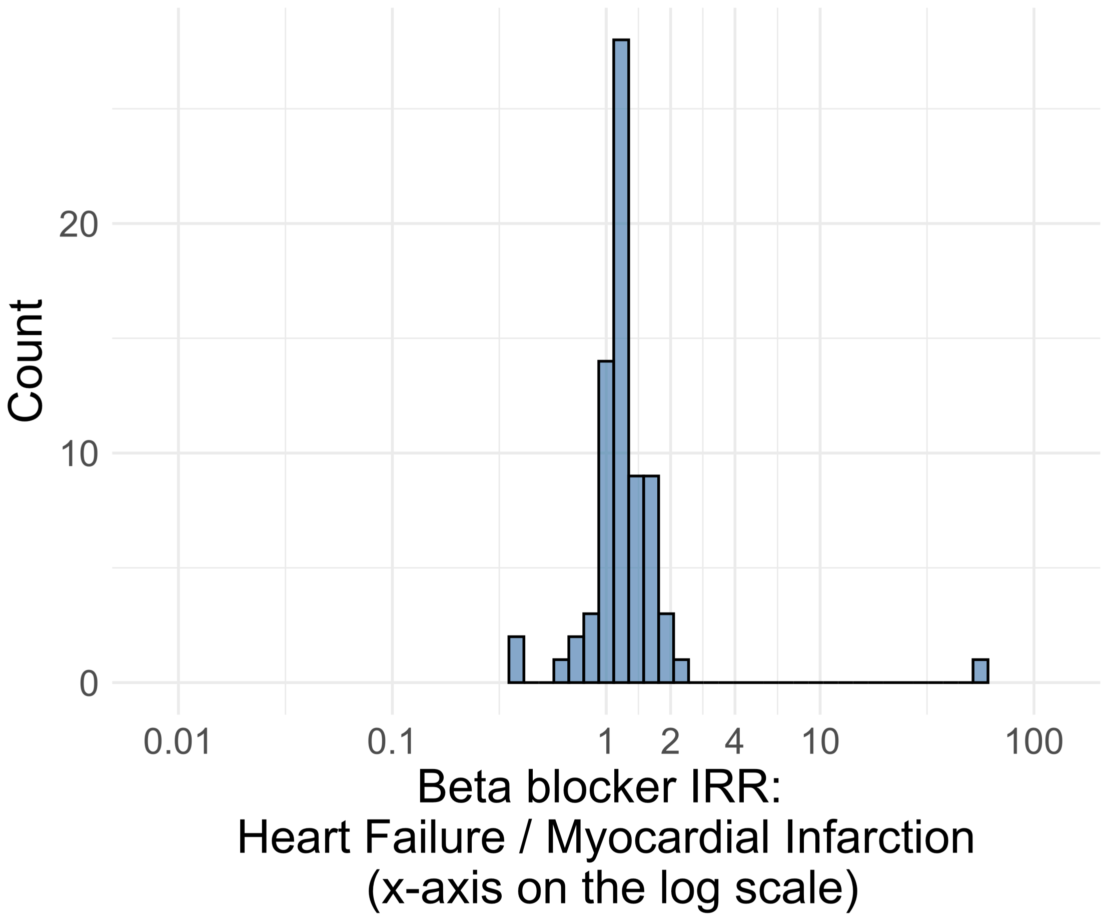


Figure S4.03. IRR distribution comparing Heart_Failure and Myocardial_Infarction in drug class: Betablocker.

*Table S4.03. Most extreme IRRs for Heart_Failure vs Myocardial_Infarction (rounded to 0.001) in drug class: Betablocker.*

| Outcome | Indication 1 | Indication 2 | IRR |
|---|---|---|---|
| Thrombosis with Thrombocytopenia (TWT) | Heart Failure | Myocardial Infarction | 58.7 |
| Hospitalization with heart failure events | Heart Failure | Myocardial Infarction | 2.1 |
| Total cardiovascular disease events (ischemic stroke, hemorrhagic stroke, heart failure, acute myocardial infarction or sudden cardiac death) | Heart Failure | Myocardial Infarction | 2.0 |
| Coronary Vessel Revascularization | Heart Failure | Myocardial Infarction | 0.4 |
| Acute Myocardial Infarction including its complications | Heart Failure | Myocardial Infarction | 0.4 |
| Myocarditis Pericarditis | Heart Failure | Myocardial Infarction | 0.6 |

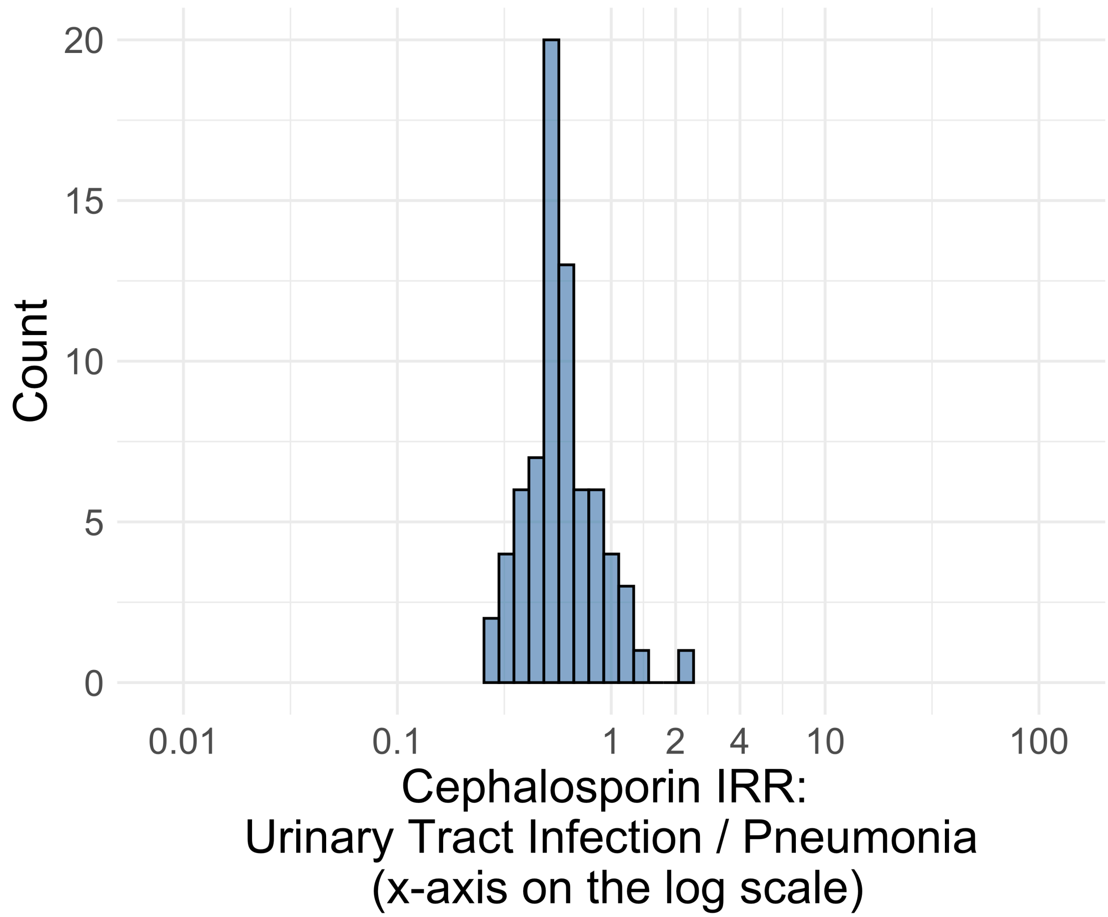


Figure S4.04. IRR distribution comparing Urinary_Tract_Infection and Pneumonia in drug class: Cephalosporin.

*Table S4.04. Most extreme IRRs for Urinary_Tract_Infection vs Pneumonia (rounded to 0.001) in drug class: Cephalosporin.*

| Outcome | Indication 1 | Indication 2 | IRR |
|---|---|---|---|
| Bladder Cancer | Urinary Tract Infection | Pneumonia | 2.3 |
| Persons with vertigo | Urinary Tract Infection | Pneumonia | 1.4 |
| Renal Cancer | Urinary Tract Infection | Pneumonia | 1.2 |
| Cardiovascular-related mortality | Urinary Tract Infection | Pneumonia | 0.3 |
| Sudden Cardiac arrest or cardiac death | Urinary Tract Infection | Pneumonia | 0.3 |
| Hospitalization with heart failure events | Urinary Tract Infection | Pneumonia | 0.3 |

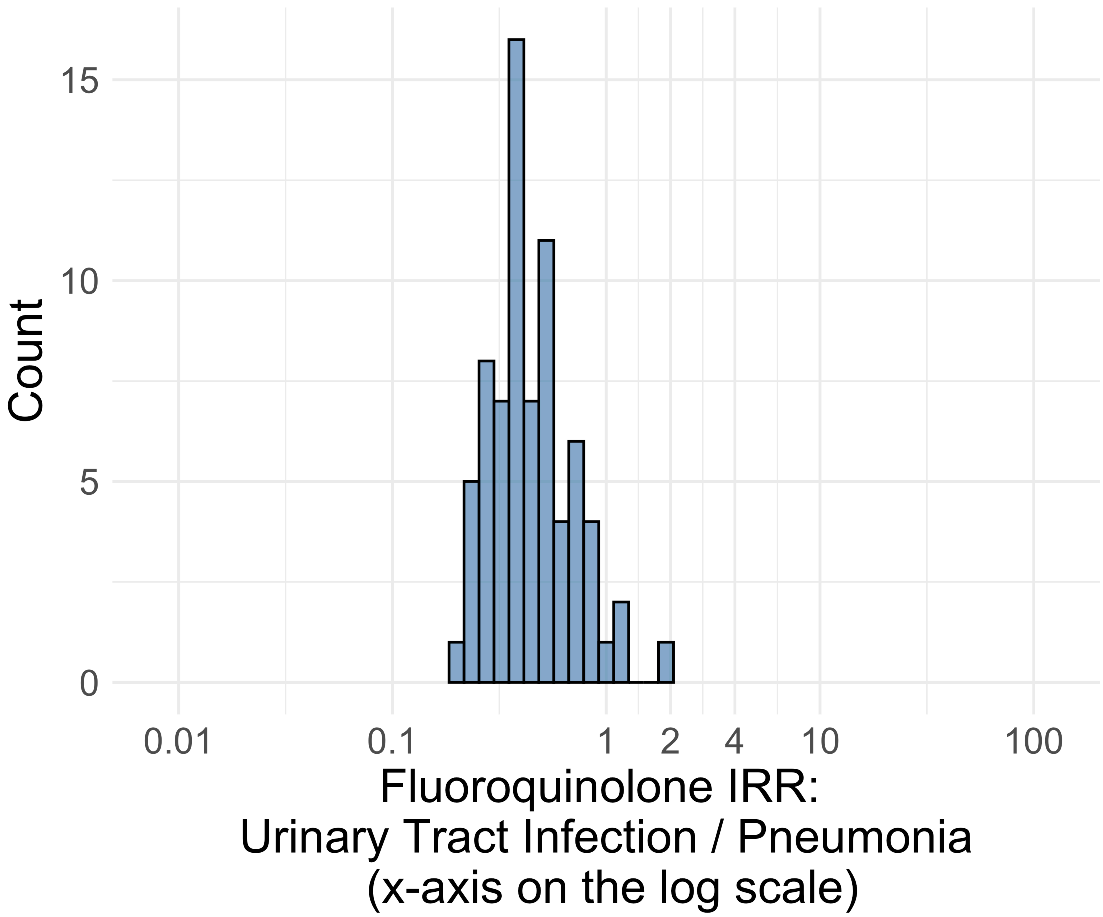


Figure S4.05. IRR distribution comparing Urinary_Tract_Infection and Pneumonia in drug class: Fluoroquinolone.

*Table S4.05. Most extreme IRRs for Urinary_Tract_Infection vs Pneumonia (rounded to 0.001) in drug class: Fluoroquinolone.*

| Outcome | Indication 1 | Indication 2 | IRR |
|---|---|---|---|
| Bladder Cancer | Urinary Tract Infection | Pneumonia | 2.0 |
| Abdominal pain events | Urinary Tract Infection | Pneumonia | 1.2 |
| Persons with vertigo | Urinary Tract Infection | Pneumonia | 1.1 |
| Cardiovascular-related mortality | Urinary Tract Infection | Pneumonia | 0.2 |
| Myocarditis Pericarditis | Urinary Tract Infection | Pneumonia | 0.2 |
| Sudden Cardiac arrest or cardiac death | Urinary Tract Infection | Pneumonia | 0.2 |

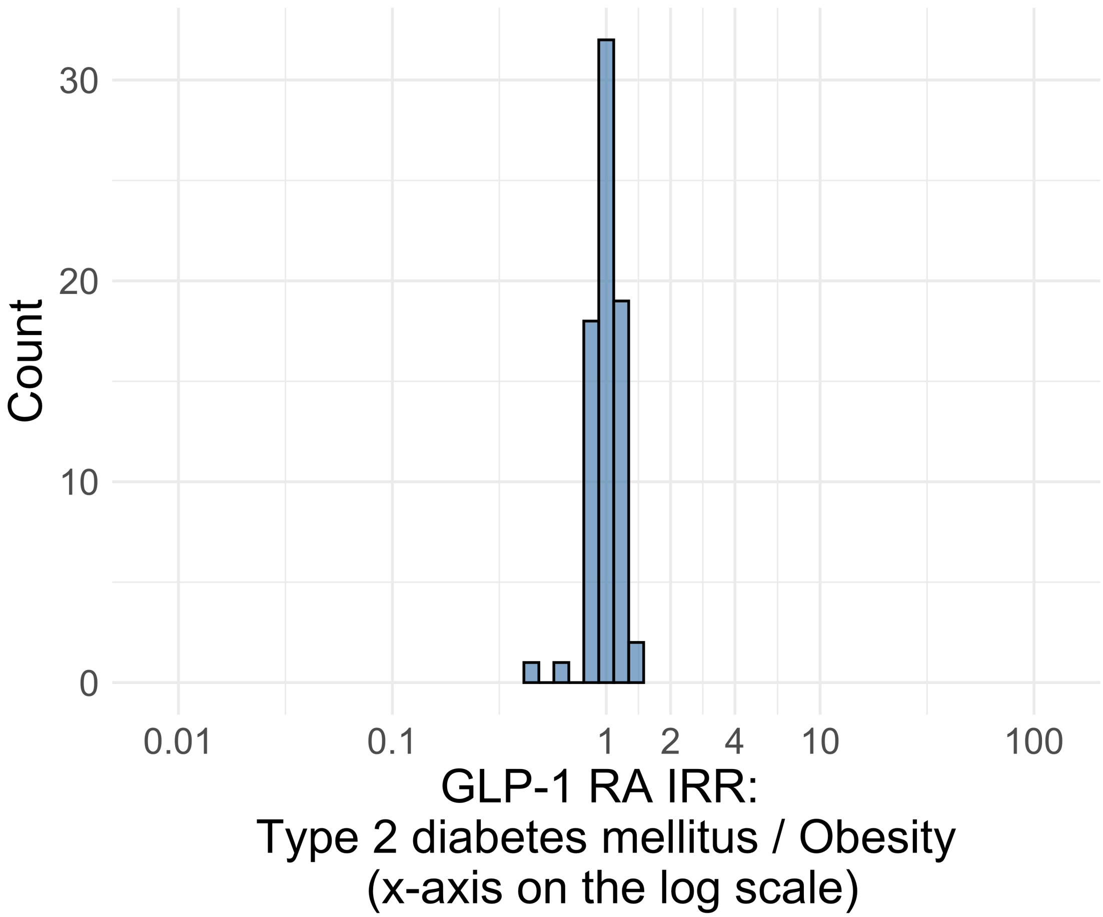


Figure S4.06. IRR distribution comparing Type_2_diabetes_mellitus and Obesity in drug class: GLP1RA.

*Table S4.06. Most extreme IRRs for Type_2_diabetes_mellitus vs Obesity (rounded to 0.001) in drug class: GLP1RA.*

| Outcome | Indication 1 | Indication 2 | IRR |
|---|---|---|---|
| Coronary Vessel Revascularization | Type 2 diabetes mellitus | Obesity | 1.4 |
| Guillain Barre Syndrome | Type 2 diabetes mellitus | Obesity | 1.3 |
| Lower extremity amputation | Type 2 diabetes mellitus | Obesity | 1.3 |
| Abnormal weight gain events | Type 2 diabetes mellitus | Obesity | 0.4 |
| Headache events | Type 2 diabetes mellitus | Obesity | 0.6 |
| Nausea events | Type 2 diabetes mellitus | Obesity | 0.8 |

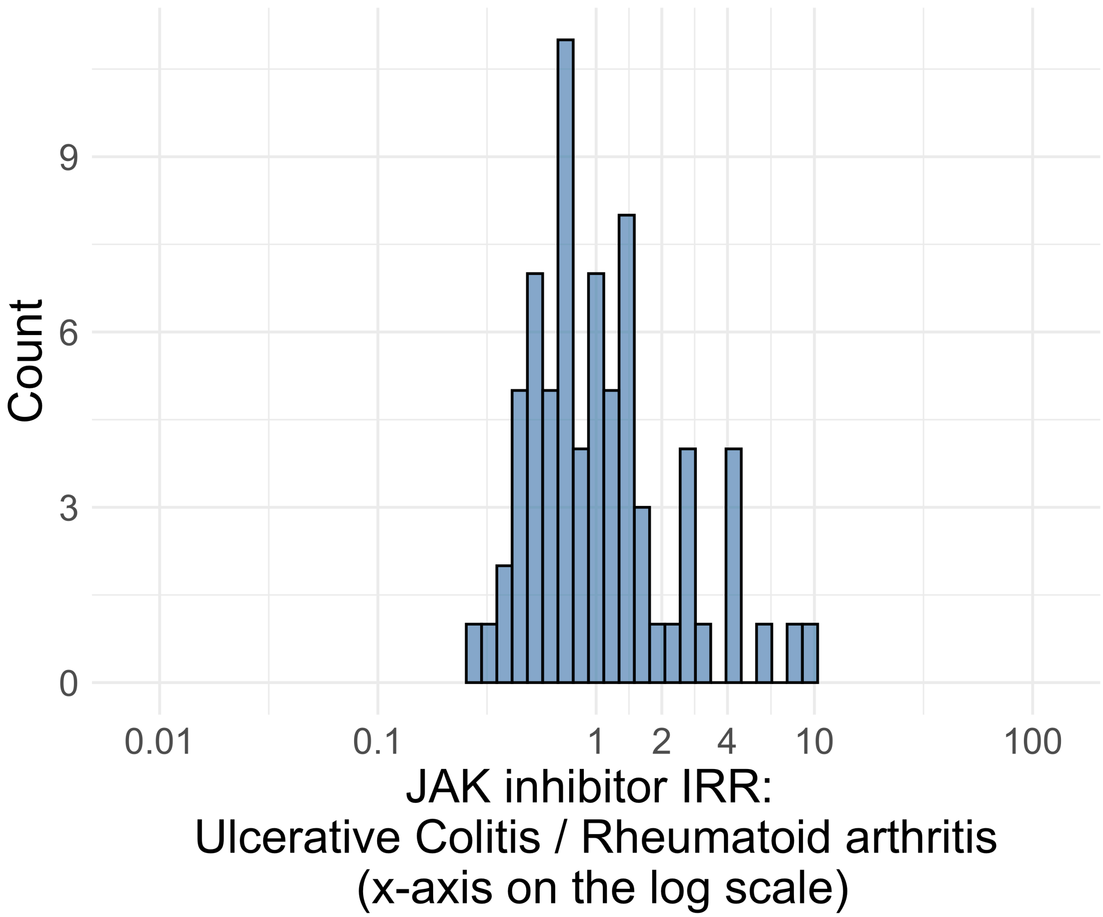


Figure S4.07. IRR distribution comparing Ulcerative_Colitis and Rheumatoid_arthritis in drug class: JAKinhibitor.

*Table S4.07. Most extreme IRRs for Ulcerative_Colitis vs Rheumatoid_arthritis (rounded to 0.001) in drug class: JAKinhibitor.*

| Outcome | Indication 1 | Indication 2 | IRR |
|---|---|---|---|
| Gastrointestinal bleeding events | Ulcerative Colitis | Rheumatoid arthritis | 9.1 |
| Acquired Pure Red Cell Aplasia | Ulcerative Colitis | Rheumatoid arthritis | 8.2 |
| Polymorphic Ventricular Tachycardia or Torsades de Pointes | Ulcerative Colitis | Rheumatoid arthritis | 5.8 |
| Lower extremity amputation | Ulcerative Colitis | Rheumatoid arthritis | 0.3 |
| Persons with gout | Ulcerative Colitis | Rheumatoid arthritis | 0.3 |
| Anaphylaxis | Ulcerative Colitis | Rheumatoid arthritis | 0.4 |

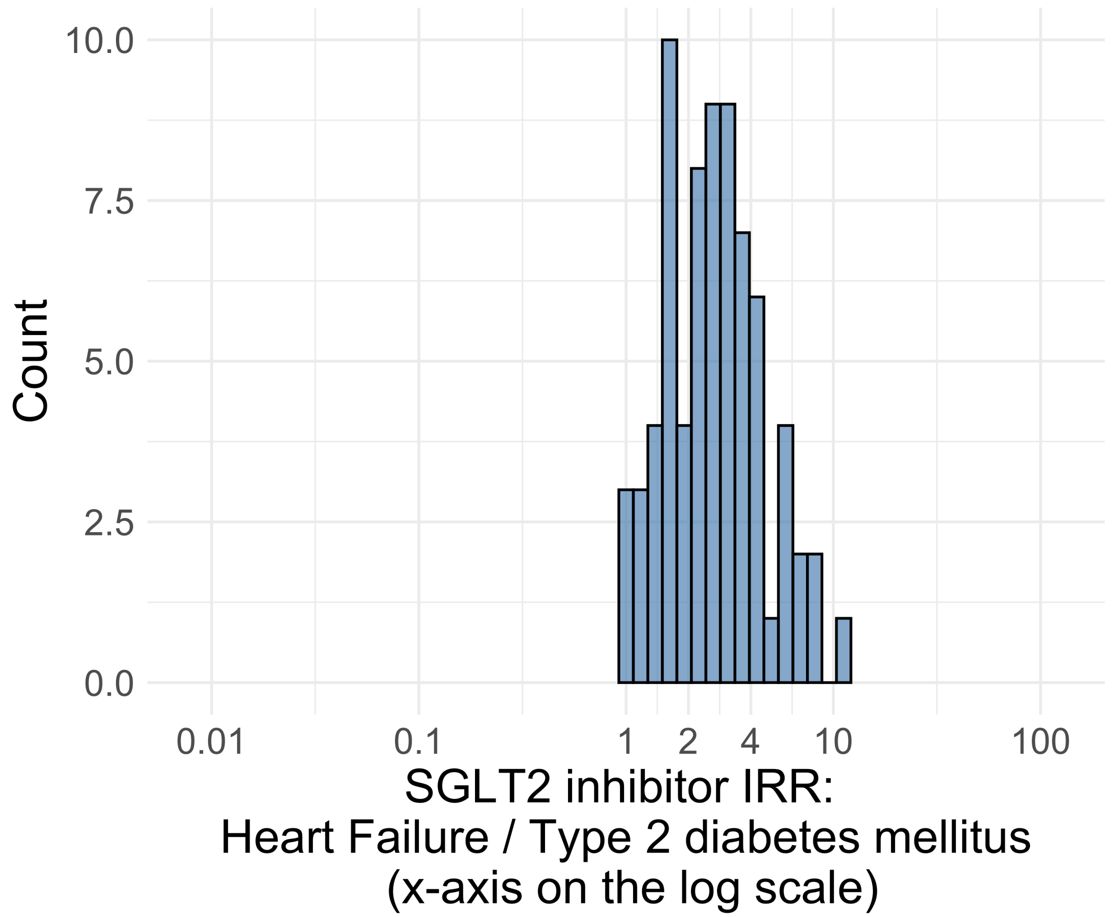


Figure S4.08. IRR distribution comparing Heart_Failure and Type_2_diabetes_mellitus in drug class: SGLT2inhibitor.

*Table S4.08. Most extreme IRRs for Heart_Failure vs Type_2_diabetes_mellitus (rounded to 0.001) in drug class: SGLT2inhibitor.*

| Outcome | Indication 1 | Indication 2 | IRR |
|---|---|---|---|
| Hospitalization with heart failure events | Heart Failure | Type 2 diabetes mellitus | 10.9 |
| Total cardiovascular disease events (ischemic stroke, hemorrhagic stroke, heart failure, acute myocardial infarction or sudden cardiac death) | Heart Failure | Type 2 diabetes mellitus | 7.8 |
| 4-point MACE | Heart Failure | Type 2 diabetes mellitus | 7.7 |
| Thyroid tumor | Heart Failure | Type 2 diabetes mellitus | 1.0 |
| All events of Autoimmune hepatitis, with a washout period of 365 days | Heart Failure | Type 2 diabetes mellitus | 1.0 |
| Bells Palsy | Heart Failure | Type 2 diabetes mellitus | 1.1 |

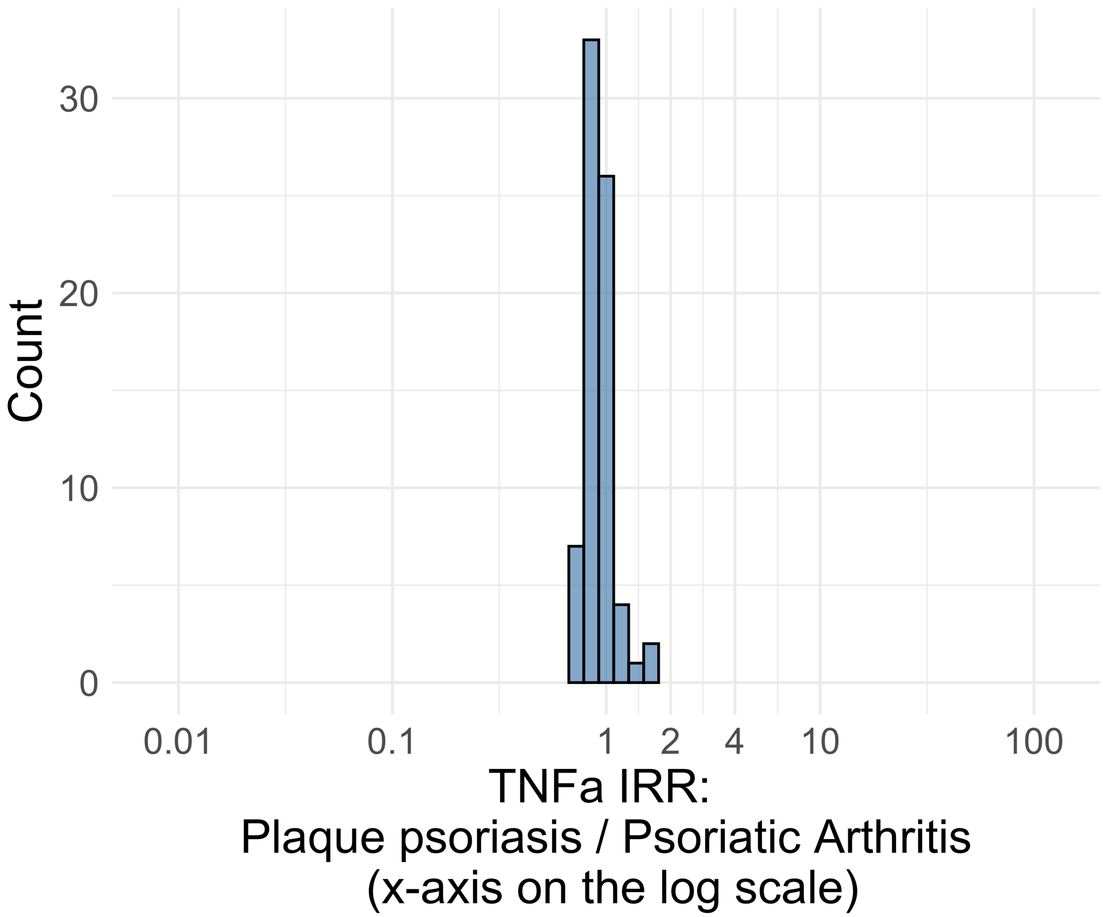


Figure S4.09. IRR distribution comparing Plaque_psoriasis and Psoriatic_Arthritis in drug class: TNFa.

*Table S4.09. Most extreme IRRs for Plaque_psoriasis vs Psoriatic_Arthritis (rounded to 0.001) in drug class: TNFa.*

| Outcome | Indication 1 | Indication 2 | IRR |
|---|---|---|---|
| Disseminated Intravascular Coagulation | Plaque psoriasis | Psoriatic Arthritis | 1.6 |
| Guillain Barre Syndrome | Plaque psoriasis | Psoriatic Arthritis | 1.5 |
| Persons with hepatic failure | Plaque psoriasis | Psoriatic Arthritis | 1.3 |
| Sudden Vision Loss | Plaque psoriasis | Psoriatic Arthritis | 0.7 |
| Thyroid tumor | Plaque psoriasis | Psoriatic Arthritis | 0.7 |
| All events Drug Rash with Eosinophilia and Systemic Symptoms (DRESS) | Plaque psoriasis | Psoriatic Arthritis | 0.7 |

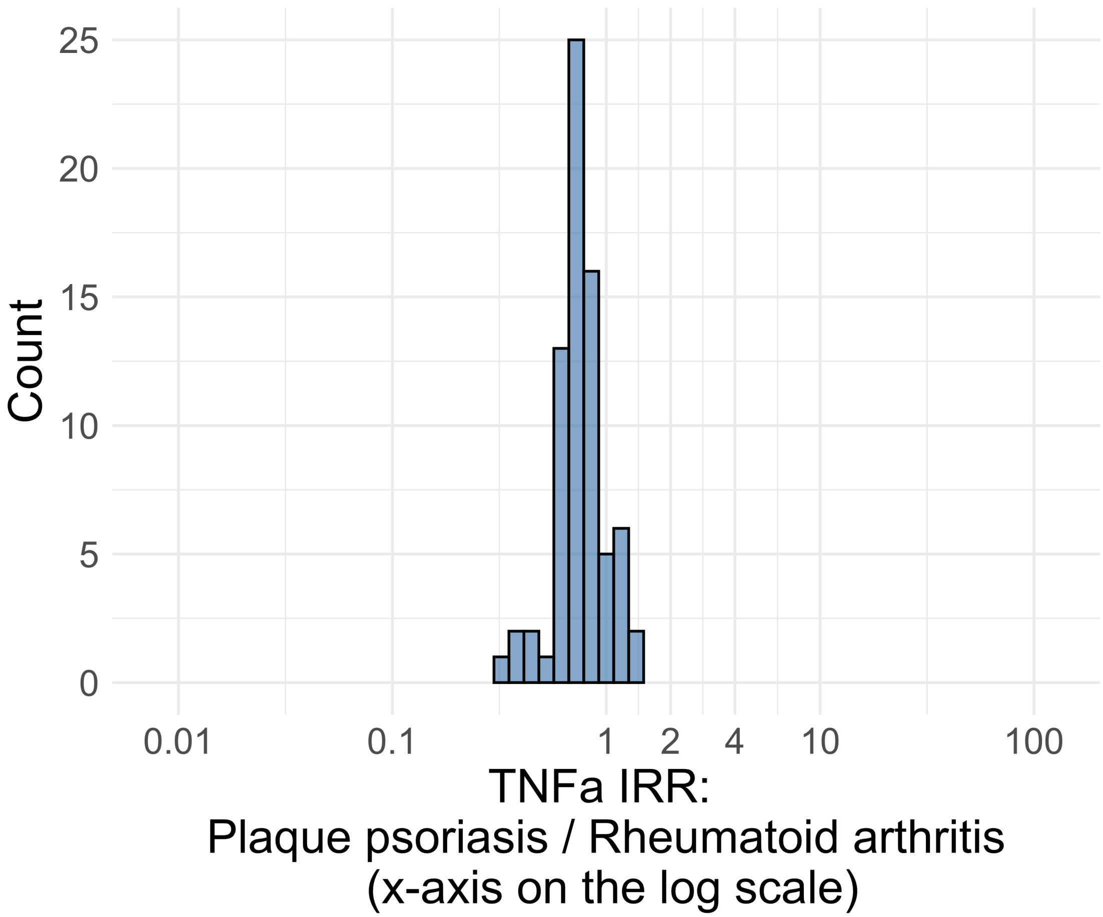


Figure S4.10. IRR distribution comparing Plaque_psoriasis and Rheumatoid_arthritis in drug class: TNFa.

*Table S4.10. Most extreme IRRs for Plaque_psoriasis vs Rheumatoid_arthritis (rounded to 0.001) in drug class: TNFa.*

| Outcome | Indication 1 | Indication 2 | IRR |
|---|---|---|---|
| Persons with hepatic failure | Plaque psoriasis | Rheumatoid arthritis | 1.4 |
| Polymorphic Ventricular Tachycardia or Torsades de Pointes | Plaque psoriasis | Rheumatoid arthritis | 1.3 |
| Progressive multifocal leukoencephalopathy | Plaque psoriasis | Rheumatoid arthritis | 1.2 |
| Aplastic Anemia | Plaque psoriasis | Rheumatoid arthritis | 0.3 |
| Cardiovascular-related mortality | Plaque psoriasis | Rheumatoid arthritis | 0.4 |
| Encephalomyelitis | Plaque psoriasis | Rheumatoid arthritis | 0.4 |

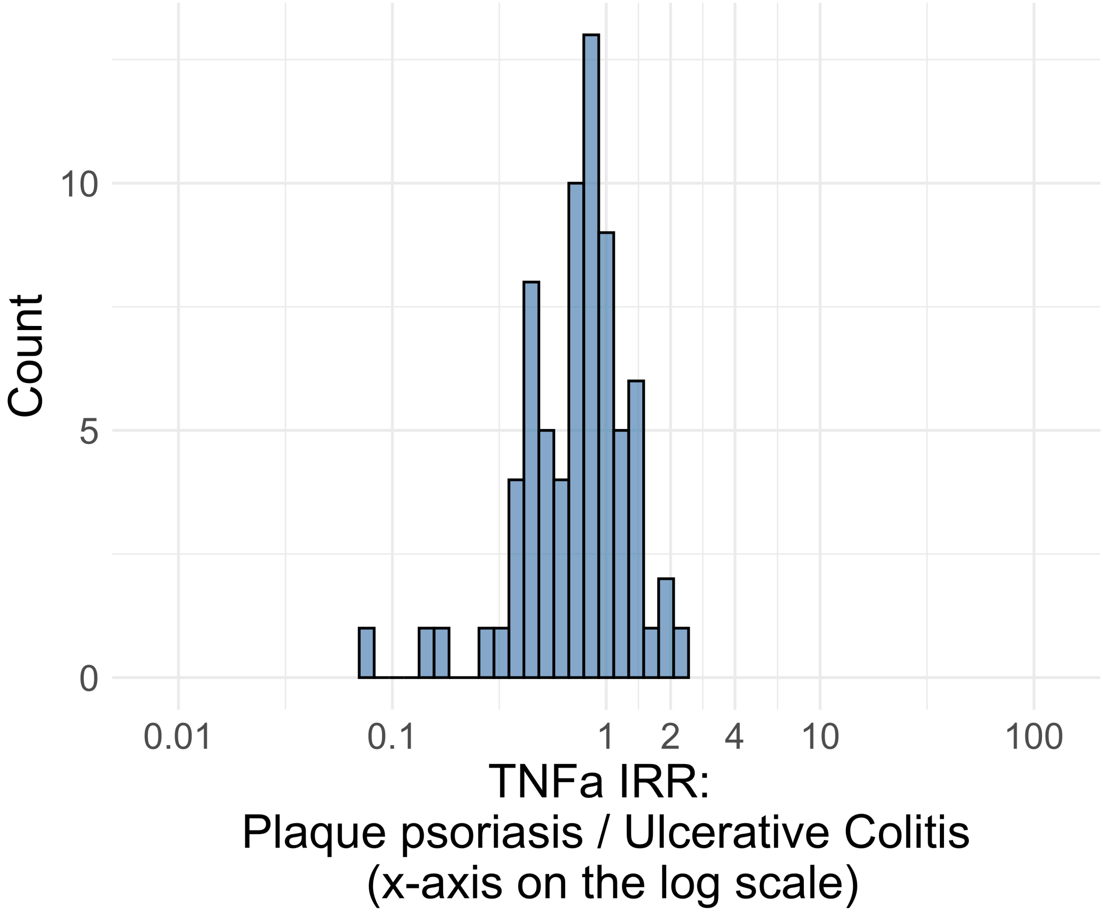


Figure S4.11. IRR distribution comparing Plaque_psoriasis and Ulcerative_Colitis in drug class: TNFa.

*Table S4.11. Most extreme IRRs for Plaque_psoriasis vs Ulcerative_Colitis (rounded to 0.001) in drug class: TNFa.*

| Outcome | Indication 1 | Indication 2 | IRR |
|---|---|---|---|
| Persons with gout | Plaque psoriasis | Ulcerative Colitis | 2.1 |
| Coronary Vessel Revascularization | Plaque psoriasis | Ulcerative Colitis | 2.0 |
| Lower extremity amputation | Plaque psoriasis | Ulcerative Colitis | 1.9 |
| Gastrointestinal bleeding events | Plaque psoriasis | Ulcerative Colitis | 0.1 |
| Aplastic Anemia | Plaque psoriasis | Ulcerative Colitis | 0.1 |
| Diarrhea events | Plaque psoriasis | Ulcerative Colitis | 0.2 |

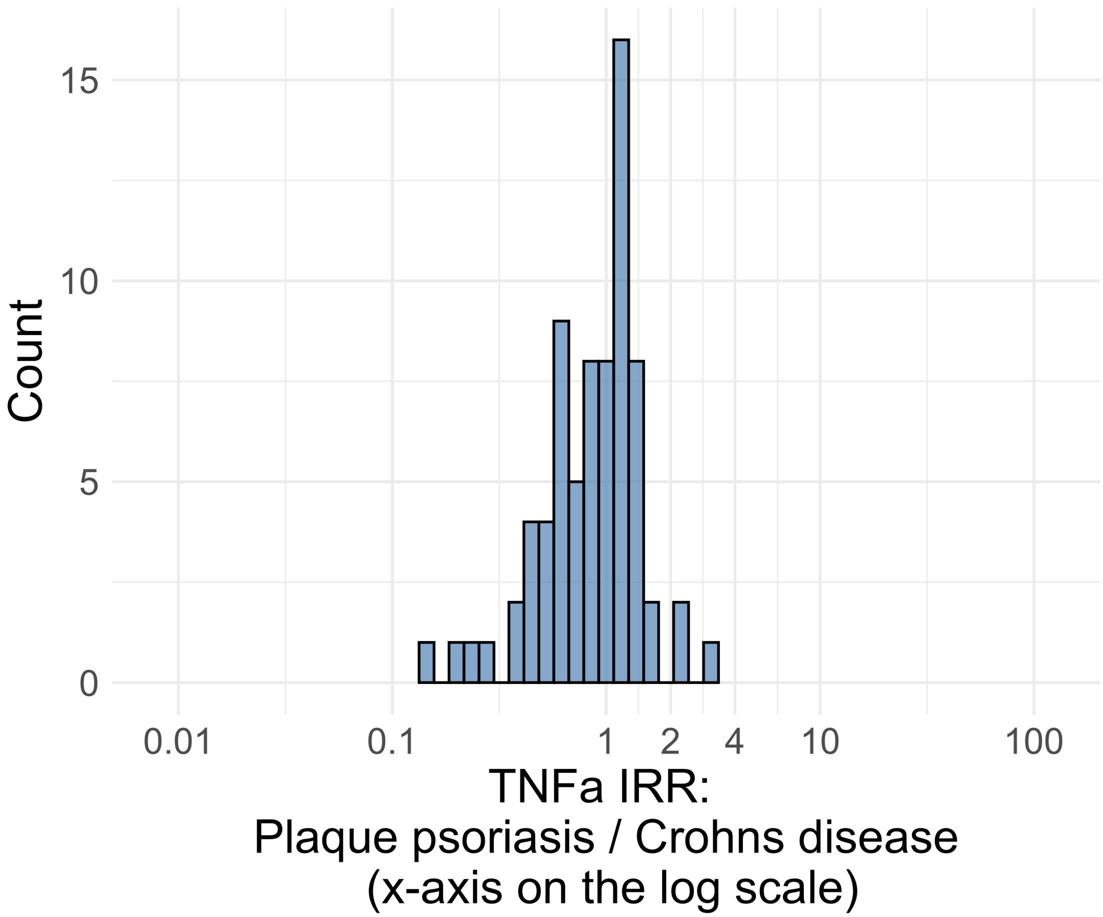


Figure S4.12. IRR distribution comparing Plaque_psoriasis and Crohns_disease in drug class: TNFa.

*Table S4.12. Most extreme IRRs for Plaque_psoriasis vs Crohns_disease (rounded to 0.001) in drug class: TNFa.*

| Outcome | Indication 1 | Indication 2 | IRR |
|---|---|---|---|
| Lower extremity amputation | Plaque psoriasis | Crohns disease | 2.9 |
| Coronary Vessel Revascularization | Plaque psoriasis | Crohns disease | 2.4 |
| Persons with gout | Plaque psoriasis | Crohns disease | 2.1 |
| Gastrointestinal bleeding events | Plaque psoriasis | Crohns disease | 0.1 |
| Aplastic Anemia | Plaque psoriasis | Crohns disease | 0.2 |
| Diarrhea events | Plaque psoriasis | Crohns disease | 0.2 |

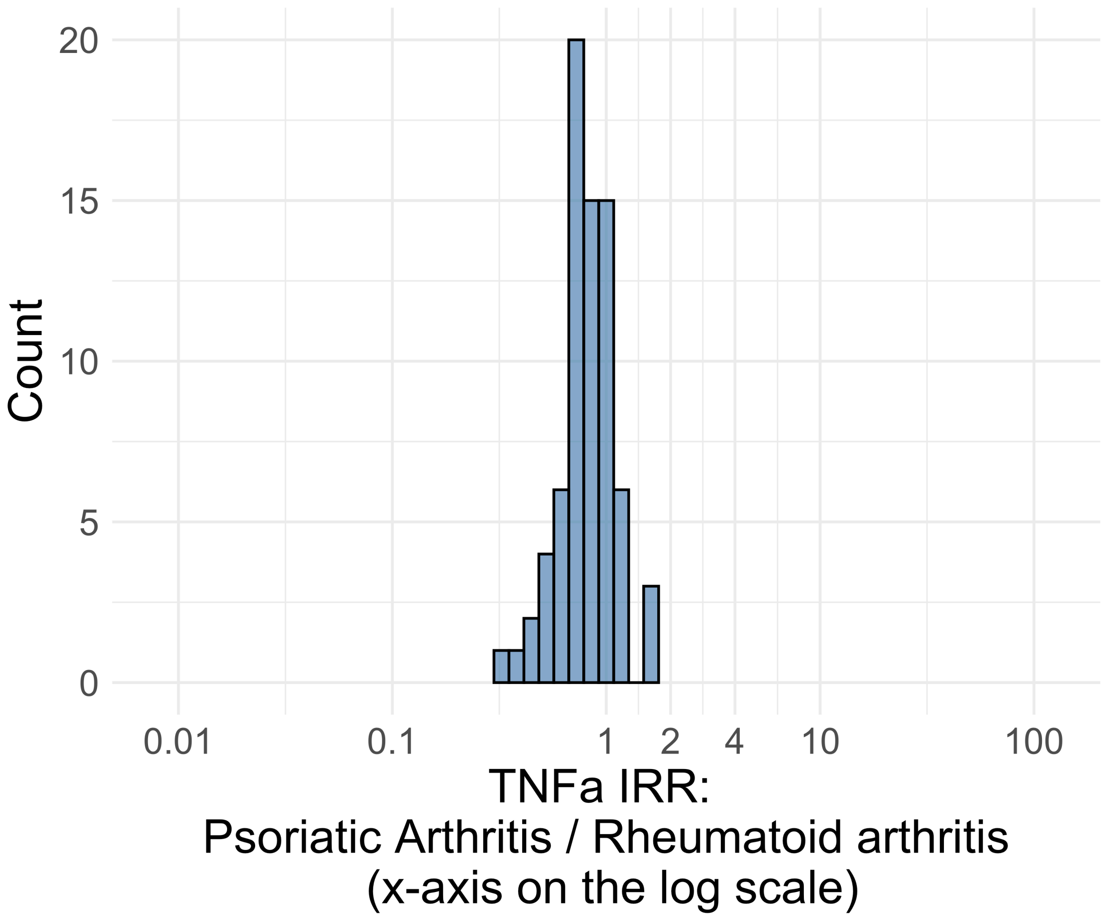


Figure S4.13. IRR distribution comparing Psoriatic_Arthritis and Rheumatoid_arthritis in drug class: TNFa.

*Table S4.13. Most extreme IRRs for Psoriatic_Arthritis vs Rheumatoid_arthritis (rounded to 0.001) in drug class: TNFa.*

| Outcome | Indication 1 | Indication 2 | IRR |
|---|---|---|---|
| Polymorphic Ventricular Tachycardia or Torsades de Pointes | Psoriatic Arthritis | Rheumatoid arthritis | 1.7 |
| Progressive multifocal leukoencephalopathy | Psoriatic Arthritis | Rheumatoid arthritis | 1.7 |
| All events Drug Rash with Eosinophilia and Systemic Symptoms (DRESS) | Psoriatic Arthritis | Rheumatoid arthritis | 1.6 |
| Cardiovascular-related mortality | Psoriatic Arthritis | Rheumatoid arthritis | 0.3 |
| Aplastic Anemia | Psoriatic Arthritis | Rheumatoid arthritis | 0.4 |
| Disseminated Intravascular Coagulation | Psoriatic Arthritis | Rheumatoid arthritis | 0.5 |

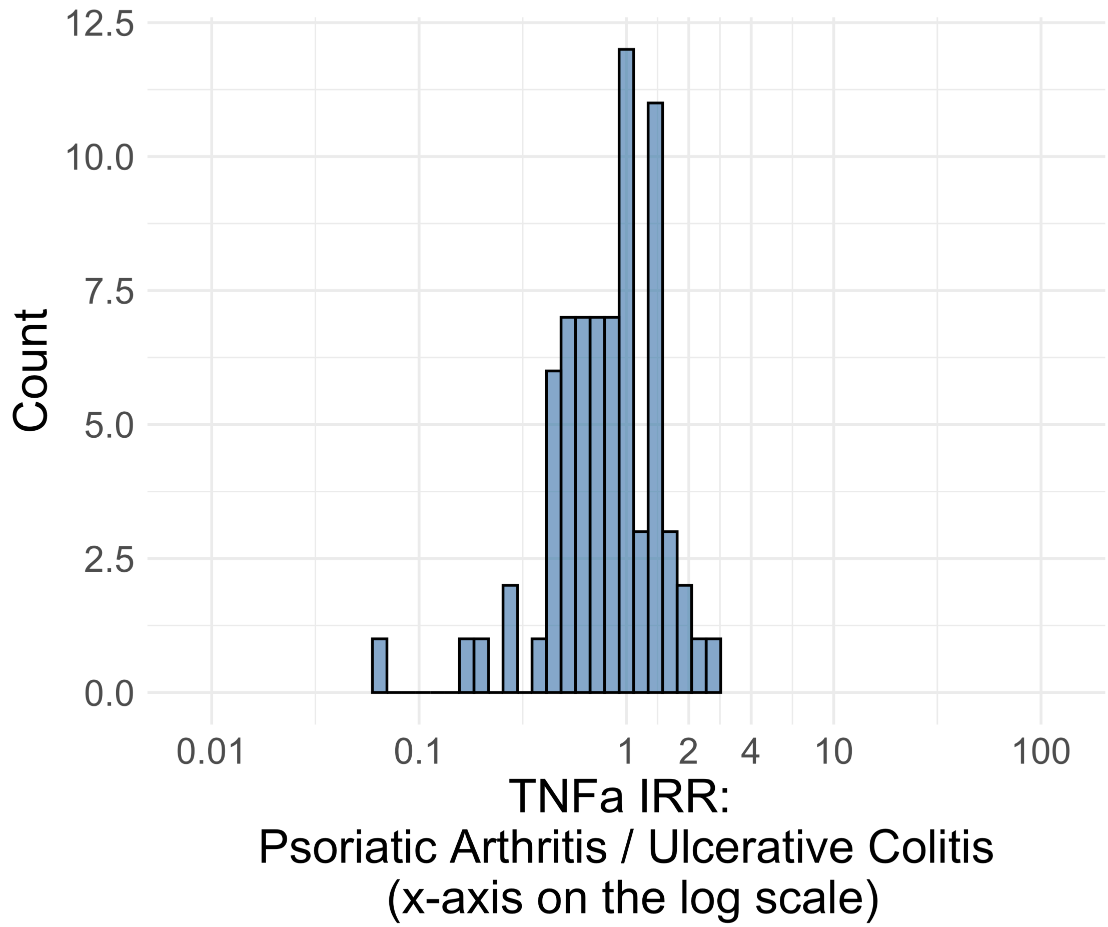


Figure S4.14. IRR distribution comparing Psoriatic_Arthritis and Ulcerative_Colitis in drug class: TNFa.

*Table S4.14. Most extreme IRRs for Psoriatic_Arthritis vs Ulcerative_Colitis (rounded to 0.001) in drug class: TNFa.*

| Outcome | Indication 1 | Indication 2 | IRR |
|---|---|---|---|
| Persons with gout | Psoriatic Arthritis | Ulcerative Colitis | 2.7 |
| Coronary Vessel Revascularization | Psoriatic Arthritis | Ulcerative Colitis | 2.1 |
| Lower extremity amputation | Psoriatic Arthritis | Ulcerative Colitis | 1.9 |
| Gastrointestinal bleeding events | Psoriatic Arthritis | Ulcerative Colitis | 0.1 |
| Aplastic Anemia | Psoriatic Arthritis | Ulcerative Colitis | 0.2 |
| Diarrhea events | Psoriatic Arthritis | Ulcerative Colitis | 0.2 |

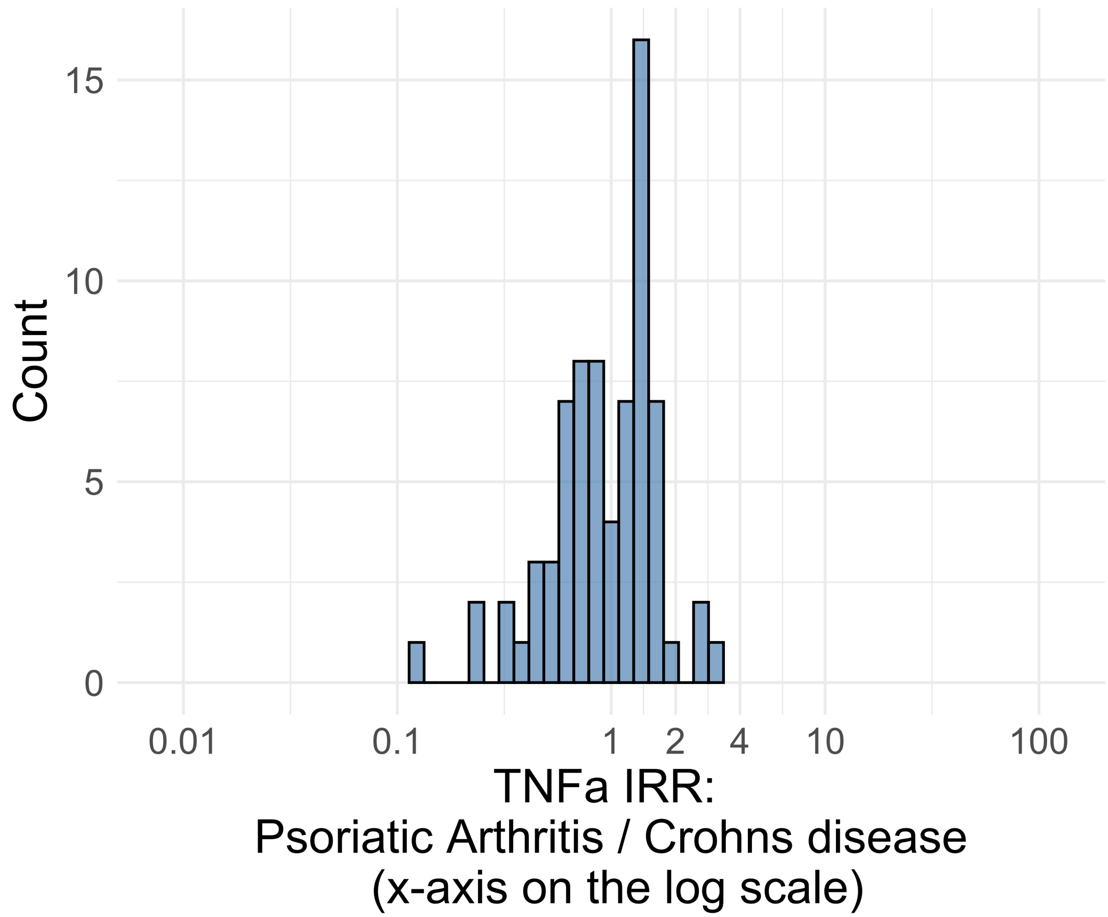


Figure S4.15. IRR distribution comparing Psoriatic_Arthritis and Crohns_disease in drug class: TNFa.

*Table S4.15. Most extreme IRRs for Psoriatic_Arthritis vs Crohns_disease (rounded to 0.001) in drug class: TNFa.*

| Outcome | Indication 1 | Indication 2 | IRR |
|---|---|---|---|
| Lower extremity amputation | Psoriatic Arthritis | Crohns disease | 2.9 |
| Coronary Vessel Revascularization | Psoriatic Arthritis | Crohns disease | 2.6 |
| Persons with gout | Psoriatic Arthritis | Crohns disease | 2.6 |
| Gastrointestinal bleeding events | Psoriatic Arthritis | Crohns disease | 0.1 |
| Aplastic Anemia | Psoriatic Arthritis | Crohns disease | 0.2 |
| Diarrhea events | Psoriatic Arthritis | Crohns disease | 0.3 |

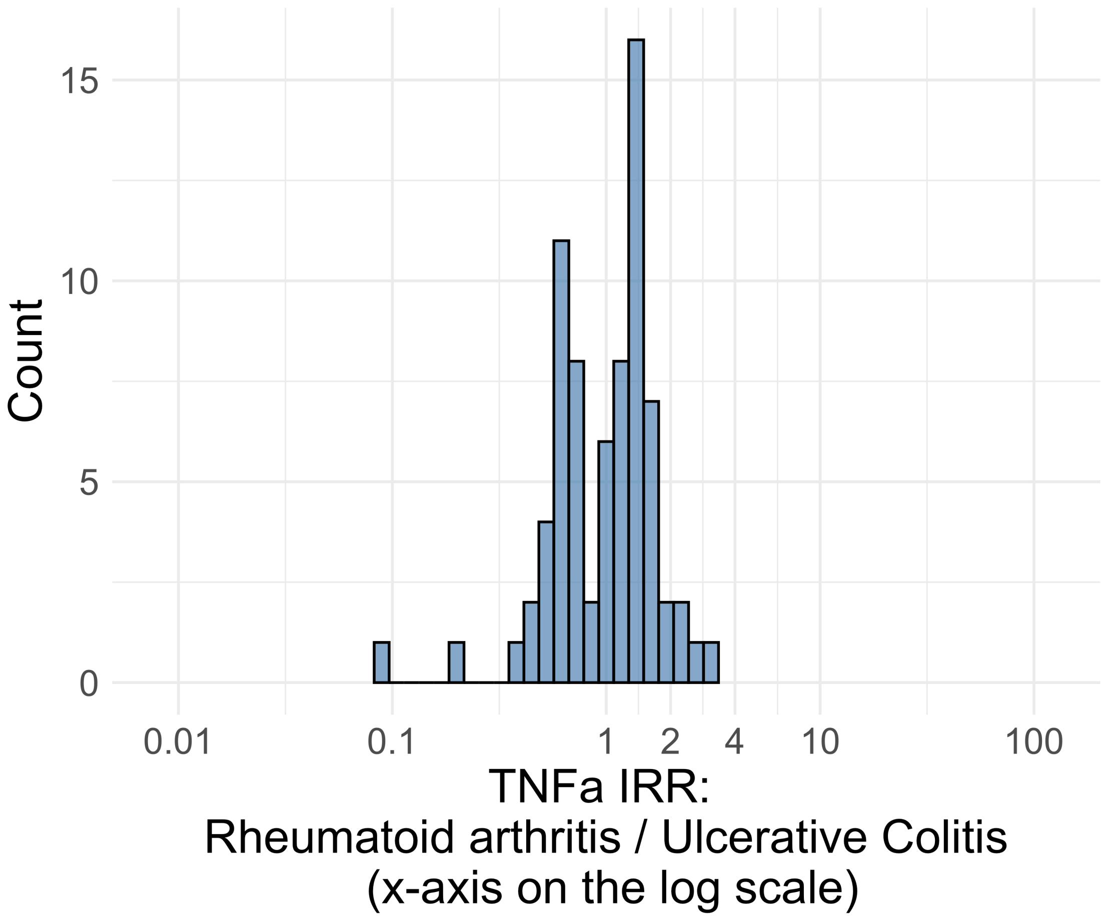


Figure S4.16. IRR distribution comparing Rheumatoid_arthritis and Ulcerative_Colitis in drug class: TNFa.

*Table S4.16. Most extreme IRRs for Rheumatoid_arthritis vs Ulcerative_Colitis (rounded to 0.001) in drug class: TNFa.*

| Outcome | Indication 1 | Indication 2 | IRR |
|---|---|---|---|
| Lower extremity amputation | Rheumatoid arthritis | Ulcerative Colitis | 3.2 |
| Rhabdomyolysis | Rheumatoid arthritis | Ulcerative Colitis | 2.8 |
| Persons with gout | Rheumatoid arthritis | Ulcerative Colitis | 2.4 |
| Gastrointestinal bleeding events | Rheumatoid arthritis | Ulcerative Colitis | 0.1 |
| Diarrhea events | Rheumatoid arthritis | Ulcerative Colitis | 0.2 |
| Abnormal weight loss events | Rheumatoid arthritis | Ulcerative Colitis | 0.4 |

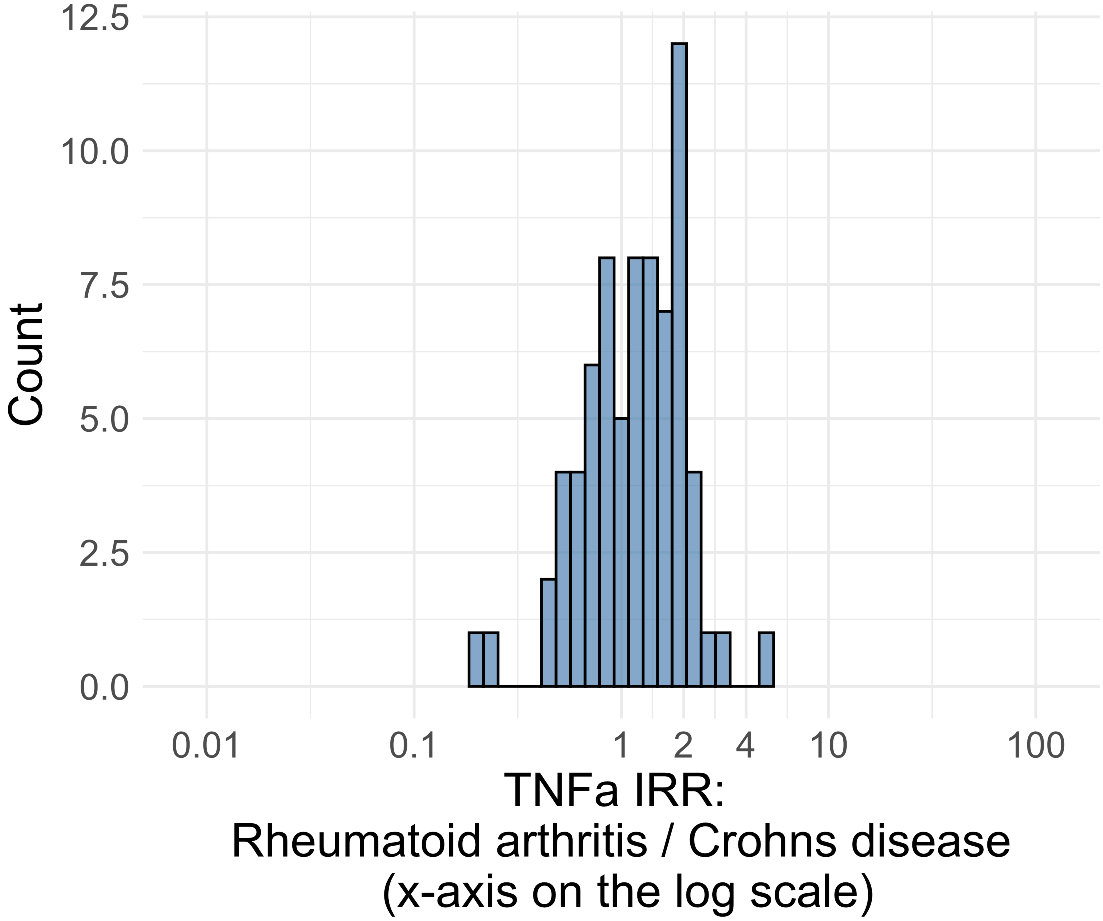


Figure S4.17. IRR distribution comparing Rheumatoid_arthritis and Crohns_disease in drug class: TNFa.

*Table S4.17. Most extreme IRRs for Rheumatoid_arthritis vs Crohns_disease (rounded to 0.001) in drug class: TNFa.*

| Outcome | Indication 1 | Indication 2 | IRR |
|---|---|---|---|
| Lower extremity amputation | Rheumatoid arthritis | Crohns disease | 5.0 |
| Rhabdomyolysis | Rheumatoid arthritis | Crohns disease | 2.9 |
| Coronary Vessel Revascularization | Rheumatoid arthritis | Crohns disease | 2.8 |
| Gastrointestinal bleeding events | Rheumatoid arthritis | Crohns disease | 0.2 |
| Diarrhea events | Rheumatoid arthritis | Crohns disease | 0.2 |
| Abnormal weight loss events | Rheumatoid arthritis | Crohns disease | 0.4 |

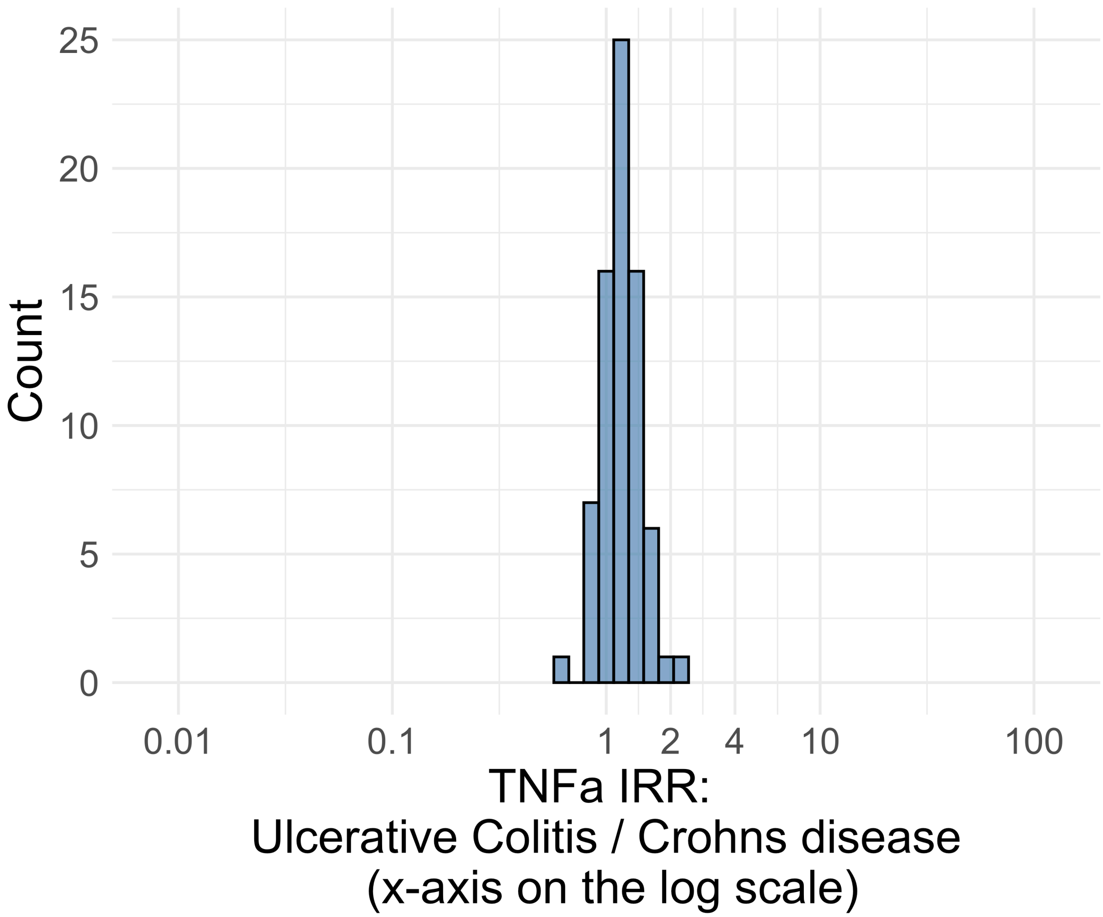


Figure S4.18. IRR distribution comparing Ulcerative_Colitis and Crohns_disease in drug class: TNFa.

*Table S4.18. Most extreme IRRs for Ulcerative_Colitis vs Crohns_disease (rounded to 0.001) in drug class: TNFa.*

| Outcome | Indication 1 | Indication 2 | IRR |
|---|---|---|---|
| Encephalomyelitis | Ulcerative Colitis | Crohns disease | 2.4 |
| Gastrointestinal bleeding events | Ulcerative Colitis | Crohns disease | 2.1 |
| Thrombosis with Thrombocytopenia (TWT) | Ulcerative Colitis | Crohns disease | 1.7 |
| Stevens-Johnson syndrome, toxic epidermal necrolysis spectrum | Ulcerative Colitis | Crohns disease | 0.6 |
| All events of Severe Cutaneous Adverse Reaction (SCAR = SJS+TEN+DRESS) | Ulcerative Colitis | Crohns disease | 0.8 |
| Anaphylaxis | Ulcerative Colitis | Crohns disease | 0.9 |

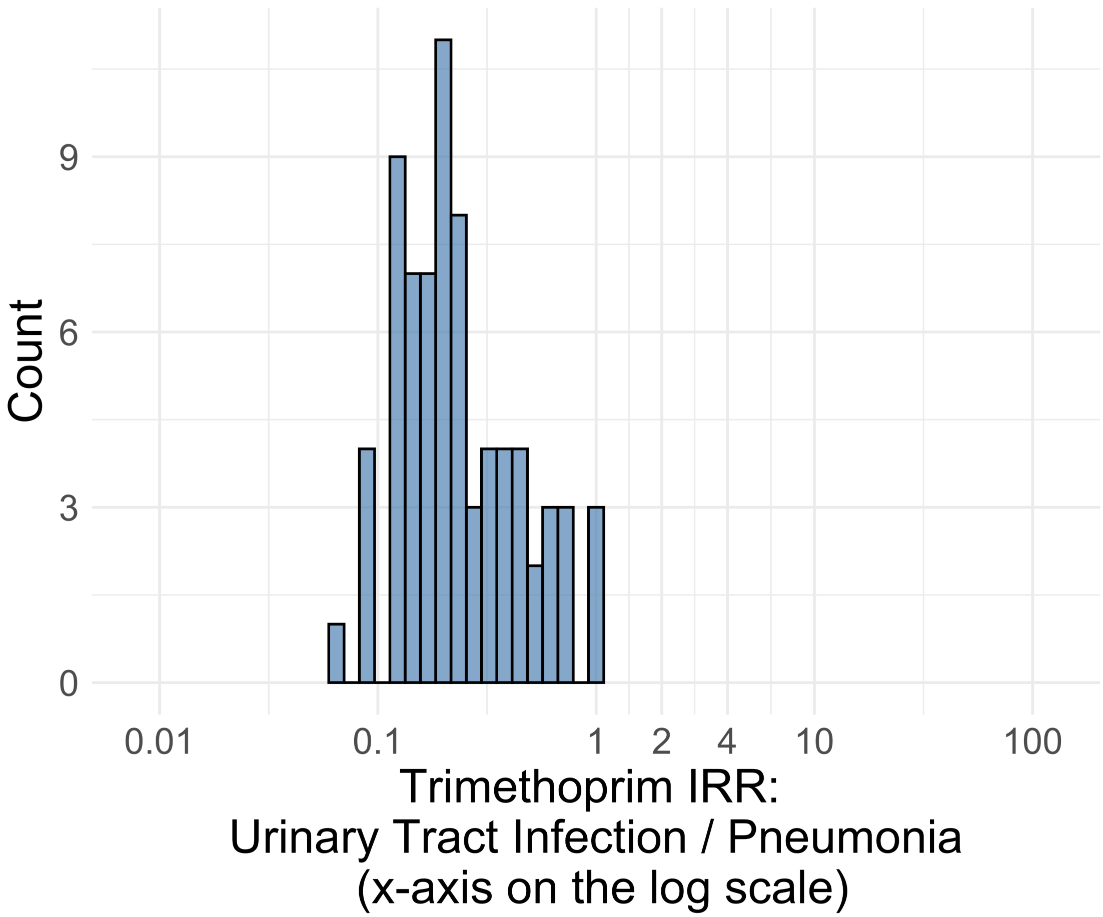


Figure S4.19. IRR distribution comparing Urinary_Tract_Infection and Pneumonia in drug class: Trimethoprim.

*Table S4.19. Most extreme IRRs for Urinary_Tract_Infection vs Pneumonia (rounded to 0.001) in drug class: Trimethoprim.*

| Outcome | Indication 1 | Indication 2 | IRR |
|---|---|---|---|
| Persons with vertigo | Urinary Tract Infection | Pneumonia | 1.1 |
| Narcolepsy | Urinary Tract Infection | Pneumonia | 1.0 |
| Bladder Cancer | Urinary Tract Infection | Pneumonia | 1.0 |
| Febrile Neutropenia or Neutropenic Fever | Urinary Tract Infection | Pneumonia | 0.1 |
| Hospitalization with heart failure events | Urinary Tract Infection | Pneumonia | 0.1 |
| Cardiovascular-related mortality | Urinary Tract Infection | Pneumonia | 0.1 |
