## Supplement S5 for "The Role Of Drug Indication On Incidence Rate Heterogeneity: A Large-Scale, Systematic Evaluation Across An International Network Of Observational Databases"

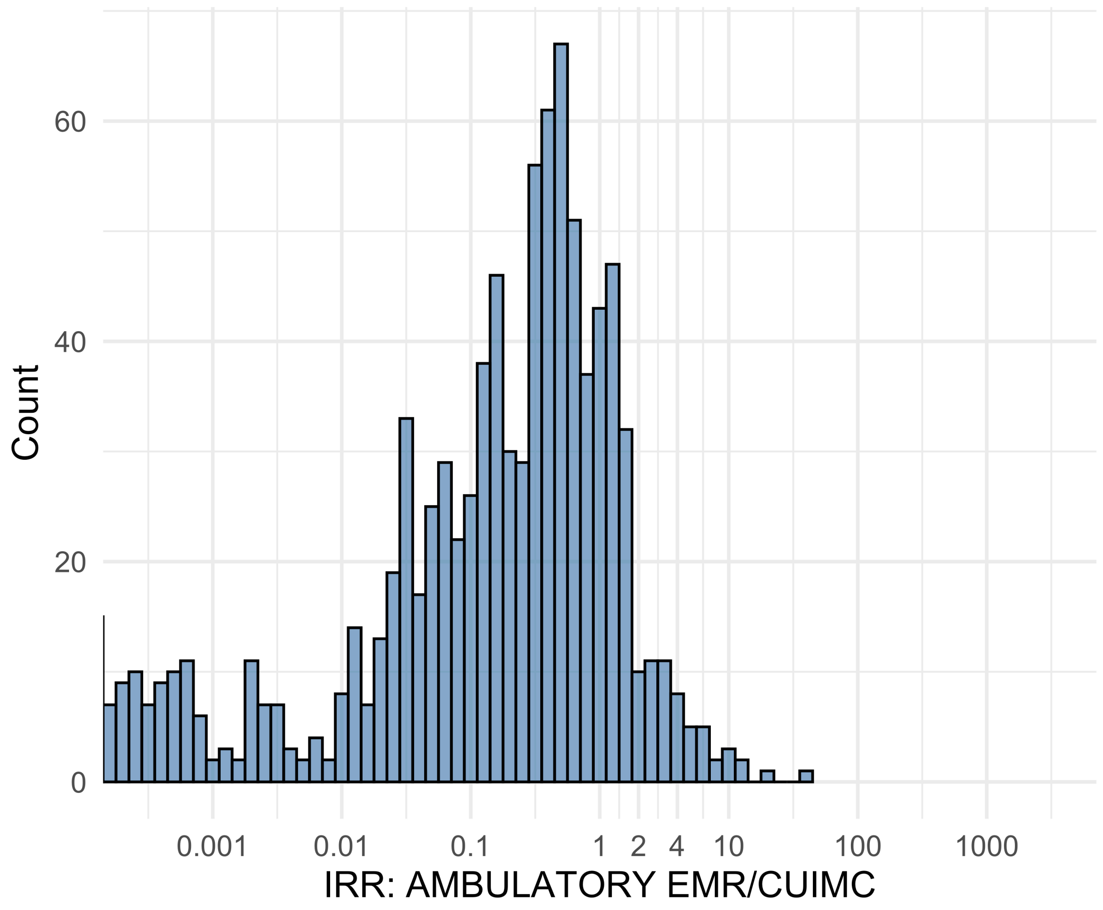


Figure S5.01. IRR distribution comparing AMBULATORY EMR and CUIMC.

*Table S5.01. Most extreme IRRs for AMBULATORY EMR vs CUIMC (rounded to 0.001).*

| source1 | source2 | Drug and Indication | Outcome | IRR |
|---|---|---|---|---|
| AMBULATORY EMR | CUIMC | New users of SGLT2 inhibitor nested in Left Heart Failure | Anaphylaxis | 35.9 |
| AMBULATORY EMR | CUIMC | New users of Tumor Necrosis Factor alpha (TNFa) inhibitors nested in Crohns disease | Anaphylaxis | 17.9 |
| AMBULATORY EMR | CUIMC | New users of SGLT2 inhibitor nested in Type 2 diabetes mellitus | Anaphylaxis | 12.5 |
| AMBULATORY EMR | CUIMC | New users of Beta blockers nested in essential hypertension | 4-point MACE | <0.001 |
| AMBULATORY EMR | CUIMC | New users of Beta blockers nested in essential hypertension | Total cardiovascular disease events (ischemic stroke, hemorrhagic stroke, heart failure, acute myocardial infarction or sudden cardiac death) | <0.001 |

| source1 | source2 | Drug and Indication | Outcome | IRR |
|---|---|---|---|---|
| AMBULATORY EMR | CUIMC | New users of Beta blockers nested in essential hypertension | Hospitalization with heart failure events | <0.001 |

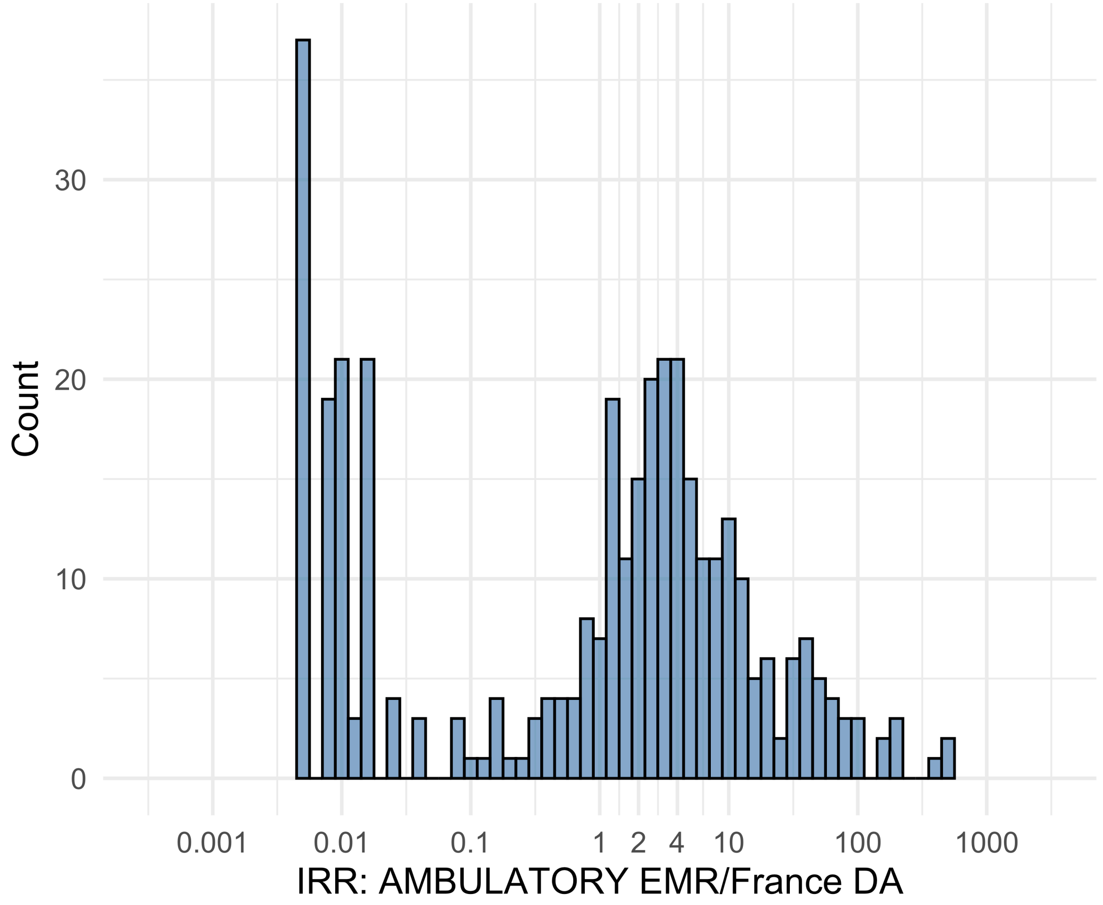


Figure S5.02. IRR distribution comparing AMBULATORY EMR and France DA.

*Table S5.02. Most extreme IRRs for AMBULATORY EMR vs France DA (rounded to 0.001).*

| source1 | source2 | Drug and Indication | Outcome | IRR |
|---|---|---|---|---|
| AMBULATORY EMR | France DA | New users of Fluoroquinolone systemic nested in Urinary Tract Infection | Diarrhea events | 496.9 |
| AMBULATORY EMR | France DA | New users of Trimethoprim systemetic nested in Urinary Tract Infection | Diarrhea events | 476.8 |
| AMBULATORY EMR | France DA | New users of Beta blockers nested in essential hypertension | Diarrhea events | 394.4 |
| AMBULATORY EMR | France DA | New users of Trimethoprim systemetic nested in Urinary Tract Infection | Febrile Neutropenia or Neutropenic Fever | 0.0 |
| AMBULATORY EMR | France DA | New users of Trimethoprim systemetic nested in Urinary Tract Infection | Rhabdomyolysis | 0.0 |
| AMBULATORY EMR | France DA | New users of Trimethoprim systemetic nested in Urinary Tract Infection | Sudden Cardiac arrest or cardiac death | 0.0 |

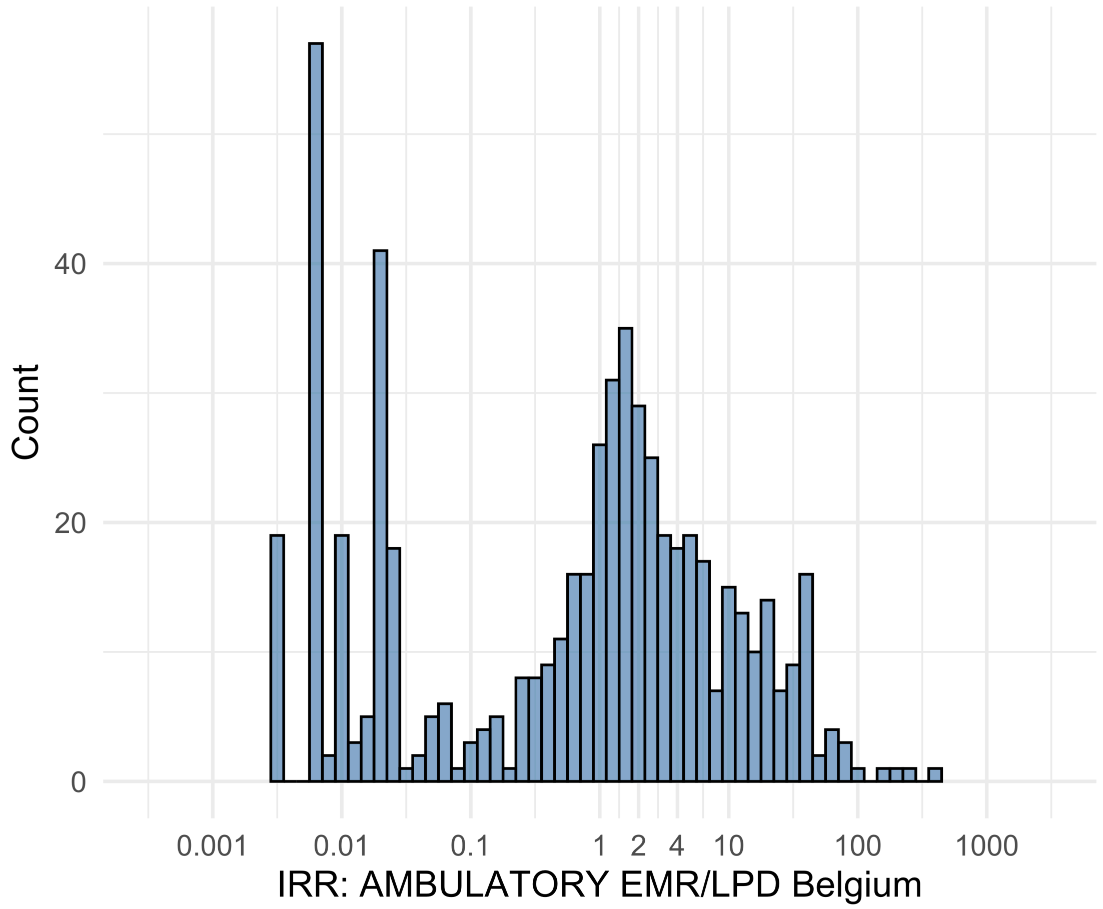


Figure S5.03. IRR distribution comparing AMBULATORY EMR and LPD Belgium.

*Table S5.03. Most extreme IRRs for AMBULATORY EMR vs LPD Belgium (rounded to 0.001).*

| source1 | source2 | Drug and Indication | Outcome | IRR |
|---|---|---|---|---|
| AMBULATORY EMR | LPD Belgium | New users of Beta blockers nested in essential hypertension | Deep Vein Thrombosis (DVT) | 358.0 |
| AMBULATORY EMR | LPD Belgium | New users of Beta blockers nested in essential hypertension | Anaphylaxis | 277.4 |
| AMBULATORY EMR | LPD Belgium | New users of Fluoroquinolone systemic nested in Urinary Tract Infection | Anaphylaxis | 204.4 |
| AMBULATORY EMR | LPD Belgium | New users of GLP-1 receptor antagonists nested in obesity | Febrile Neutropenia or Neutropenic Fever | 0.0 |
| AMBULATORY EMR | LPD Belgium | New users of GLP-1 receptor antagonists nested in obesity | Rhabdomyolysis | 0.0 |
| AMBULATORY EMR | LPD Belgium | New users of GLP-1 receptor antagonists nested in obesity | Sudden Cardiac arrest or cardiac death | 0.0 |

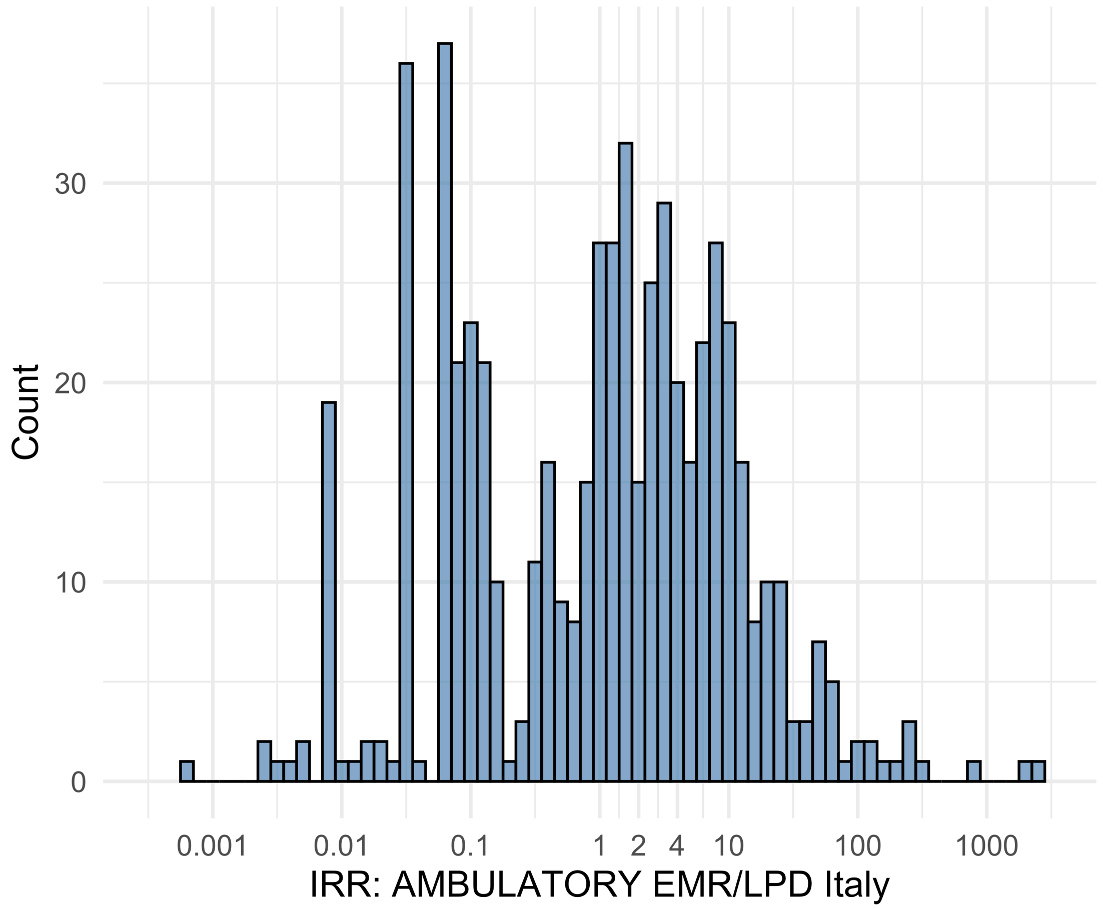


Figure S5.04. IRR distribution comparing AMBULATORY EMR and LPD Italy.

*Table S5.04. Most extreme IRRs for AMBULATORY EMR vs LPD Italy (rounded to 0.001).*

| source1 | source2 | Drug and Indication | Outcome | IRR |
|---|---|---|---|---|
| AMBULATORY EMR | LPD Italy | New users of Fluoroquinolone systemic nested in Urinary Tract Infection | Fall events | 2413.8 |
| AMBULATORY EMR | LPD Italy | New users of Beta blockers nested in essential hypertension | Fall events | 2215.4 |
| AMBULATORY EMR | LPD Italy | New users of Cephalosporin systemetic nested in Urinary Tract Infection | Fall events | 869.3 |
| AMBULATORY EMR | LPD Italy | New users of Beta blockers nested in essential hypertension | Cardiovascular-related mortality | 0.0 |
| AMBULATORY EMR | LPD Italy | New users of Cephalosporin systemetic nested in Urinary Tract Infection | Cardiovascular-related mortality | 0.0 |
| AMBULATORY EMR | LPD Italy | New users of Beta blockers nested in essential hypertension | Progressive multifocal leukoencephalopathy | 0.0 |

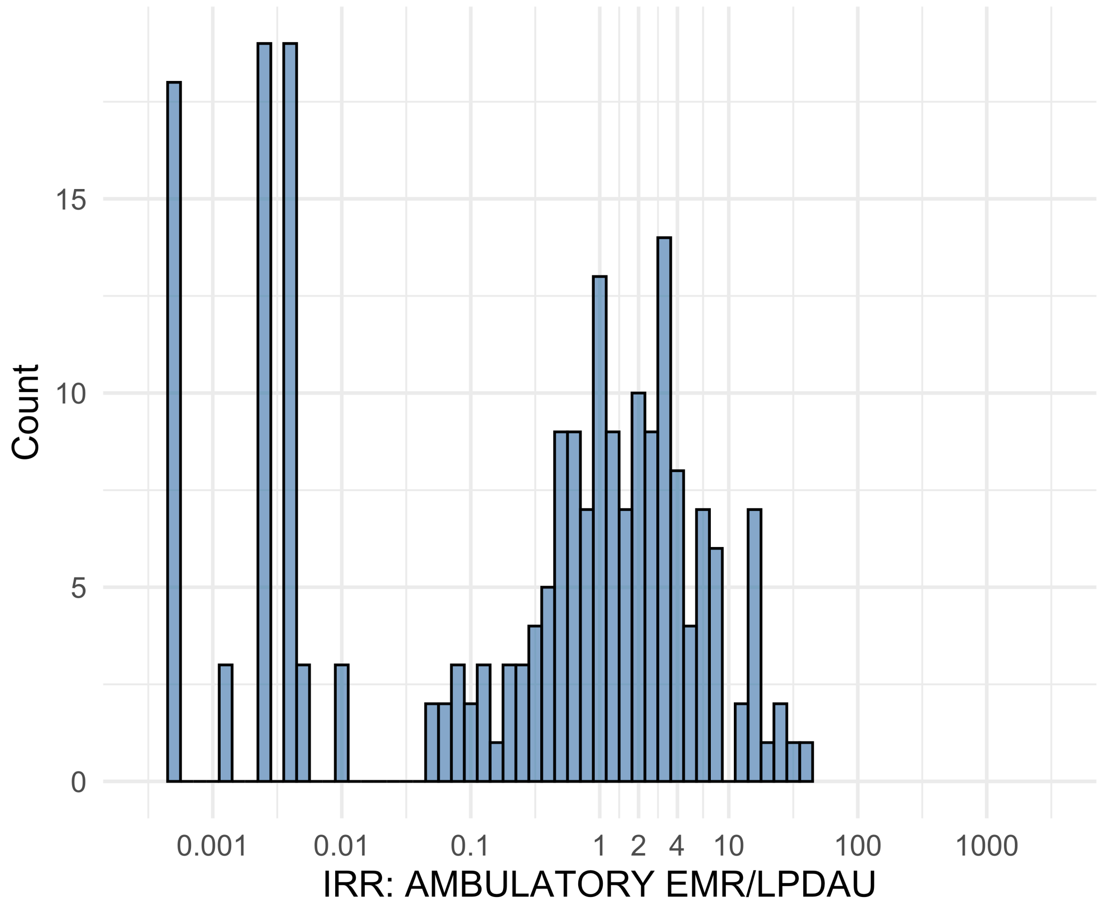


Figure S5.05. IRR distribution comparing AMBULATORY EMR and LPDAU.

*Table S5.05. Most extreme IRRs for AMBULATORY EMR vs LPDAU (rounded to 0.001).*

| source1 | source2 | Drug and Indication | Outcome | IRR |
|---|---|---|---|---|
| AMBULATORY EMR | LPDAU | New users of Cephalosporin systemetic nested in Urinary Tract Infection | Hypokalemia events | 36.5 |
| AMBULATORY EMR | LPDAU | New users of Cephalosporin systemetic nested in Urinary Tract Infection | Acute Kidney Injury AKI | 32.3 |
| AMBULATORY EMR | LPDAU | New users of Trimethoprim systemetic nested in Urinary Tract Infection | Abnormal weight loss events | 27.4 |
| AMBULATORY EMR | LPDAU | New users of Beta blockers nested in essential hypertension | Febrile Neutropenia or Neutropenic Fever | 0.0 |
| AMBULATORY EMR | LPDAU | New users of Beta blockers nested in essential hypertension | Rhabdomyolysis | 0.0 |
| AMBULATORY EMR | LPDAU | New users of Beta blockers nested in essential hypertension | Sudden Cardiac arrest or cardiac death | 0.0 |

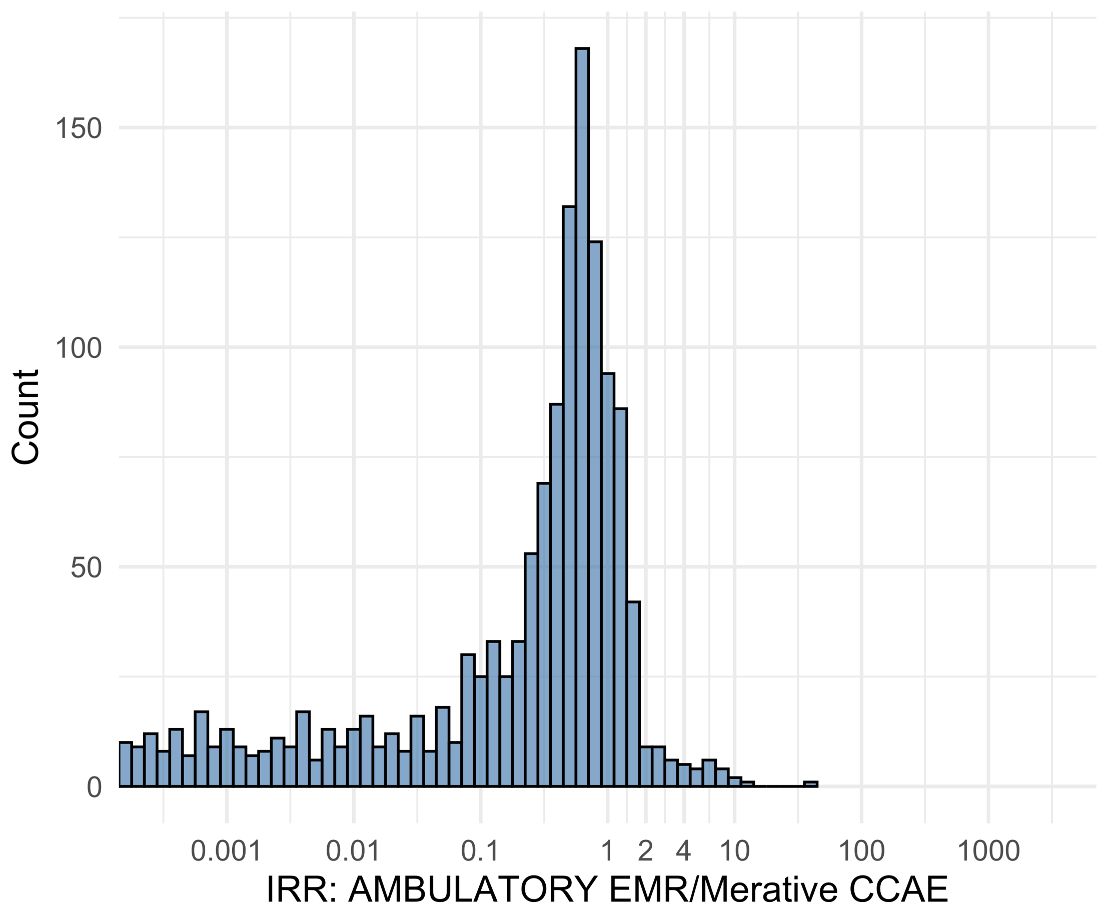


Figure S5.06. IRR distribution comparing AMBULATORY EMR and Merative CCAE.

*Table S5.06. Most extreme IRRs for AMBULATORY EMR vs Merative CCAE (rounded to 0.001).*

| source1 | source2 | Drug and Indication | Outcome | IRR |
|---|---|---|---|---|
| AMBULATORY EMR | Merative CCAE | New users of Beta blockers nested in essential hypertension | Polymorphic Ventricular Tachycardia or Torsades de Pointes | 38.2 |
| AMBULATORY EMR | Merative CCAE | New users of SGLT2 inhibitor nested in Left Heart Failure | Anaphylaxis | 11.8 |
| AMBULATORY EMR | Merative CCAE | New users of Beta blockers nested in Left Heart Failure | Polymorphic Ventricular Tachycardia or Torsades de Pointes | 10.5 |
| AMBULATORY EMR | Merative CCAE | New users of Beta blockers nested in essential hypertension | Hospitalization with heart failure events | <0.001 |
| AMBULATORY EMR | Merative CCAE | New users of Beta blockers nested in essential hypertension | 4-point MACE | <0.001 |
| AMBULATORY EMR | Merative CCAE | New users of Beta blockers nested in essential hypertension | Total cardiovascular disease events (ischemic stroke, hemorrhagic stroke, heart failure, acute myocardial infarction or sudden cardiac death) | <0.001 |

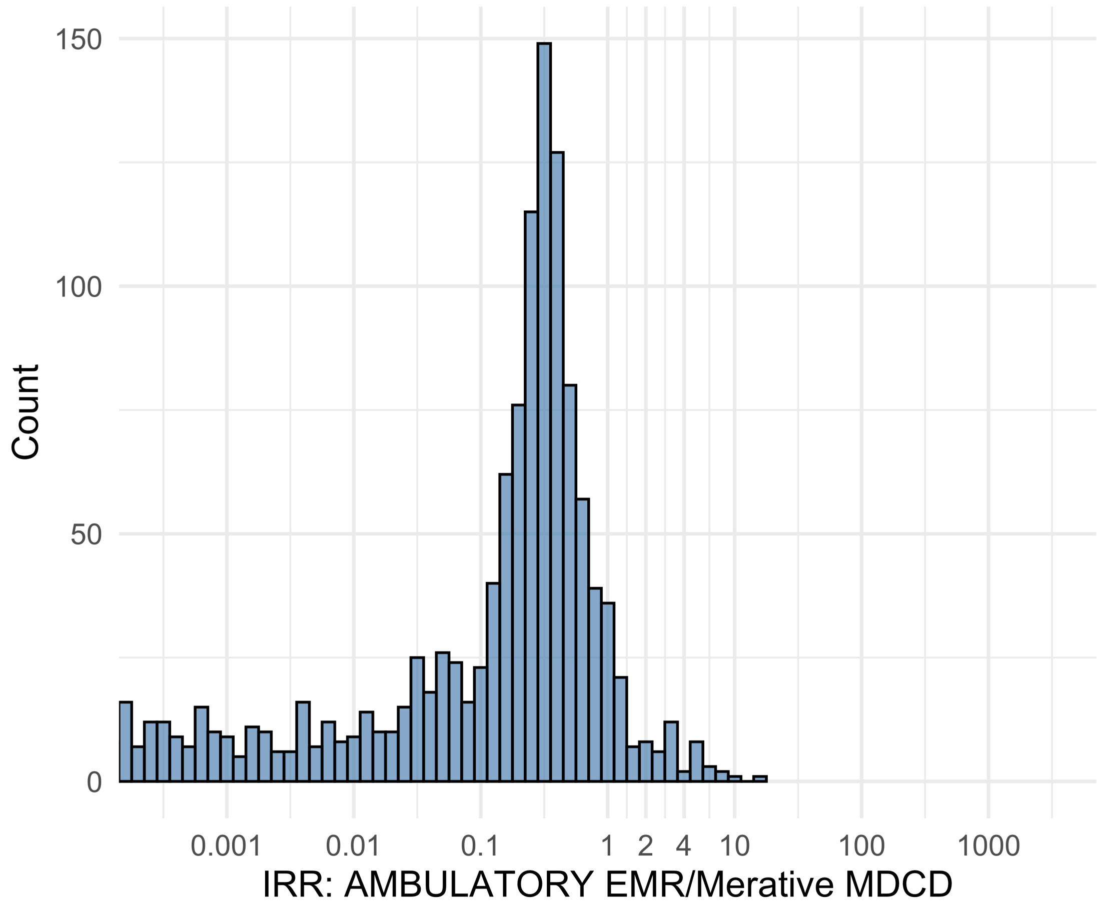


Figure S5.07. IRR distribution comparing AMBULATORY EMR and Merative MDCD.

*Table S5.07. Most extreme IRRs for AMBULATORY EMR vs Merative MDCD (rounded to 0.001).*

| source1 | source2 | Drug and Indication | Outcome | IRR |
|---|---|---|---|---|
| AMBULATORY EMR | Merative MDCD | New users of JAK inhibitors nested in Rheumatoid arthritis | Anaphylaxis | 16.9 |
| AMBULATORY EMR | Merative MDCD | New users of Beta blockers nested in Left Heart Failure | Polymorphic Ventricular Tachycardia or Torsades de Pointes | 9.0 |
| AMBULATORY EMR | Merative MDCD | New users of Trimethoprim systemetic nested in Acute Typical Pneumonia | Anaphylaxis | 8.5 |
| AMBULATORY EMR | Merative MDCD | New users of Beta blockers nested in essential hypertension | Hospitalization with heart failure events | <0.001 |
| AMBULATORY EMR | Merative MDCD | New users of Beta blockers nested in essential hypertension | 4-point MACE | <0.001 |
| AMBULATORY EMR | Merative MDCD | New users of Beta blockers nested in essential hypertension | Total cardiovascular disease events (ischemic stroke, hemorrhagic stroke, heart failure, acute myocardial infarction or sudden cardiac death) | <0.001 |

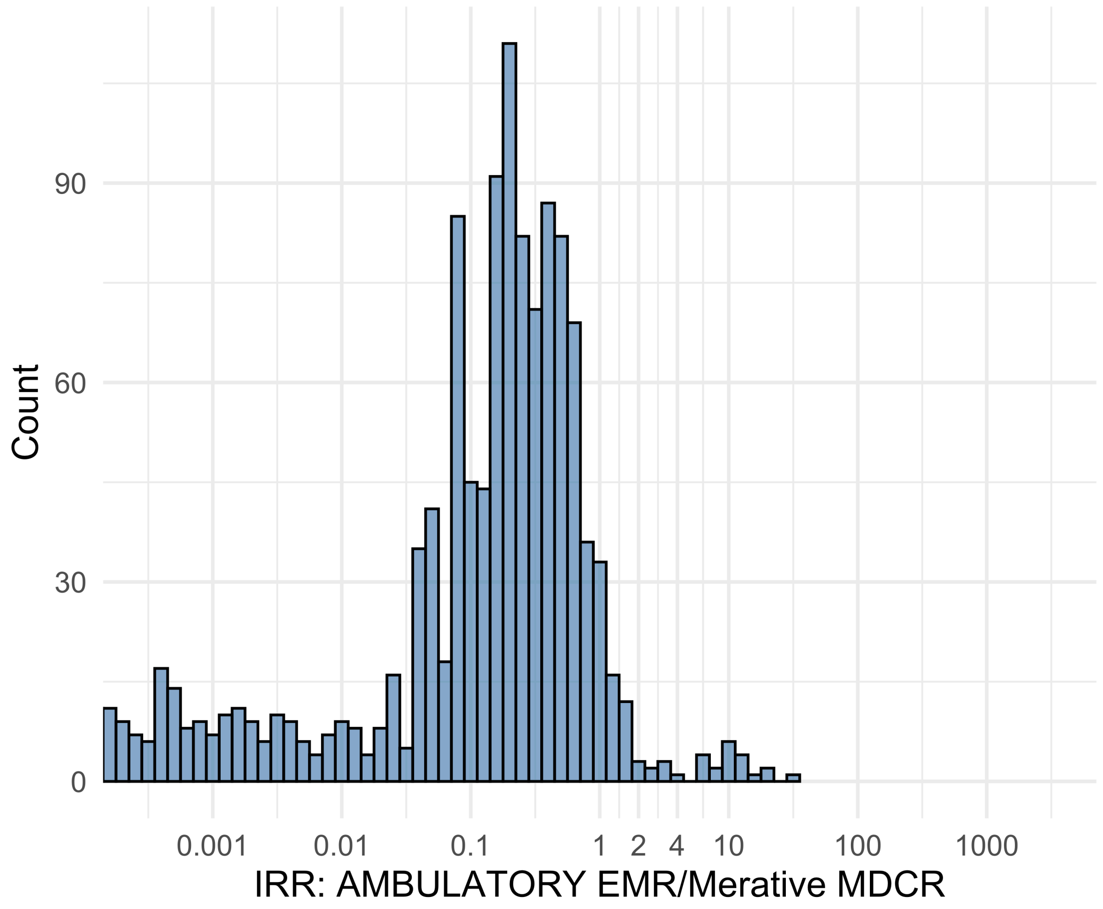


Figure S5.08. IRR distribution comparing AMBULATORY EMR and Merative MDCR.

*Table S5.08. Most extreme IRRs for AMBULATORY EMR vs Merative MDCR (rounded to 0.001).*

| source1 | source2 | Drug and Indication | Outcome | IRR |
|---|---|---|---|---|
| AMBULATORY EMR | Merative MDCR | New users of Tumor Necrosis Factor alpha (TNFa) inhibitors nested in Rheumatoid arthritis | Anaphylaxis | 33.8 |
| AMBULATORY EMR | Merative MDCR | New users of Beta blockers nested in Left Heart Failure | Polymorphic Ventricular Tachycardia or Torsades de Pointes | 21.3 |
| AMBULATORY EMR | Merative MDCR | New users of Trimethoprim systemetic nested in Acute Typical Pneumonia | Anaphylaxis | 19.1 |
| AMBULATORY EMR | Merative MDCR | New users of Beta blockers nested in essential hypertension | Hospitalization with heart failure events | <0.001 |
| AMBULATORY EMR | Merative MDCR | New users of Beta blockers nested in essential hypertension | 4-point MACE | <0.001 |
| AMBULATORY EMR | Merative MDCR | New users of Beta blockers nested in essential hypertension | Total cardiovascular disease events (ischemic stroke, hemorrhagic stroke, heart failure, acute myocardial infarction or sudden cardiac death) | <0.001 |

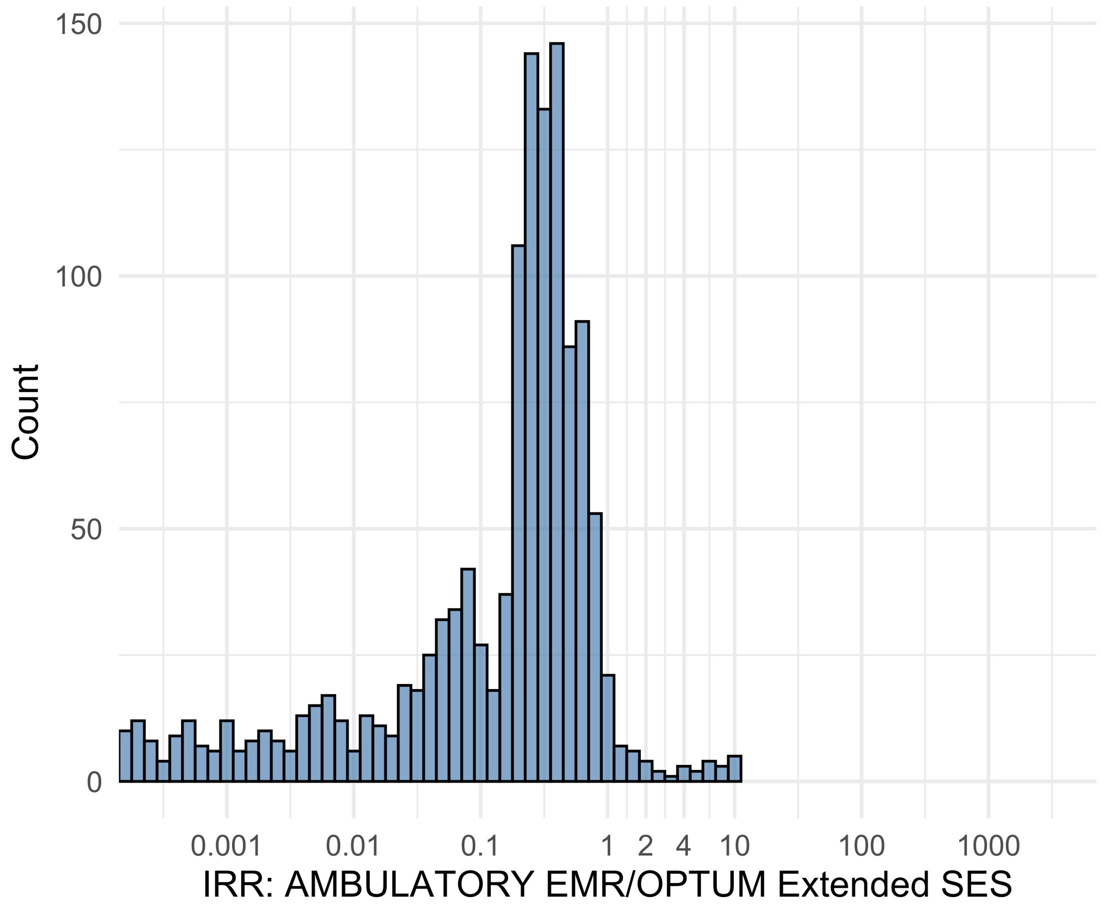


Figure S5.09. IRR distribution comparing AMBULATORY EMR and OPTUM Extended SES.

*Table S5.09. Most extreme IRRs for AMBULATORY EMR vs OPTUM Extended SES (rounded to 0.001).*

| source1 | source2 | Drug and Indication | Outcome | IRR |
|---|---|---|---|---|
| AMBULATORY EMR | OPTUM Extended SES | New users of SGLT2 inhibitor nested in Left Heart Failure | Anaphylaxis | 10.7 |
| AMBULATORY EMR | OPTUM Extended SES | New users of Trimethoprim systemetic nested in Acute Typical Pneumonia | Anaphylaxis | 9.7 |
| AMBULATORY EMR | OPTUM Extended SES | New users of SGLT2 inhibitor nested in Type 2 diabetes mellitus | Anaphylaxis | 9.6 |
| AMBULATORY EMR | OPTUM Extended SES | New users of Beta blockers nested in essential hypertension | Hospitalization with heart failure events | <0.001 |
| AMBULATORY EMR | OPTUM Extended SES | New users of Beta blockers nested in essential hypertension | 4-point MACE | <0.001 |
| AMBULATORY EMR | OPTUM Extended SES | New users of Beta blockers nested in essential hypertension | Total cardiovascular disease events (ischemic stroke, hemorrhagic stroke, heart failure, acute myocardial infarction or sudden cardiac death) | <0.001 |

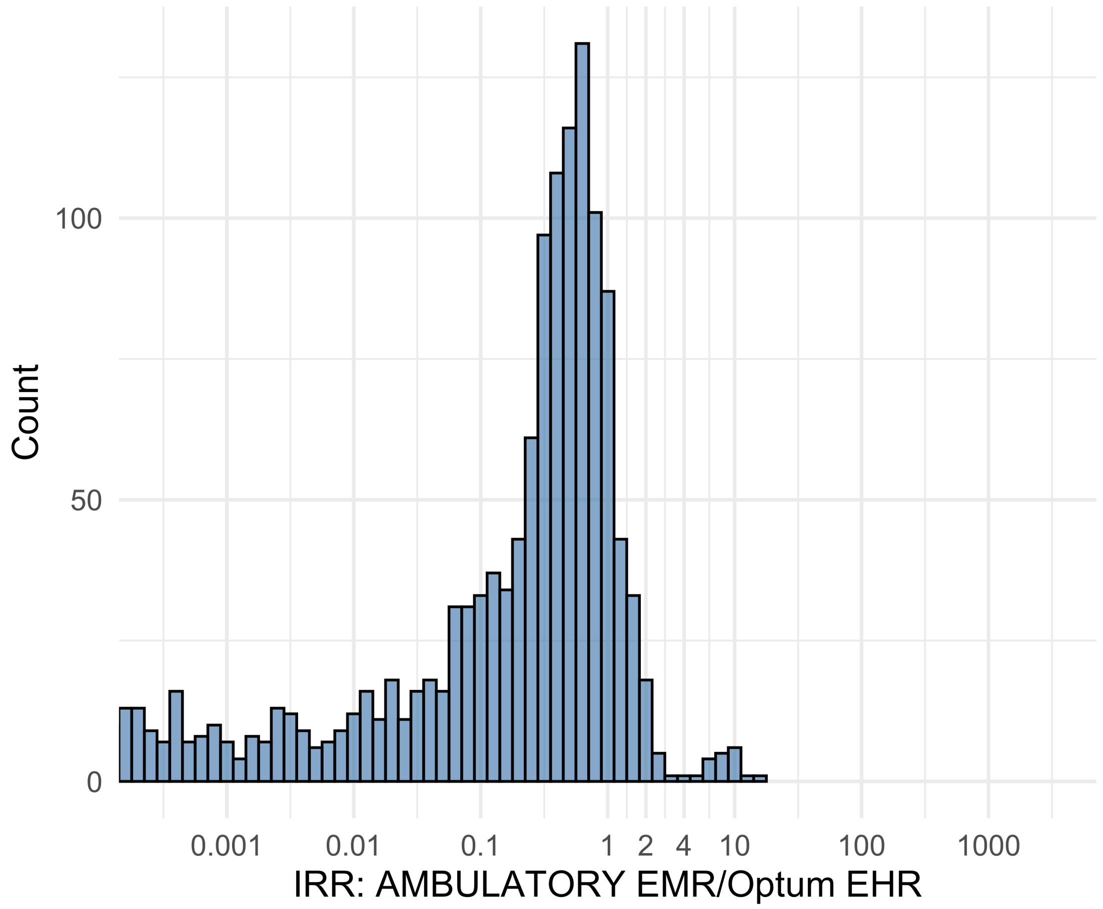


Figure S5.10. IRR distribution comparing AMBULATORY EMR and Optum EHR.

*Table S5.10. Most extreme IRRs for AMBULATORY EMR vs Optum EHR (rounded to 0.001).*

| source1 | source2 | Drug and Indication | Outcome | IRR |
|---|---|---|---|---|
| AMBULATORY EMR | Optum EHR | New users of Tumor Necrosis Factor alpha (TNFa) inhibitors nested in Psoriatic Arthritis | Anaphylaxis | 15.4 |
| AMBULATORY EMR | Optum EHR | New users of Trimethoprim systemetic nested in Acute Typical Pneumonia | Anaphylaxis | 11.7 |
| AMBULATORY EMR | Optum EHR | New users of Cephalosporin systemetic nested in Urinary Tract Infection | Anaphylaxis | 10.5 |
| AMBULATORY EMR | Optum EHR | New users of Beta blockers nested in essential hypertension | Hospitalization with heart failure events | <0.001 |
| AMBULATORY EMR | Optum EHR | New users of Beta blockers nested in essential hypertension | 4-point MACE | <0.001 |
| AMBULATORY EMR | Optum EHR | New users of Beta blockers nested in essential hypertension | Total cardiovascular disease events (ischemic stroke, hemorrhagic stroke, heart failure, acute myocardial infarction or sudden cardiac death) | <0.001 |

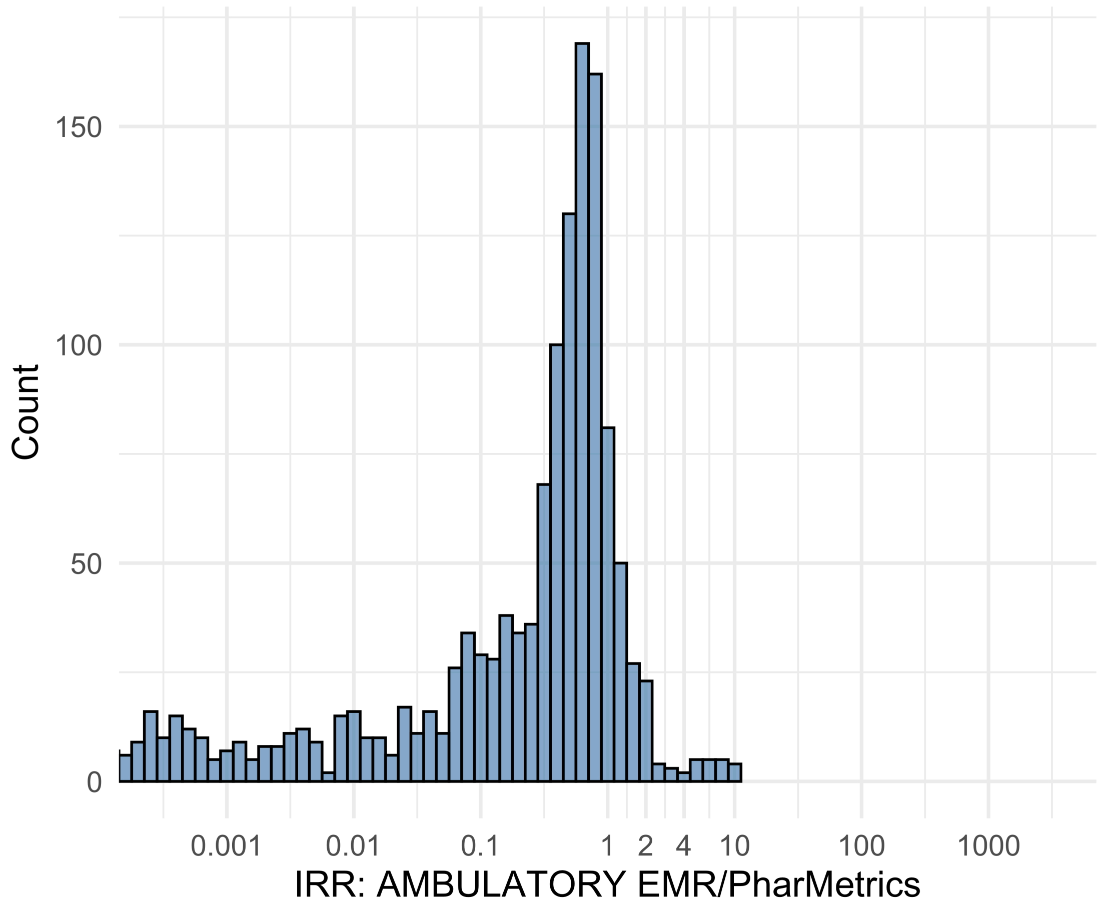


Figure S5.11. IRR distribution comparing AMBULATORY EMR and PharMetrics.

*Table S5.11. Most extreme IRRs for AMBULATORY EMR vs PharMetrics (rounded to 0.001).*

| source1 | source2 | Drug and Indication | Outcome | IRR |
|---|---|---|---|---|
| AMBULATORY EMR | PharMetrics | New users of Trimethoprim systemetic nested in Acute Typical Pneumonia | Anaphylaxis | 10.6 |
| AMBULATORY EMR | PharMetrics | New users of SGLT2 inhibitor nested in Type 2 diabetes mellitus | Anaphylaxis | 10.1 |
| AMBULATORY EMR | PharMetrics | New users of SGLT2 inhibitor nested in Left Heart Failure | Anaphylaxis | 9.6 |
| AMBULATORY EMR | PharMetrics | New users of Beta blockers nested in essential hypertension | Hospitalization with heart failure events | <0.001 |
| AMBULATORY EMR | PharMetrics | New users of Beta blockers nested in essential hypertension | 4-point MACE | <0.001 |
| AMBULATORY EMR | PharMetrics | New users of Beta blockers nested in essential hypertension | Total cardiovascular disease events (ischemic stroke, hemorrhagic stroke, heart failure, acute myocardial infarction or sudden cardiac death) | <0.001 |

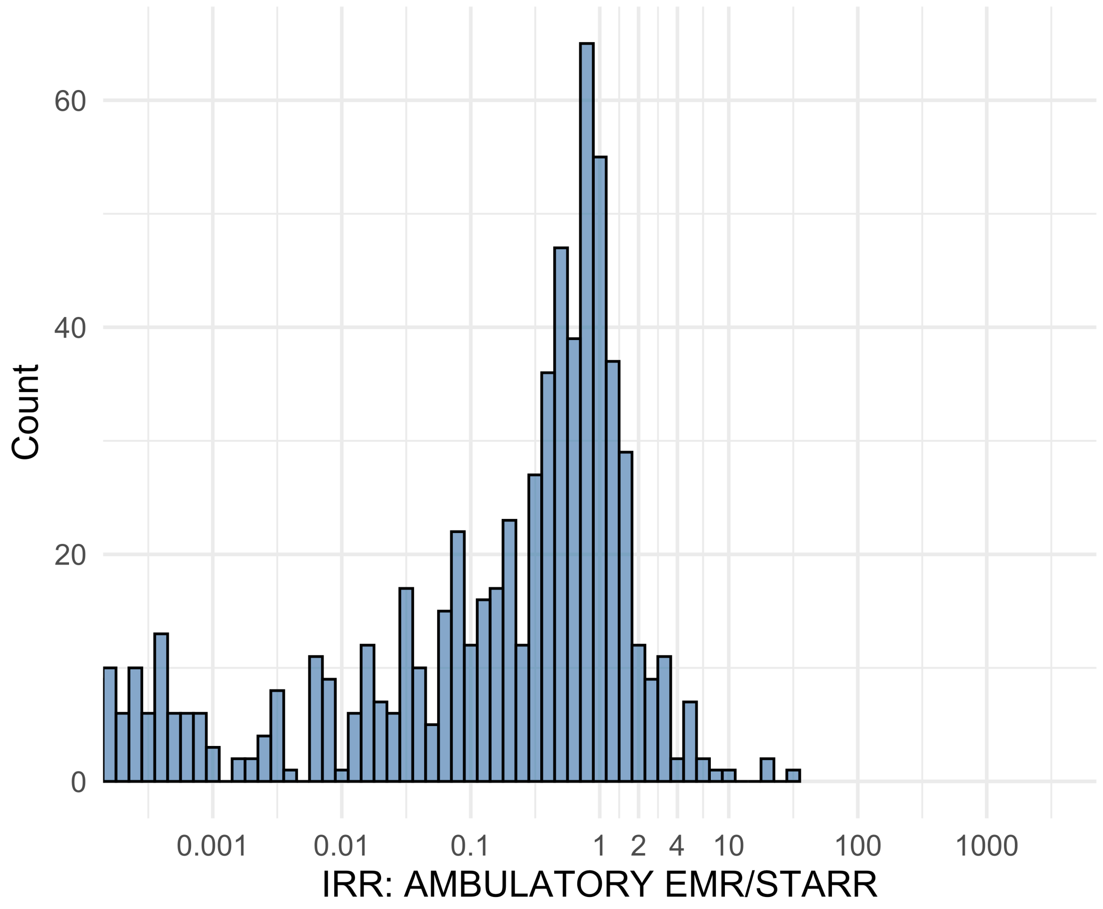


Figure S5.12. IRR distribution comparing AMBULATORY EMR and STARR.

*Table S5.12. Most extreme IRRs for AMBULATORY EMR vs STARR (rounded to 0.001).*

| source1 | source2 | Drug and Indication | Outcome | IRR |
|---|---|---|---|---|
| AMBULATORY EMR | STARR | New users of SGLT2 inhibitor nested in Type 2 diabetes mellitus | Anaphylaxis | 33.3 |
| AMBULATORY EMR | STARR | New users of GLP-1 receptor antagonists nested in obesity | Anaphylaxis | 19.7 |
| AMBULATORY EMR | STARR | New users of GLP-1 receptor antagonists nested in Type 2 diabetes mellitus | Anaphylaxis | 19.4 |
| AMBULATORY EMR | STARR | New users of Beta blockers nested in essential hypertension | Hospitalization with heart failure events | <0.001 |
| AMBULATORY EMR | STARR | New users of Beta blockers nested in essential hypertension | 4-point MACE | <0.001 |
| AMBULATORY EMR | STARR | New users of Beta blockers nested in essential hypertension | Total cardiovascular disease events (ischemic stroke, hemorrhagic stroke, heart failure, acute myocardial infarction or sudden cardiac death) | <0.001 |

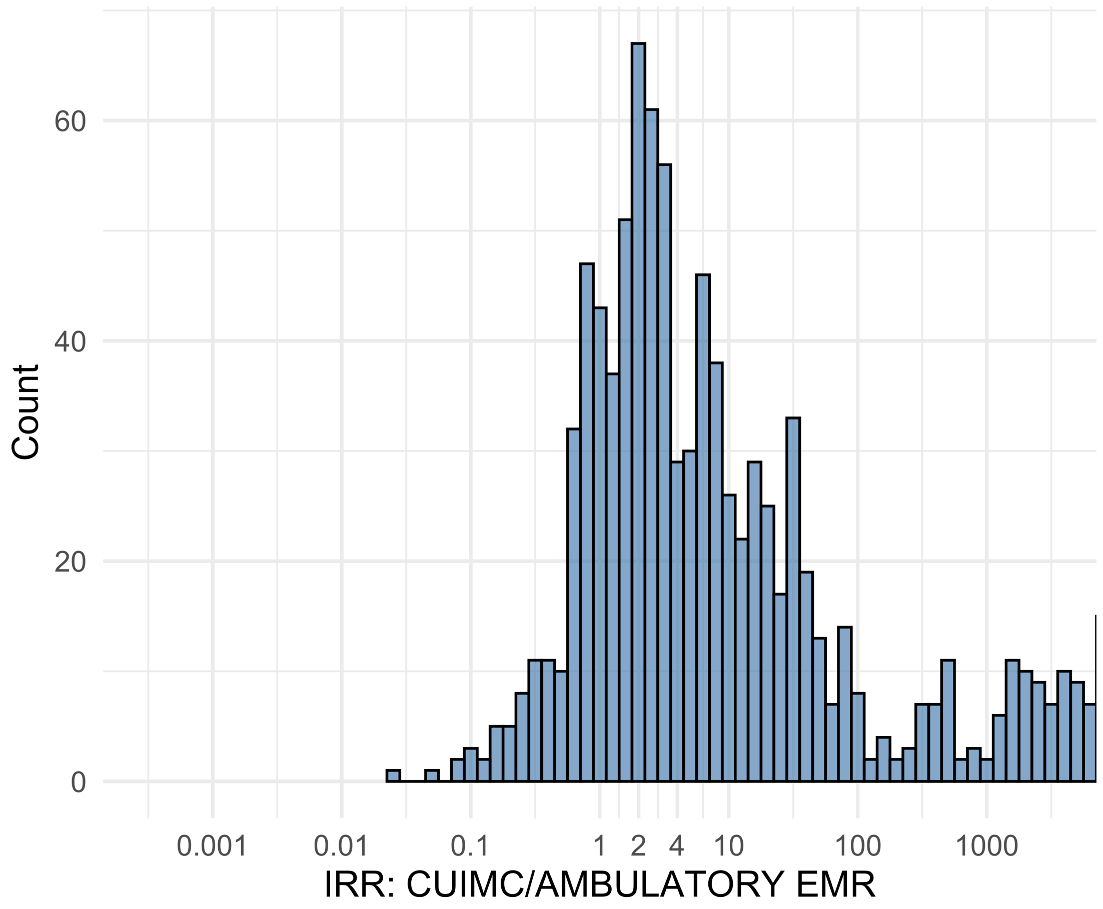


Figure S5.13. IRR distribution comparing CUIMC and AMBULATORY EMR.

*Table S5.13. Most extreme IRRs for CUIMC vs AMBULATORY EMR (rounded to 0.001).*

| source1 | source2 | Drug and Indication | Outcome | IRR |
|---|---|---|---|---|
| CUIMC | AMBULATORY EMR | New users of Beta blockers nested in essential hypertension | 4-point MACE | 248200.8 |
| CUIMC | AMBULATORY EMR | New users of Beta blockers nested in essential hypertension | Total cardiovascular disease events (ischemic stroke, hemorrhagic stroke, heart failure, acute myocardial infarction or sudden cardiac death) | 245925.5 |
| CUIMC | AMBULATORY EMR | New users of Beta blockers nested in essential hypertension | Hospitalization with heart failure events | 226639.2 |
| CUIMC | AMBULATORY EMR | New users of SGLT2 inhibitor nested in Left Heart Failure | Anaphylaxis | 0.0 |
| CUIMC | AMBULATORY EMR | New users of Tumor Necrosis Factor alpha (TNFa) inhibitors nested in Crohns disease | Anaphylaxis | 0.1 |
| CUIMC | AMBULATORY EMR | New users of SGLT2 inhibitor nested in Type 2 diabetes mellitus | Anaphylaxis | 0.1 |

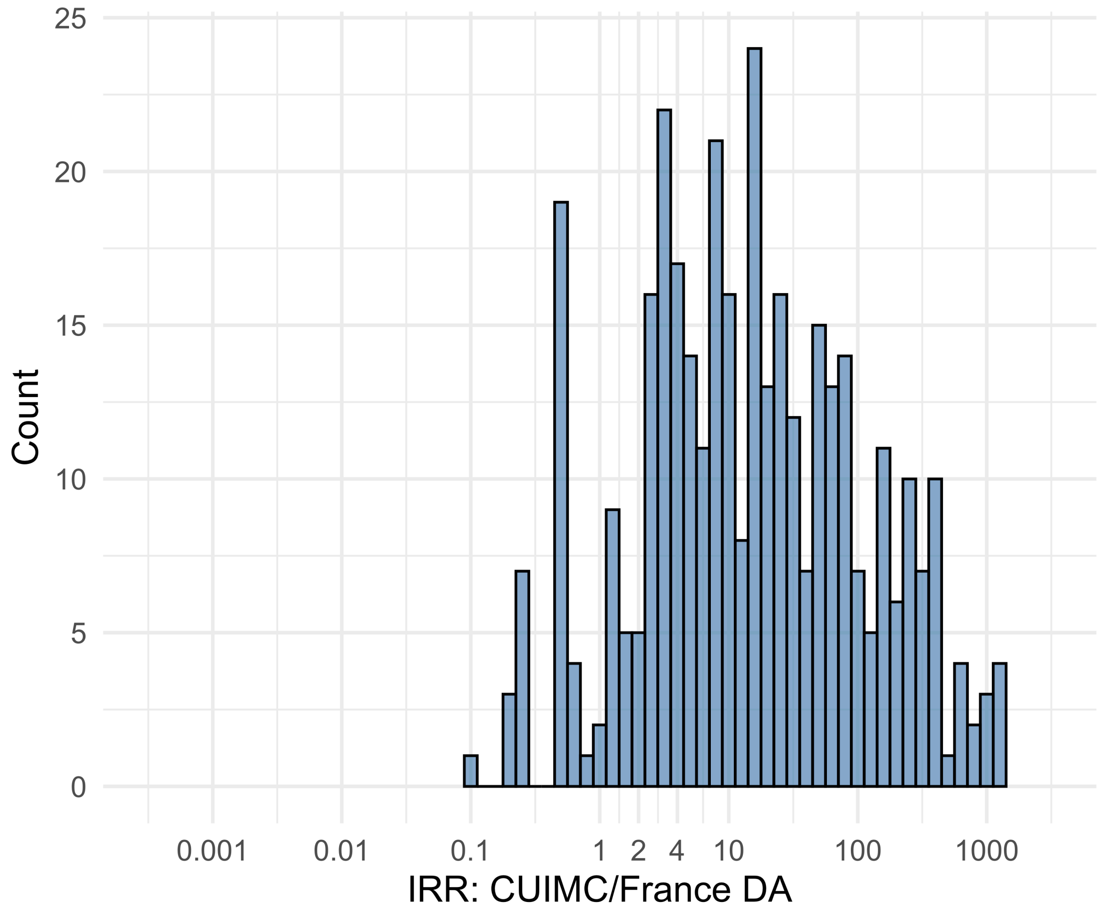


Figure S5.14. IRR distribution comparing CUIMC and France DA.

*Table S5.14. Most extreme IRRs for CUIMC vs France DA (rounded to 0.001).*

| source1 | source2 | Drug and Indication | Outcome | IRR |
|---|---|---|---|---|
| CUIMC | France DA | New users of Beta blockers nested in essential hypertension | Acute Kidney Injury AKI | 1375.7 |
| CUIMC | France DA | New users of Fluoroquinolone systemic nested in Urinary Tract Infection | Acute Kidney Injury AKI | 1306.4 |
| CUIMC | France DA | New users of Beta blockers nested in essential hypertension | 4-point MACE | 1219.3 |
| CUIMC | France DA | New users of Beta blockers nested in essential hypertension | Polymorphic Ventricular Tachycardia or Torsades de Pointes | 0.1 |
| CUIMC | France DA | New users of GLP-1 receptor antagonists nested in Type 2 diabetes mellitus | Progressive multifocal leukoencephalopathy | 0.2 |
| CUIMC | France DA | New users of GLP-1 receptor antagonists nested in Type 2 diabetes mellitus | Polymorphic Ventricular Tachycardia or Torsades de Pointes | 0.2 |

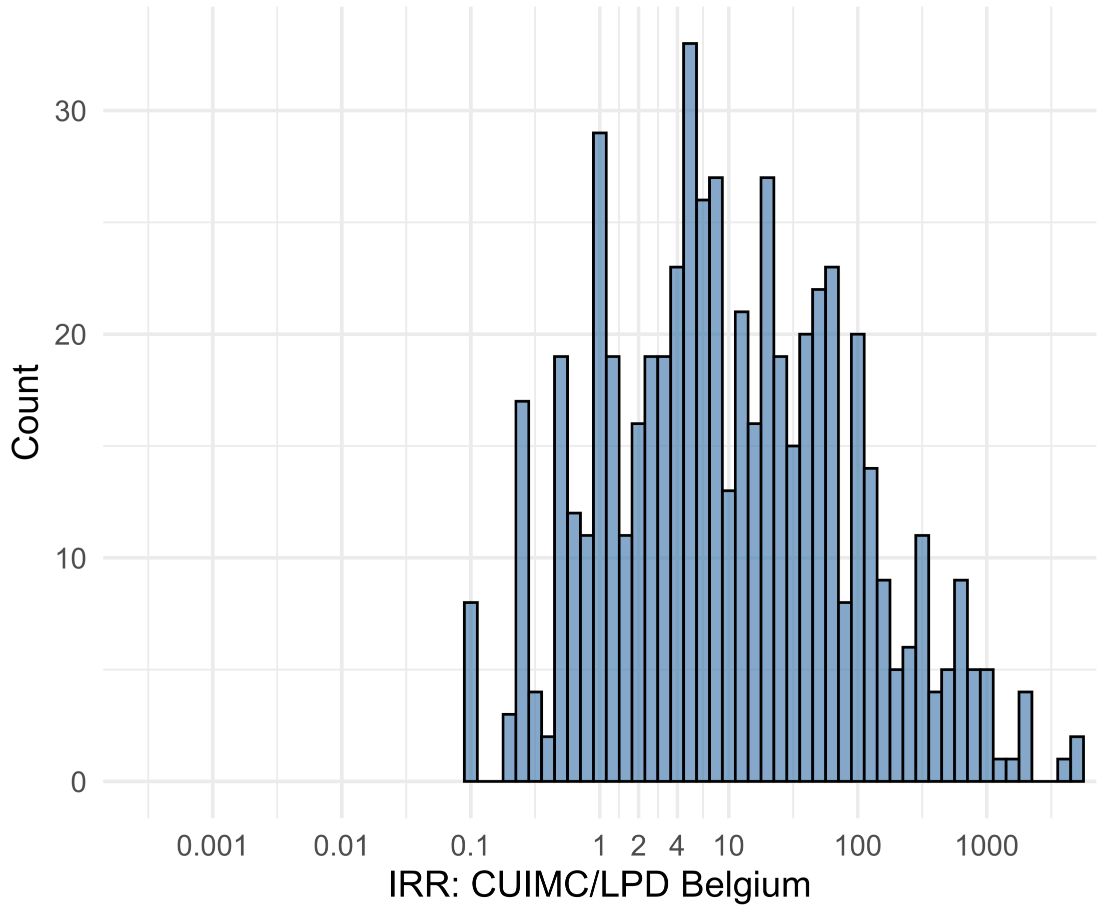


Figure S5.15. IRR distribution comparing CUIMC and LPD Belgium.

*Table S5.15. Most extreme IRRs for CUIMC vs LPD Belgium (rounded to 0.001).*

| source1 | source2 | Drug and Indication | Outcome | IRR |
|---|---|---|---|---|
| CUIMC | LPD Belgium | New users of Beta blockers nested in essential hypertension | 4-point MACE | 4745.1 |
| CUIMC | LPD Belgium | New users of Beta blockers nested in essential hypertension | Total cardiovascular disease events (ischemic stroke, hemorrhagic stroke, heart failure, acute myocardial infarction or sudden cardiac death) | 4701.6 |
| CUIMC | LPD Belgium | New users of Beta blockers nested in essential hypertension | Hospitalization with heart failure events | 4332.9 |
| CUIMC | LPD Belgium | New users of GLP-1 receptor antagonists nested in obesity | Polymorphic Ventricular Tachycardia or Torsades de Pointes | 0.1 |
| CUIMC | LPD Belgium | New users of GLP-1 receptor antagonists nested in obesity | Progressive multifocal leukoencephalopathy | 0.1 |
| CUIMC | LPD Belgium | New users of GLP-1 receptor antagonists nested in obesity | Stevens-Johnson syndrome, toxic epidermal necrolysis spectrum | 0.1 |

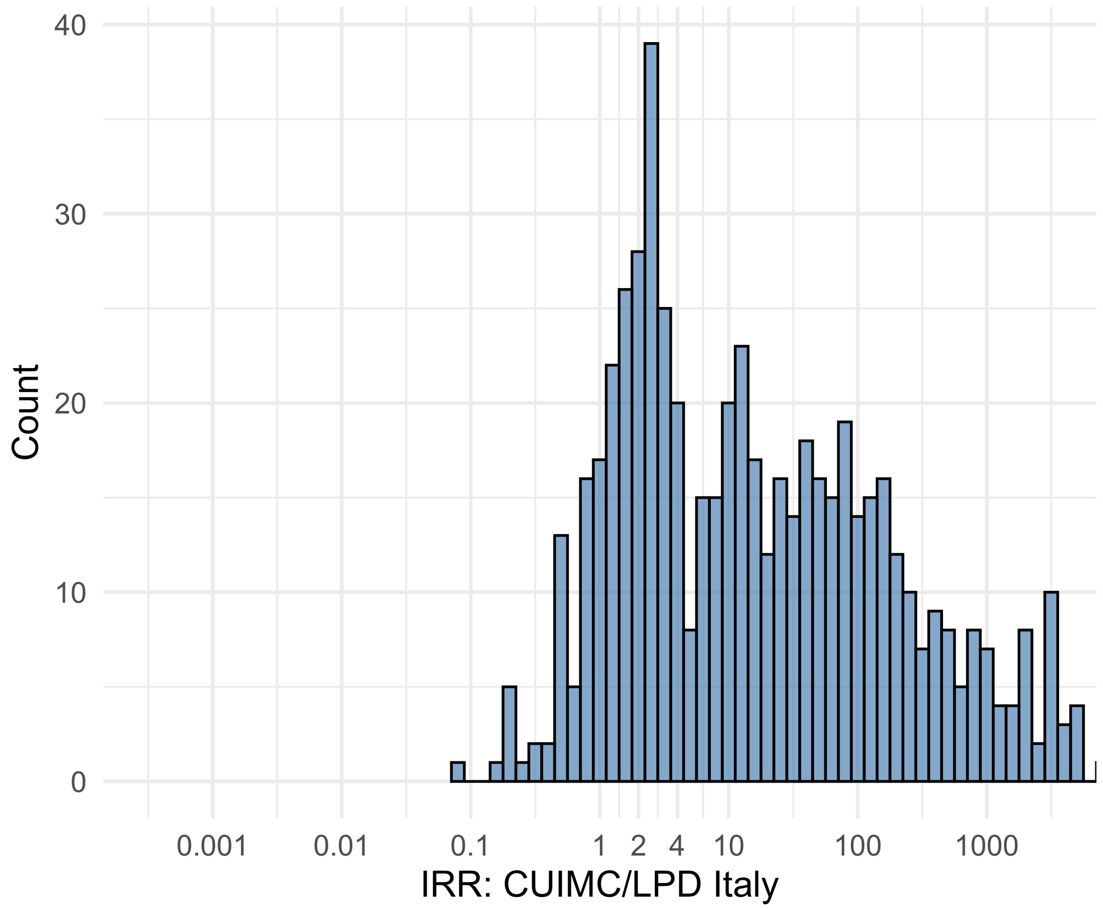


Figure S5.16. IRR distribution comparing CUIMC and LPD Italy.

*Table S5.16. Most extreme IRRs for CUIMC vs LPD Italy (rounded to 0.001).*

| source1 | source2 | Drug and Indication | Outcome | IRR |
|---|---|---|---|---|
| CUIMC | LPD Italy | New users of Beta blockers nested in essential hypertension | 4-point MACE | 15427.8 |
| CUIMC | LPD Italy | New users of Beta blockers nested in essential hypertension | Total cardiovascular disease events (ischemic stroke, hemorrhagic stroke, heart failure, acute myocardial infarction or sudden cardiac death) | 15286.3 |
| CUIMC | LPD Italy | New users of Beta blockers nested in essential hypertension | Hospitalization with heart failure events | 14087.5 |
| CUIMC | LPD Italy | New users of Cephalosporin systemetic nested in Urinary Tract Infection | Progressive multifocal leukoencephalopathy | 0.1 |
| CUIMC | LPD Italy | New users of SGLT2 inhibitor nested in Type 2 diabetes mellitus | Autoimmune hemolytic anemia | 0.1 |
| CUIMC | LPD Italy | New users of Beta blockers nested in essential hypertension | Progressive multifocal leukoencephalopathy | 0.2 |

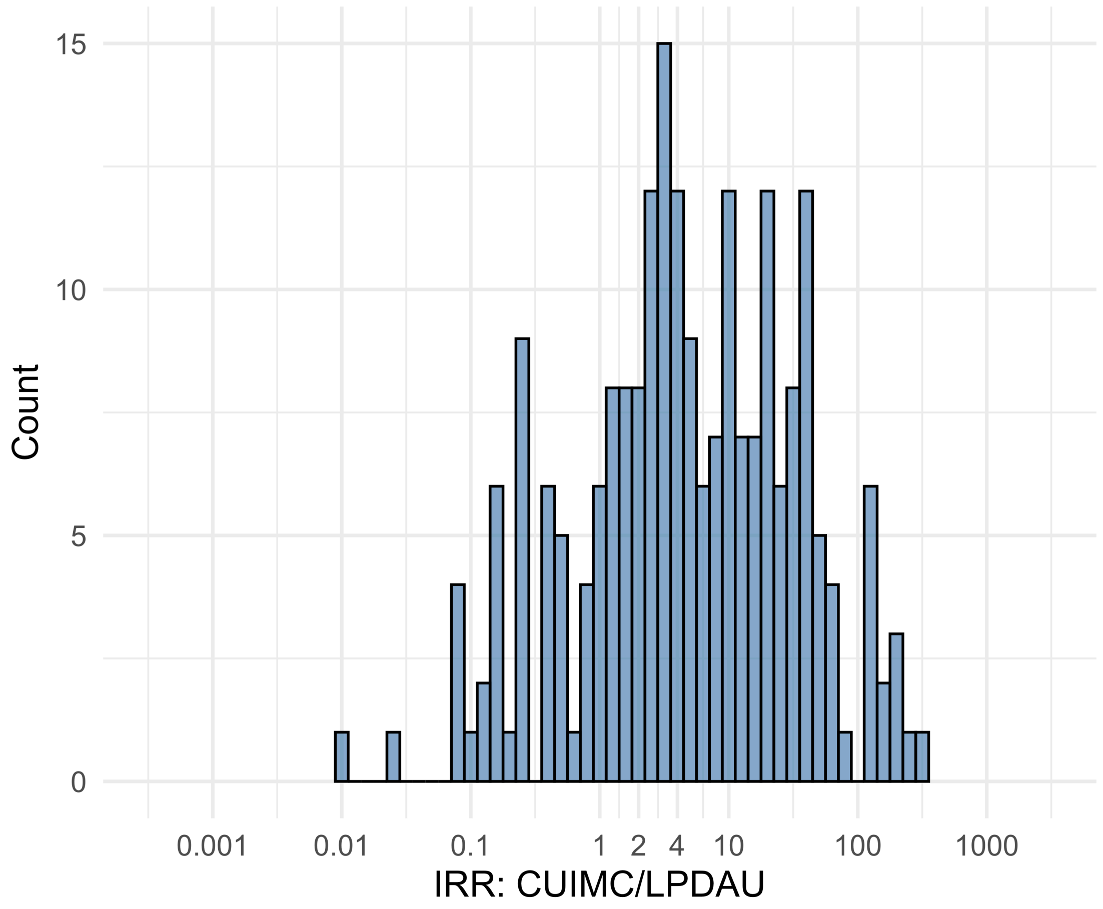


Figure S5.17. IRR distribution comparing CUIMC and LPDAU.

*Table S5.17. Most extreme IRRs for CUIMC vs LPDAU (rounded to 0.001).*

| source1 | source2 | Drug and Indication | Outcome | IRR |
|---|---|---|---|---|
| CUIMC | LPDAU | New users of Cephalosporin systemetic nested in Urinary Tract Infection | Acute Kidney Injury AKI | 283.0 |
| CUIMC | LPDAU | New users of Trimethoprim systemetic nested in Urinary Tract Infection | Acute Kidney Injury AKI | 236.1 |
| CUIMC | LPDAU | New users of Cephalosporin systemetic nested in Urinary Tract Infection | Hospitalization with heart failure events | 189.8 |
| CUIMC | LPDAU | New users of Beta blockers nested in essential hypertension | Polymorphic Ventricular Tachycardia or Torsades de Pointes | 0.0 |
| CUIMC | LPDAU | New users of Beta blockers nested in essential hypertension | All events Drug Rash with Eosinophilia and Systemic Symptoms (DRESS) | 0.0 |
| CUIMC | LPDAU | New users of Cephalosporin systemetic nested in Urinary Tract Infection | Polymorphic Ventricular Tachycardia or Torsades de Pointes | 0.1 |

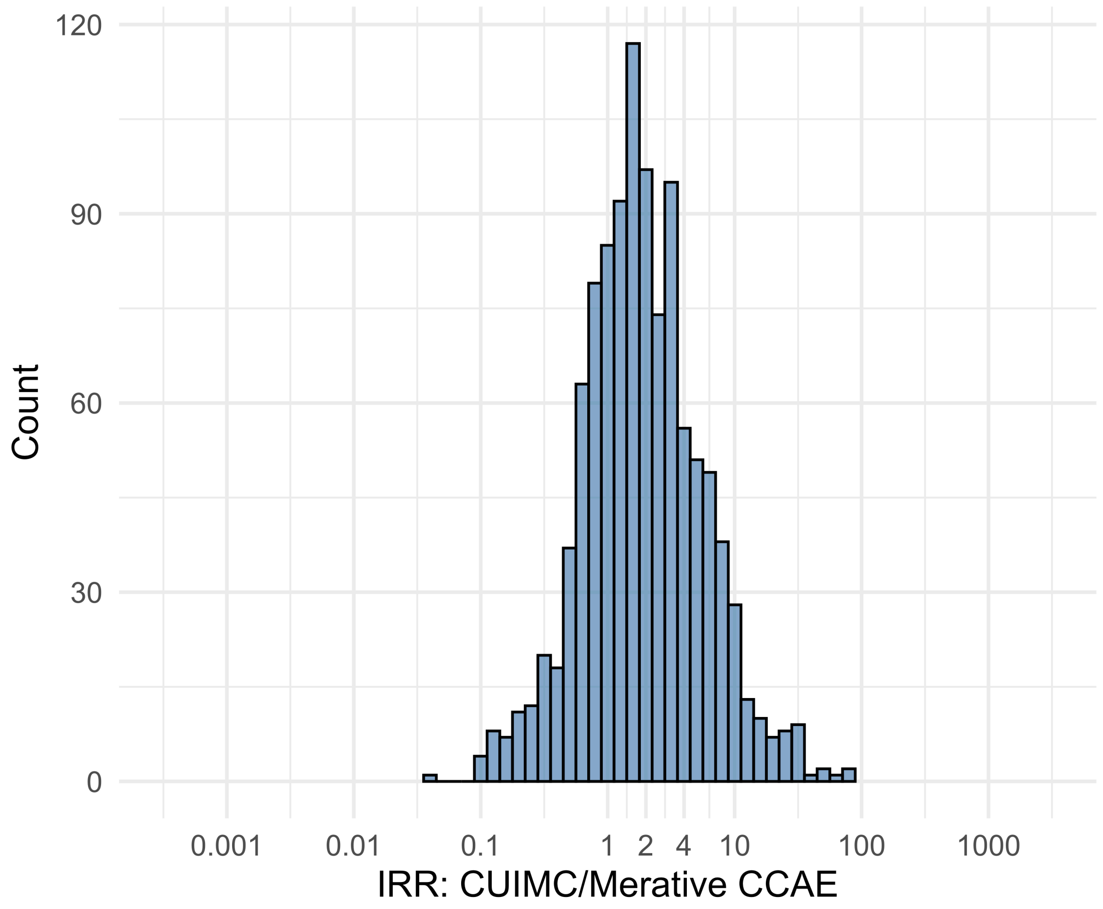


Figure S5.18. IRR distribution comparing CUIMC and Merative CCAE.

*Table S5.18. Most extreme IRRs for CUIMC vs Merative CCAE (rounded to 0.001).*

| source1 | source2 | Drug and Indication | Outcome | IRR |
|---|---|---|---|---|
| CUIMC | Merative CCAE | New users of Trimethoprim systemetic nested in Urinary Tract Infection | Polymorphic Ventricular Tachycardia or Torsades de Pointes | 78.2 |
| CUIMC | Merative CCAE | New users of Fluoroquinolone systemic nested in Urinary Tract Infection | Polymorphic Ventricular Tachycardia or Torsades de Pointes | 71.9 |
| CUIMC | Merative CCAE | New users of Trimethoprim systemetic nested in Urinary Tract Infection | Cardiovascular-related mortality | 59.6 |
| CUIMC | Merative CCAE | New users of Beta blockers nested in Acute Myocardial Infarction | Abnormal weight gain events | 0.0 |
| CUIMC | Merative CCAE | New users of Cephalosporin systemetic nested in Acute Typical Pneumonia | Abnormal weight gain events | 0.1 |
| CUIMC | Merative CCAE | New users of Tumor Necrosis Factor alpha (TNFa) inhibitors nested in Rheumatoid arthritis | All events of Acute Liver Injury | 0.1 |

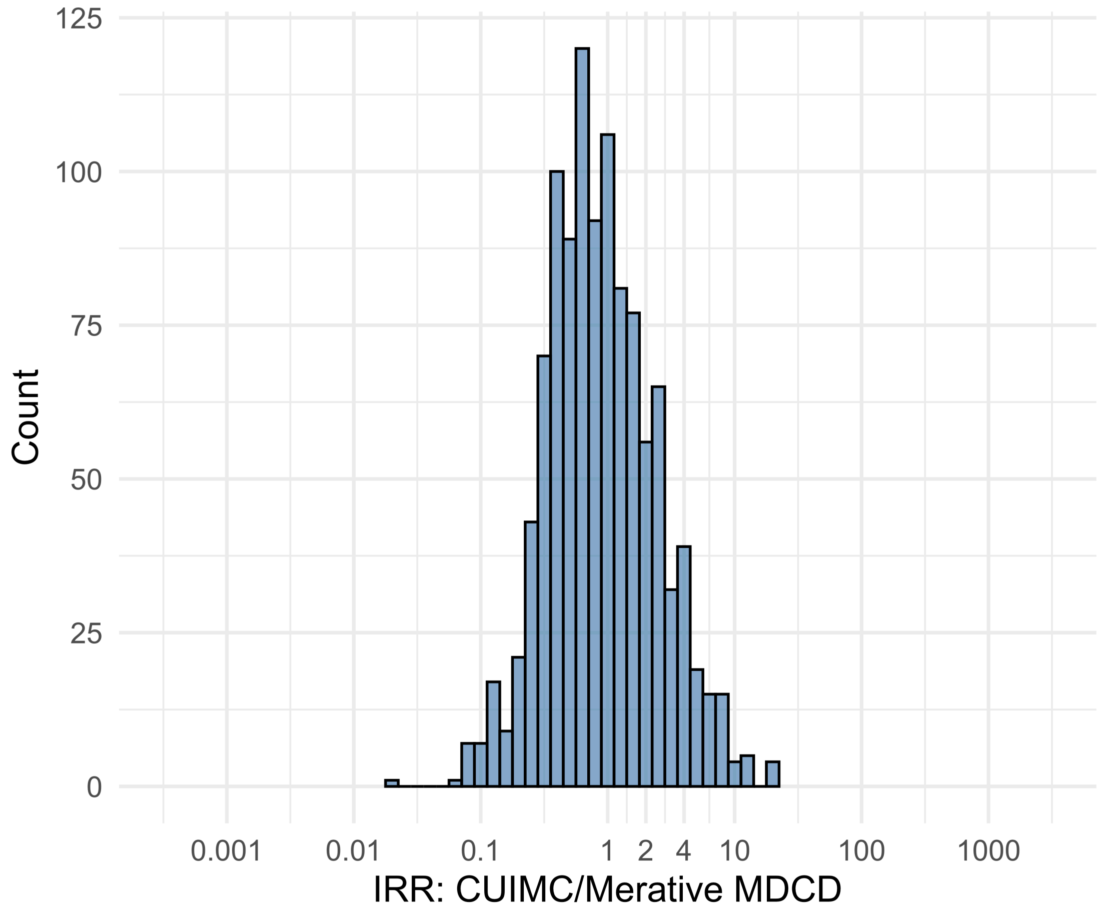


Figure S5.19. IRR distribution comparing CUIMC and Merative MDCD.

*Table S5.19. Most extreme IRRs for CUIMC vs Merative MDCD (rounded to 0.001).*

| source1 | source2 | Drug and Indication | Outcome | IRR |
|---|---|---|---|---|
| CUIMC | Merative MDCD | New users of Fluoroquinolone systemic nested in Urinary Tract Infection | Polymorphic Ventricular Tachycardia or Torsades de Pointes | 22.2 |
| CUIMC | Merative MDCD | New users of SGLT2 inhibitor nested in Left Heart Failure | All events of Autoimmune hepatitis, with a washout period of 365 days | 19.7 |
| CUIMC | Merative MDCD | New users of Trimethoprim systemetic nested in Urinary Tract Infection | Progressive multifocal leukoencephalopathy | 18.6 |
| CUIMC | Merative MDCD | New users of Beta blockers nested in Acute Myocardial Infarction | Abnormal weight gain events | 0.0 |
| CUIMC | Merative MDCD | New users of Tumor Necrosis Factor alpha (TNFa) inhibitors nested in Rheumatoid arthritis | Acute Myocardial Infarction including its complications | 0.1 |
| CUIMC | Merative MDCD | New users of Tumor Necrosis Factor alpha (TNFa) inhibitors nested in Rheumatoid arthritis | Coronary Vessel Revascularization | 0.1 |

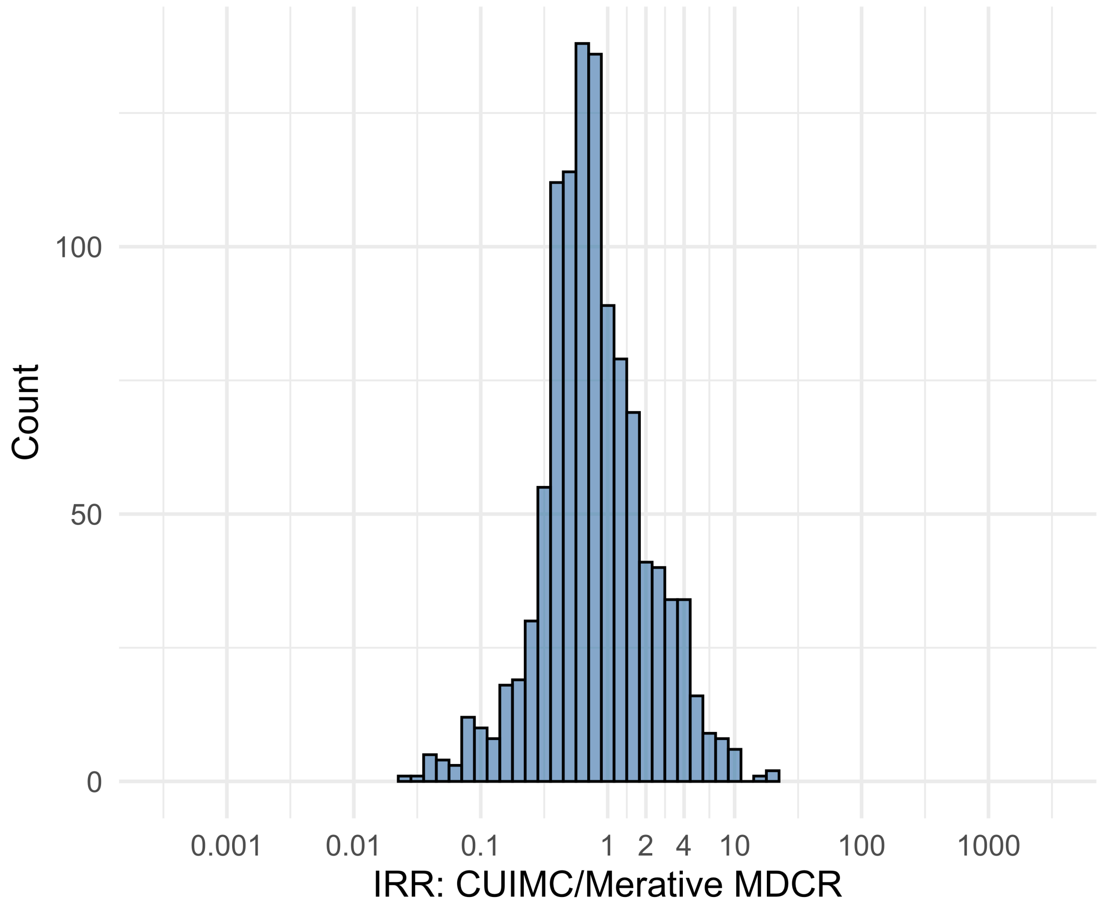


Figure S5.20. IRR distribution comparing CUIMC and Merative MDCR.

*Table S5.20. Most extreme IRRs for CUIMC vs Merative MDCR (rounded to 0.001).*

| source1 | source2 | Drug and Indication | Outcome | IRR |
|---|---|---|---|---|
| CUIMC | Merative MDCR | New users of Tumor Necrosis Factor alpha (TNFa) inhibitors nested in Rheumatoid arthritis | Myocarditis Pericarditis | 21.4 |
| CUIMC | Merative MDCR | New users of Trimethoprim systemetic nested in Acute Typical Pneumonia | Earliest event of Thrombotic microangiopathy (TMA) or Microangiopathic hemolytic anemia (MAHA) | 18.7 |
| CUIMC | Merative MDCR | New users of Beta blockers nested in essential hypertension | Progressive multifocal leukoencephalopathy | 14.4 |
| CUIMC | Merative MDCR | New users of Tumor Necrosis Factor alpha (TNFa) inhibitors nested in Crohns disease | Stroke (ischemic or hemorrhagic) events | 0.0 |
| CUIMC | Merative MDCR | New users of Tumor Necrosis Factor alpha (TNFa) inhibitors nested in Rheumatoid arthritis | Coronary Vessel Revascularization | 0.0 |
| CUIMC | Merative MDCR | New users of Tumor Necrosis Factor alpha (TNFa) inhibitors nested in Rheumatoid arthritis | Acute Myocardial Infarction including its complications | 0.0 |

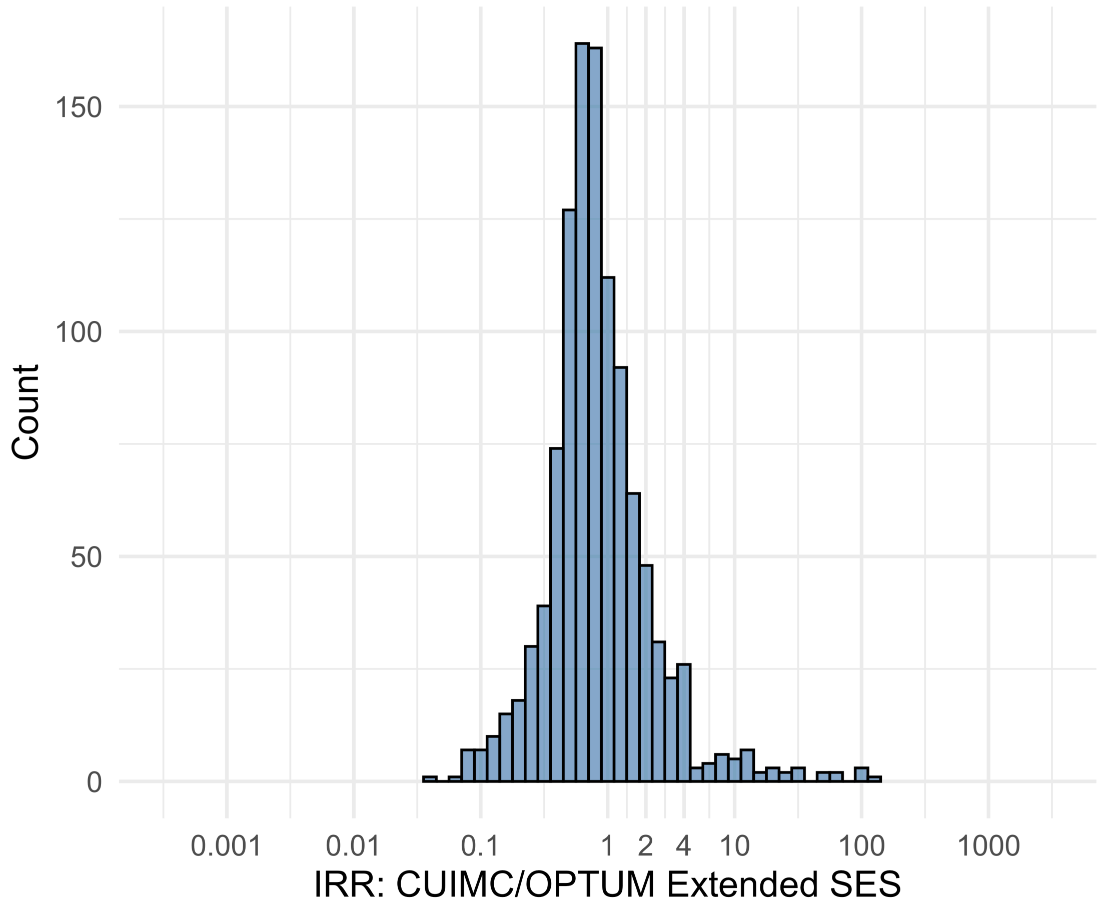


Figure S5.21. IRR distribution comparing CUIMC and OPTUM Extended SES.

*Table S5.21. Most extreme IRRs for CUIMC vs OPTUM Extended SES (rounded to 0.001).*

| source1 | source2 | Drug and Indication | Outcome | IRR |
|---|---|---|---|---|
| CUIMC | OPTUM Extended SES | New users of Tumor Necrosis Factor alpha (TNFa) inhibitors nested in Rheumatoid arthritis | Cardiovascular-related mortality | 126.5 |
| CUIMC | OPTUM Extended SES | New users of Fluoroquinolone systemic nested in Acute Typical Pneumonia | Cardiovascular-related mortality | 100.0 |
| CUIMC | OPTUM Extended SES | New users of Cephalosporin systemetic nested in Acute Typical Pneumonia | Cardiovascular-related mortality | 94.8 |
| CUIMC | OPTUM Extended SES | New users of Beta blockers nested in Acute Myocardial Infarction | Abnormal weight gain events | 0.0 |
| CUIMC | OPTUM Extended SES | New users of SGLT2 inhibitor nested in Left Heart Failure | Rhabdomyolysis | 0.1 |
| CUIMC | OPTUM Extended SES | New users of Tumor Necrosis Factor alpha (TNFa) inhibitors nested in Rheumatoid arthritis | All events of Acute Liver Injury | 0.1 |

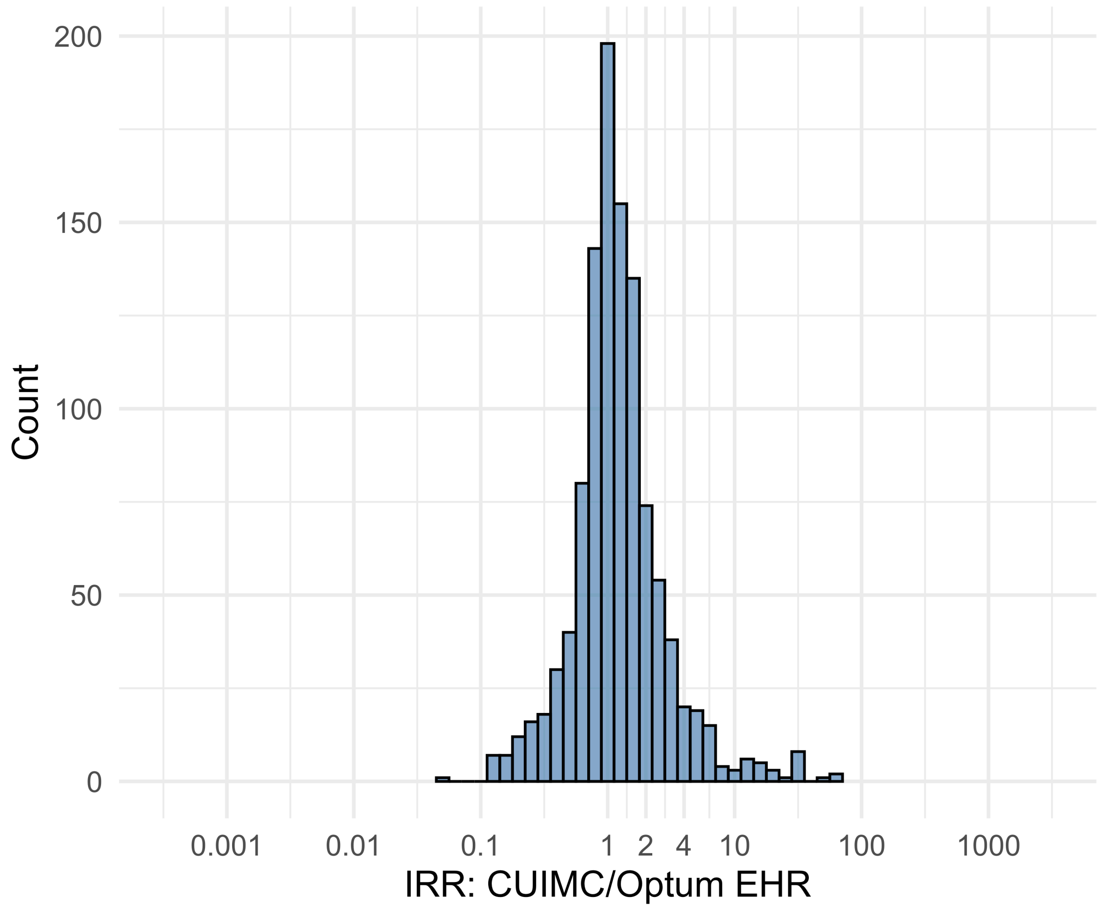


Figure S5.22. IRR distribution comparing CUIMC and Optum EHR.

*Table S5.22. Most extreme IRRs for CUIMC vs Optum EHR (rounded to 0.001).*

| source1 | source2 | Drug and Indication | Outcome | IRR |
|---|---|---|---|---|
| CUIMC | Optum EHR | New users of Tumor Necrosis Factor alpha (TNFa) inhibitors nested in Rheumatoid arthritis | All events Drug Rash with Eosinophilia and Systemic Symptoms (DRESS) | 60.8 |
| CUIMC | Optum EHR | New users of Tumor Necrosis Factor alpha (TNFa) inhibitors nested in Rheumatoid arthritis | Polymorphic Ventricular Tachycardia or Torsades de Pointes | 60.7 |
| CUIMC | Optum EHR | New users of Trimethoprim systemetic nested in Urinary Tract Infection | Polymorphic Ventricular Tachycardia or Torsades de Pointes | 51.5 |
| CUIMC | Optum EHR | New users of Beta blockers nested in Acute Myocardial Infarction | Abnormal weight gain events | 0.1 |
| CUIMC | Optum EHR | New users of Tumor Necrosis Factor alpha (TNFa) inhibitors nested in Rheumatoid arthritis | Coronary Vessel Revascularization | 0.1 |
| CUIMC | Optum EHR | New users of SGLT2 inhibitor nested in Left Heart Failure | Rhabdomyolysis | 0.1 |

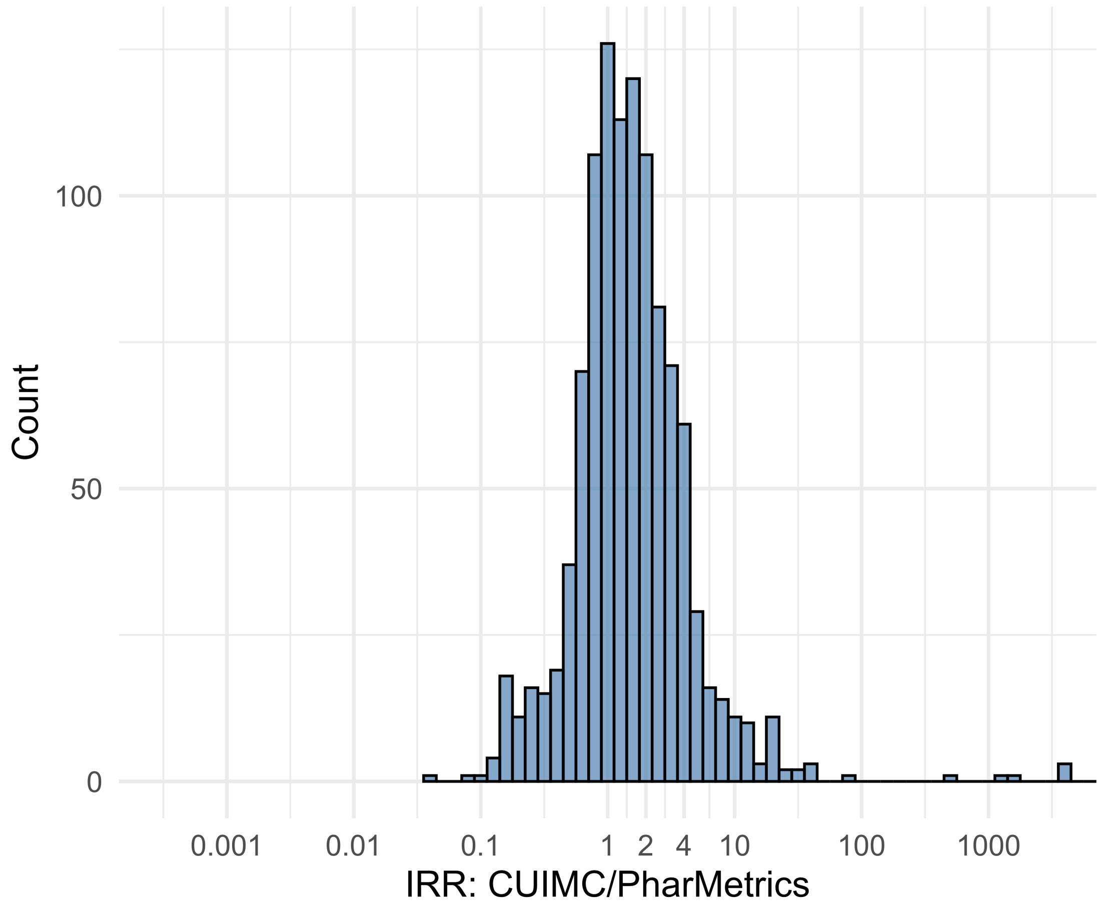


Figure S5.23. IRR distribution comparing CUIMC and PharMetrics.

*Table S5.23. Most extreme IRRs for CUIMC vs PharMetrics (rounded to 0.001).*

| source1 | source2 | Drug and Indication | Outcome | IRR |
|---|---|---|---|---|
| CUIMC | PharMetrics | New users of Cephalosporin systemetic nested in Acute Typical Pneumonia | Cardiovascular-related mortality | 43892.3 |
| CUIMC | PharMetrics | New users of Beta blockers nested in essential hypertension | Cardiovascular-related mortality | 38334.4 |
| CUIMC | PharMetrics | New users of Fluoroquinolone systemic nested in Acute Typical Pneumonia | Cardiovascular-related mortality | 29294.6 |
| CUIMC | PharMetrics | New users of Beta blockers nested in Acute Myocardial Infarction | Abnormal weight gain events | 0.0 |
| CUIMC | PharMetrics | New users of SGLT2 inhibitor nested in Left Heart Failure | Rhabdomyolysis | 0.1 |
| CUIMC | PharMetrics | New users of Cephalosporin systemetic nested in Acute Typical Pneumonia | Abnormal weight gain events | 0.1 |

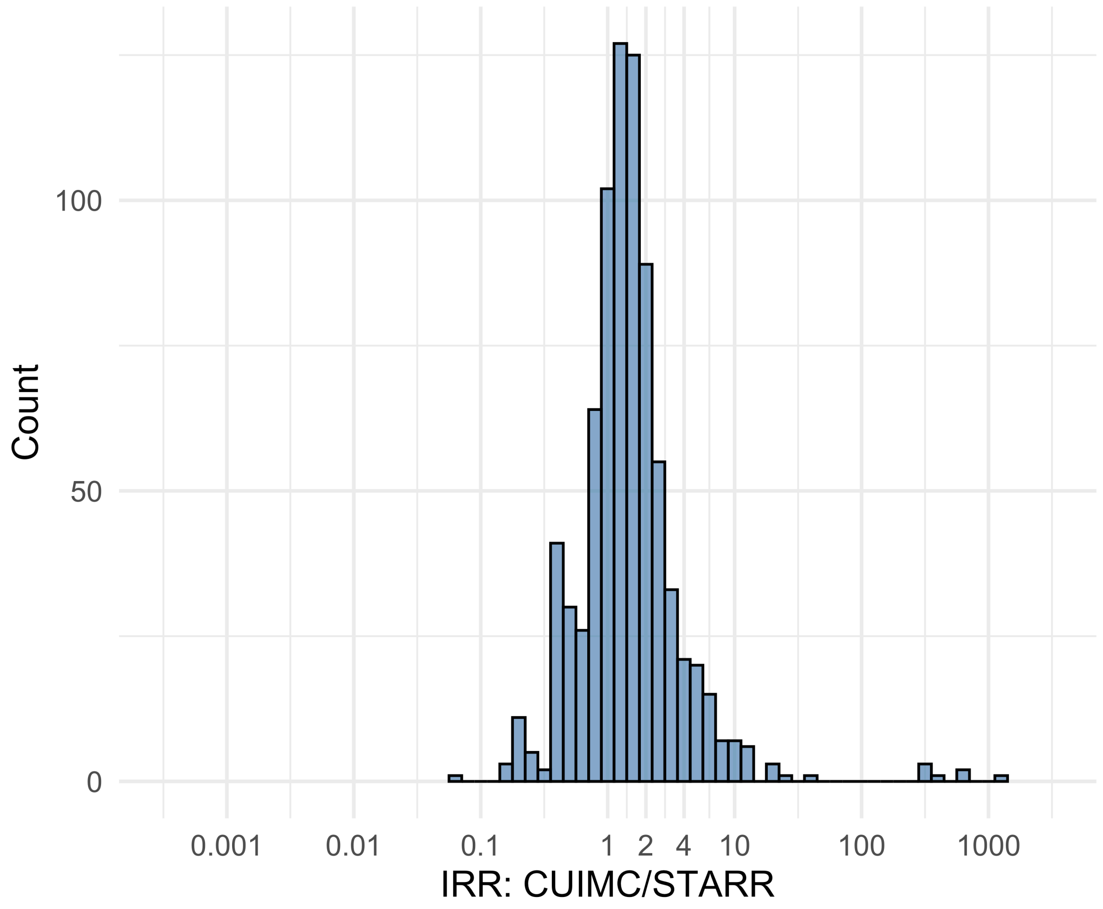


Figure S5.24. IRR distribution comparing CUIMC and STARR.

*Table S5.24. Most extreme IRRs for CUIMC vs STARR (rounded to 0.001).*

| source1 | source2 | Drug and Indication | Outcome | IRR |
|---|---|---|---|---|
| CUIMC | STARR | New users of Beta blockers nested in essential hypertension | Febrile Neutropenia or Neutropenic Fever | 1202.7 |
| CUIMC | STARR | New users of Fluoroquinolone systemic nested in Acute Typical Pneumonia | Febrile Neutropenia or Neutropenic Fever | 596.6 |
| CUIMC | STARR | New users of Fluoroquinolone systemic nested in Urinary Tract Infection | Febrile Neutropenia or Neutropenic Fever | 576.6 |
| CUIMC | STARR | New users of GLP-1 receptor antagonists nested in obesity | Abnormal weight loss events | 0.1 |
| CUIMC | STARR | New users of Fluoroquinolone systemic nested in Acute Typical Pneumonia | Earliest event of Thrombotic microangiopathy (TMA) or Microangiopathic hemolytic anemia (MAHA) | 0.1 |
| CUIMC | STARR | New users of Beta blockers nested in Acute Myocardial Infarction | Acquired Pure Red Cell Aplasia | 0.2 |

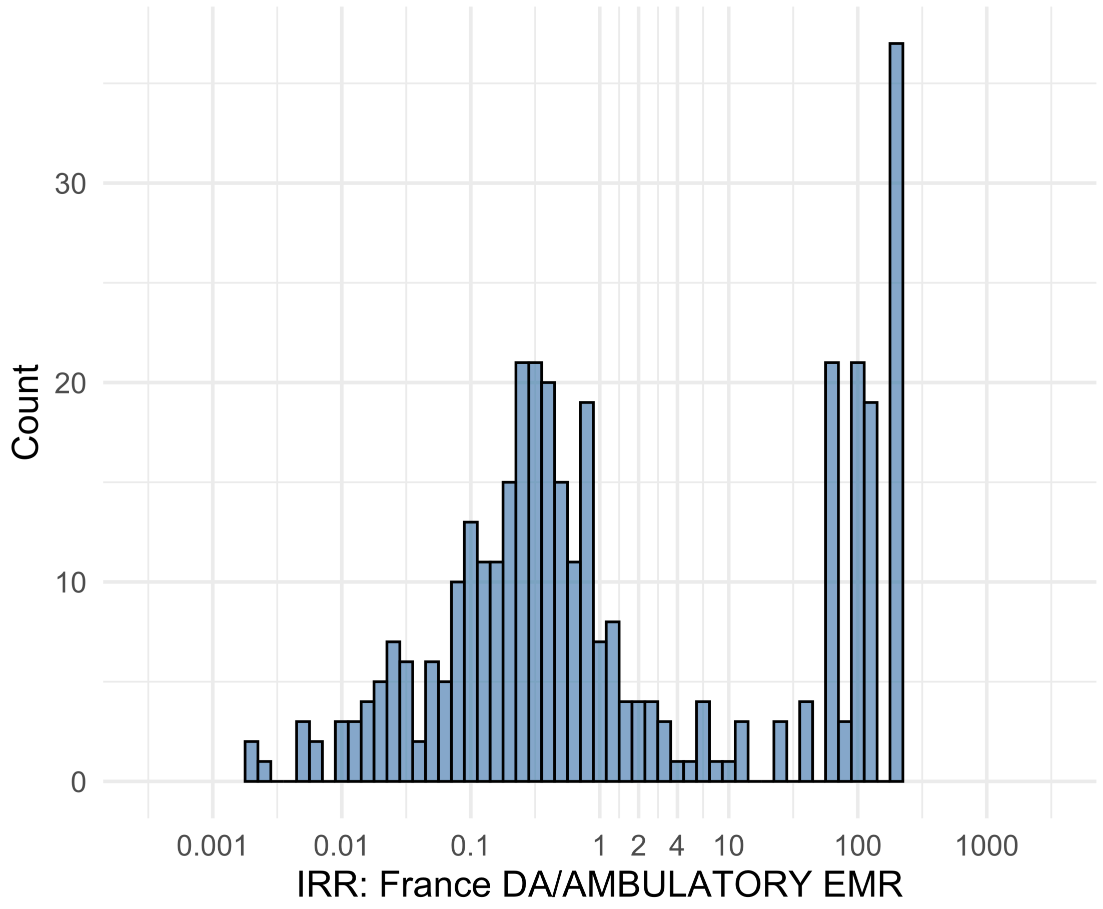


Figure S5.25. IRR distribution comparing France DA and AMBULATORY EMR.

*Table S5.25. Most extreme IRRs for France DA vs AMBULATORY EMR (rounded to 0.001).*

| source1 | source2 | Drug and Indication | Outcome | IRR |
|---|---|---|---|---|
| France DA | AMBULATORY EMR | New users of Trimethoprim systemetic nested in Urinary Tract Infection | Febrile Neutropenia or Neutropenic Fever | 216.0 |
| France DA | AMBULATORY EMR | New users of Trimethoprim systemetic nested in Urinary Tract Infection | Rhabdomyolysis | 216.0 |
| France DA | AMBULATORY EMR | New users of Trimethoprim systemetic nested in Urinary Tract Infection | Sudden Cardiac arrest or cardiac death | 216.0 |
| France DA | AMBULATORY EMR | New users of Fluoroquinolone systemic nested in Urinary Tract Infection | Diarrhea events | 0.0 |
| France DA | AMBULATORY EMR | New users of Trimethoprim systemetic nested in Urinary Tract Infection | Diarrhea events | 0.0 |
| France DA | AMBULATORY EMR | New users of Beta blockers nested in essential hypertension | Diarrhea events | 0.0 |

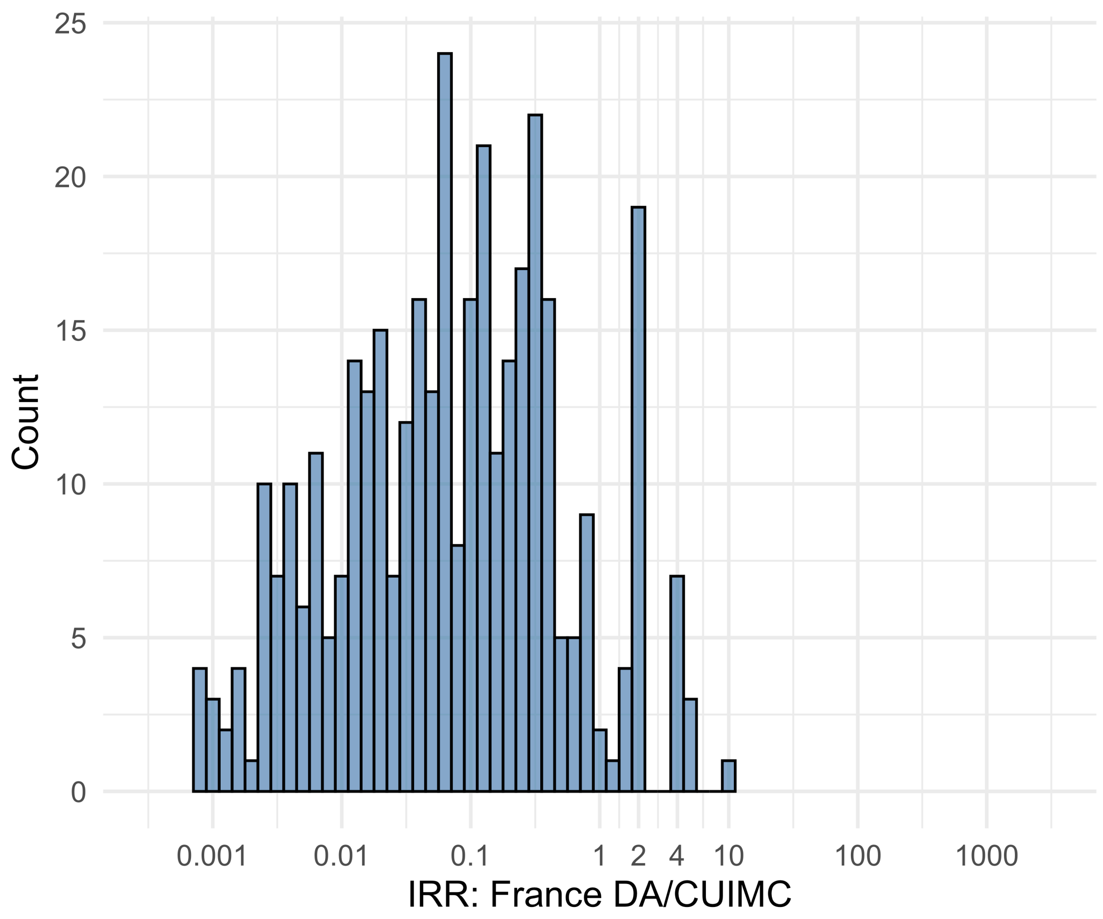


Figure S5.26. IRR distribution comparing France DA and CUIMC.

*Table S5.26. Most extreme IRRs for France DA vs CUIMC (rounded to 0.001).*

| source1 | source2 | Drug and Indication | Outcome | IRR |
|---|---|---|---|---|
| France DA | CUIMC | New users of Beta blockers nested in essential hypertension | Polymorphic Ventricular Tachycardia or Torsades de Pointes | 10.4 |
| France DA | CUIMC | New users of GLP-1 receptor antagonists nested in Type 2 diabetes mellitus | Progressive multifocal leukoencephalopathy | 4.8 |
| France DA | CUIMC | New users of GLP-1 receptor antagonists nested in Type 2 diabetes mellitus | Polymorphic Ventricular Tachycardia or Torsades de Pointes | 4.8 |
| France DA | CUIMC | New users of Beta blockers nested in essential hypertension | Acute Kidney Injury AKI | 0.0 |
| France DA | CUIMC | New users of Fluoroquinolone systemic nested in Urinary Tract Infection | Acute Kidney Injury AKI | 0.0 |
| France DA | CUIMC | New users of Beta blockers nested in essential hypertension | 4-point MACE | 0.0 |

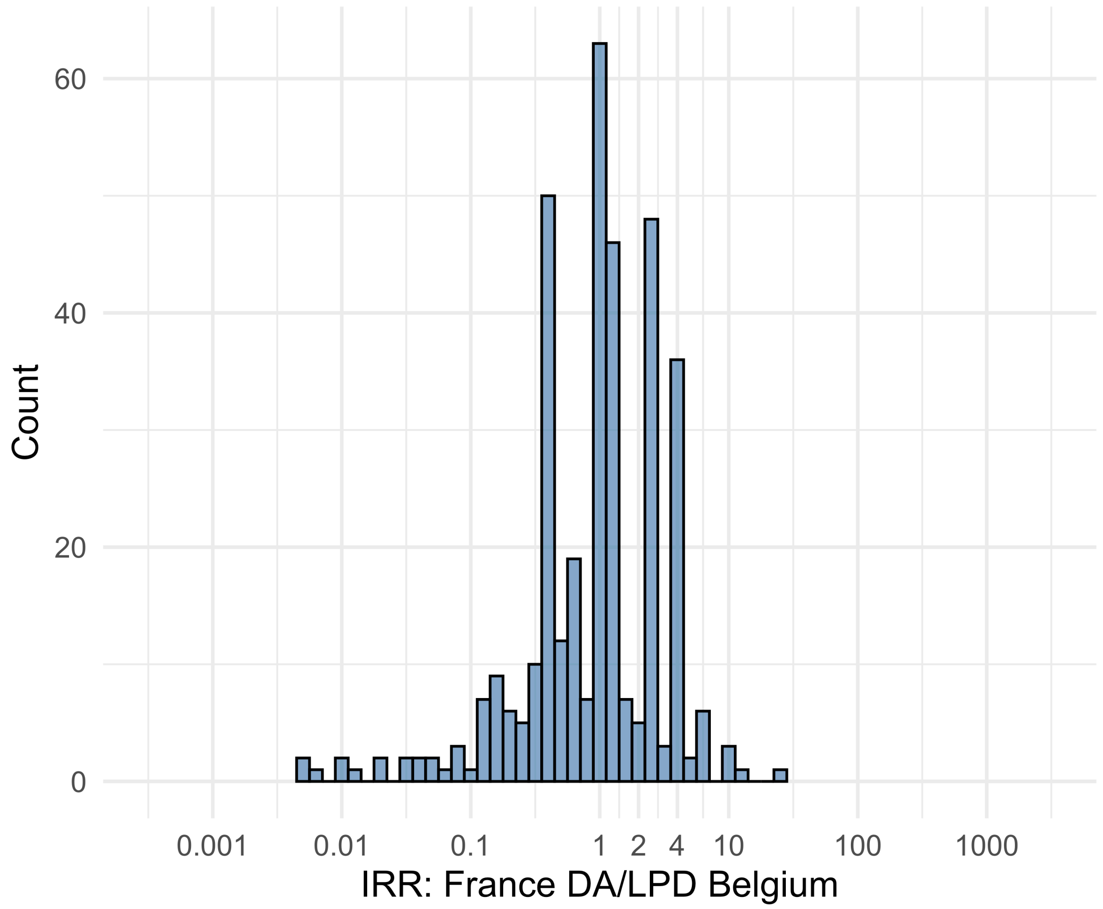


Figure S5.27. IRR distribution comparing France DA and LPD Belgium.

*Table S5.27. Most extreme IRRs for France DA vs LPD Belgium (rounded to 0.001).*

| source1 | source2 | Drug and Indication | Outcome | IRR |
|---|---|---|---|---|
| France DA | LPD Belgium | New users of Fluoroquinolone systemic nested in Urinary Tract Infection | Bladder Cancer | 26.6 |
| France DA | LPD Belgium | New users of Trimethoprim systemetic nested in Urinary Tract Infection | Abnormal weight loss events | 13.1 |
| France DA | LPD Belgium | New users of Beta blockers nested in essential hypertension | Deep Vein Thrombosis (DVT) | 9.5 |
| France DA | LPD Belgium | New users of Fluoroquinolone systemic nested in Urinary Tract Infection | Diarrhea events | 0.0 |
| France DA | LPD Belgium | New users of Trimethoprim systemetic nested in Urinary Tract Infection | Diarrhea events | 0.0 |
| France DA | LPD Belgium | New users of Beta blockers nested in essential hypertension | Diarrhea events | 0.0 |

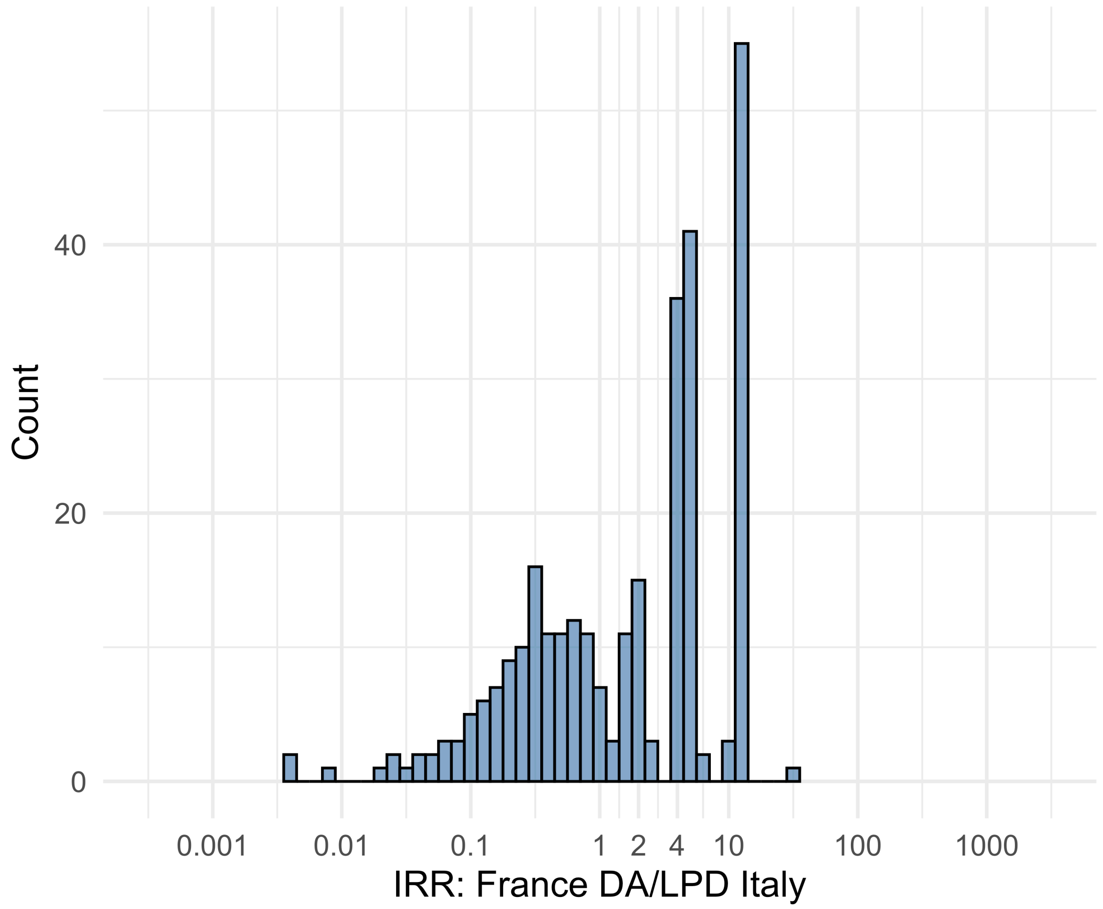


Figure S5.28. IRR distribution comparing France DA and LPD Italy.

*Table S5.28. Most extreme IRRs for France DA vs LPD Italy (rounded to 0.001).*

| source1 | source2 | Drug and Indication | Outcome | IRR |
|---|---|---|---|---|
| France DA | LPD Italy | New users of Beta blockers nested in essential hypertension | All events of Autoimmune hepatitis, with a washout period of 365 days | 31.0 |
| France DA | LPD Italy | New users of Beta blockers nested in essential hypertension | Earliest event of Acute Hepatic Failure, NO viral hepatitis or alcoholic hepatic failure | 12.7 |
| France DA | LPD Italy | New users of Beta blockers nested in essential hypertension | Earliest event of Acute Hepatic Failure | 12.7 |
| France DA | LPD Italy | New users of Beta blockers nested in essential hypertension | Diarrhea events | 0.0 |
| France DA | LPD Italy | New users of Fluoroquinolone systemic nested in Urinary Tract Infection | Diarrhea events | 0.0 |
| France DA | LPD Italy | New users of GLP-1 receptor antagonists nested in Type 2 diabetes mellitus | Diarrhea events | 0.0 |

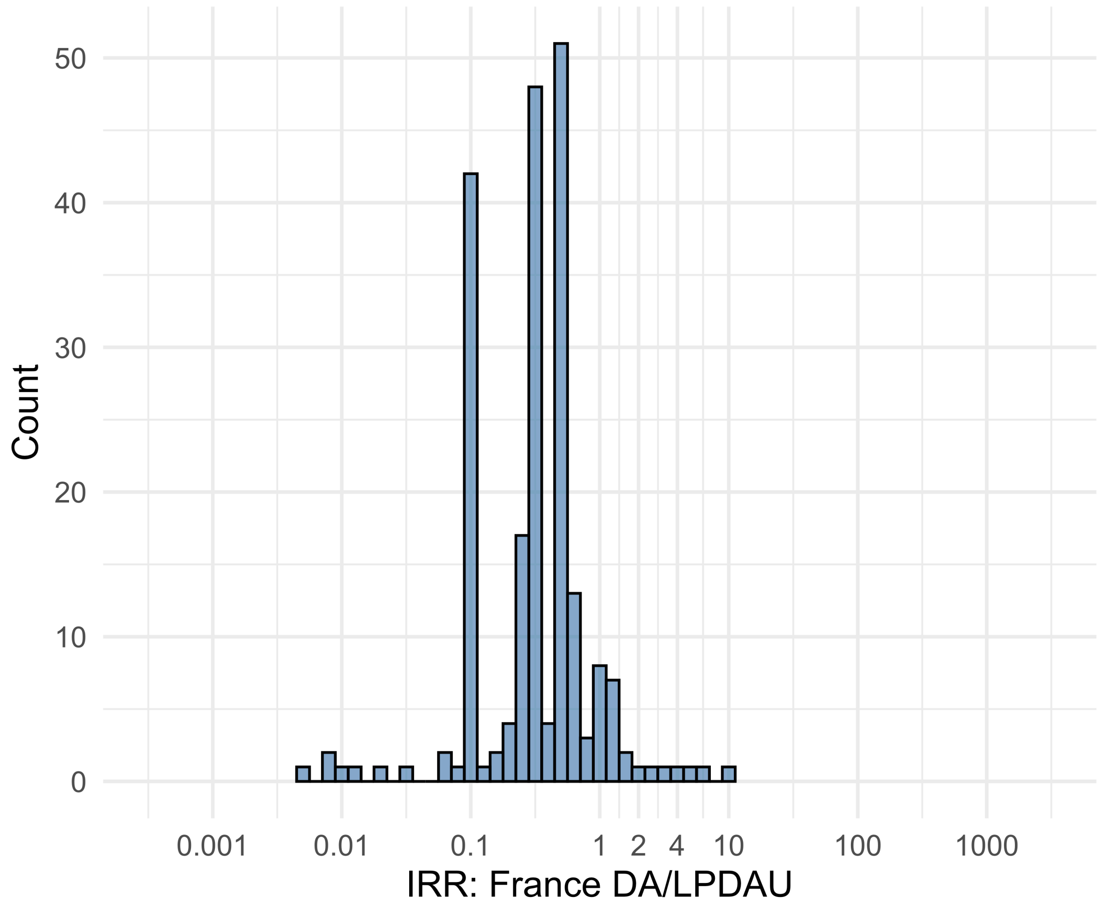


Figure S5.29. IRR distribution comparing France DA and LPDAU.

*Table S5.29. Most extreme IRRs for France DA vs LPDAU (rounded to 0.001).*

| source1 | source2 | Drug and Indication | Outcome | IRR |
|---|---|---|---|---|
| France DA | LPDAU | New users of Trimethoprim systemetic nested in Urinary Tract Infection | Abnormal weight loss events | 9.9 |
| France DA | LPDAU | New users of Trimethoprim systemetic nested in Urinary Tract Infection | Bladder Cancer | 6.9 |
| France DA | LPDAU | New users of Beta blockers nested in essential hypertension | Abnormal weight loss events | 5.0 |
| France DA | LPDAU | New users of Beta blockers nested in essential hypertension | Fall events | 0.0 |
| France DA | LPDAU | New users of Beta blockers nested in essential hypertension | Diarrhea events | 0.0 |
| France DA | LPDAU | New users of Trimethoprim systemetic nested in Urinary Tract Infection | Diarrhea events | 0.0 |

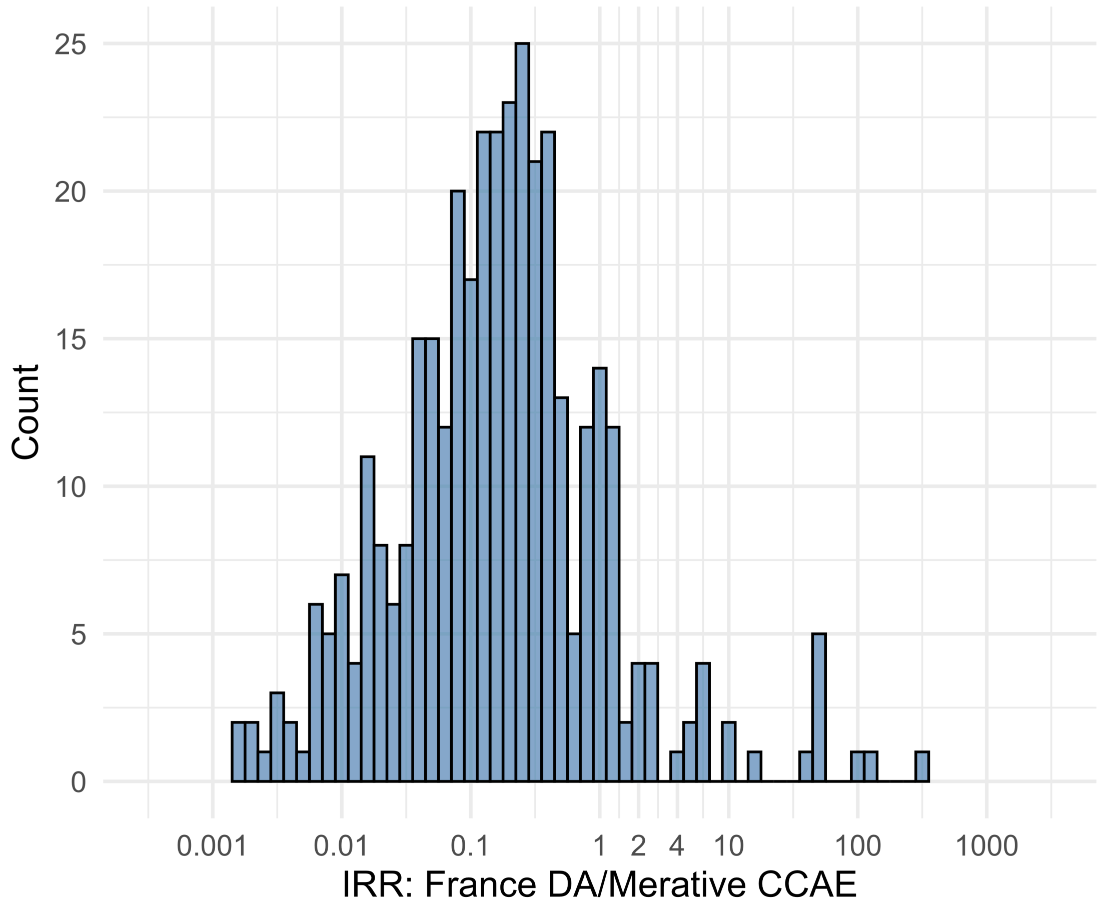


Figure S5.30. IRR distribution comparing France DA and Merative CCAE.

*Table S5.30. Most extreme IRRs for France DA vs Merative CCAE (rounded to 0.001).*

| source1 | source2 | Drug and Indication | Outcome | IRR |
|---|---|---|---|---|
| France DA | Merative CCAE | New users of Trimethoprim systemetic nested in Urinary Tract Infection | Polymorphic Ventricular Tachycardia or Torsades de Pointes | 309.3 |
| France DA | Merative CCAE | New users of Fluoroquinolone systemic nested in Urinary Tract Infection | Polymorphic Ventricular Tachycardia or Torsades de Pointes | 131.8 |
| France DA | Merative CCAE | New users of Cephalosporin systemetic nested in Urinary Tract Infection | Polymorphic Ventricular Tachycardia or Torsades de Pointes | 89.8 |
| France DA | Merative CCAE | New users of Beta blockers nested in essential hypertension | Hospitalization with heart failure events | 0.0 |
| France DA | Merative CCAE | New users of Beta blockers nested in essential hypertension | Acute Kidney Injury AKI | 0.0 |
| France DA | Merative CCAE | New users of Beta blockers nested in essential hypertension | 4-point MACE | 0.0 |

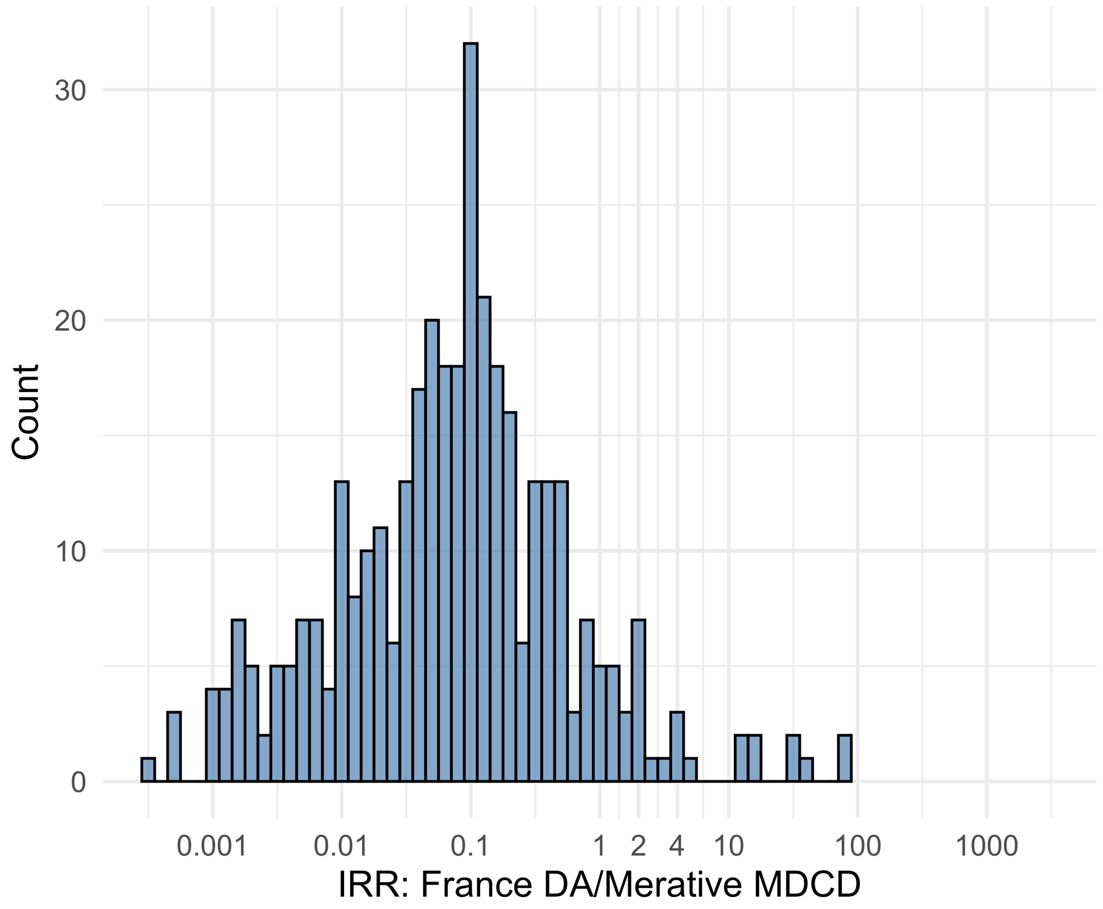


Figure S5.31. IRR distribution comparing France DA and Merative MDCD.

*Table S5.31. Most extreme IRRs for France DA vs Merative MDCD (rounded to 0.001).*

| source1 | source2 | Drug and Indication | Outcome | IRR |
|---|---|---|---|---|
| France DA | Merative MDCD | New users of Trimethoprim systemetic nested in Urinary Tract Infection | Polymorphic Ventricular Tachycardia or Torsades de Pointes | 73.7 |
| France DA | Merative MDCD | New users of Trimethoprim systemetic nested in Urinary Tract Infection | Progressive multifocal leukoencephalopathy | 73.7 |
| France DA | Merative MDCD | New users of Fluoroquinolone systemic nested in Urinary Tract Infection | Polymorphic Ventricular Tachycardia or Torsades de Pointes | 40.8 |
| France DA | Merative MDCD | New users of Beta blockers nested in essential hypertension | Hospitalization with heart failure events | <0.001 |
| France DA | Merative MDCD | New users of Beta blockers nested in essential hypertension | Acute Kidney Injury AKI | <0.001 |
| France DA | Merative MDCD | New users of Beta blockers nested in essential hypertension | 4-point MACE | 0.0 |

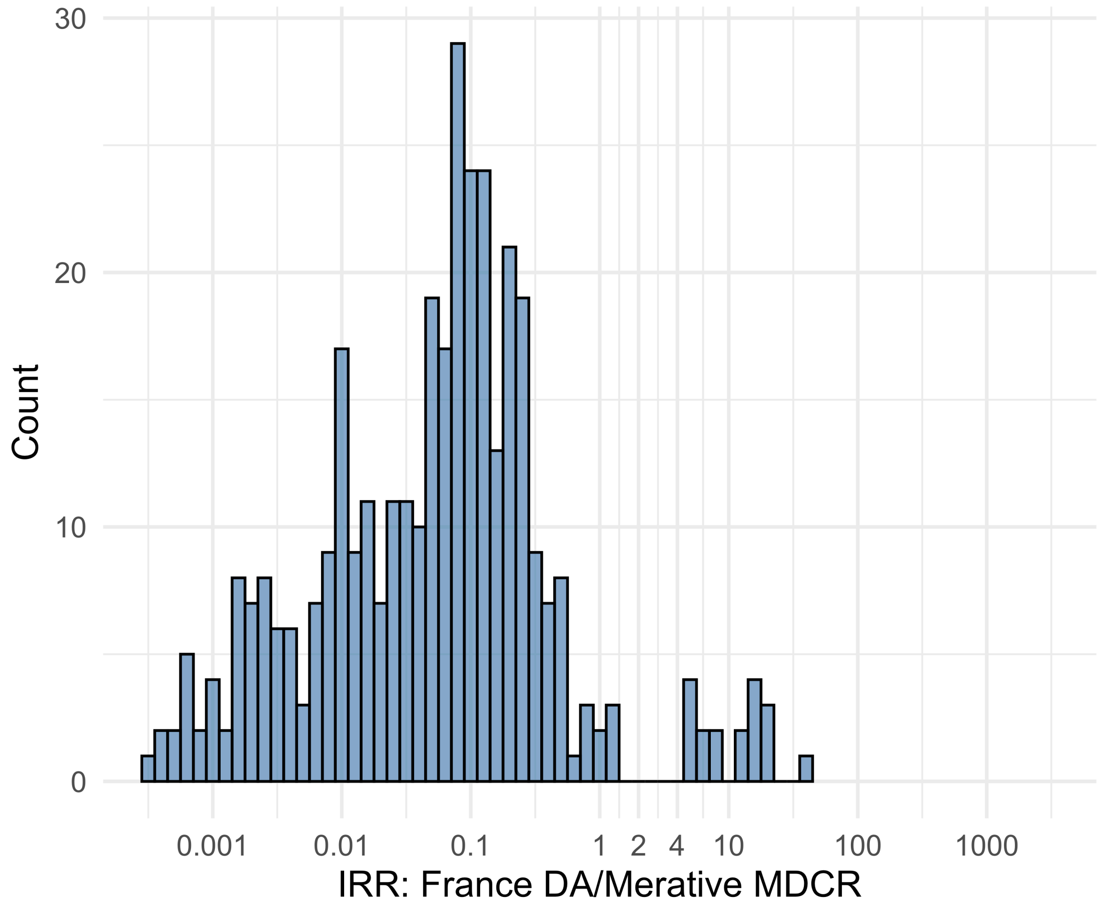


Figure S5.32. IRR distribution comparing France DA and Merative MDCR.

*Table S5.32. Most extreme IRRs for France DA vs Merative MDCR (rounded to 0.001).*

| source1 | source2 | Drug and Indication | Outcome | IRR |
|---|---|---|---|---|
| France DA | Merative MDCR | New users of Trimethoprim systemetic nested in Urinary Tract Infection | Polymorphic Ventricular Tachycardia or Torsades de Pointes | 38.0 |
| France DA | Merative MDCR | New users of Trimethoprim systemetic nested in Urinary Tract Infection | Progressive multifocal leukoencephalopathy | 19.0 |
| France DA | Merative MDCR | New users of Trimethoprim systemetic nested in Urinary Tract Infection | All events Drug Rash with Eosinophilia and Systemic Symptoms (DRESS) | 19.0 |
| France DA | Merative MDCR | New users of Beta blockers nested in essential hypertension | Hospitalization with heart failure events | <0.001 |
| France DA | Merative MDCR | New users of Beta blockers nested in essential hypertension | 4-point MACE | <0.001 |
| France DA | Merative MDCR | New users of Beta blockers nested in essential hypertension | Total cardiovascular disease events (ischemic stroke, hemorrhagic stroke, heart failure, acute myocardial infarction or sudden cardiac death) | <0.001 |

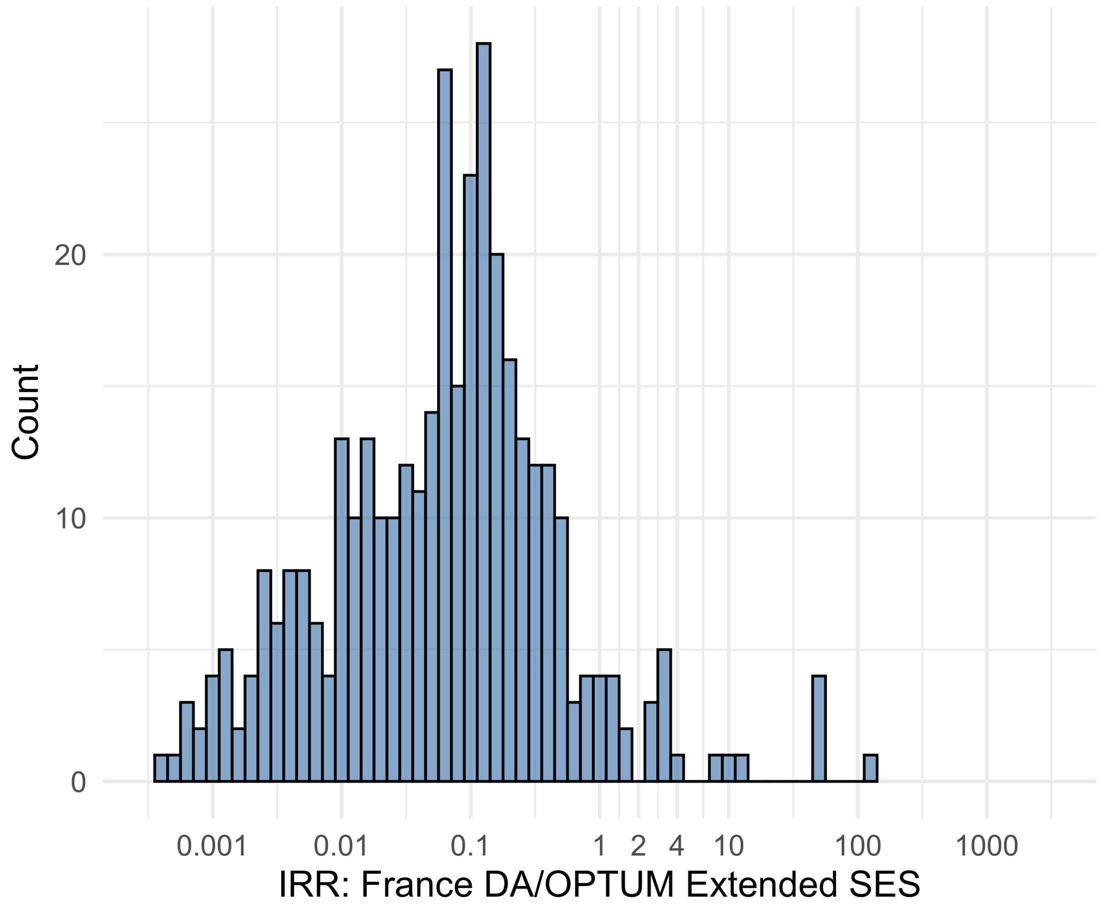


Figure S5.33. IRR distribution comparing France DA and OPTUM Extended SES.

*Table S5.33. Most extreme IRRs for France DA vs OPTUM Extended SES (rounded to 0.001).*

| source1 | source2 | Drug and Indication | Outcome | IRR |
|---|---|---|---|---|
| France DA | OPTUM Extended SES | New users of Trimethoprim systemetic nested in Urinary Tract Infection | Polymorphic Ventricular Tachycardia or Torsades de Pointes | 117.9 |
| France DA | OPTUM Extended SES | New users of Fluoroquinolone systemic nested in Urinary Tract Infection | Polymorphic Ventricular Tachycardia or Torsades de Pointes | 44.9 |
| France DA | OPTUM Extended SES | New users of GLP-1 receptor antagonists nested in Type 2 diabetes mellitus | Polymorphic Ventricular Tachycardia or Torsades de Pointes | 44.7 |
| France DA | OPTUM Extended SES | New users of Beta blockers nested in essential hypertension | Hospitalization with heart failure events | <0.001 |
| France DA | OPTUM Extended SES | New users of Beta blockers nested in essential hypertension | Acute Kidney Injury AKI | 0.0 |
| France DA | OPTUM Extended SES | New users of Beta blockers nested in essential hypertension | 4-point MACE | 0.0 |

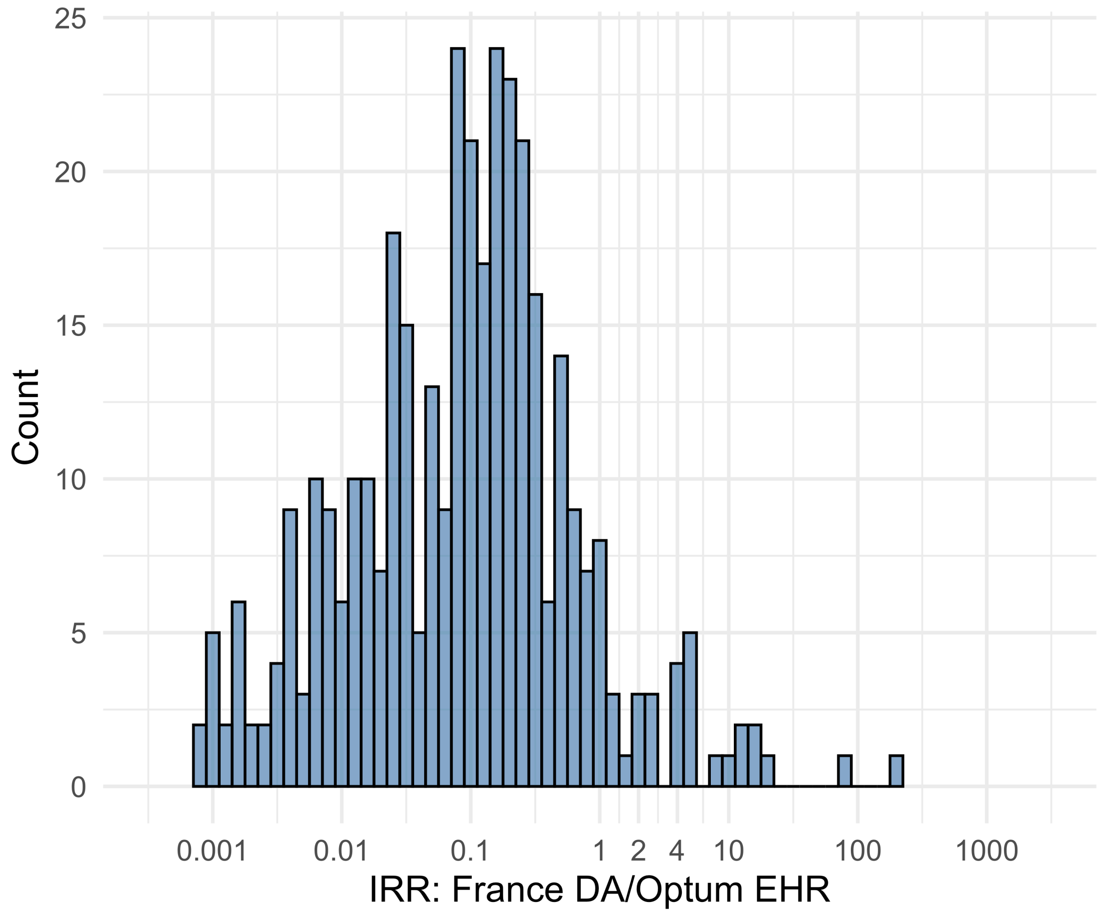


Figure S5.34. IRR distribution comparing France DA and Optum EHR.

*Table S5.34. Most extreme IRRs for France DA vs Optum EHR (rounded to 0.001).*

| source1 | source2 | Drug and Indication | Outcome | IRR |
|---|---|---|---|---|
| France DA | Optum EHR | New users of Trimethoprim systemetic nested in Urinary Tract Infection | Polymorphic Ventricular Tachycardia or Torsades de Pointes | 203.8 |
| France DA | Optum EHR | New users of GLP-1 receptor antagonists nested in Type 2 diabetes mellitus | Progressive multifocal leukoencephalopathy | 82.4 |
| France DA | Optum EHR | New users of Trimethoprim systemetic nested in Urinary Tract Infection | All events Drug Rash with Eosinophilia and Systemic Symptoms (DRESS) | 18.5 |
| France DA | Optum EHR | New users of Beta blockers nested in essential hypertension | Hospitalization with heart failure events | 0.0 |
| France DA | Optum EHR | New users of Beta blockers nested in essential hypertension | Acute Kidney Injury AKI | 0.0 |
| France DA | Optum EHR | New users of Beta blockers nested in essential hypertension | 4-point MACE | 0.0 |

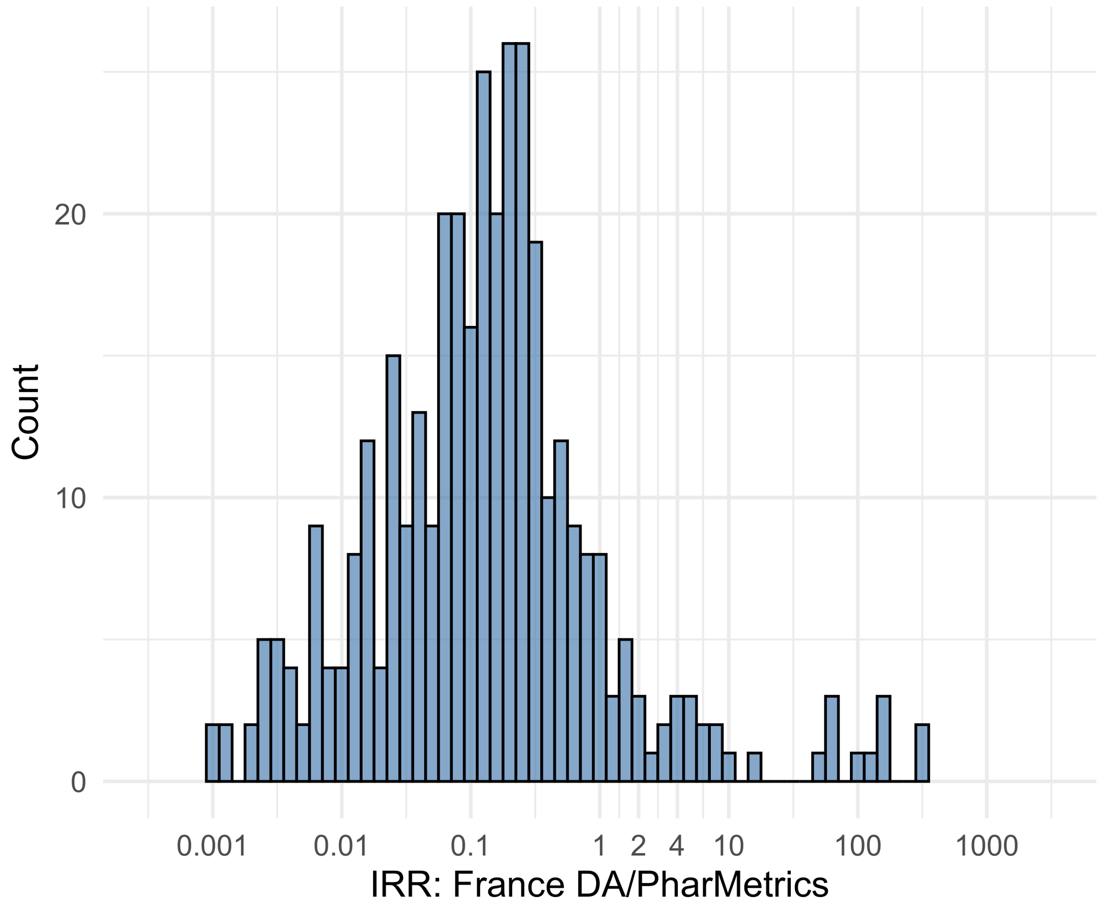


Figure S5.35. IRR distribution comparing France DA and PharMetrics.

*Table S5.35. Most extreme IRRs for France DA vs PharMetrics (rounded to 0.001).*

| source1 | source2 | Drug and Indication | Outcome | IRR |
|---|---|---|---|---|
| France DA | PharMetrics | New users of Trimethoprim systemetic nested in Urinary Tract Infection | Cardiovascular-related mortality | 284.6 |
| France DA | PharMetrics | New users of Trimethoprim systemetic nested in Urinary Tract Infection | Polymorphic Ventricular Tachycardia or Torsades de Pointes | 284.6 |
| France DA | PharMetrics | New users of Beta blockers nested in essential hypertension | Cardiovascular-related mortality | 168.1 |
| France DA | PharMetrics | New users of Beta blockers nested in essential hypertension | Hospitalization with heart failure events | 0.0 |
| France DA | PharMetrics | New users of Beta blockers nested in essential hypertension | Acute Kidney Injury AKI | 0.0 |
| France DA | PharMetrics | New users of Beta blockers nested in essential hypertension | 4-point MACE | 0.0 |

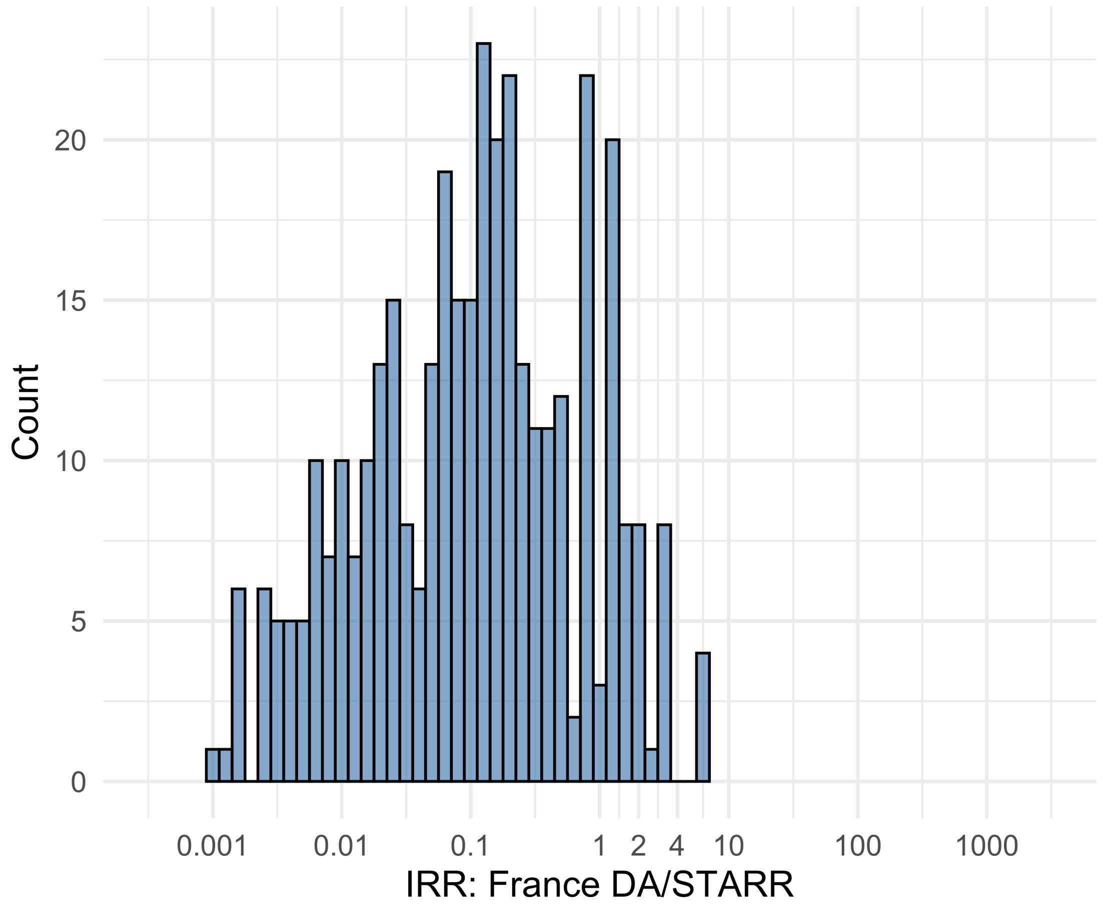


Figure S5.36. IRR distribution comparing France DA and STARR.

*Table S5.36. Most extreme IRRs for France DA vs STARR (rounded to 0.001).*

| source1 | source2 | Drug and Indication | Outcome | IRR |
|---|---|---|---|---|
| France DA | STARR | New users of Beta blockers nested in essential hypertension | Febrile Neutropenia or Neutropenic Fever | 7.0 |
| France DA | STARR | New users of Beta blockers nested in essential hypertension | Polymorphic Ventricular Tachycardia or Torsades de Pointes | 7.0 |
| France DA | STARR | New users of Beta blockers nested in essential hypertension | All events Drug Rash with Eosinophilia and Systemic Symptoms (DRESS) | 7.0 |
| France DA | STARR | New users of Beta blockers nested in essential hypertension | Hospitalization with heart failure events | 0.0 |
| France DA | STARR | New users of Cephalosporin systemetic nested in Urinary Tract Infection | Hospitalization with heart failure events | 0.0 |
| France DA | STARR | New users of Beta blockers nested in essential hypertension | 4-point MACE | 0.0 |

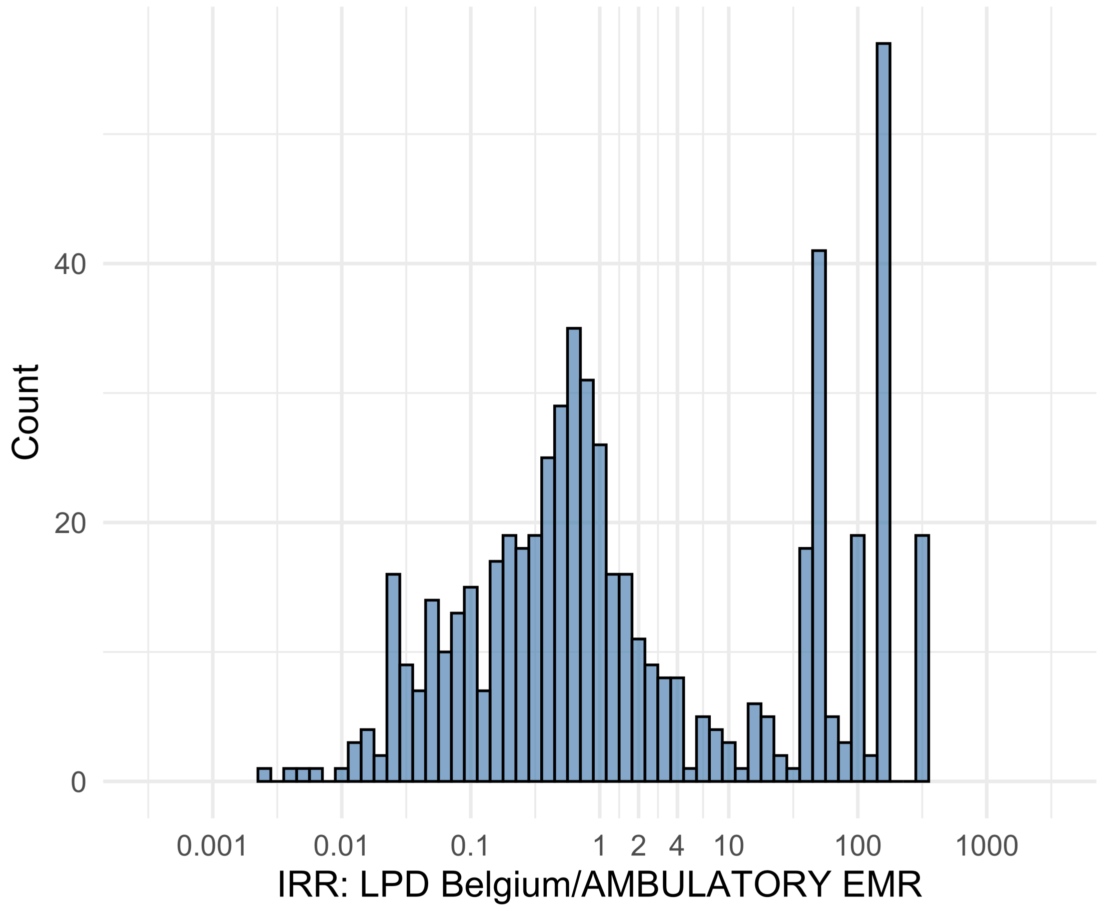


Figure S5.37. IRR distribution comparing LPD Belgium and AMBULATORY EMR.

*Table S5.37. Most extreme IRRs for LPD Belgium vs AMBULATORY EMR (rounded to 0.001).*

| source1 | source2 | Drug and Indication | Outcome | IRR |
|---|---|---|---|---|
| LPD Belgium | AMBULATORY EMR | New users of GLP-1 receptor antagonists nested in obesity | Febrile Neutropenia or Neutropenic Fever | 321.3 |
| LPD Belgium | AMBULATORY EMR | New users of GLP-1 receptor antagonists nested in obesity | Rhabdomyolysis | 321.3 |
| LPD Belgium | AMBULATORY EMR | New users of GLP-1 receptor antagonists nested in obesity | Sudden Cardiac arrest or cardiac death | 321.3 |
| LPD Belgium | AMBULATORY EMR | New users of Beta blockers nested in essential hypertension | Deep Vein Thrombosis (DVT) | 0.0 |
| LPD Belgium | AMBULATORY EMR | New users of Beta blockers nested in essential hypertension | Anaphylaxis | 0.0 |
| LPD Belgium | AMBULATORY EMR | New users of Fluoroquinolone systemic nested in Urinary Tract Infection | Anaphylaxis | 0.0 |

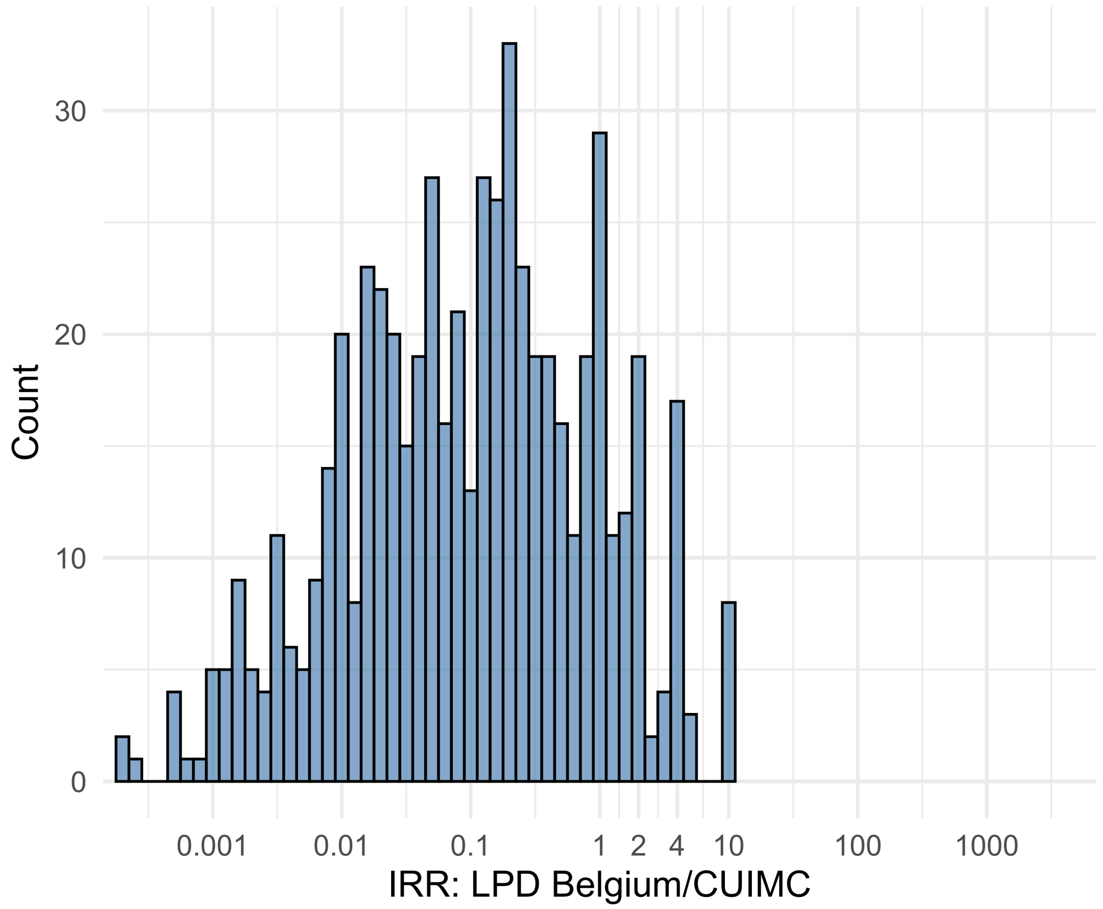


Figure S5.38. IRR distribution comparing LPD Belgium and CUIMC.

*Table S5.38. Most extreme IRRs for LPD Belgium vs CUIMC (rounded to 0.001).*

| source1 | source2 | Drug and Indication | Outcome | IRR |
|---|---|---|---|---|
| LPD Belgium | CUIMC | New users of GLP-1 receptor antagonists nested in obesity | Polymorphic Ventricular Tachycardia or Torsades de Pointes | 10.9 |
| LPD Belgium | CUIMC | New users of GLP-1 receptor antagonists nested in obesity | Progressive multifocal leukoencephalopathy | 10.9 |
| LPD Belgium | CUIMC | New users of GLP-1 receptor antagonists nested in obesity | Stevens-Johnson syndrome, toxic epidermal necrolysis spectrum | 10.9 |
| LPD Belgium | CUIMC | New users of Beta blockers nested in essential hypertension | 4-point MACE | <0.001 |
| LPD Belgium | CUIMC | New users of Beta blockers nested in essential hypertension | Total cardiovascular disease events (ischemic stroke, hemorrhagic stroke, heart failure, acute myocardial infarction or sudden cardiac death) | <0.001 |
| LPD Belgium | CUIMC | New users of Beta blockers nested in essential hypertension | Hospitalization with heart failure events | <0.001 |

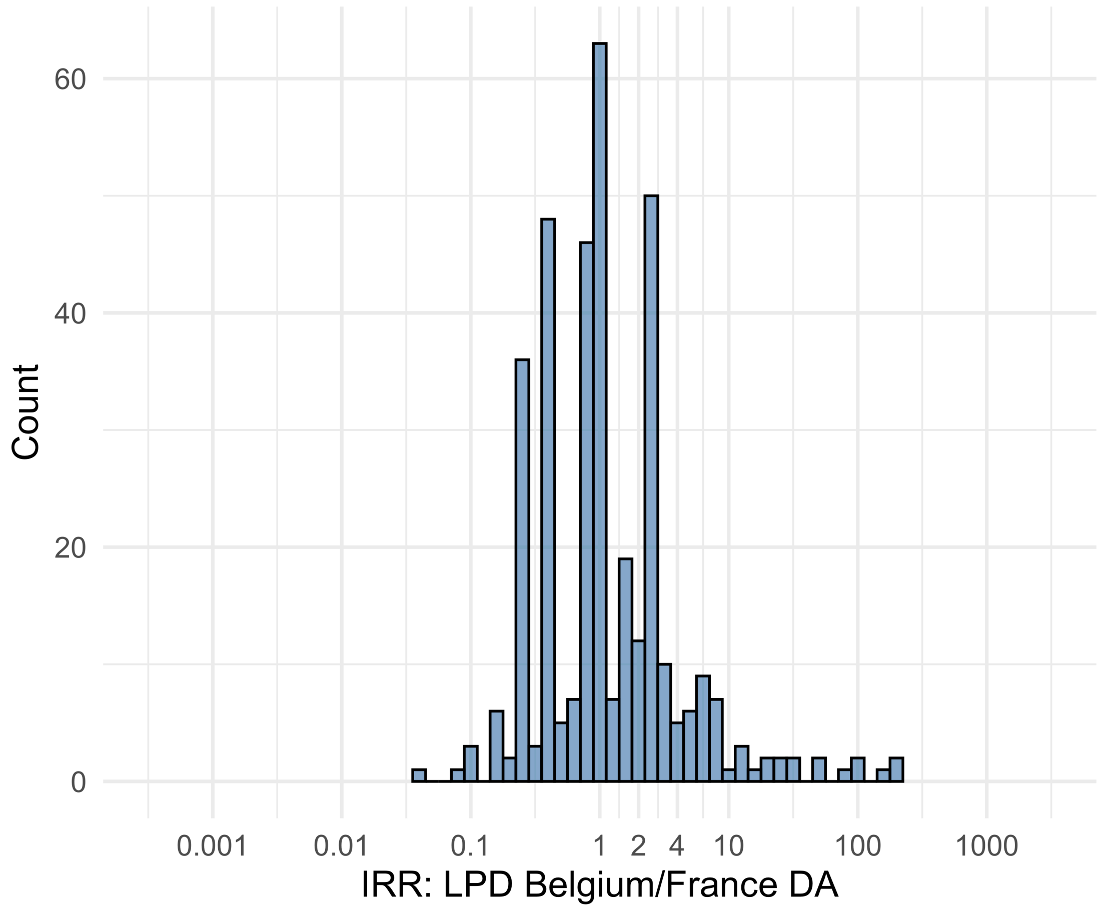


Figure S5.39. IRR distribution comparing LPD Belgium and France DA.

*Table S5.39. Most extreme IRRs for LPD Belgium vs France DA (rounded to 0.001).*

| source1 | source2 | Drug and Indication | Outcome | IRR |
|---|---|---|---|---|
| LPD Belgium | France DA | New users of Fluoroquinolone systemic nested in Urinary Tract Infection | Diarrhea events | 181.9 |
| LPD Belgium | France DA | New users of Trimethoprim systemetic nested in Urinary Tract Infection | Diarrhea events | 180.9 |
| LPD Belgium | France DA | New users of Beta blockers nested in essential hypertension | Diarrhea events | 161.7 |
| LPD Belgium | France DA | New users of Fluoroquinolone systemic nested in Urinary Tract Infection | Bladder Cancer | 0.0 |
| LPD Belgium | France DA | New users of Trimethoprim systemetic nested in Urinary Tract Infection | Abnormal weight loss events | 0.1 |
| LPD Belgium | France DA | New users of Beta blockers nested in essential hypertension | Deep Vein Thrombosis (DVT) | 0.1 |

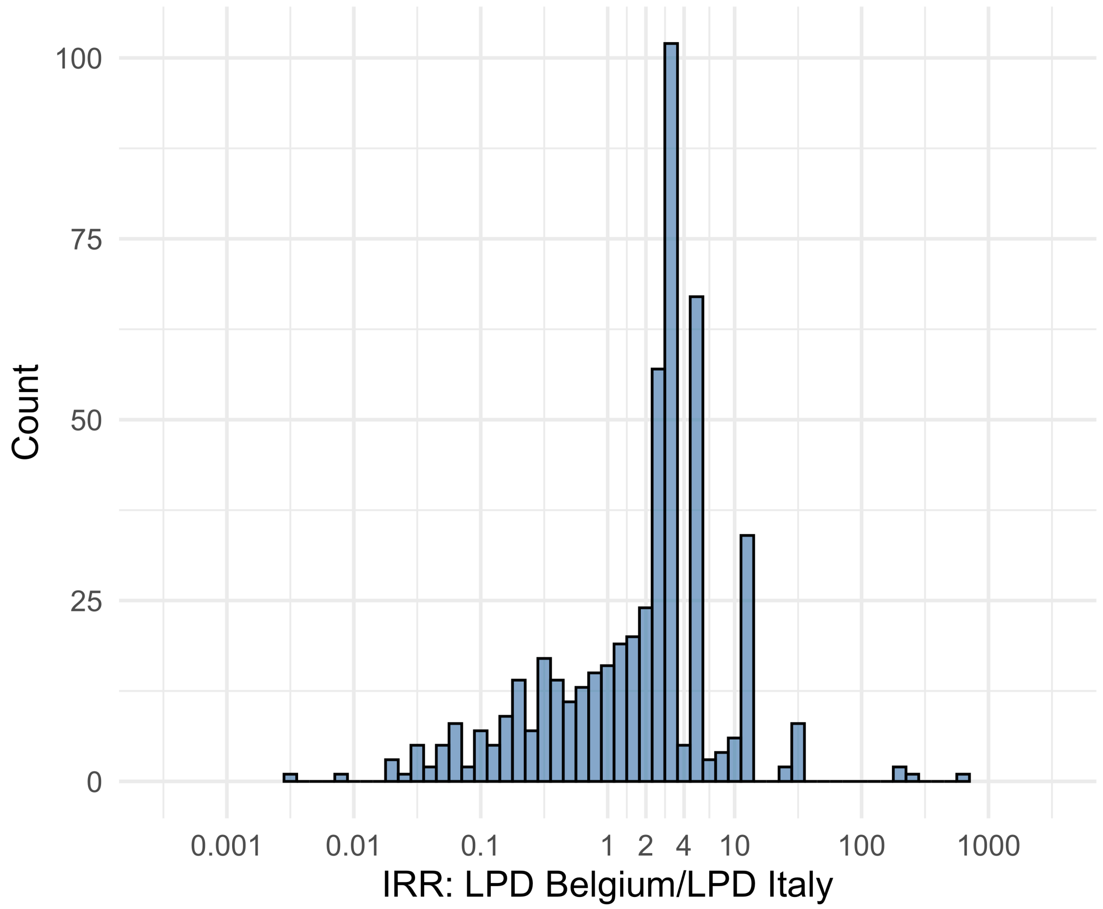


Figure S5.40. IRR distribution comparing LPD Belgium and LPD Italy.

*Table S5.40. Most extreme IRRs for LPD Belgium vs LPD Italy (rounded to 0.001).*

| source1 | source2 | Drug and Indication | Outcome | IRR |
|---|---|---|---|---|
| LPD Belgium | LPD Italy | New users of GLP-1 receptor antagonists nested in obesity | Abnormal weight gain events | 594.1 |
| LPD Belgium | LPD Italy | New users of Beta blockers nested in essential hypertension | Fall events | 240.8 |
| LPD Belgium | LPD Italy | New users of GLP-1 receptor antagonists nested in Type 2 diabetes mellitus | Abnormal weight gain events | 200.9 |
| LPD Belgium | LPD Italy | New users of Beta blockers nested in essential hypertension | Deep Vein Thrombosis (DVT) | 0.0 |
| LPD Belgium | LPD Italy | New users of Fluoroquinolone systemic nested in Urinary Tract Infection | Deep Vein Thrombosis (DVT) | 0.0 |
| LPD Belgium | LPD Italy | New users of Beta blockers nested in essential hypertension | Bladder Cancer | 0.0 |

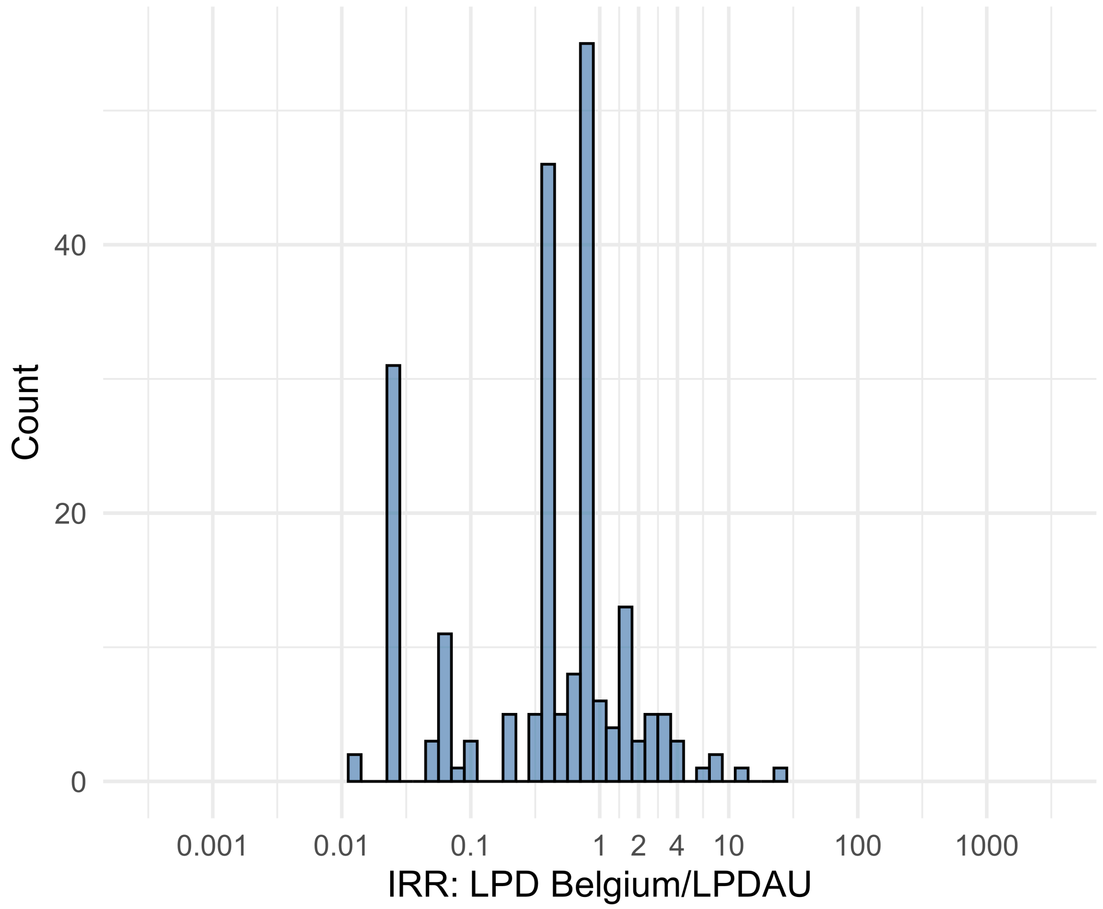


Figure S5.41. IRR distribution comparing LPD Belgium and LPDAU.

*Table S5.41. Most extreme IRRs for LPD Belgium vs LPDAU (rounded to 0.001).*

| source1 | source2 | Drug and Indication | Outcome | IRR |
|---|---|---|---|---|
| LPD Belgium | LPDAU | New users of Cephalosporin systemetic nested in Urinary Tract Infection | Acute Kidney Injury AKI | 25.8 |
| LPD Belgium | LPDAU | New users of Trimethoprim systemetic nested in Urinary Tract Infection | Acute Kidney Injury AKI | 12.1 |
| LPD Belgium | LPDAU | New users of Beta blockers nested in essential hypertension | Acute Kidney Injury AKI | 8.2 |
| LPD Belgium | LPDAU | New users of Beta blockers nested in essential hypertension | Deep Vein Thrombosis (DVT) | 0.0 |
| LPD Belgium | LPDAU | New users of Beta blockers nested in essential hypertension | Anaphylaxis | 0.0 |
| LPD Belgium | LPDAU | New users of Trimethoprim systemetic nested in Urinary Tract Infection | Deep Vein Thrombosis (DVT) | 0.0 |

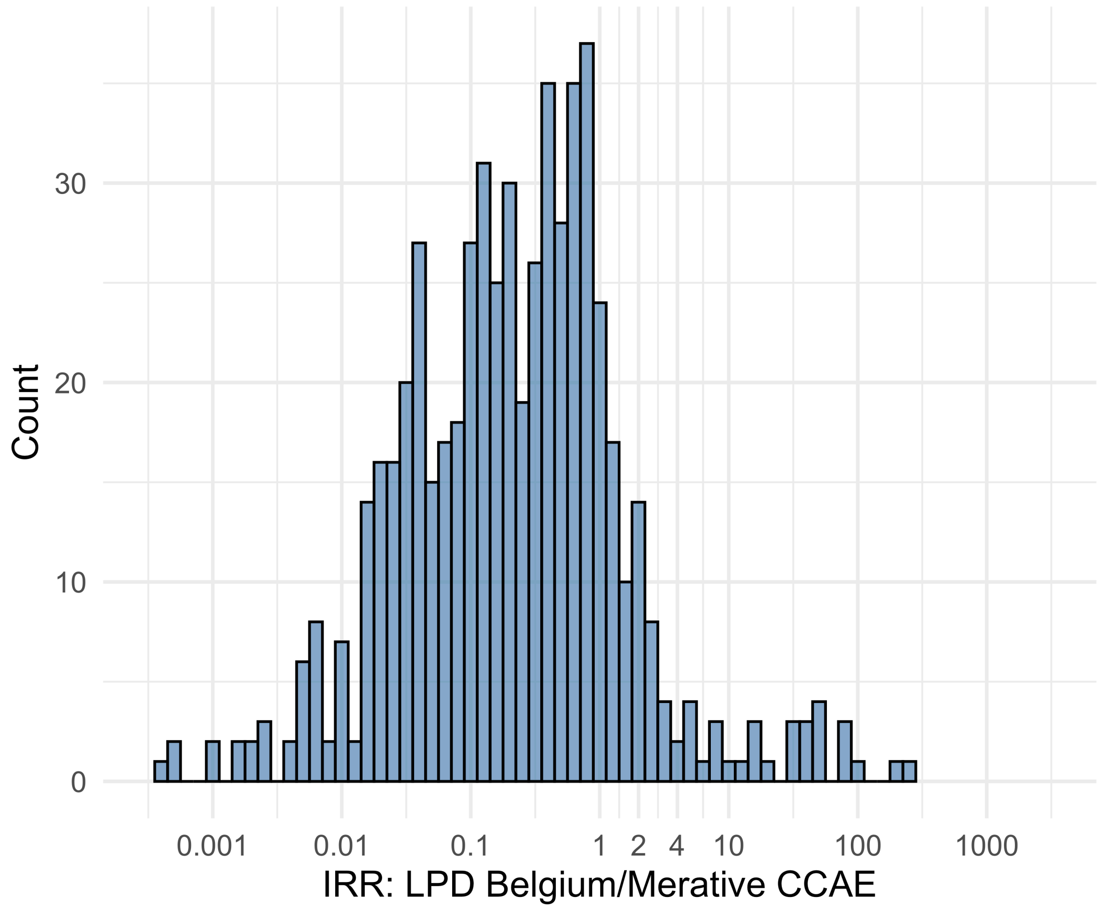


Figure S5.42. IRR distribution comparing LPD Belgium and Merative CCAE.

*Table S5.42. Most extreme IRRs for LPD Belgium vs Merative CCAE (rounded to 0.001).*

| source1 | source2 | Drug and Indication | Outcome | IRR |
|---|---|---|---|---|
| LPD Belgium | Merative CCAE | New users of Trimethoprim systemetic nested in Urinary Tract Infection | Polymorphic Ventricular Tachycardia or Torsades de Pointes | 236.1 |
| LPD Belgium | Merative CCAE | New users of Cephalosporin systemetic nested in Urinary Tract Infection | Polymorphic Ventricular Tachycardia or Torsades de Pointes | 223.3 |
| LPD Belgium | Merative CCAE | New users of Cephalosporin systemetic nested in Urinary Tract Infection | Progressive multifocal leukoencephalopathy | 91.1 |
| LPD Belgium | Merative CCAE | New users of Beta blockers nested in essential hypertension | Hospitalization with heart failure events | <0.001 |
| LPD Belgium | Merative CCAE | New users of Beta blockers nested in essential hypertension | 4-point MACE | <0.001 |
| LPD Belgium | Merative CCAE | New users of Beta blockers nested in essential hypertension | Total cardiovascular disease events (ischemic stroke, hemorrhagic stroke, heart failure, acute myocardial infarction or sudden cardiac death) | <0.001 |

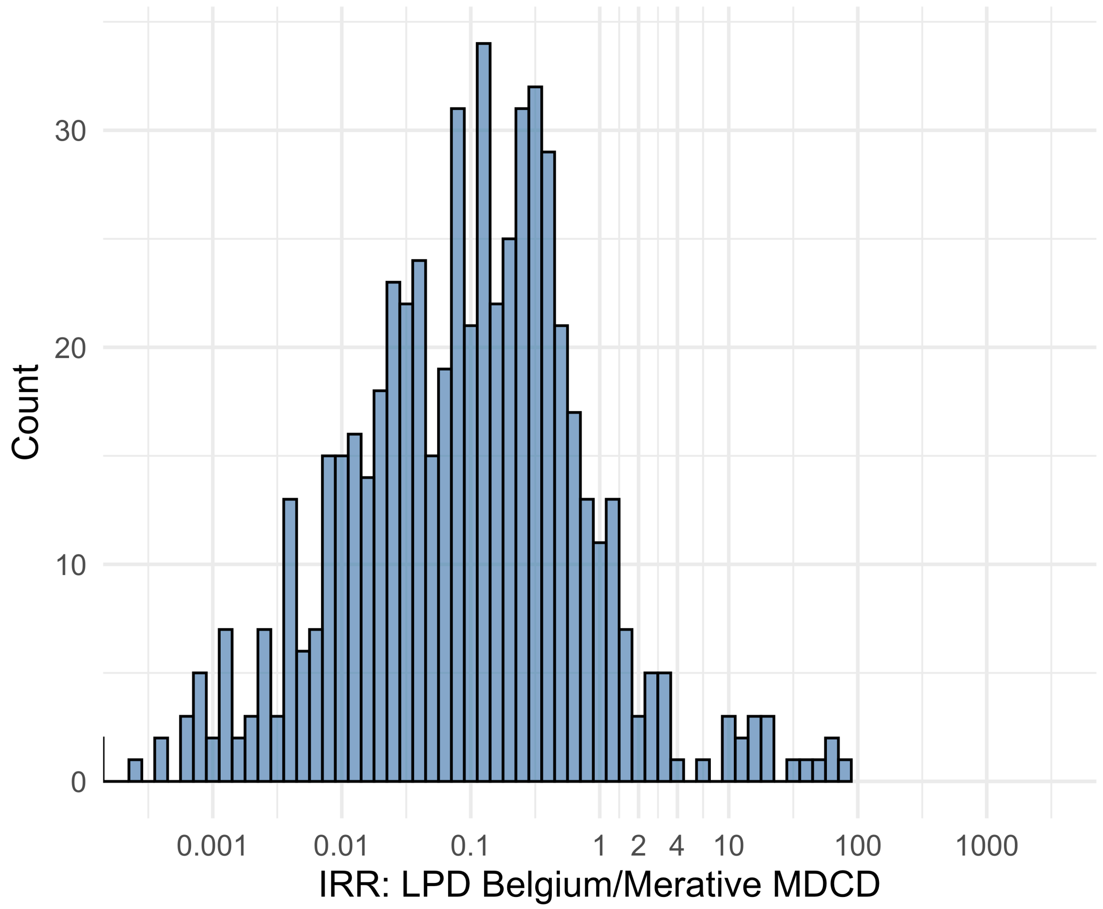


Figure S5.43. IRR distribution comparing LPD Belgium and Merative MDCD.

*Table S5.43. Most extreme IRRs for LPD Belgium vs Merative MDCD (rounded to 0.001).*

| source1 | source2 | Drug and Indication | Outcome | IRR |
|---|---|---|---|---|
| LPD Belgium | Merative MDCD | New users of Cephalosporin systemetic nested in Urinary Tract Infection | Polymorphic Ventricular Tachycardia or Torsades de Pointes | 74.5 |
| LPD Belgium | Merative MDCD | New users of Trimethoprim systemetic nested in Urinary Tract Infection | Polymorphic Ventricular Tachycardia or Torsades de Pointes | 56.3 |
| LPD Belgium | Merative MDCD | New users of Trimethoprim systemetic nested in Urinary Tract Infection | Progressive multifocal leukoencephalopathy | 56.3 |
| LPD Belgium | Merative MDCD | New users of Beta blockers nested in essential hypertension | Hospitalization with heart failure events | <0.001 |
| LPD Belgium | Merative MDCD | New users of Beta blockers nested in essential hypertension | 4-point MACE | <0.001 |
| LPD Belgium | Merative MDCD | New users of Beta blockers nested in essential hypertension | Total cardiovascular disease events (ischemic stroke, hemorrhagic stroke, heart failure, acute myocardial infarction or sudden cardiac death) | <0.001 |

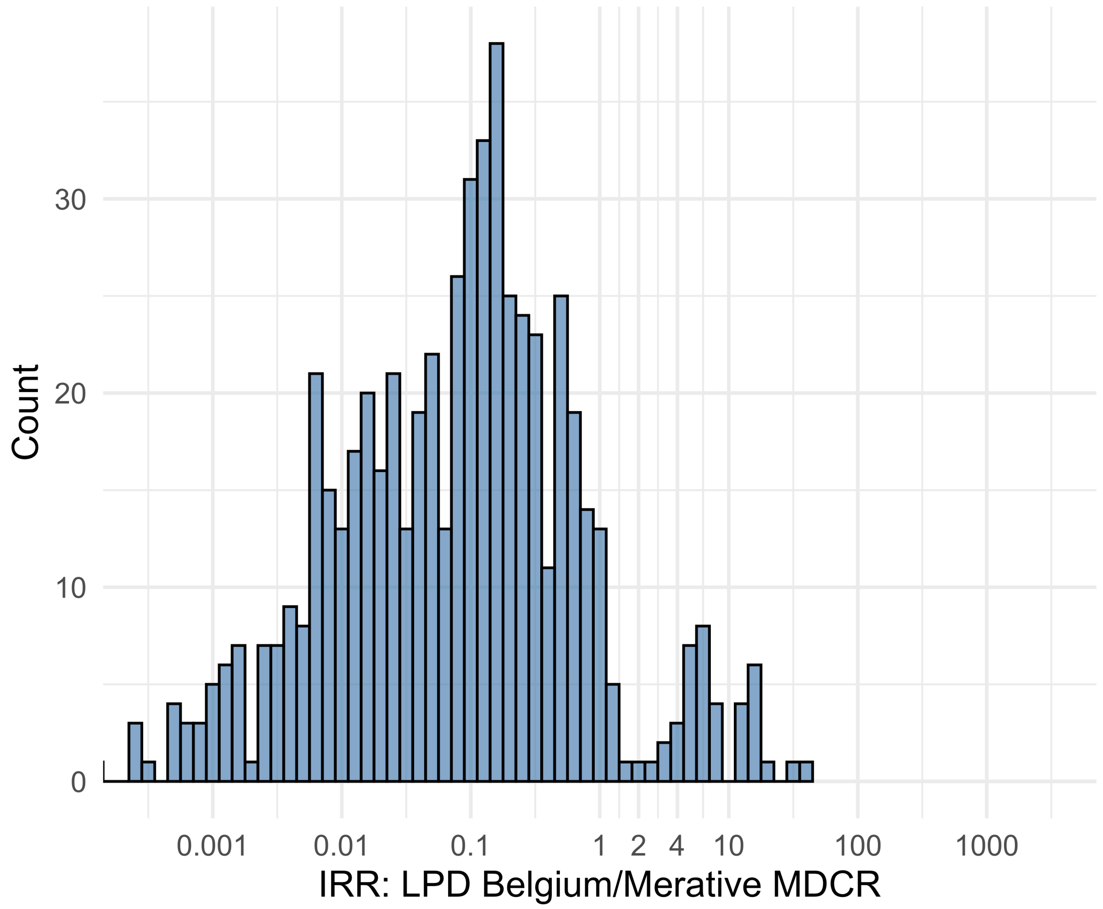


Figure S5.44. IRR distribution comparing LPD Belgium and Merative MDCR.

*Table S5.44. Most extreme IRRs for LPD Belgium vs Merative MDCR (rounded to 0.001).*

| source1 | source2 | Drug and Indication | Outcome | IRR |
|---|---|---|---|---|
| LPD Belgium | Merative MDCR | New users of Cephalosporin systemetic nested in Urinary Tract Infection | Polymorphic Ventricular Tachycardia or Torsades de Pointes | 40.5 |
| LPD Belgium | Merative MDCR | New users of Trimethoprim systemetic nested in Urinary Tract Infection | Polymorphic Ventricular Tachycardia or Torsades de Pointes | 29.0 |
| LPD Belgium | Merative MDCR | New users of Fluoroquinolone systemic nested in Acute Typical Pneumonia | Polymorphic Ventricular Tachycardia or Torsades de Pointes | 20.0 |
| LPD Belgium | Merative MDCR | New users of Beta blockers nested in essential hypertension | Hospitalization with heart failure events | <0.001 |
| LPD Belgium | Merative MDCR | New users of Beta blockers nested in essential hypertension | 4-point MACE | <0.001 |
| LPD Belgium | Merative MDCR | New users of Beta blockers nested in essential hypertension | Total cardiovascular disease events (ischemic stroke, hemorrhagic stroke, heart failure, acute myocardial infarction or sudden cardiac death) | <0.001 |

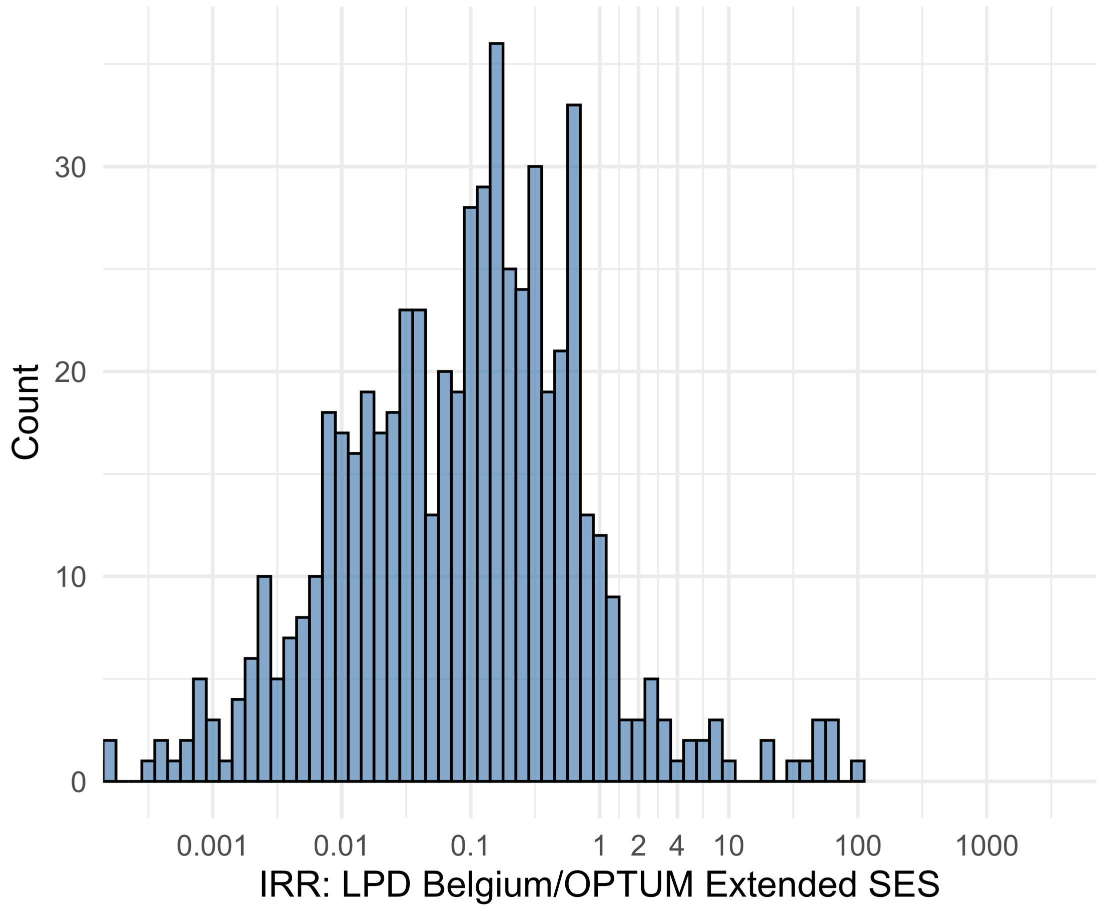


Figure S5.45. IRR distribution comparing LPD Belgium and OPTUM Extended SES.

*Table S5.45. Most extreme IRRs for LPD Belgium vs OPTUM Extended SES (rounded to 0.001).*

| source1 | source2 | Drug and Indication | Outcome | IRR |
|---|---|---|---|---|
| LPD Belgium | OPTUM Extended SES | New users of Trimethoprim systemetic nested in Urinary Tract Infection | Polymorphic Ventricular Tachycardia or Torsades de Pointes | 90.0 |
| LPD Belgium | OPTUM Extended SES | New users of GLP-1 receptor antagonists nested in obesity | Polymorphic Ventricular Tachycardia or Torsades de Pointes | 62.2 |
| LPD Belgium | OPTUM Extended SES | New users of GLP-1 receptor antagonists nested in obesity | All events Drug Rash with Eosinophilia and Systemic Symptoms (DRESS) | 62.2 |
| LPD Belgium | OPTUM Extended SES | New users of Beta blockers nested in essential hypertension | Hospitalization with heart failure events | <0.001 |
| LPD Belgium | OPTUM Extended SES | New users of Beta blockers nested in essential hypertension | 4-point MACE | <0.001 |
| LPD Belgium | OPTUM Extended SES | New users of Beta blockers nested in essential hypertension | Total cardiovascular disease events (ischemic stroke, hemorrhagic stroke, heart failure, acute myocardial infarction or sudden cardiac death) | <0.001 |

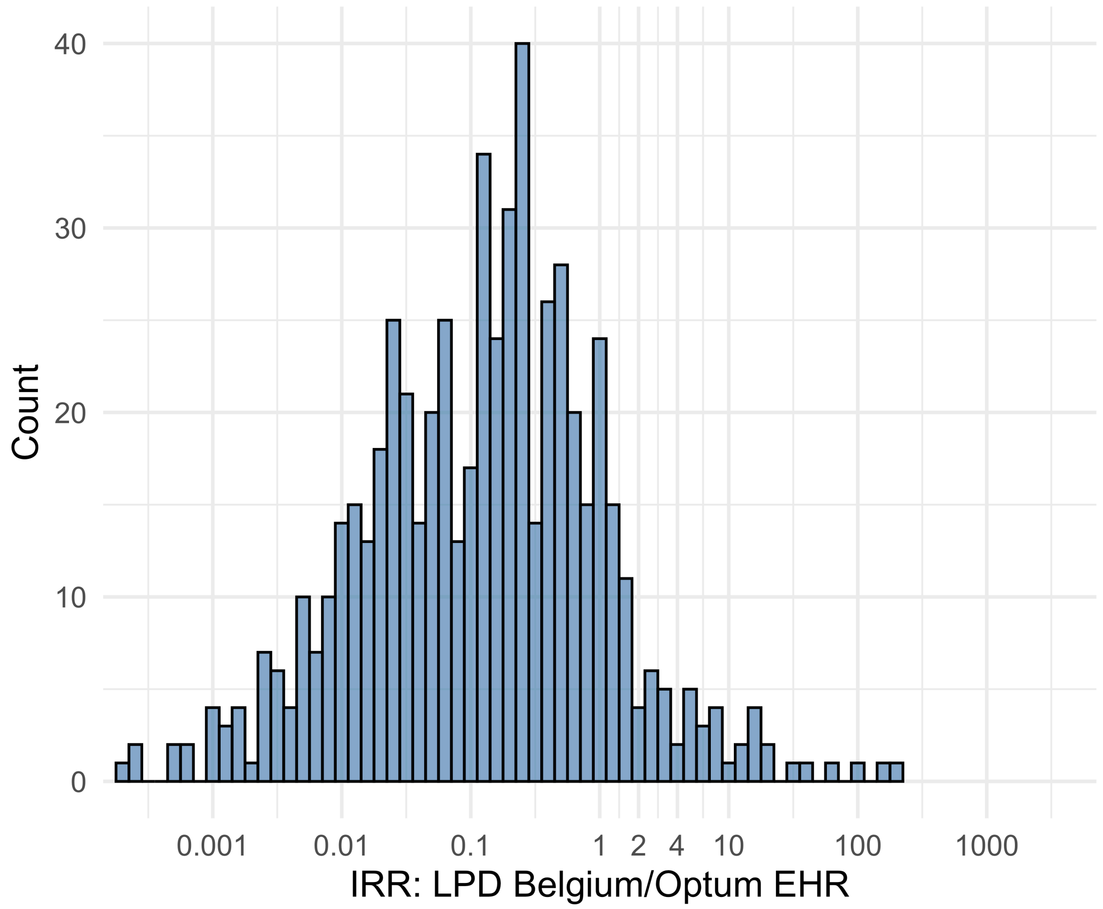


Figure S5.46. IRR distribution comparing LPD Belgium and Optum EHR.

*Table S5.46. Most extreme IRRs for LPD Belgium vs Optum EHR (rounded to 0.001).*

| source1 | source2 | Drug and Indication | Outcome | IRR |
|---|---|---|---|---|
| LPD Belgium | Optum EHR | New users of GLP-1 receptor antagonists nested in obesity | Progressive multifocal leukoencephalopathy | 186.5 |
| LPD Belgium | Optum EHR | New users of Trimethoprim systemetic nested in Urinary Tract Infection | Polymorphic Ventricular Tachycardia or Torsades de Pointes | 155.6 |
| LPD Belgium | Optum EHR | New users of GLP-1 receptor antagonists nested in Type 2 diabetes mellitus | Progressive multifocal leukoencephalopathy | 89.9 |
| LPD Belgium | Optum EHR | New users of Beta blockers nested in essential hypertension | Hospitalization with heart failure events | <0.001 |
| LPD Belgium | Optum EHR | New users of Beta blockers nested in essential hypertension | 4-point MACE | <0.001 |
| LPD Belgium | Optum EHR | New users of Beta blockers nested in essential hypertension | Total cardiovascular disease events (ischemic stroke, hemorrhagic stroke, heart failure, acute myocardial infarction or sudden cardiac death) | <0.001 |

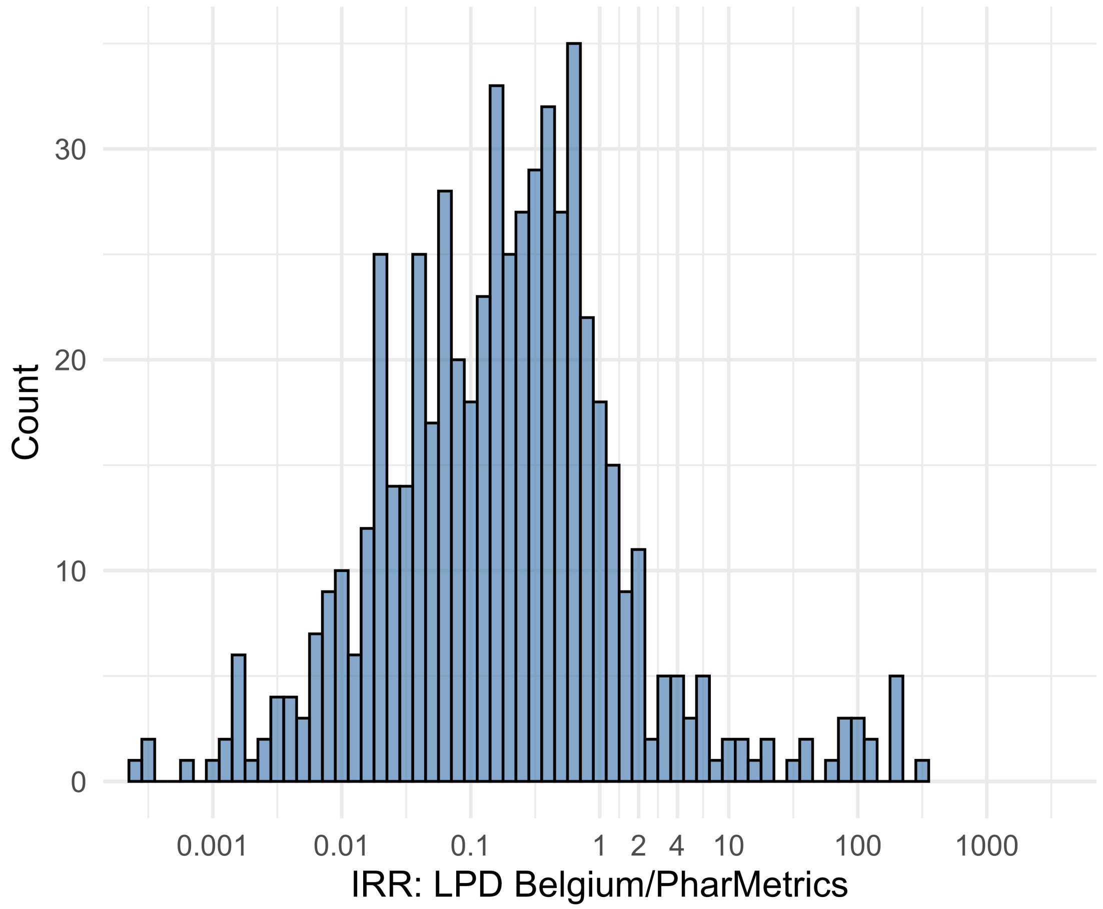


Figure S5.47. IRR distribution comparing LPD Belgium and PharMetrics.

*Table S5.47. Most extreme IRRs for LPD Belgium vs PharMetrics (rounded to 0.001).*

| source1 | source2 | Drug and Indication | Outcome | IRR |
|---|---|---|---|---|
| LPD Belgium | PharMetrics | New users of Cephalosporin systemetic nested in Urinary Tract Infection | Cardiovascular-related mortality | 346.0 |
| LPD Belgium | PharMetrics | New users of GLP-1 receptor antagonists nested in obesity | Cardiovascular-related mortality | 223.4 |
| LPD Belgium | PharMetrics | New users of GLP-1 receptor antagonists nested in obesity | Progressive multifocal leukoencephalopathy | 223.4 |
| LPD Belgium | PharMetrics | New users of Beta blockers nested in essential hypertension | Hospitalization with heart failure events | <0.001 |
| LPD Belgium | PharMetrics | New users of Beta blockers nested in essential hypertension | 4-point MACE | <0.001 |
| LPD Belgium | PharMetrics | New users of Beta blockers nested in essential hypertension | Total cardiovascular disease events (ischemic stroke, hemorrhagic stroke, heart failure, acute myocardial infarction or sudden cardiac death) | <0.001 |

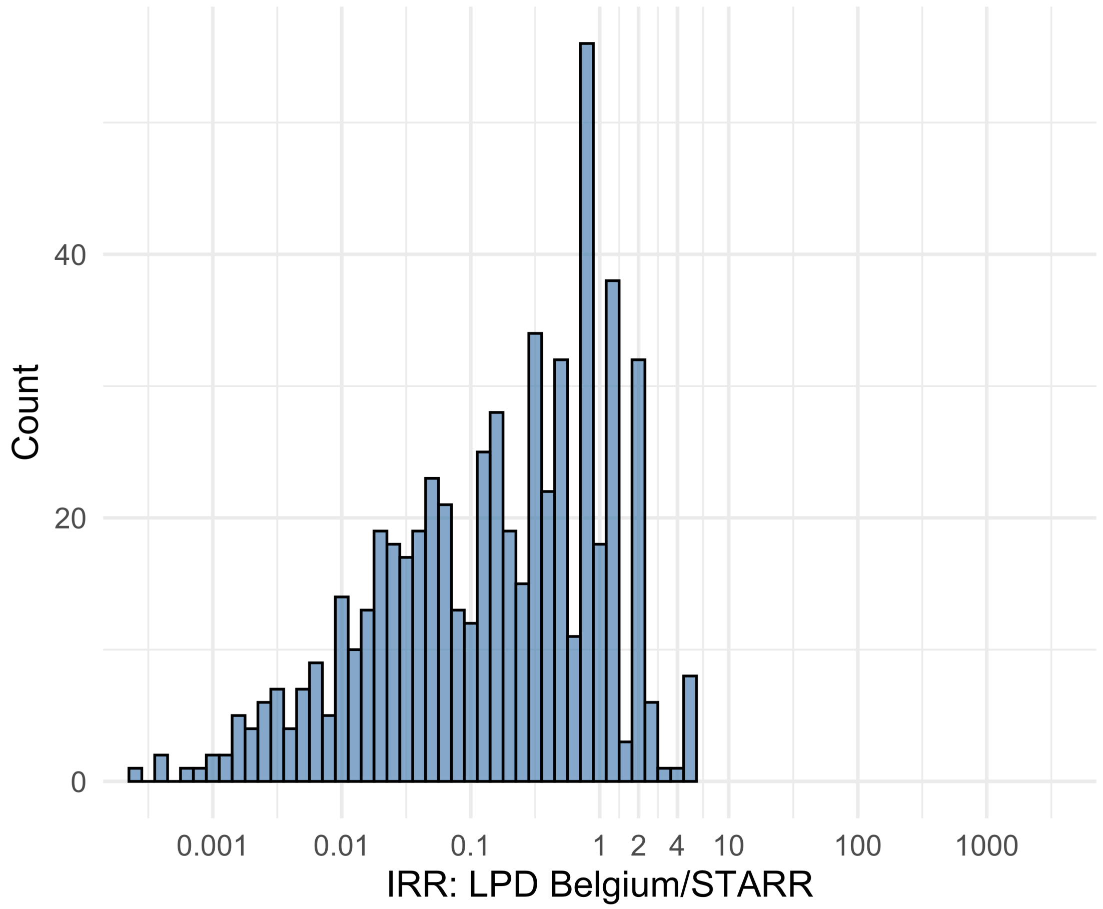


Figure S5.48. IRR distribution comparing LPD Belgium and STARR.

*Table S5.48. Most extreme IRRs for LPD Belgium vs STARR (rounded to 0.001).*

| source1 | source2 | Drug and Indication | Outcome | IRR |
|---|---|---|---|---|
| LPD Belgium | STARR | New users of GLP-1 receptor antagonists nested in obesity | Narcolepsy | 5.4 |
| LPD Belgium | STARR | New users of Cephalosporin systemetic nested in Urinary Tract Infection | Febrile Neutropenia or Neutropenic Fever | 5.1 |
| LPD Belgium | STARR | New users of Cephalosporin systemetic nested in Urinary Tract Infection | Polymorphic Ventricular Tachycardia or Torsades de Pointes | 5.1 |
| LPD Belgium | STARR | New users of Beta blockers nested in essential hypertension | Hospitalization with heart failure events | <0.001 |
| LPD Belgium | STARR | New users of Beta blockers nested in essential hypertension | 4-point MACE | <0.001 |
| LPD Belgium | STARR | New users of Beta blockers nested in essential hypertension | Total cardiovascular disease events (ischemic stroke, hemorrhagic stroke, heart failure, acute myocardial infarction or sudden cardiac death) | <0.001 |

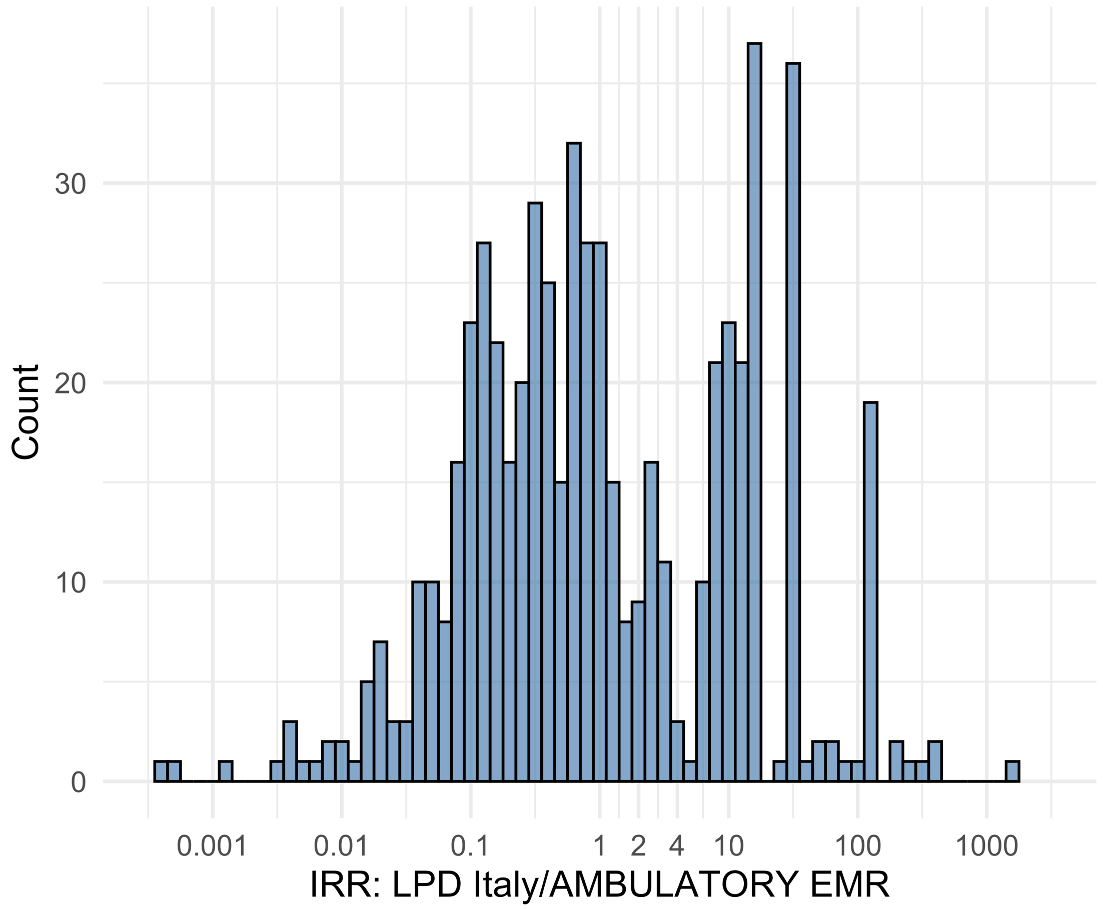


Figure S5.49. IRR distribution comparing LPD Italy and AMBULATORY EMR.

*Table S5.49. Most extreme IRRs for LPD Italy vs AMBULATORY EMR (rounded to 0.001).*

| source1 | source2 | Drug and Indication | Outcome | IRR |
|---|---|---|---|---|
| LPD Italy | AMBULATORY EMR | New users of Beta blockers nested in essential hypertension | Cardiovascular-related mortality | 1641.0 |
| LPD Italy | AMBULATORY EMR | New users of Cephalosporin systemetic nested in Urinary Tract Infection | Cardiovascular-related mortality | 444.9 |
| LPD Italy | AMBULATORY EMR | New users of Beta blockers nested in essential hypertension | Progressive multifocal leukoencephalopathy | 394.4 |
| LPD Italy | AMBULATORY EMR | New users of Fluoroquinolone systemic nested in Urinary Tract Infection | Fall events | <0.001 |
| LPD Italy | AMBULATORY EMR | New users of Beta blockers nested in essential hypertension | Fall events | <0.001 |
| LPD Italy | AMBULATORY EMR | New users of Cephalosporin systemetic nested in Urinary Tract Infection | Fall events | 0.0 |

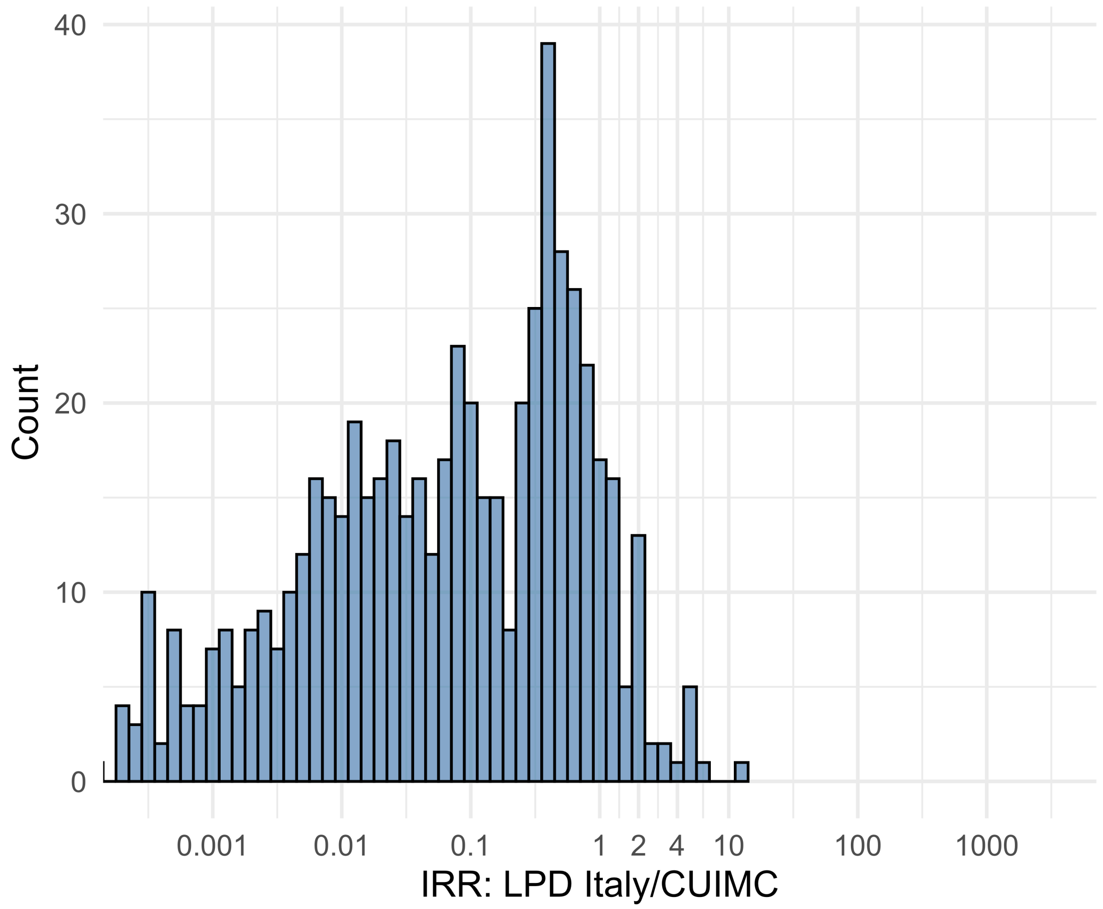


Figure S5.50. IRR distribution comparing LPD Italy and CUIMC.

*Table S5.50. Most extreme IRRs for LPD Italy vs CUIMC (rounded to 0.001).*

| source1 | source2 | Drug and Indication | Outcome | IRR |
|---|---|---|---|---|
| LPD Italy | CUIMC | New users of Cephalosporin systemetic nested in Urinary Tract Infection | Progressive multifocal leukoencephalopathy | 12.1 |
| LPD Italy | CUIMC | New users of SGLT2 inhibitor nested in Type 2 diabetes mellitus | Autoimmune hemolytic anemia | 6.9 |
| LPD Italy | CUIMC | New users of Beta blockers nested in essential hypertension | Progressive multifocal leukoencephalopathy | 4.9 |
| LPD Italy | CUIMC | New users of Beta blockers nested in essential hypertension | 4-point MACE | <0.001 |
| LPD Italy | CUIMC | New users of Beta blockers nested in essential hypertension | Total cardiovascular disease events (ischemic stroke, hemorrhagic stroke, heart failure, acute myocardial infarction or sudden cardiac death) | <0.001 |
| LPD Italy | CUIMC | New users of Beta blockers nested in essential hypertension | Hospitalization with heart failure events | <0.001 |

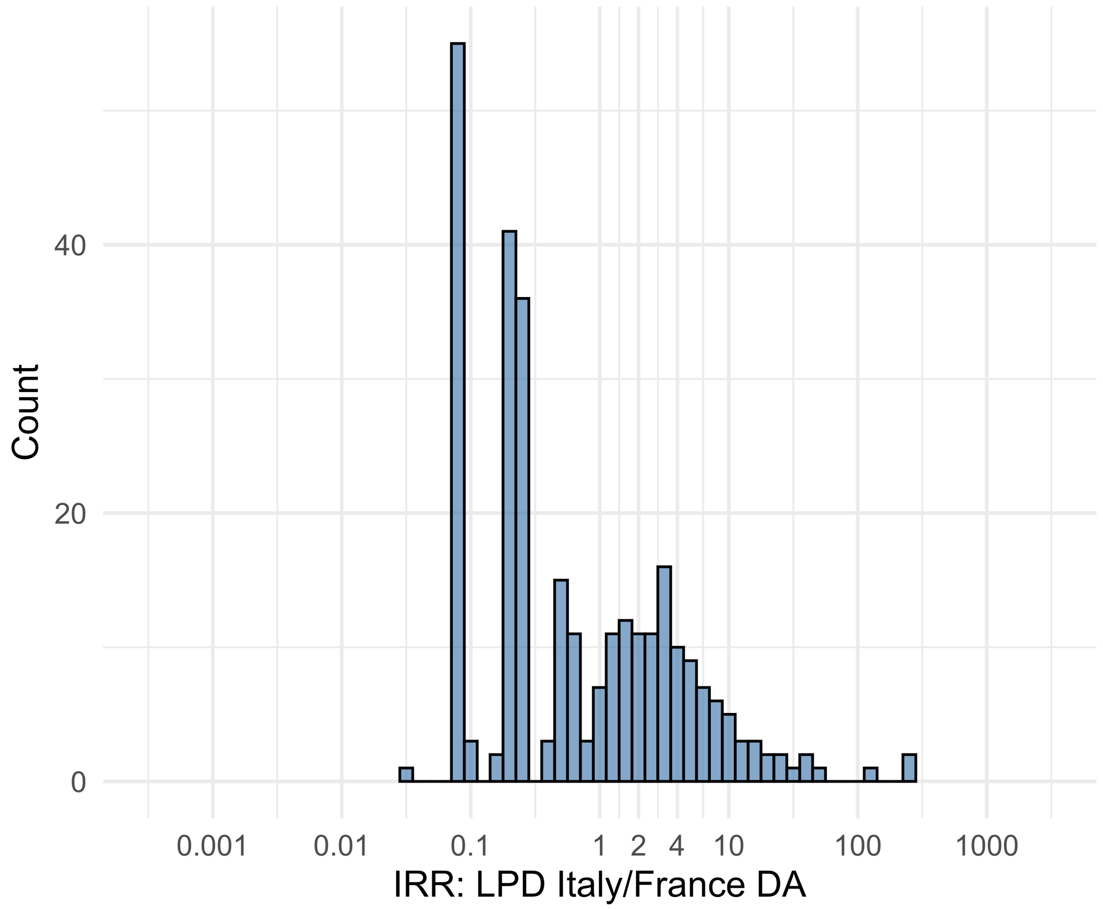


Figure S5.51. IRR distribution comparing LPD Italy and France DA.

*Table S5.51. Most extreme IRRs for LPD Italy vs France DA (rounded to 0.001).*

| source1 | source2 | Drug and Indication | Outcome | IRR |
|---|---|---|---|---|
| LPD Italy | France DA | New users of Beta blockers nested in essential hypertension | Diarrhea events | 275.4 |
| LPD Italy | France DA | New users of Fluoroquinolone systemic nested in Urinary Tract Infection | Diarrhea events | 243.6 |
| LPD Italy | France DA | New users of GLP-1 receptor antagonists nested in Type 2 diabetes mellitus | Diarrhea events | 119.2 |
| LPD Italy | France DA | New users of Beta blockers nested in essential hypertension | All events of Autoimmune hepatitis, with a washout period of 365 days | 0.0 |
| LPD Italy | France DA | New users of Beta blockers nested in essential hypertension | Earliest event of Acute Hepatic Failure, NO viral hepatitis or alcoholic hepatic failure | 0.1 |
| LPD Italy | France DA | New users of Beta blockers nested in essential hypertension | Earliest event of Acute Hepatic Failure | 0.1 |

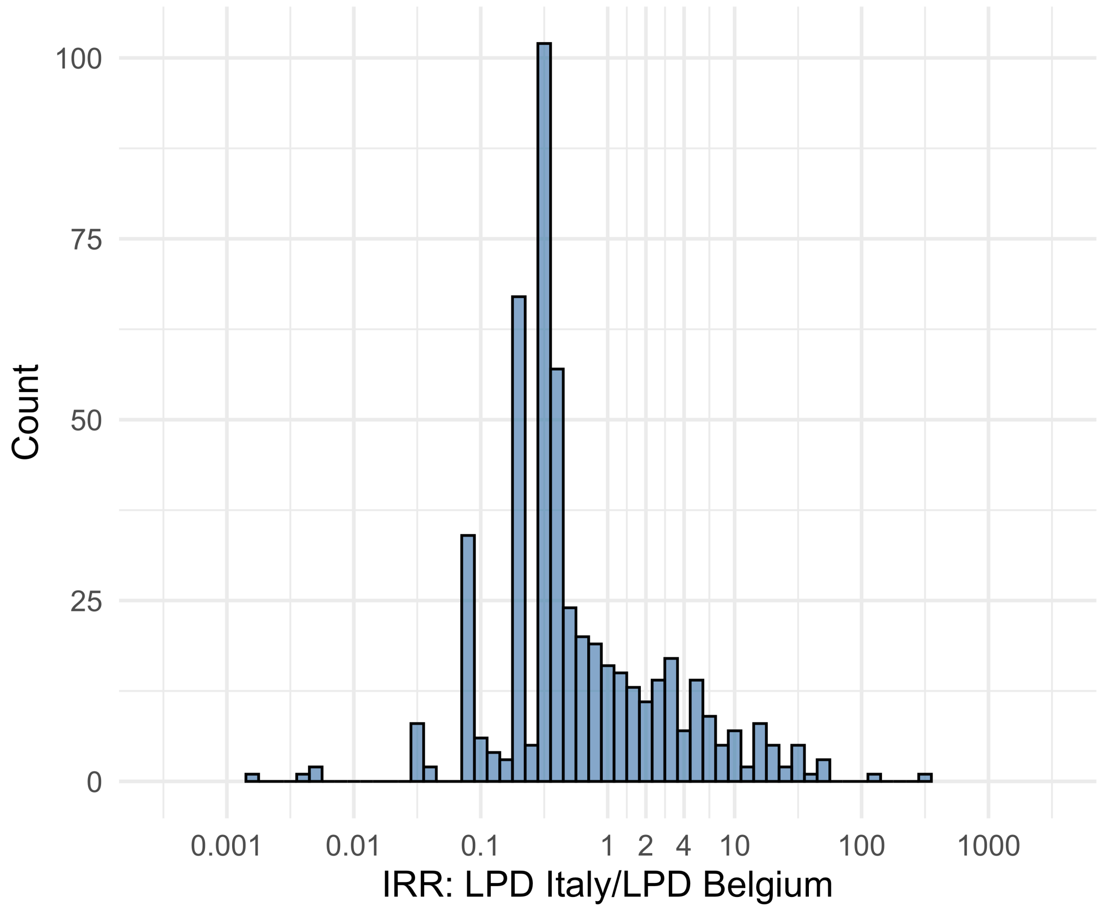


Figure S5.52. IRR distribution comparing LPD Italy and LPD Belgium.

*Table S5.52. Most extreme IRRs for LPD Italy vs LPD Belgium (rounded to 0.001).*

| source1 | source2 | Drug and Indication | Outcome | IRR |
|---|---|---|---|---|
| LPD Italy | LPD Belgium | New users of Beta blockers nested in essential hypertension | Deep Vein Thrombosis (DVT) | 340.5 |
| LPD Italy | LPD Belgium | New users of Fluoroquinolone systemic nested in Urinary Tract Infection | Deep Vein Thrombosis (DVT) | 124.1 |
| LPD Italy | LPD Belgium | New users of Beta blockers nested in essential hypertension | Bladder Cancer | 54.8 |
| LPD Italy | LPD Belgium | New users of GLP-1 receptor antagonists nested in obesity | Abnormal weight gain events | 0.0 |
| LPD Italy | LPD Belgium | New users of Beta blockers nested in essential hypertension | Fall events | 0.0 |
| LPD Italy | LPD Belgium | New users of GLP-1 receptor antagonists nested in Type 2 diabetes mellitus | Abnormal weight gain events | 0.0 |

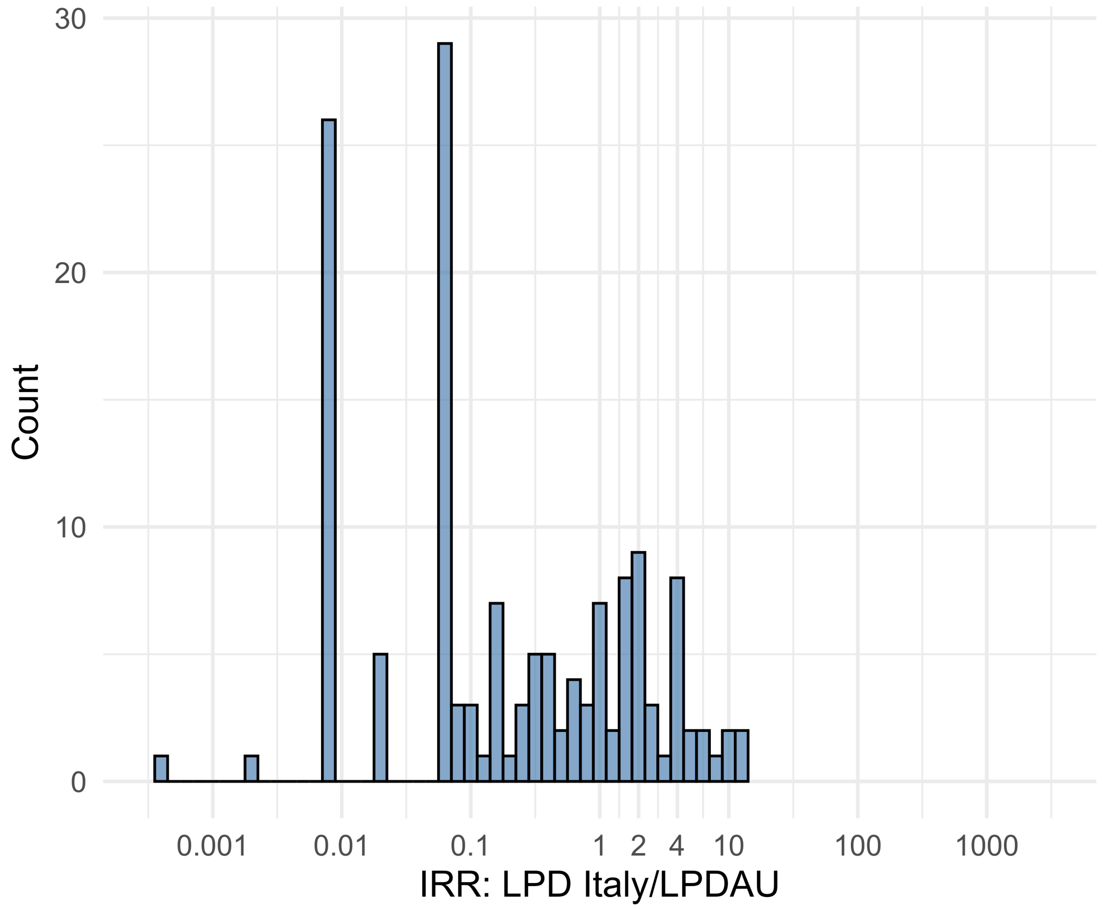


Figure S5.53. IRR distribution comparing LPD Italy and LPDAU.

*Table S5.53. Most extreme IRRs for LPD Italy vs LPDAU (rounded to 0.001).*

| source1 | source2 | Drug and Indication | Outcome | IRR |
|---|---|---|---|---|
| LPD Italy | LPDAU | New users of Cephalosporin systemetic nested in Urinary Tract Infection | Persons with chronic kidney disease | 12.9 |
| LPD Italy | LPDAU | New users of Cephalosporin systemetic nested in Urinary Tract Infection | Abnormal weight loss events | 11.3 |
| LPD Italy | LPDAU | New users of Beta blockers nested in essential hypertension | Persons with chronic kidney disease | 10.6 |
| LPD Italy | LPDAU | New users of Beta blockers nested in essential hypertension | Fall events | <0.001 |
| LPD Italy | LPDAU | New users of Cephalosporin systemetic nested in Urinary Tract Infection | Fall events | 0.0 |
| LPD Italy | LPDAU | New users of Beta blockers nested in essential hypertension | Sudden Vision Loss | 0.0 |

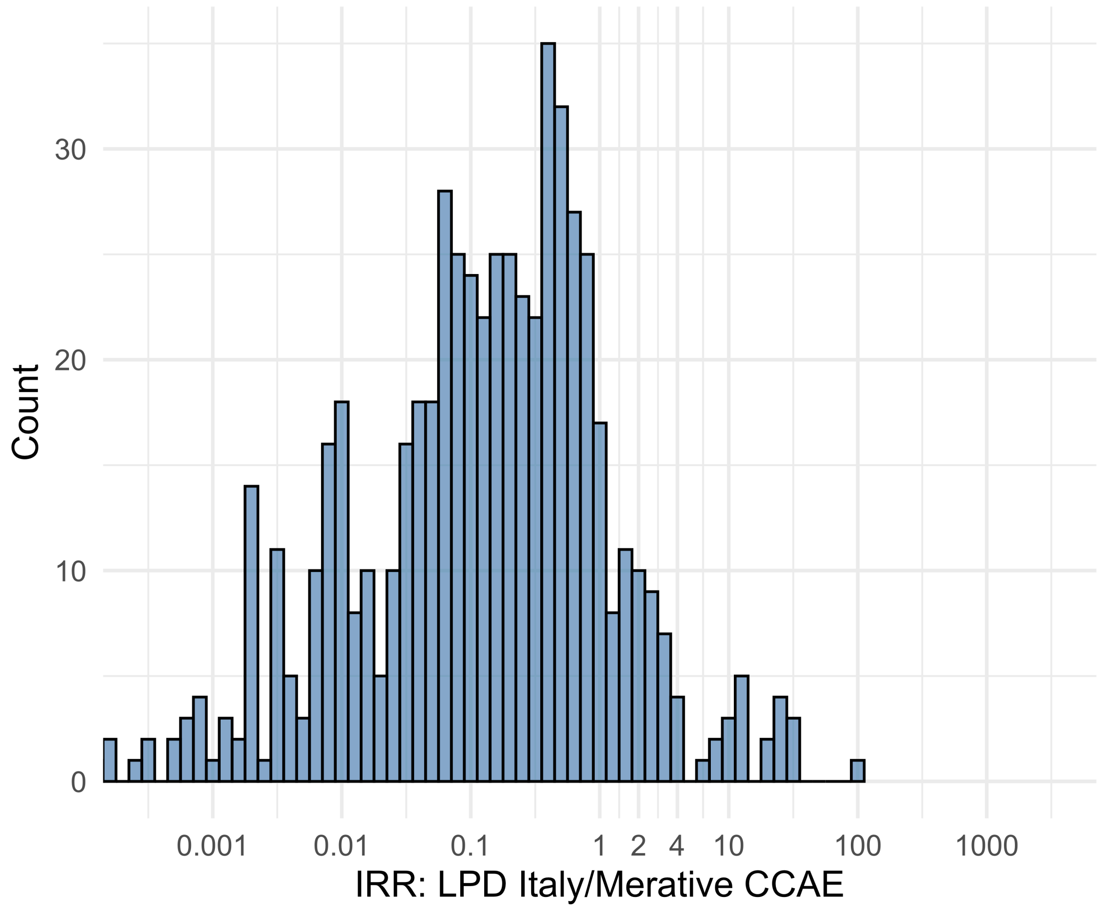


Figure S5.54. IRR distribution comparing LPD Italy and Merative CCAE.

*Table S5.54. Most extreme IRRs for LPD Italy vs Merative CCAE (rounded to 0.001).*

| source1 | source2 | Drug and Indication | Outcome | IRR |
|---|---|---|---|---|
| LPD Italy | Merative CCAE | New users of Cephalosporin systemetic nested in Urinary Tract Infection | Progressive multifocal leukoencephalopathy | 109.6 |
| LPD Italy | Merative CCAE | New users of GLP-1 receptor antagonists nested in obesity | Progressive multifocal leukoencephalopathy | 31.2 |
| LPD Italy | Merative CCAE | New users of GLP-1 receptor antagonists nested in obesity | Polymorphic Ventricular Tachycardia or Torsades de Pointes | 31.2 |
| LPD Italy | Merative CCAE | New users of Beta blockers nested in essential hypertension | Hospitalization with heart failure events | <0.001 |
| LPD Italy | Merative CCAE | New users of Beta blockers nested in essential hypertension | 4-point MACE | <0.001 |
| LPD Italy | Merative CCAE | New users of Beta blockers nested in essential hypertension | Total cardiovascular disease events (ischemic stroke, hemorrhagic stroke, heart failure, acute myocardial infarction or sudden cardiac death) | <0.001 |

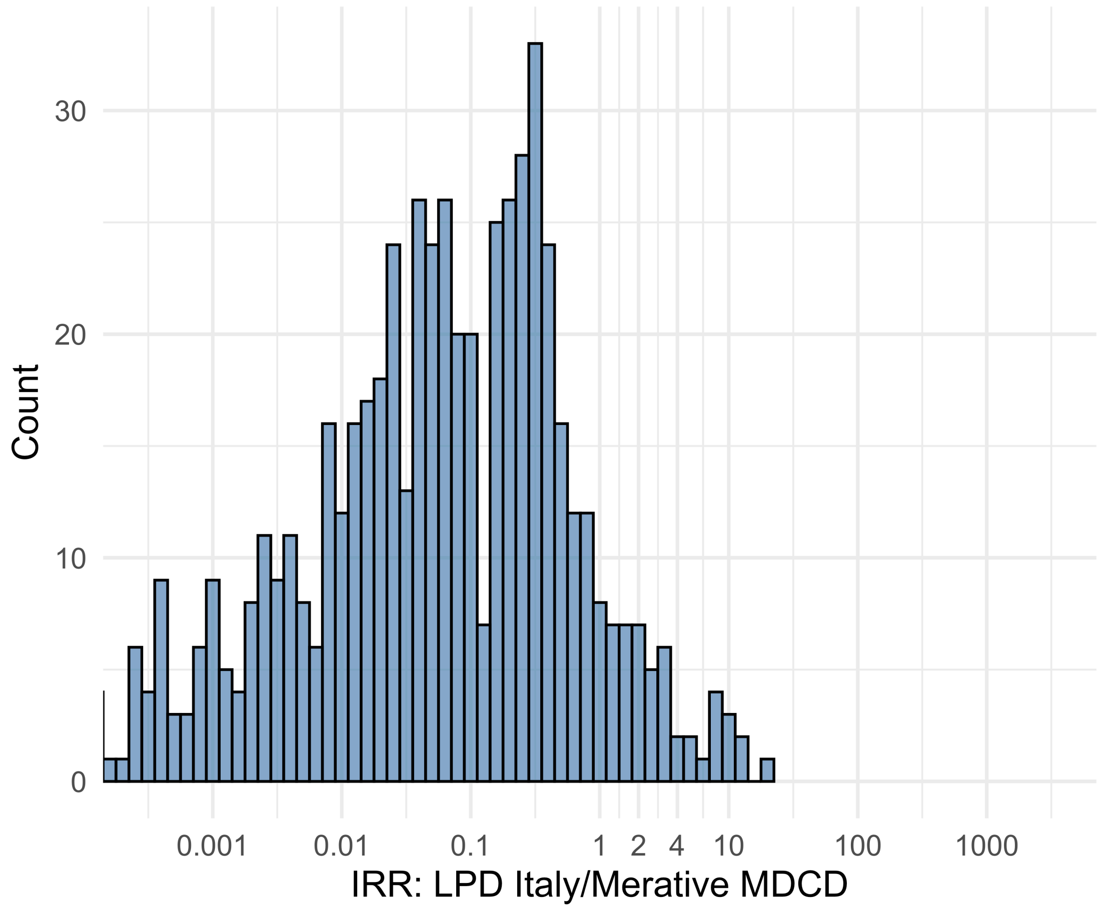


Figure S5.55. IRR distribution comparing LPD Italy and Merative MDCD.

*Table S5.55. Most extreme IRRs for LPD Italy vs Merative MDCD (rounded to 0.001).*

| source1 | source2 | Drug and Indication | Outcome | IRR |
|---|---|---|---|---|
| LPD Italy | Merative MDCD | New users of GLP-1 receptor antagonists nested in obesity | Polymorphic Ventricular Tachycardia or Torsades de Pointes | 19.9 |
| LPD Italy | Merative MDCD | New users of Cephalosporin systemetic nested in Acute Typical Pneumonia | Progressive multifocal leukoencephalopathy | 13.3 |
| LPD Italy | Merative MDCD | New users of SGLT2 inhibitor nested in Type 2 diabetes mellitus | Progressive multifocal leukoencephalopathy | 12.8 |
| LPD Italy | Merative MDCD | New users of Beta blockers nested in essential hypertension | Hospitalization with heart failure events | <0.001 |
| LPD Italy | Merative MDCD | New users of Beta blockers nested in essential hypertension | 4-point MACE | <0.001 |
| LPD Italy | Merative MDCD | New users of Beta blockers nested in essential hypertension | Total cardiovascular disease events (ischemic stroke, hemorrhagic stroke, heart failure, acute myocardial infarction or sudden cardiac death) | <0.001 |

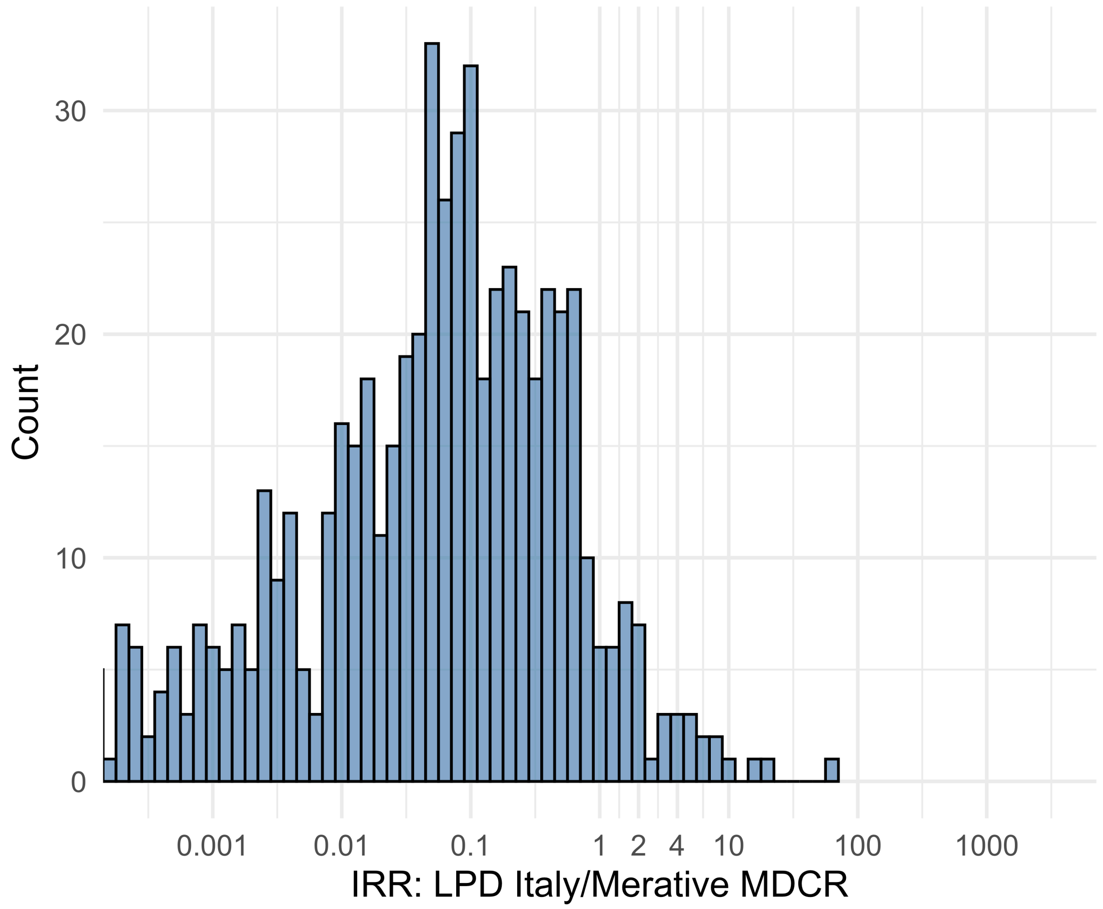


Figure S5.56. IRR distribution comparing LPD Italy and Merative MDCR.

*Table S5.56. Most extreme IRRs for LPD Italy vs Merative MDCR (rounded to 0.001).*

| source1 | source2 | Drug and Indication | Outcome | IRR |
|---|---|---|---|---|
| LPD Italy | Merative MDCR | New users of Beta blockers nested in essential hypertension | Progressive multifocal leukoencephalopathy | 70.7 |
| LPD Italy | Merative MDCR | New users of Cephalosporin systemetic nested in Urinary Tract Infection | Progressive multifocal leukoencephalopathy | 19.9 |
| LPD Italy | Merative MDCR | New users of Fluoroquinolone systemic nested in Urinary Tract Infection | Progressive multifocal leukoencephalopathy | 16.2 |
| LPD Italy | Merative MDCR | New users of Beta blockers nested in essential hypertension | Hospitalization with heart failure events | <0.001 |
| LPD Italy | Merative MDCR | New users of Beta blockers nested in essential hypertension | 4-point MACE | <0.001 |
| LPD Italy | Merative MDCR | New users of Beta blockers nested in essential hypertension | Total cardiovascular disease events (ischemic stroke, hemorrhagic stroke, heart failure, acute myocardial infarction or sudden cardiac death) | <0.001 |

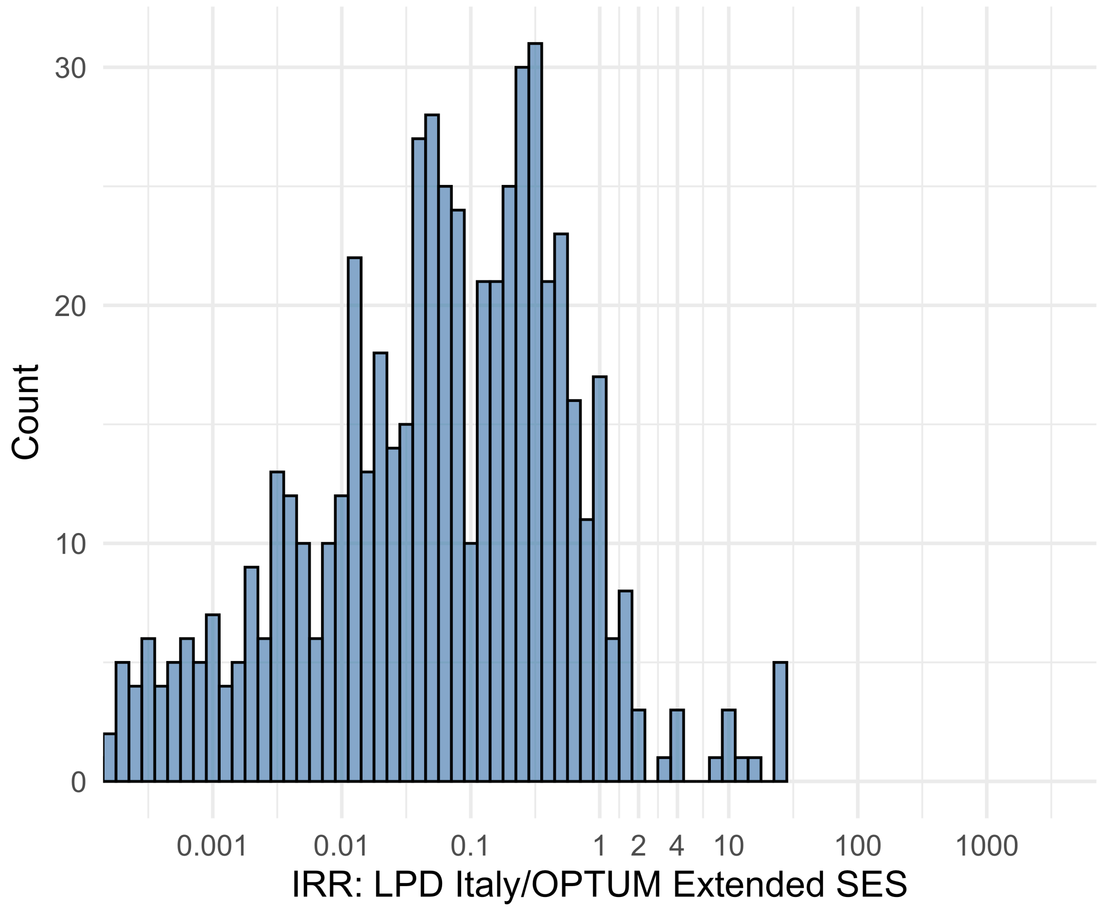


Figure S5.57. IRR distribution comparing LPD Italy and OPTUM Extended SES.

*Table S5.57. Most extreme IRRs for LPD Italy vs OPTUM Extended SES (rounded to 0.001).*

| source1 | source2 | Drug and Indication | Outcome | IRR |
|---|---|---|---|---|
| LPD Italy | OPTUM Extended SES | New users of GLP-1 receptor antagonists nested in obesity | Progressive multifocal leukoencephalopathy | 25.6 |
| LPD Italy | OPTUM Extended SES | New users of GLP-1 receptor antagonists nested in obesity | Polymorphic Ventricular Tachycardia or Torsades de Pointes | 25.5 |
| LPD Italy | OPTUM Extended SES | New users of GLP-1 receptor antagonists nested in obesity | All events Drug Rash with Eosinophilia and Systemic Symptoms (DRESS) | 25.5 |
| LPD Italy | OPTUM Extended SES | New users of Beta blockers nested in essential hypertension | Hospitalization with heart failure events | <0.001 |
| LPD Italy | OPTUM Extended SES | New users of Beta blockers nested in essential hypertension | 4-point MACE | <0.001 |
| LPD Italy | OPTUM Extended SES | New users of Beta blockers nested in essential hypertension | Total cardiovascular disease events (ischemic stroke, hemorrhagic stroke, heart failure, acute myocardial infarction or sudden cardiac death) | <0.001 |

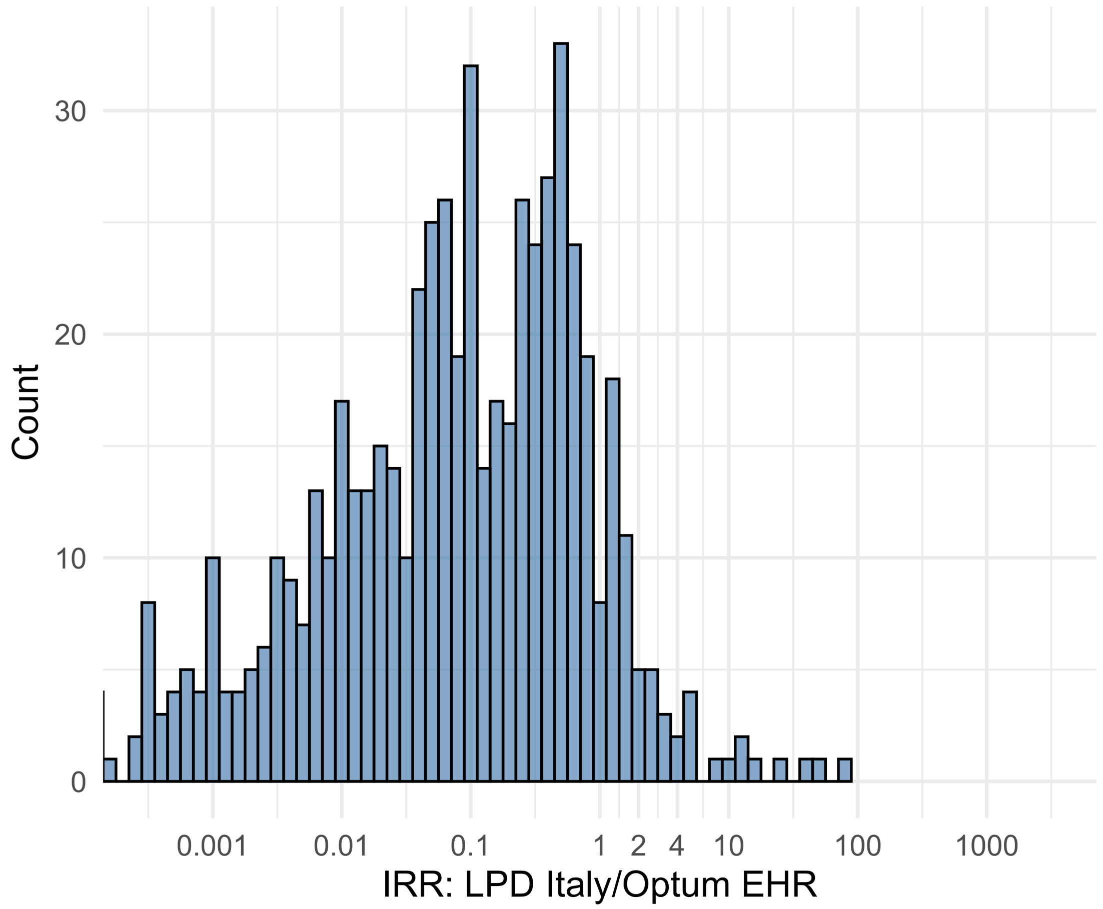


Figure S5.58. IRR distribution comparing LPD Italy and Optum EHR.

*Table S5.58. Most extreme IRRs for LPD Italy vs Optum EHR (rounded to 0.001).*

| source1 | source2 | Drug and Indication | Outcome | IRR |
|---|---|---|---|---|
| LPD Italy | Optum EHR | New users of GLP-1 receptor antagonists nested in obesity | Progressive multifocal leukoencephalopathy | 76.6 |
| LPD Italy | Optum EHR | New users of GLP-1 receptor antagonists nested in Type 2 diabetes mellitus | Progressive multifocal leukoencephalopathy | 46.3 |
| LPD Italy | Optum EHR | New users of SGLT2 inhibitor nested in Type 2 diabetes mellitus | Progressive multifocal leukoencephalopathy | 41.8 |
| LPD Italy | Optum EHR | New users of Beta blockers nested in essential hypertension | Hospitalization with heart failure events | <0.001 |
| LPD Italy | Optum EHR | New users of Beta blockers nested in essential hypertension | 4-point MACE | <0.001 |
| LPD Italy | Optum EHR | New users of Beta blockers nested in essential hypertension | Total cardiovascular disease events (ischemic stroke, hemorrhagic stroke, heart failure, acute myocardial infarction or sudden cardiac death) | <0.001 |

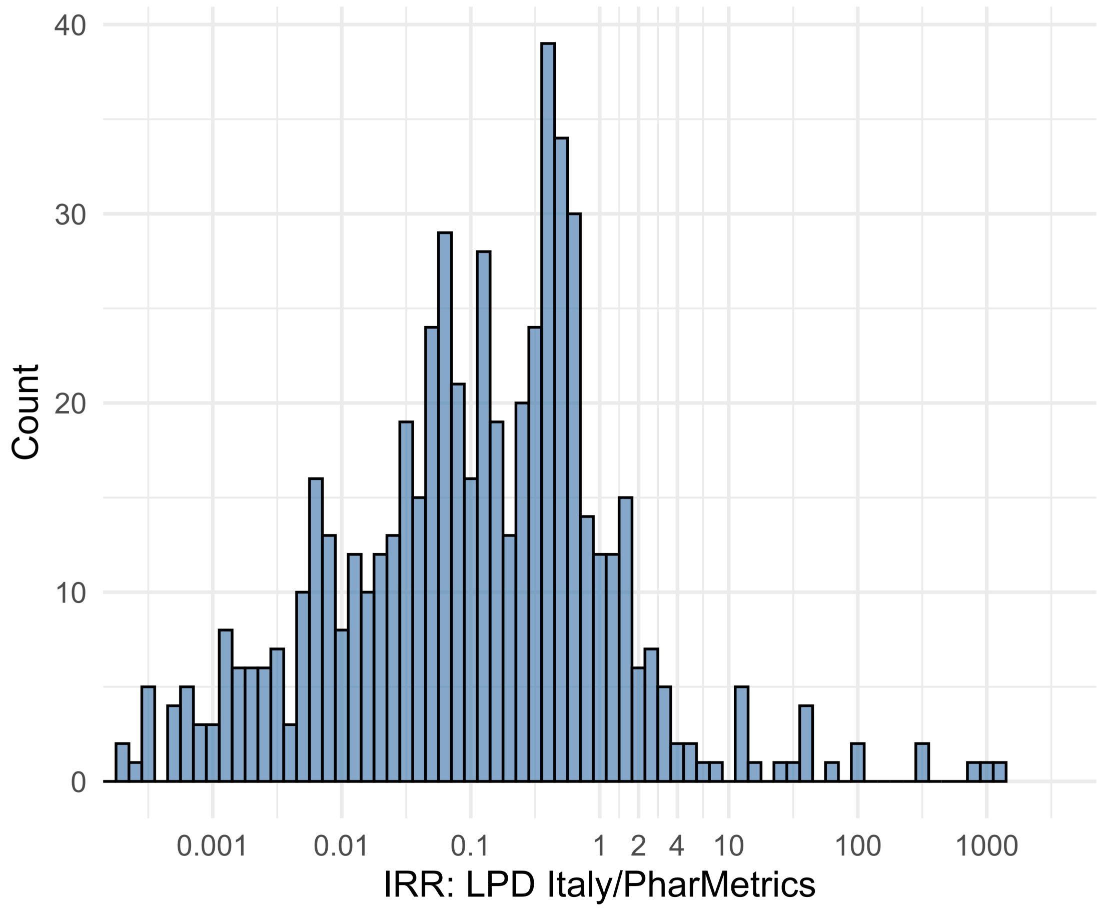


Figure S5.59. IRR distribution comparing LPD Italy and PharMetrics.

*Table S5.59. Most extreme IRRs for LPD Italy vs PharMetrics (rounded to 0.001).*

| source1 | source2 | Drug and Indication | Outcome | IRR |
|---|---|---|---|---|
| LPD Italy | PharMetrics | New users of Beta blockers nested in essential hypertension | Cardiovascular-related mortality | 1354.9 |
| LPD Italy | PharMetrics | New users of Cephalosporin systemetic nested in Urinary Tract Infection | Cardiovascular-related mortality | 950.4 |
| LPD Italy | PharMetrics | New users of Cephalosporin systemetic nested in Acute Typical Pneumonia | Cardiovascular-related mortality | 756.3 |
| LPD Italy | PharMetrics | New users of Beta blockers nested in essential hypertension | Hospitalization with heart failure events | <0.001 |
| LPD Italy | PharMetrics | New users of Beta blockers nested in essential hypertension | 4-point MACE | <0.001 |
| LPD Italy | PharMetrics | New users of Beta blockers nested in essential hypertension | Total cardiovascular disease events (ischemic stroke, hemorrhagic stroke, heart failure, acute myocardial infarction or sudden cardiac death) | <0.001 |

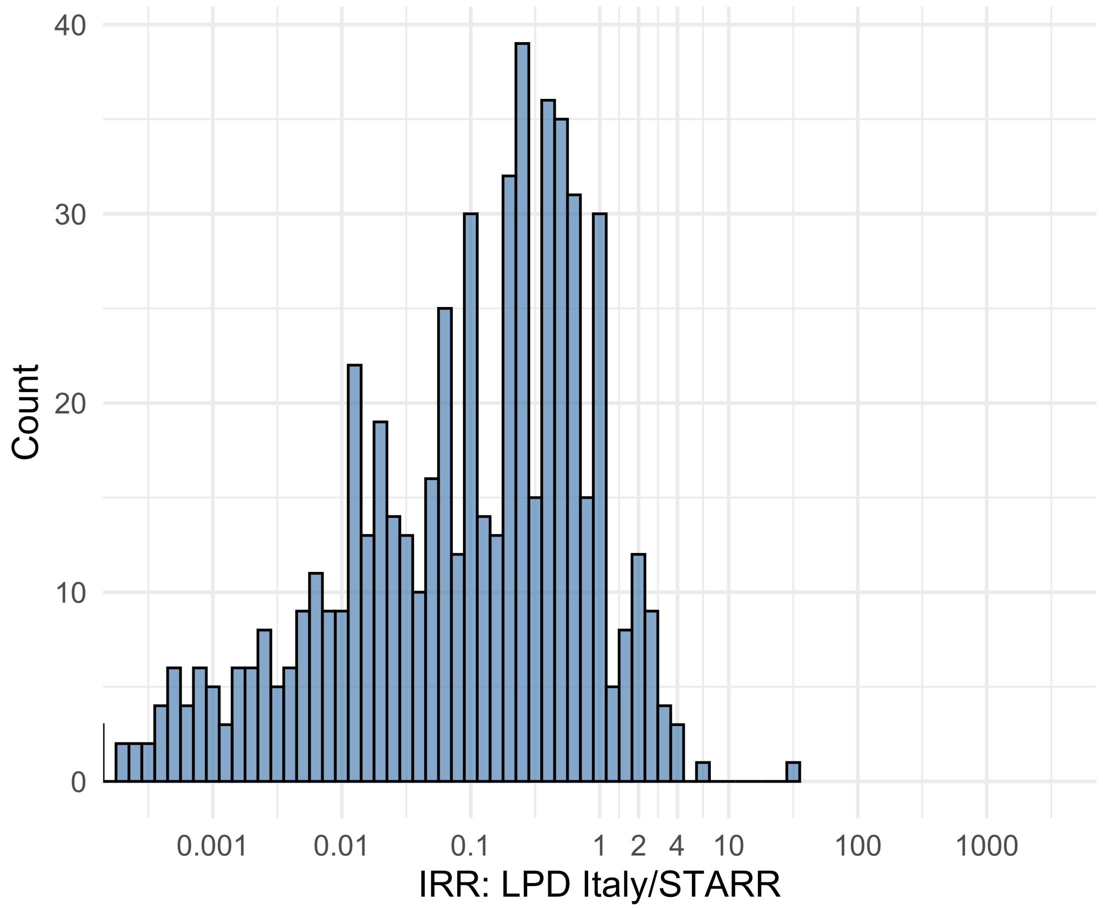


Figure S5.60. IRR distribution comparing LPD Italy and STARR.

*Table S5.60. Most extreme IRRs for LPD Italy vs STARR (rounded to 0.001).*

| source1 | source2 | Drug and Indication | Outcome | IRR |
|---|---|---|---|---|
| LPD Italy | STARR | New users of Beta blockers nested in essential hypertension | Progressive multifocal leukoencephalopathy | 33.3 |
| LPD Italy | STARR | New users of Cephalosporin systemetic nested in Urinary Tract Infection | Progressive multifocal leukoencephalopathy | 6.2 |
| LPD Italy | STARR | New users of Cephalosporin systemetic nested in Acute Typical Pneumonia | Renal Cancer | 4.4 |
| LPD Italy | STARR | New users of Beta blockers nested in essential hypertension | Hospitalization with heart failure events | <0.001 |
| LPD Italy | STARR | New users of Beta blockers nested in essential hypertension | 4-point MACE | <0.001 |
| LPD Italy | STARR | New users of Beta blockers nested in essential hypertension | Total cardiovascular disease events (ischemic stroke, hemorrhagic stroke, heart failure, acute myocardial infarction or sudden cardiac death) | <0.001 |

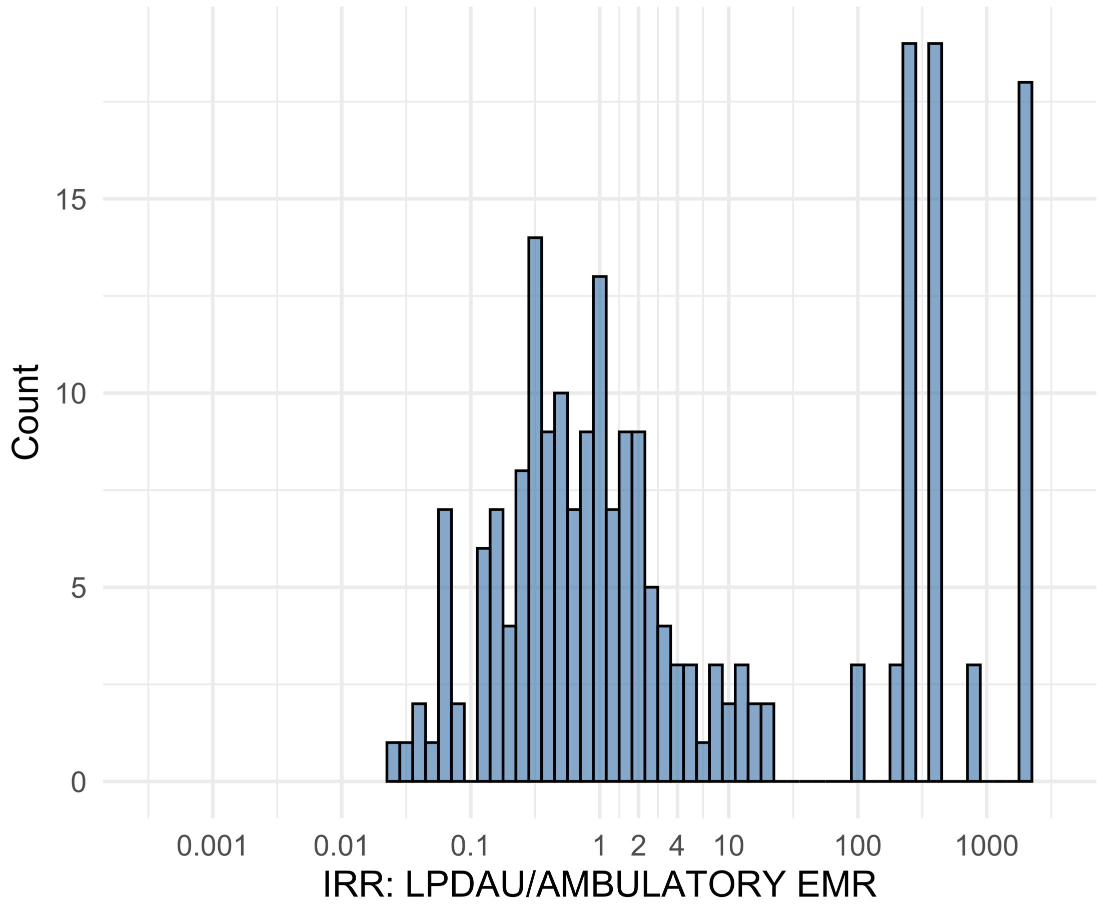


Figure S5.61. IRR distribution comparing LPDAU and AMBULATORY EMR.

*Table S5.61. Most extreme IRRs for LPDAU vs AMBULATORY EMR (rounded to 0.001).*

| source1 | source2 | Drug and Indication | Outcome | IRR |
|---|---|---|---|---|
| LPDAU | AMBULATORY EMR | New users of Beta blockers nested in essential hypertension | Febrile Neutropenia or Neutropenic Fever | 1858.9 |
| LPDAU | AMBULATORY EMR | New users of Beta blockers nested in essential hypertension | Rhabdomyolysis | 1858.9 |
| LPDAU | AMBULATORY EMR | New users of Beta blockers nested in essential hypertension | Sudden Cardiac arrest or cardiac death | 1858.9 |
| LPDAU | AMBULATORY EMR | New users of Cephalosporin systemetic nested in Urinary Tract Infection | Hypokalemia events | 0.0 |
| LPDAU | AMBULATORY EMR | New users of Cephalosporin systemetic nested in Urinary Tract Infection | Acute Kidney Injury AKI | 0.0 |
| LPDAU | AMBULATORY EMR | New users of Trimethoprim systemetic nested in Urinary Tract Infection | Abnormal weight loss events | 0.0 |

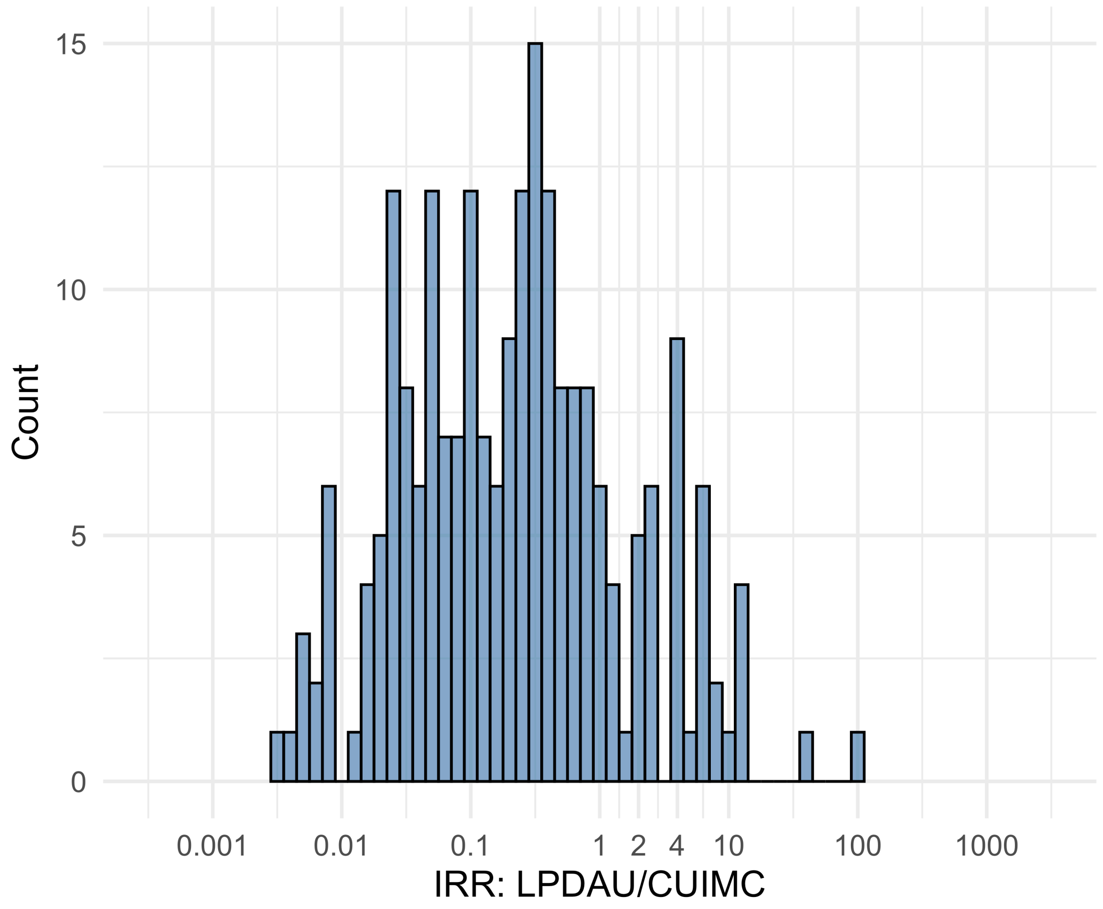


Figure S5.62. IRR distribution comparing LPDAU and CUIMC.

*Table S5.62. Most extreme IRRs for LPDAU vs CUIMC (rounded to 0.001).*

| source1 | source2 | Drug and Indication | Outcome | IRR |
|---|---|---|---|---|
| LPDAU | CUIMC | New users of Beta blockers nested in essential hypertension | Polymorphic Ventricular Tachycardia or Torsades de Pointes | 94.5 |
| LPDAU | CUIMC | New users of Beta blockers nested in essential hypertension | All events Drug Rash with Eosinophilia and Systemic Symptoms (DRESS) | 38.6 |
| LPDAU | CUIMC | New users of Cephalosporin systemetic nested in Urinary Tract Infection | Polymorphic Ventricular Tachycardia or Torsades de Pointes | 14.0 |
| LPDAU | CUIMC | New users of Cephalosporin systemetic nested in Urinary Tract Infection | Acute Kidney Injury AKI | 0.0 |
| LPDAU | CUIMC | New users of Trimethoprim systemetic nested in Urinary Tract Infection | Acute Kidney Injury AKI | 0.0 |
| LPDAU | CUIMC | New users of Cephalosporin systemetic nested in Urinary Tract Infection | Hospitalization with heart failure events | 0.0 |

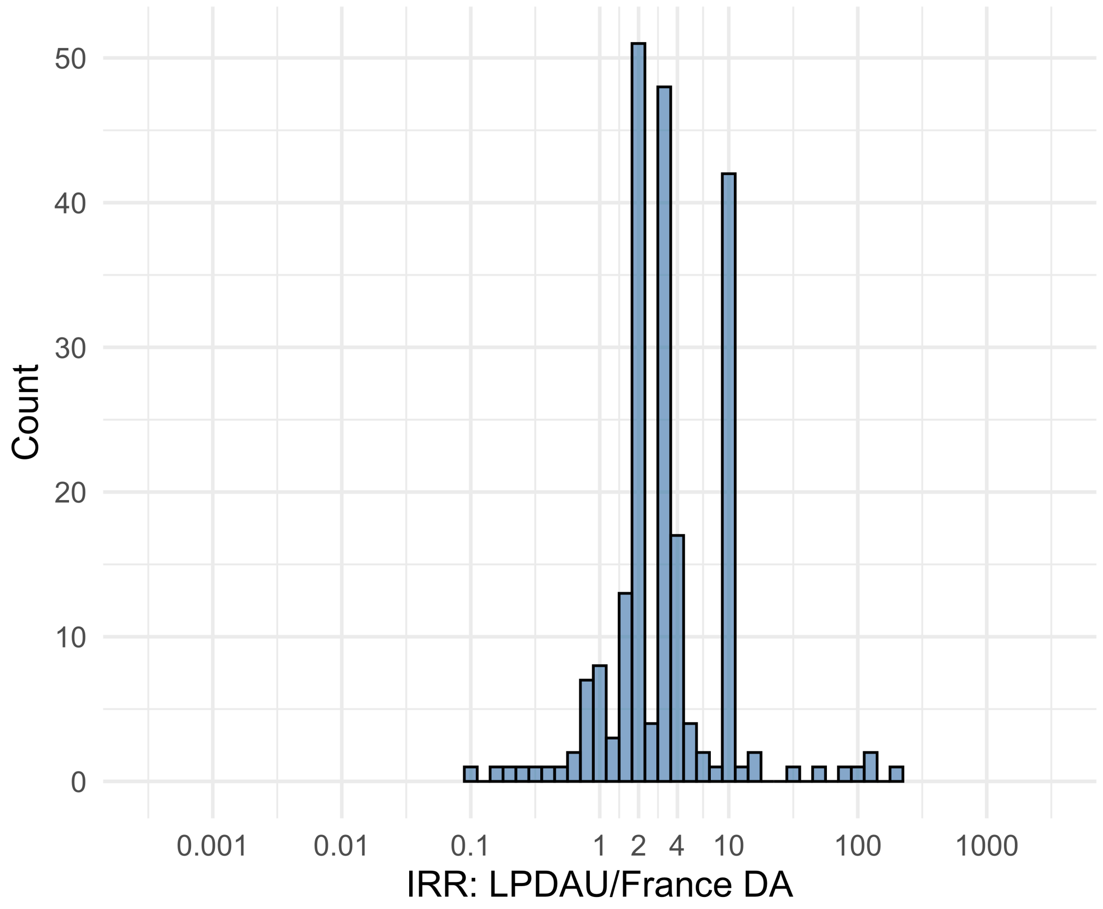


Figure S5.63. IRR distribution comparing LPDAU and France DA.

*Table S5.63. Most extreme IRRs for LPDAU vs France DA (rounded to 0.001).*

| source1 | source2 | Drug and Indication | Outcome | IRR |
|---|---|---|---|---|
| LPDAU | France DA | New users of Beta blockers nested in essential hypertension | Fall events | 202.1 |
| LPDAU | France DA | New users of Beta blockers nested in essential hypertension | Diarrhea events | 134.4 |
| LPDAU | France DA | New users of Trimethoprim systemetic nested in Urinary Tract Infection | Diarrhea events | 121.4 |
| LPDAU | France DA | New users of Trimethoprim systemetic nested in Urinary Tract Infection | Abnormal weight loss events | 0.1 |
| LPDAU | France DA | New users of Trimethoprim systemetic nested in Urinary Tract Infection | Bladder Cancer | 0.1 |
| LPDAU | France DA | New users of Beta blockers nested in essential hypertension | Abnormal weight loss events | 0.2 |

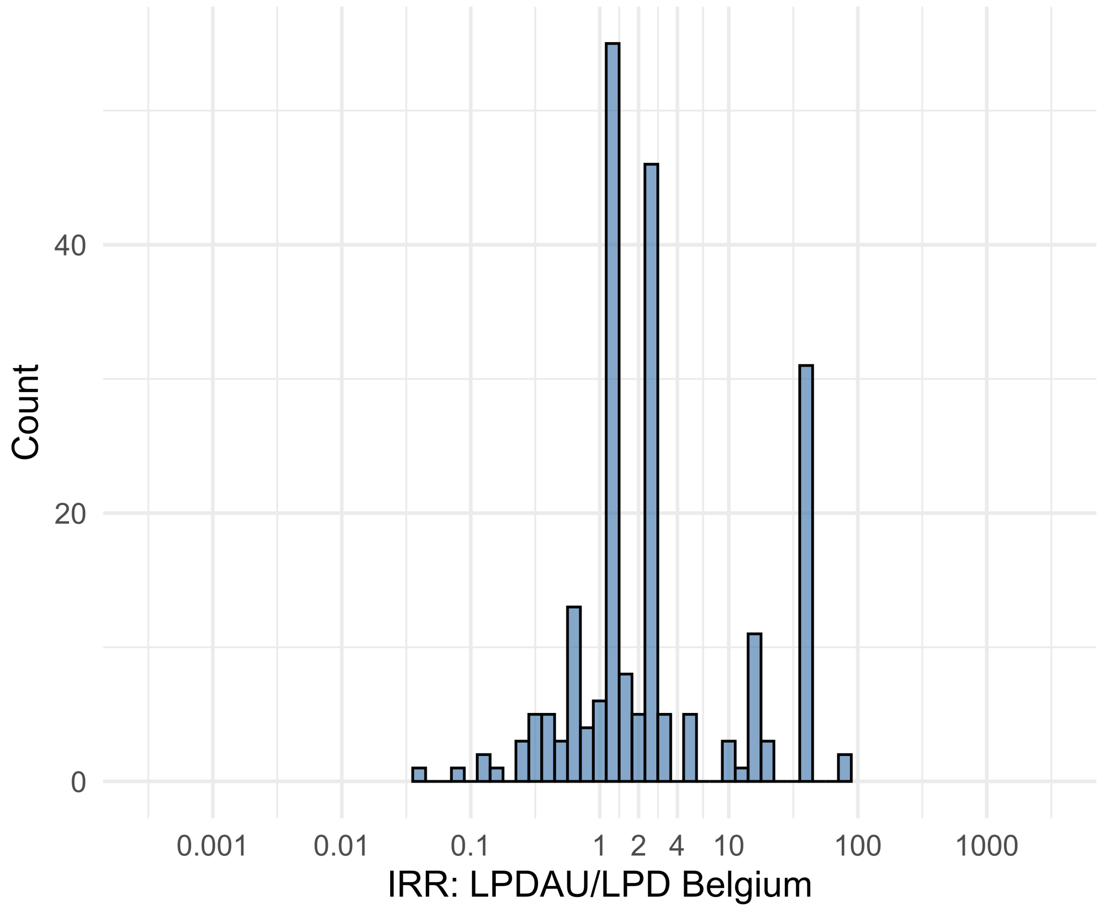


Figure S5.64. IRR distribution comparing LPDAU and LPD Belgium.

*Table S5.64. Most extreme IRRs for LPDAU vs LPD Belgium (rounded to 0.001).*

| source1 | source2 | Drug and Indication | Outcome | IRR |
|---|---|---|---|---|
| LPDAU | LPD Belgium | New users of Beta blockers nested in essential hypertension | Deep Vein Thrombosis (DVT) | 87.7 |
| LPDAU | LPD Belgium | New users of Beta blockers nested in essential hypertension | Anaphylaxis | 87.1 |
| LPDAU | LPD Belgium | New users of Trimethoprim systemetic nested in Urinary Tract Infection | Deep Vein Thrombosis (DVT) | 37.2 |
| LPDAU | LPD Belgium | New users of Cephalosporin systemetic nested in Urinary Tract Infection | Acute Kidney Injury AKI | 0.0 |
| LPDAU | LPD Belgium | New users of Trimethoprim systemetic nested in Urinary Tract Infection | Acute Kidney Injury AKI | 0.1 |
| LPDAU | LPD Belgium | New users of Beta blockers nested in essential hypertension | Acute Kidney Injury AKI | 0.1 |

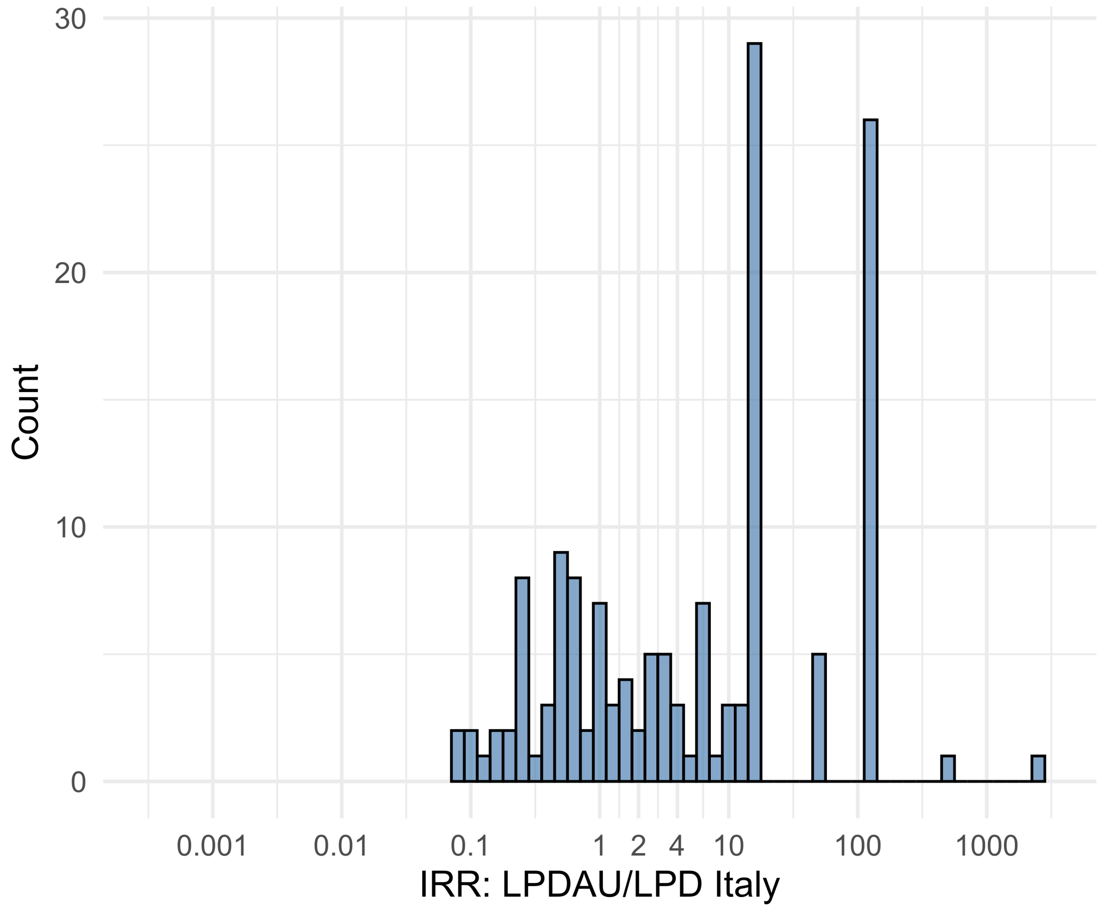


Figure S5.65. IRR distribution comparing LPDAU and LPD Italy.

*Table S5.65. Most extreme IRRs for LPDAU vs LPD Italy (rounded to 0.001).*

| source1 | source2 | Drug and Indication | Outcome | IRR |
|---|---|---|---|---|
| LPDAU | LPD Italy | New users of Beta blockers nested in essential hypertension | Fall events | 2557.8 |
| LPDAU | LPD Italy | New users of Cephalosporin systemetic nested in Urinary Tract Infection | Fall events | 523.6 |
| LPDAU | LPD Italy | New users of Beta blockers nested in essential hypertension | Sudden Vision Loss | 115.7 |
| LPDAU | LPD Italy | New users of Cephalosporin systemetic nested in Urinary Tract Infection | Persons with chronic kidney disease | 0.1 |
| LPDAU | LPD Italy | New users of Cephalosporin systemetic nested in Urinary Tract Infection | Abnormal weight loss events | 0.1 |
| LPDAU | LPD Italy | New users of Beta blockers nested in essential hypertension | Persons with chronic kidney disease | 0.1 |

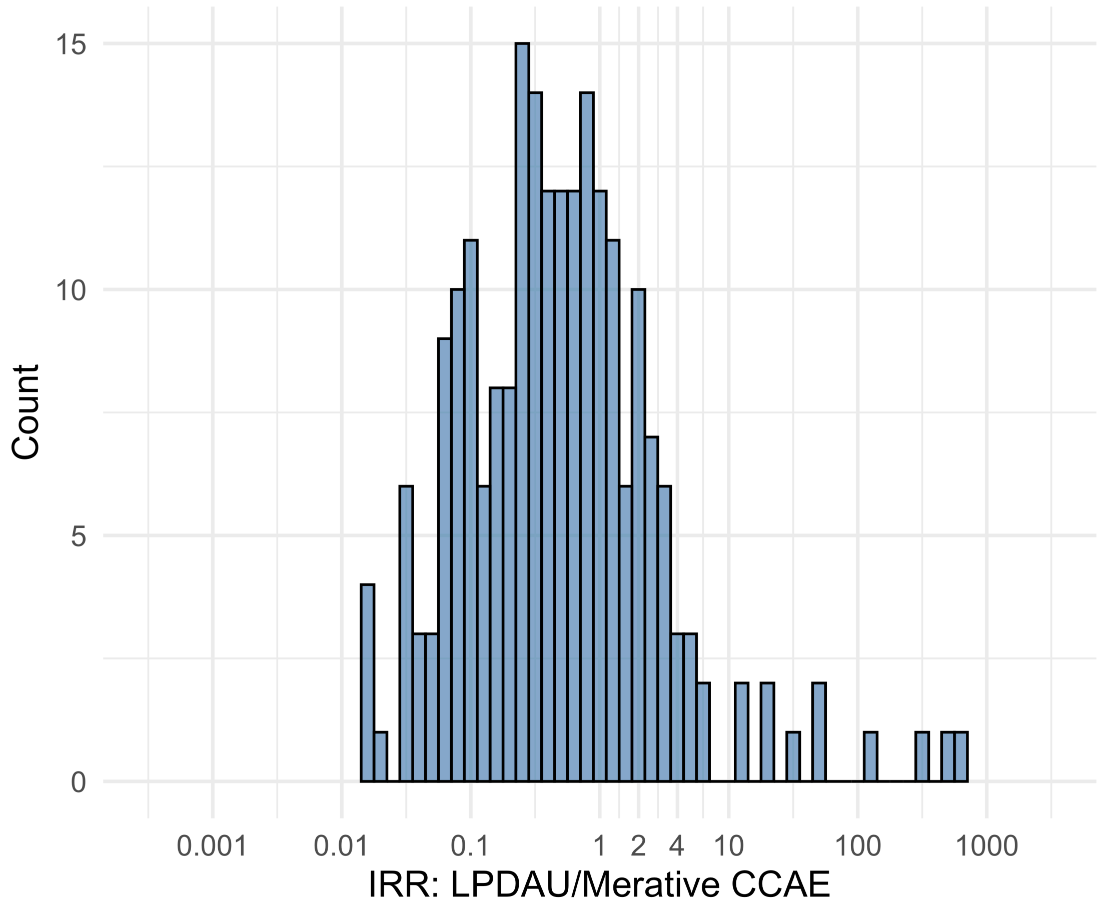


Figure S5.66. IRR distribution comparing LPDAU and Merative CCAE.

*Table S5.66. Most extreme IRRs for LPDAU vs Merative CCAE (rounded to 0.001).*

| source1 | source2 | Drug and Indication | Outcome | IRR |
|---|---|---|---|---|
| LPDAU | Merative CCAE | New users of Trimethoprim systemetic nested in Urinary Tract Infection | Polymorphic Ventricular Tachycardia or Torsades de Pointes | 624.5 |
| LPDAU | Merative CCAE | New users of Beta blockers nested in essential hypertension | Polymorphic Ventricular Tachycardia or Torsades de Pointes | 506.8 |
| LPDAU | Merative CCAE | New users of Cephalosporin systemetic nested in Urinary Tract Infection | Polymorphic Ventricular Tachycardia or Torsades de Pointes | 311.6 |
| LPDAU | Merative CCAE | New users of Beta blockers nested in essential hypertension | Hospitalization with heart failure events | 0.0 |
| LPDAU | Merative CCAE | New users of Beta blockers nested in essential hypertension | Acute Kidney Injury AKI | 0.0 |
| LPDAU | Merative CCAE | New users of Beta blockers nested in essential hypertension | 4-point MACE | 0.0 |

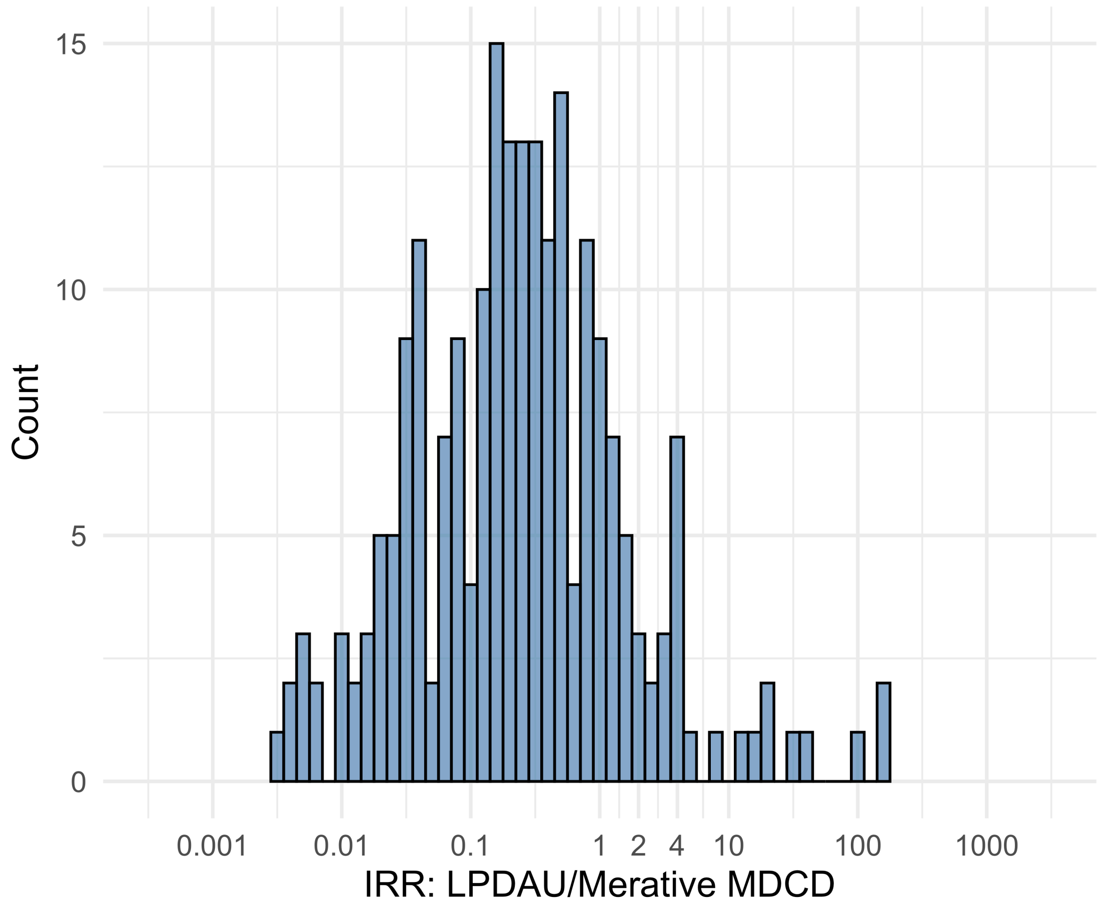


Figure S5.67. IRR distribution comparing LPDAU and Merative MDCD.

*Table S5.67. Most extreme IRRs for LPDAU vs Merative MDCD (rounded to 0.001).*

| source1 | source2 | Drug and Indication | Outcome | IRR |
|---|---|---|---|---|
| LPDAU | Merative MDCD | New users of Trimethoprim systemetic nested in Urinary Tract Infection | Polymorphic Ventricular Tachycardia or Torsades de Pointes | 148.8 |
| LPDAU | Merative MDCD | New users of Trimethoprim systemetic nested in Urinary Tract Infection | Progressive multifocal leukoencephalopathy | 148.8 |
| LPDAU | Merative MDCD | New users of Cephalosporin systemetic nested in Urinary Tract Infection | Polymorphic Ventricular Tachycardia or Torsades de Pointes | 104.0 |
| LPDAU | Merative MDCD | New users of Beta blockers nested in essential hypertension | Hospitalization with heart failure events | 0.0 |
| LPDAU | Merative MDCD | New users of Cephalosporin systemetic nested in Urinary Tract Infection | Hospitalization with heart failure events | 0.0 |
| LPDAU | Merative MDCD | New users of Cephalosporin systemetic nested in Urinary Tract Infection | Acute Kidney Injury AKI | 0.0 |

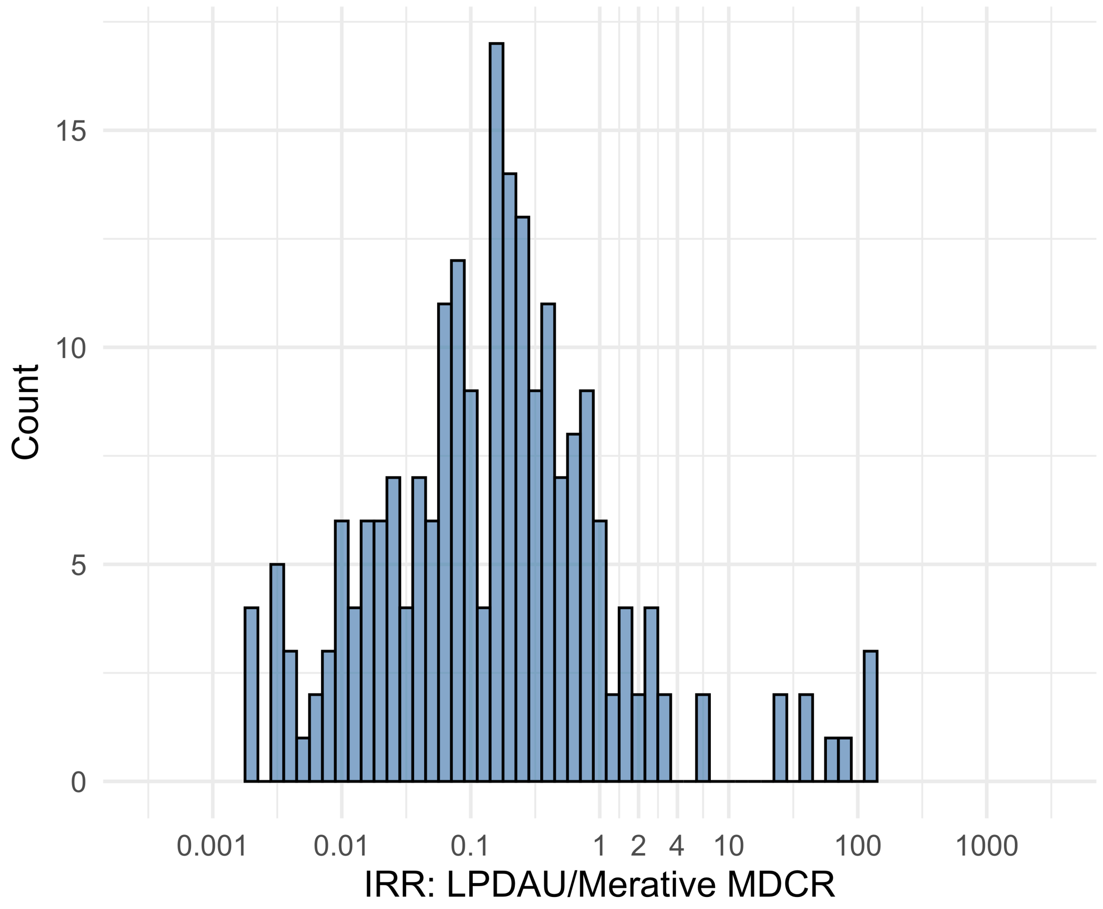


Figure S5.68. IRR distribution comparing LPDAU and Merative MDCR.

*Table S5.68. Most extreme IRRs for LPDAU vs Merative MDCR (rounded to 0.001).*

| source1 | source2 | Drug and Indication | Outcome | IRR |
|---|---|---|---|---|
| LPDAU | Merative MDCR | New users of Beta blockers nested in essential hypertension | Polymorphic Ventricular Tachycardia or Torsades de Pointes | 135.9 |
| LPDAU | Merative MDCR | New users of Beta blockers nested in essential hypertension | All events Drug Rash with Eosinophilia and Systemic Symptoms (DRESS) | 135.9 |
| LPDAU | Merative MDCR | New users of Beta blockers nested in essential hypertension | Progressive multifocal leukoencephalopathy | 135.9 |
| LPDAU | Merative MDCR | New users of Cephalosporin systemetic nested in Urinary Tract Infection | Hospitalization with heart failure events | 0.0 |
| LPDAU | Merative MDCR | New users of Cephalosporin systemetic nested in Urinary Tract Infection | Acute Kidney Injury AKI | 0.0 |
| LPDAU | Merative MDCR | New users of Cephalosporin systemetic nested in Urinary Tract Infection | 4-point MACE | 0.0 |

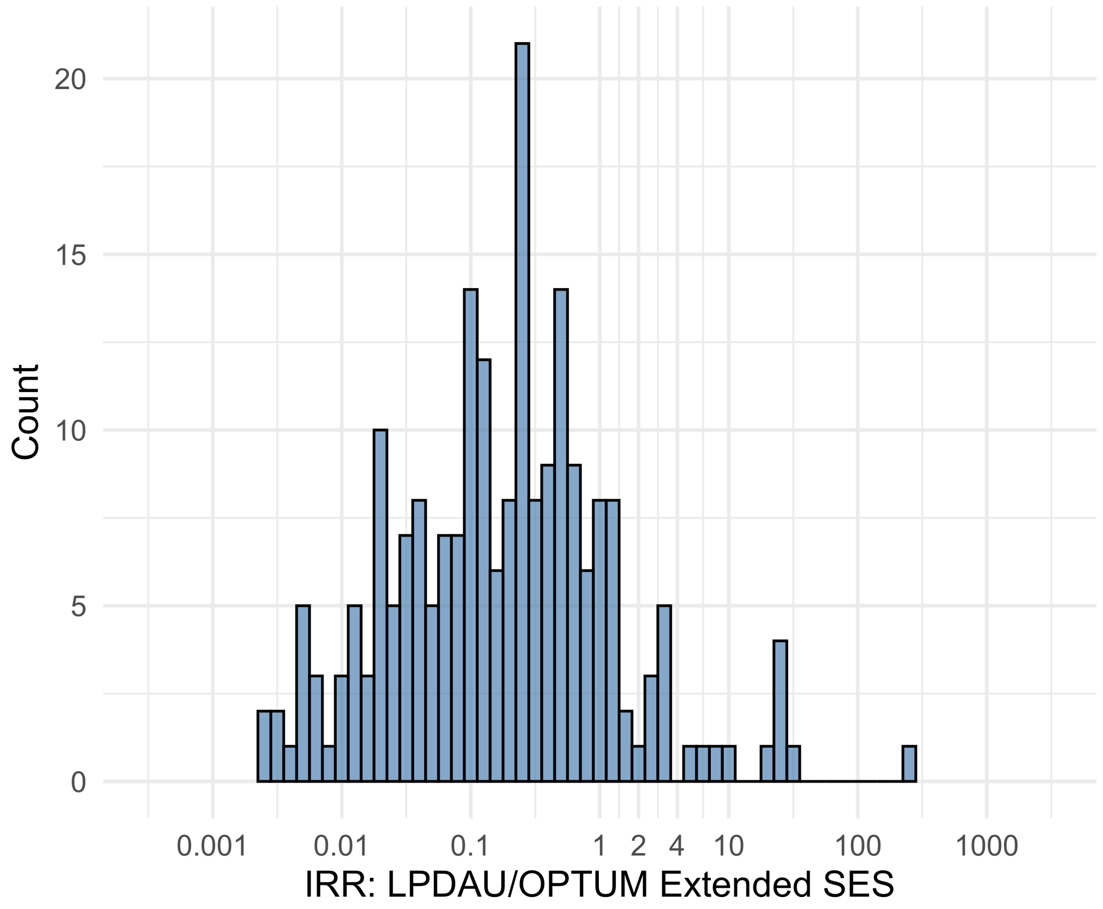


Figure S5.69. IRR distribution comparing LPDAU and OPTUM Extended SES.

*Table S5.69. Most extreme IRRs for LPDAU vs OPTUM Extended SES (rounded to 0.001).*

| source1 | source2 | Drug and Indication | Outcome | IRR |
|---|---|---|---|---|
| LPDAU | OPTUM Extended SES | New users of Trimethoprim systemetic nested in Urinary Tract Infection | Polymorphic Ventricular Tachycardia or Torsades de Pointes | 238.1 |
| LPDAU | OPTUM Extended SES | New users of Beta blockers nested in essential hypertension | Progressive multifocal leukoencephalopathy | 31.9 |
| LPDAU | OPTUM Extended SES | New users of Cephalosporin systemetic nested in Urinary Tract Infection | Polymorphic Ventricular Tachycardia or Torsades de Pointes | 26.3 |
| LPDAU | OPTUM Extended SES | New users of Cephalosporin systemetic nested in Urinary Tract Infection | Acute Kidney Injury AKI | 0.0 |
| LPDAU | OPTUM Extended SES | New users of Cephalosporin systemetic nested in Urinary Tract Infection | Hospitalization with heart failure events | 0.0 |
| LPDAU | OPTUM Extended SES | New users of Cephalosporin systemetic nested in Urinary Tract Infection | 4-point MACE | 0.0 |

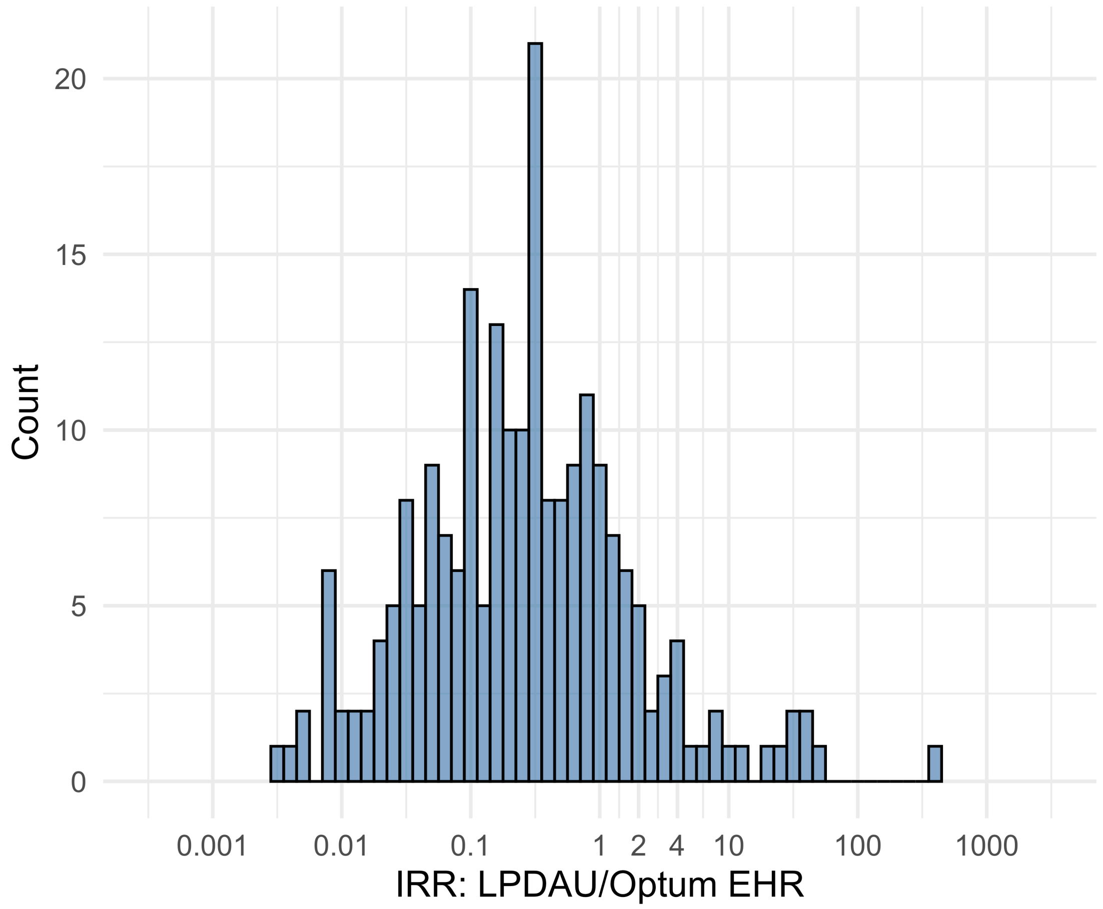


Figure S5.70. IRR distribution comparing LPDAU and Optum EHR.

*Table S5.70. Most extreme IRRs for LPDAU vs Optum EHR (rounded to 0.001).*

| source1 | source2 | Drug and Indication | Outcome | IRR |
|---|---|---|---|---|
| LPDAU | Optum EHR | New users of Trimethoprim systemetic nested in Urinary Tract Infection | Polymorphic Ventricular Tachycardia or Torsades de Pointes | 411.4 |
| LPDAU | Optum EHR | New users of Beta blockers nested in essential hypertension | Progressive multifocal leukoencephalopathy | 50.1 |
| LPDAU | Optum EHR | New users of Beta blockers nested in essential hypertension | All events Drug Rash with Eosinophilia and Systemic Symptoms (DRESS) | 43.5 |
| LPDAU | Optum EHR | New users of Cephalosporin systemetic nested in Urinary Tract Infection | Acute Kidney Injury AKI | 0.0 |
| LPDAU | Optum EHR | New users of Cephalosporin systemetic nested in Urinary Tract Infection | Hospitalization with heart failure events | 0.0 |
| LPDAU | Optum EHR | New users of Cephalosporin systemetic nested in Urinary Tract Infection | 4-point MACE | 0.0 |

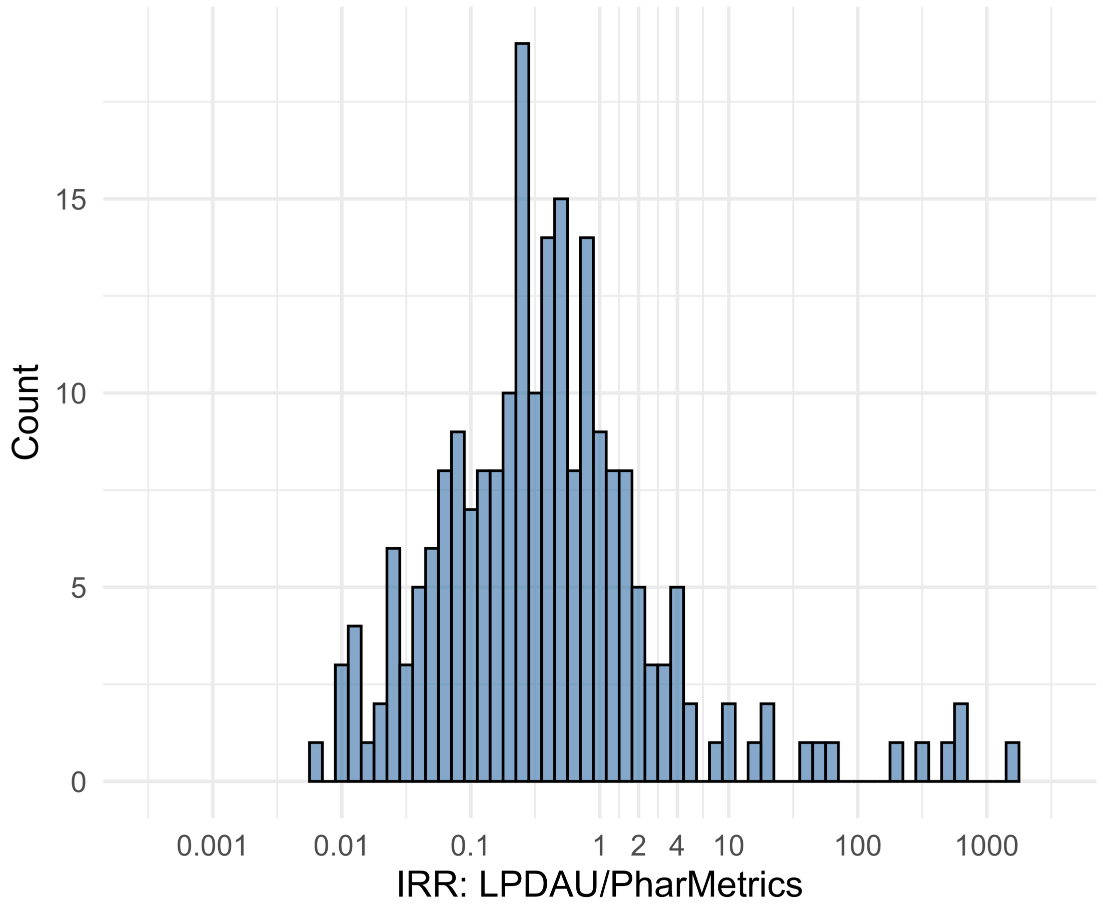


Figure S5.71. IRR distribution comparing LPDAU and PharMetrics.

*Table S5.71. Most extreme IRRs for LPDAU vs PharMetrics (rounded to 0.001).*

| source1 | source2 | Drug and Indication | Outcome | IRR |
|---|---|---|---|---|
| LPDAU | PharMetrics | New users of Beta blockers nested in essential hypertension | Cardiovascular-related mortality | 1534.8 |
| LPDAU | PharMetrics | New users of Trimethoprim systemetic nested in Urinary Tract Infection | Cardiovascular-related mortality | 574.7 |
| LPDAU | PharMetrics | New users of Trimethoprim systemetic nested in Urinary Tract Infection | Polymorphic Ventricular Tachycardia or Torsades de Pointes | 574.7 |
| LPDAU | PharMetrics | New users of Cephalosporin systemetic nested in Urinary Tract Infection | Acute Kidney Injury AKI | 0.0 |
| LPDAU | PharMetrics | New users of Beta blockers nested in essential hypertension | Hospitalization with heart failure events | 0.0 |
| LPDAU | PharMetrics | New users of Cephalosporin systemetic nested in Urinary Tract Infection | Hospitalization with heart failure events | 0.0 |

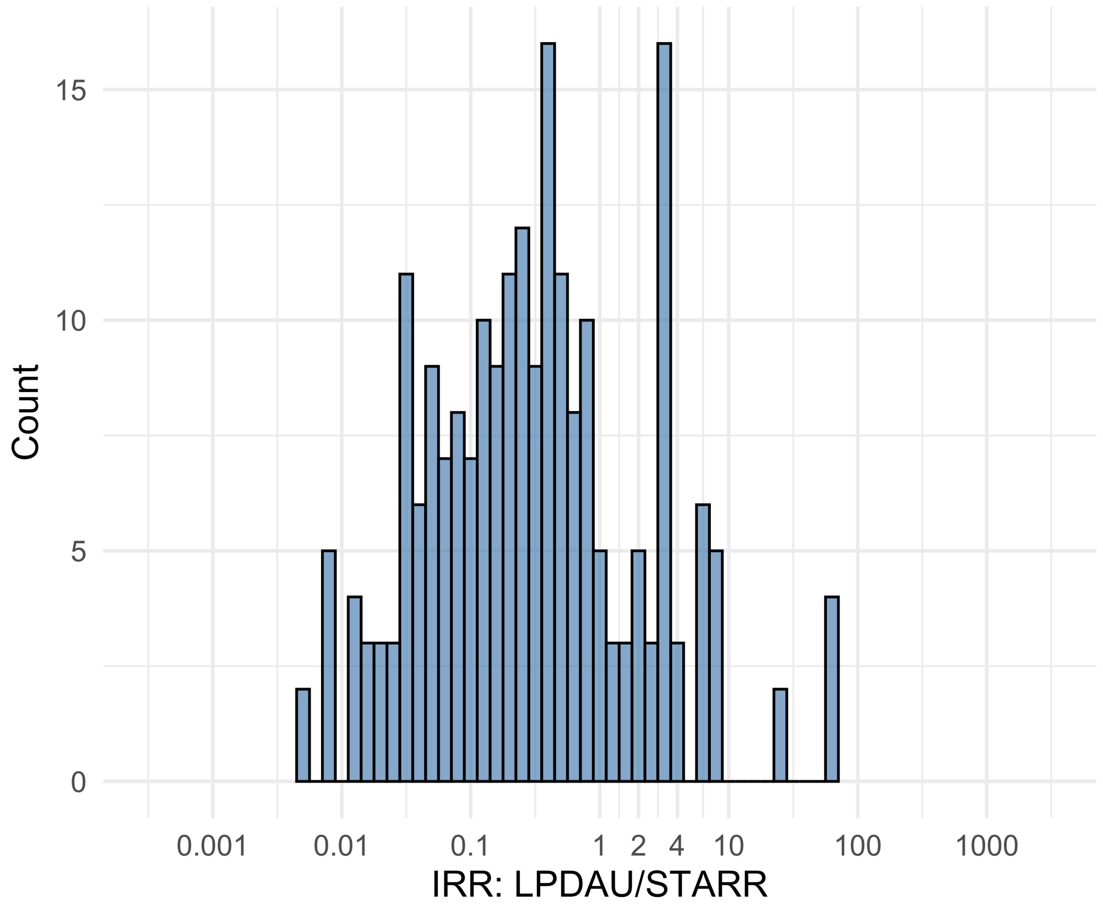


Figure S5.72. IRR distribution comparing LPDAU and STARR.

*Table S5.72. Most extreme IRRs for LPDAU vs STARR (rounded to 0.001).*

| source1 | source2 | Drug and Indication | Outcome | IRR |
|---|---|---|---|---|
| LPDAU | STARR | New users of Beta blockers nested in essential hypertension | Febrile Neutropenia or Neutropenic Fever | 64.1 |
| LPDAU | STARR | New users of Beta blockers nested in essential hypertension | Polymorphic Ventricular Tachycardia or Torsades de Pointes | 64.1 |
| LPDAU | STARR | New users of Beta blockers nested in essential hypertension | All events Drug Rash with Eosinophilia and Systemic Symptoms (DRESS) | 64.1 |
| LPDAU | STARR | New users of Cephalosporin systemetic nested in Urinary Tract Infection | Hospitalization with heart failure events | 0.0 |
| LPDAU | STARR | New users of Cephalosporin systemetic nested in Urinary Tract Infection | Acute Kidney Injury AKI | 0.0 |
| LPDAU | STARR | New users of Trimethoprim systemetic nested in Urinary Tract Infection | Hospitalization with heart failure events | 0.0 |

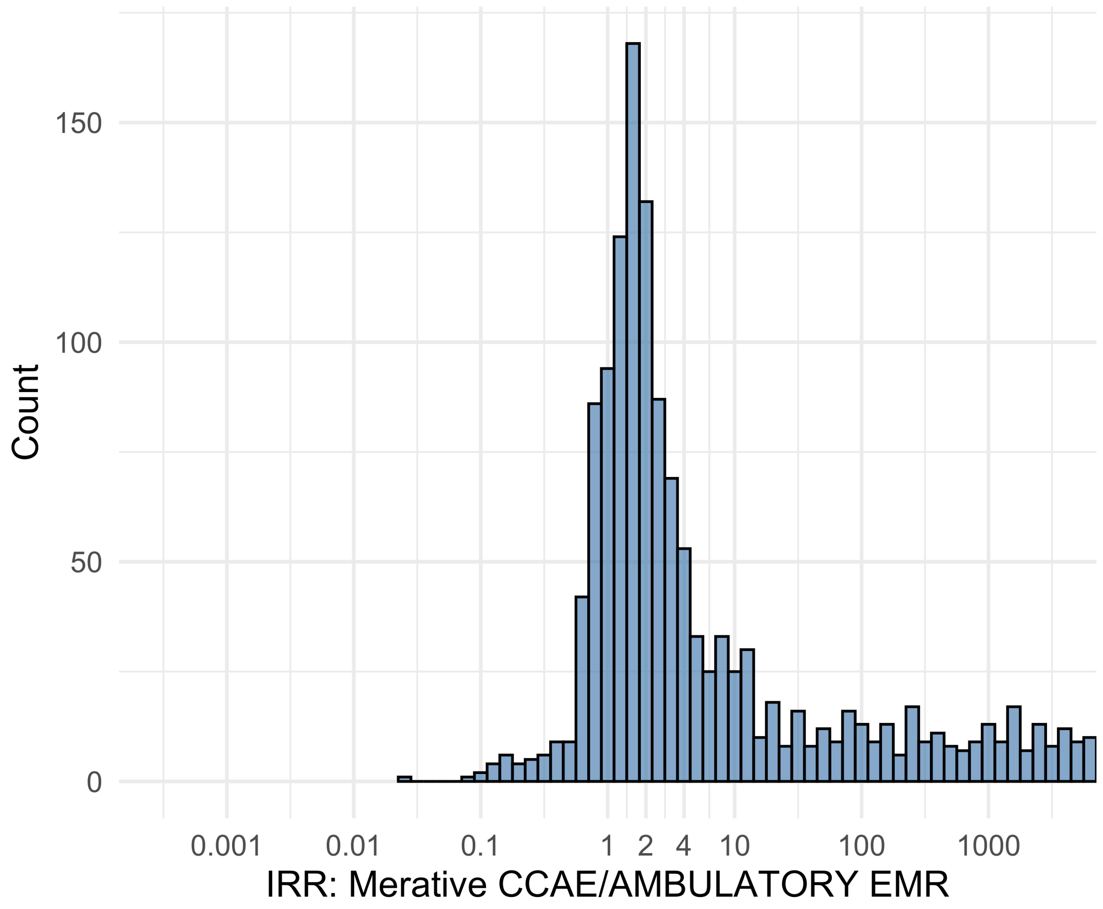


Figure S5.73. IRR distribution comparing Merative CCAE and AMBULATORY EMR.

*Table S5.73. Most extreme IRRs for Merative CCAE vs AMBULATORY EMR (rounded to 0.001).*

| source1 | source2 | Drug and Indication | Outcome | IRR |
|---|---|---|---|---|
| Merative CCAE | AMBULATORY EMR | New users of Beta blockers nested in essential hypertension | Hospitalization with heart failure events | 121646.9 |
| Merative CCAE | AMBULATORY EMR | New users of Beta blockers nested in essential hypertension | 4-point MACE | 113336.0 |
| Merative CCAE | AMBULATORY EMR | New users of Beta blockers nested in essential hypertension | Total cardiovascular disease events (ischemic stroke, hemorrhagic stroke, heart failure, acute myocardial infarction or sudden cardiac death) | 112041.4 |
| Merative CCAE | AMBULATORY EMR | New users of Beta blockers nested in essential hypertension | Polymorphic Ventricular Tachycardia or Torsades de Pointes | 0.0 |
| Merative CCAE | AMBULATORY EMR | New users of SGLT2 inhibitor nested in Left Heart Failure | Anaphylaxis | 0.1 |
| Merative CCAE | AMBULATORY EMR | New users of Beta blockers nested in Left Heart Failure | Polymorphic Ventricular Tachycardia or Torsades de Pointes | 0.1 |

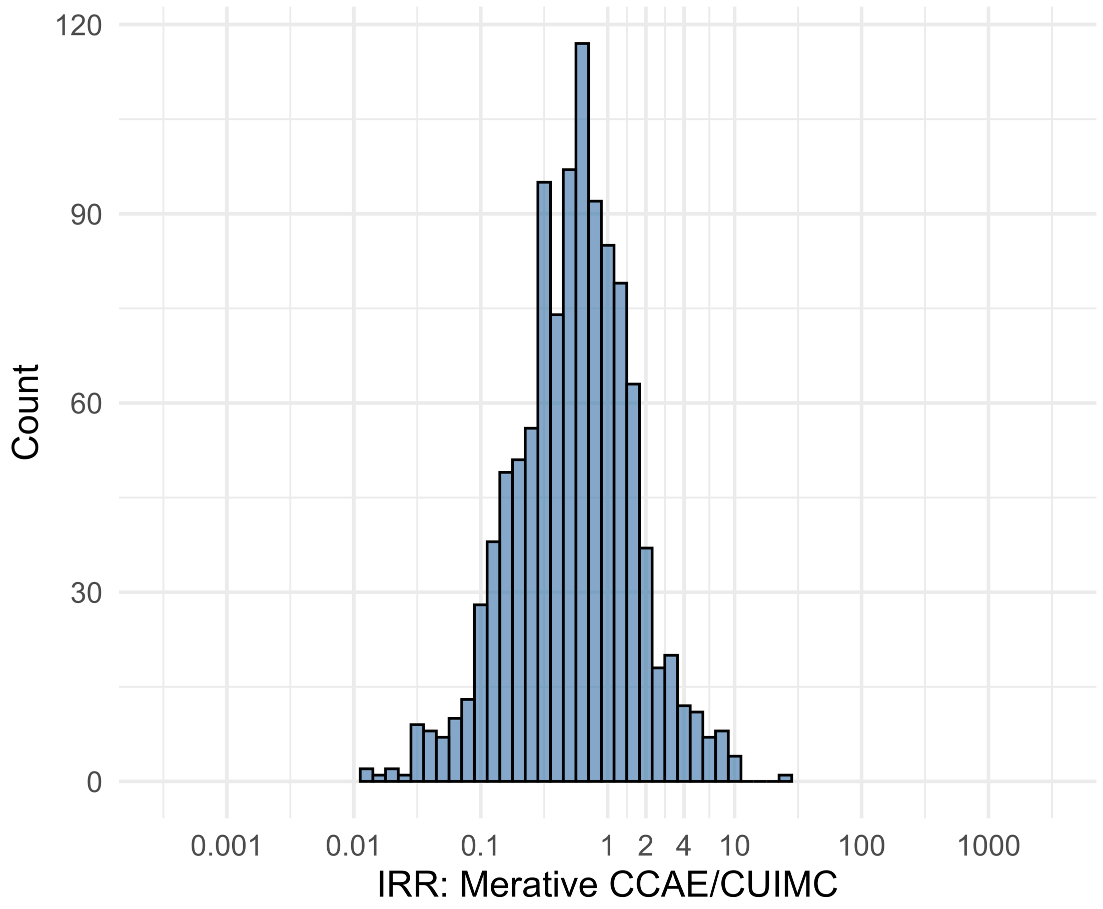


Figure S5.74. IRR distribution comparing Merative CCAE and CUIMC.

*Table S5.74. Most extreme IRRs for Merative CCAE vs CUIMC (rounded to 0.001).*

| source1 | source2 | Drug and Indication | Outcome | IRR |
|---|---|---|---|---|
| Merative CCAE | CUIMC | New users of Beta blockers nested in Acute Myocardial Infarction | Abnormal weight gain events | 25.4 |
| Merative CCAE | CUIMC | New users of Cephalosporin systemetic nested in Acute Typical Pneumonia | Abnormal weight gain events | 11.2 |
| Merative CCAE | CUIMC | New users of Tumor Necrosis Factor alpha (TNFa) inhibitors nested in Rheumatoid arthritis | All events of Acute Liver Injury | 10.5 |
| Merative CCAE | CUIMC | New users of Trimethoprim systemetic nested in Urinary Tract Infection | Polymorphic Ventricular Tachycardia or Torsades de Pointes | 0.0 |
| Merative CCAE | CUIMC | New users of Fluoroquinolone systemic nested in Urinary Tract Infection | Polymorphic Ventricular Tachycardia or Torsades de Pointes | 0.0 |
| Merative CCAE | CUIMC | New users of Trimethoprim systemetic nested in Urinary Tract Infection | Cardiovascular-related mortality | 0.0 |

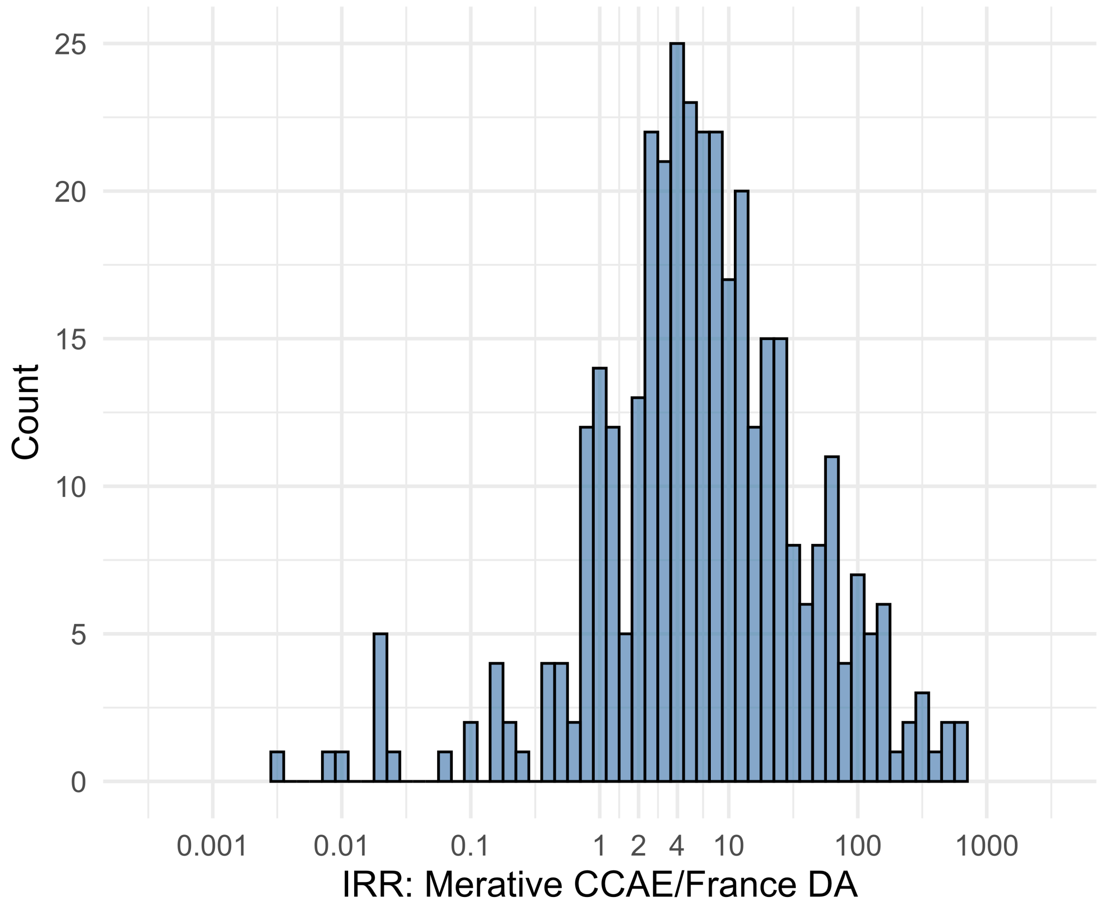


Figure S5.75. IRR distribution comparing Merative CCAE and France DA.

*Table S5.75. Most extreme IRRs for Merative CCAE vs France DA (rounded to 0.001).*

| source1 | source2 | Drug and Indication | Outcome | IRR |
|---|---|---|---|---|
| Merative CCAE | France DA | New users of Beta blockers nested in essential hypertension | Hospitalization with heart failure events | 597.6 |
| Merative CCAE | France DA | New users of Beta blockers nested in essential hypertension | Acute Kidney Injury AKI | 573.7 |
| Merative CCAE | France DA | New users of Beta blockers nested in essential hypertension | 4-point MACE | 556.8 |
| Merative CCAE | France DA | New users of Trimethoprim systemetic nested in Urinary Tract Infection | Polymorphic Ventricular Tachycardia or Torsades de Pointes | 0.0 |
| Merative CCAE | France DA | New users of Fluoroquinolone systemic nested in Urinary Tract Infection | Polymorphic Ventricular Tachycardia or Torsades de Pointes | 0.0 |
| Merative CCAE | France DA | New users of Cephalosporin systemetic nested in Urinary Tract Infection | Polymorphic Ventricular Tachycardia or Torsades de Pointes | 0.0 |

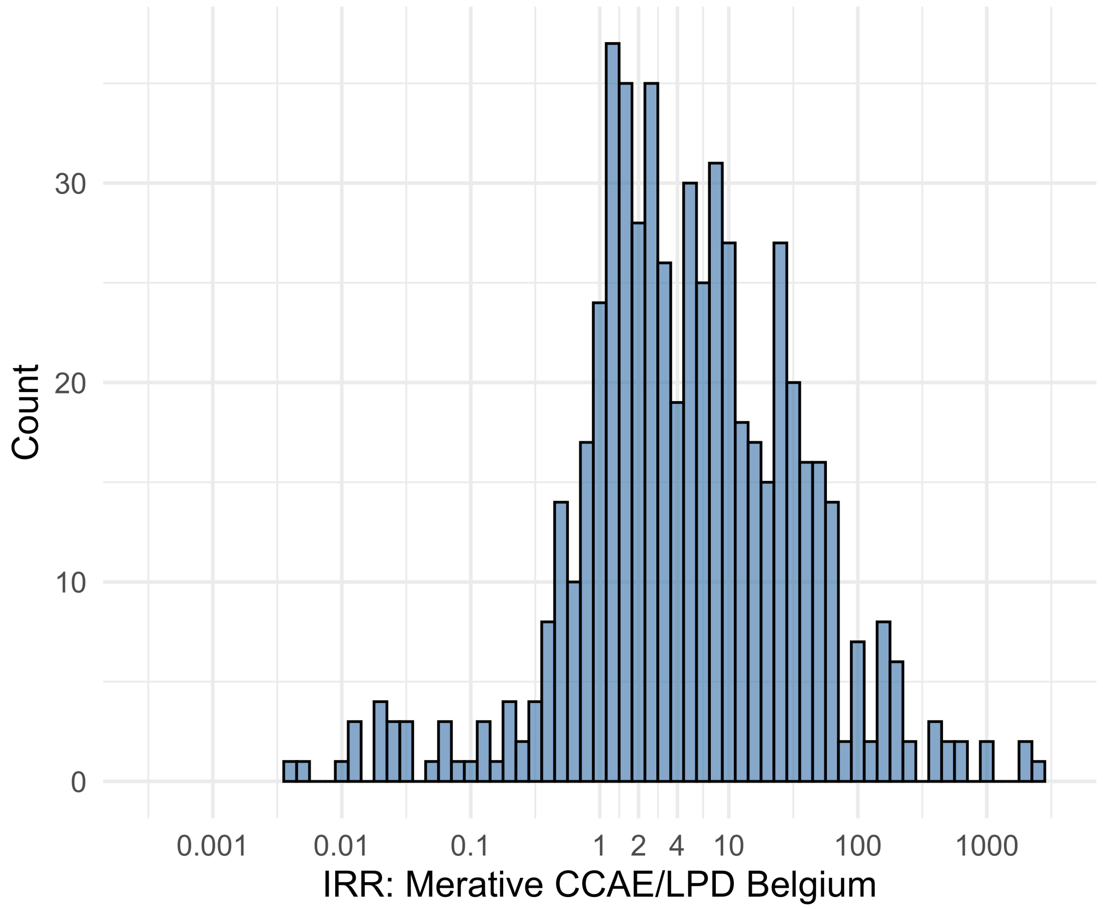


Figure S5.76. IRR distribution comparing Merative CCAE and LPD Belgium.

*Table S5.76. Most extreme IRRs for Merative CCAE vs LPD Belgium (rounded to 0.001).*

| source1 | source2 | Drug and Indication | Outcome | IRR |
|---|---|---|---|---|
| Merative CCAE | LPD Belgium | New users of Beta blockers nested in essential hypertension | Hospitalization with heart failure events | 2325.7 |
| Merative CCAE | LPD Belgium | New users of Beta blockers nested in essential hypertension | 4-point MACE | 2166.8 |
| Merative CCAE | LPD Belgium | New users of Beta blockers nested in essential hypertension | Total cardiovascular disease events (ischemic stroke, hemorrhagic stroke, heart failure, acute myocardial infarction or sudden cardiac death) | 2142.0 |
| Merative CCAE | LPD Belgium | New users of Trimethoprim systemetic nested in Urinary Tract Infection | Polymorphic Ventricular Tachycardia or Torsades de Pointes | 0.0 |
| Merative CCAE | LPD Belgium | New users of Cephalosporin systemetic nested in Urinary Tract Infection | Polymorphic Ventricular Tachycardia or Torsades de Pointes | 0.0 |
| Merative CCAE | LPD Belgium | New users of Cephalosporin systemetic nested in Urinary Tract Infection | Progressive multifocal leukoencephalopathy | 0.0 |

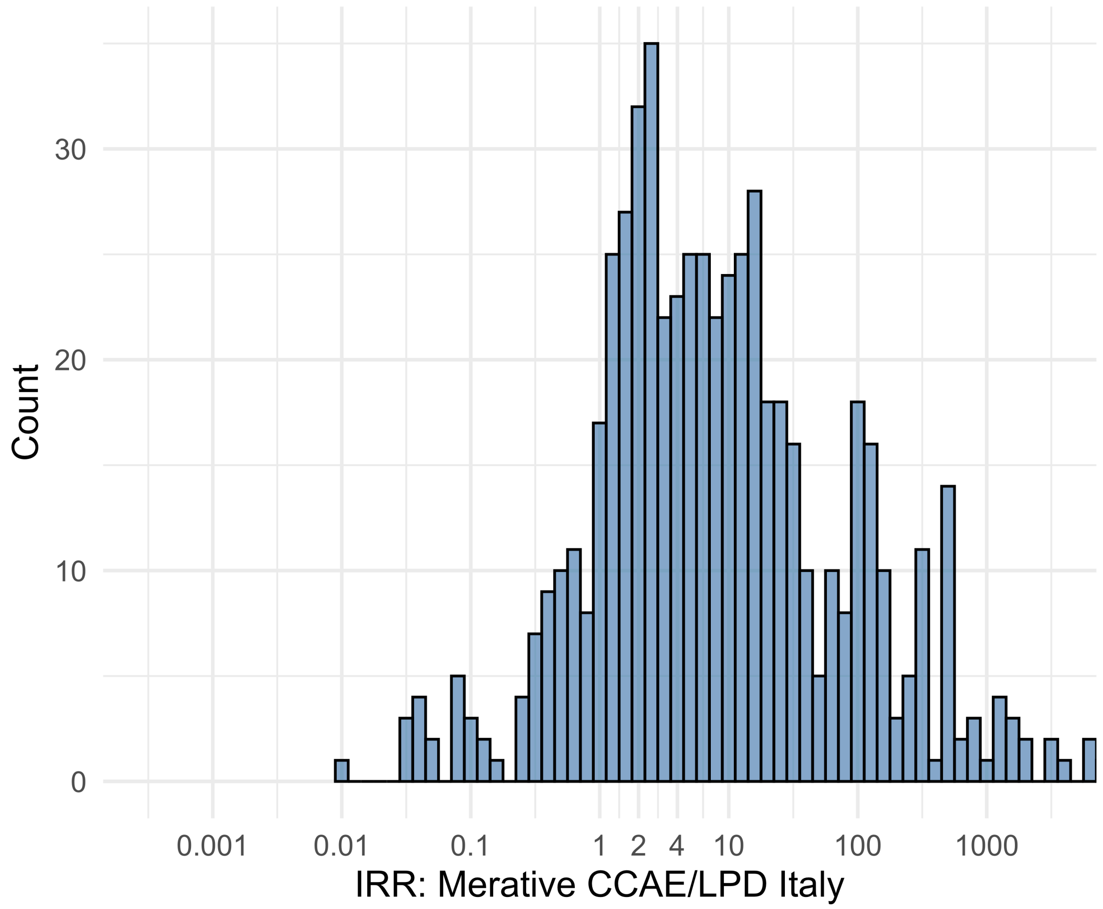


Figure S5.77. IRR distribution comparing Merative CCAE and LPD Italy.

*Table S5.77. Most extreme IRRs for Merative CCAE vs LPD Italy (rounded to 0.001).*

| source1 | source2 | Drug and Indication | Outcome | IRR |
|---|---|---|---|---|
| Merative CCAE | LPD Italy | New users of Beta blockers nested in essential hypertension | Hospitalization with heart failure events | 7561.4 |
| Merative CCAE | LPD Italy | New users of Beta blockers nested in essential hypertension | 4-point MACE | 7044.8 |
| Merative CCAE | LPD Italy | New users of Beta blockers nested in essential hypertension | Total cardiovascular disease events (ischemic stroke, hemorrhagic stroke, heart failure, acute myocardial infarction or sudden cardiac death) | 6964.3 |
| Merative CCAE | LPD Italy | New users of Cephalosporin systemetic nested in Urinary Tract Infection | Progressive multifocal leukoencephalopathy | 0.0 |
| Merative CCAE | LPD Italy | New users of GLP-1 receptor antagonists nested in obesity | Progressive multifocal leukoencephalopathy | 0.0 |
| Merative CCAE | LPD Italy | New users of GLP-1 receptor antagonists nested in obesity | Polymorphic Ventricular Tachycardia or Torsades de Pointes | 0.0 |

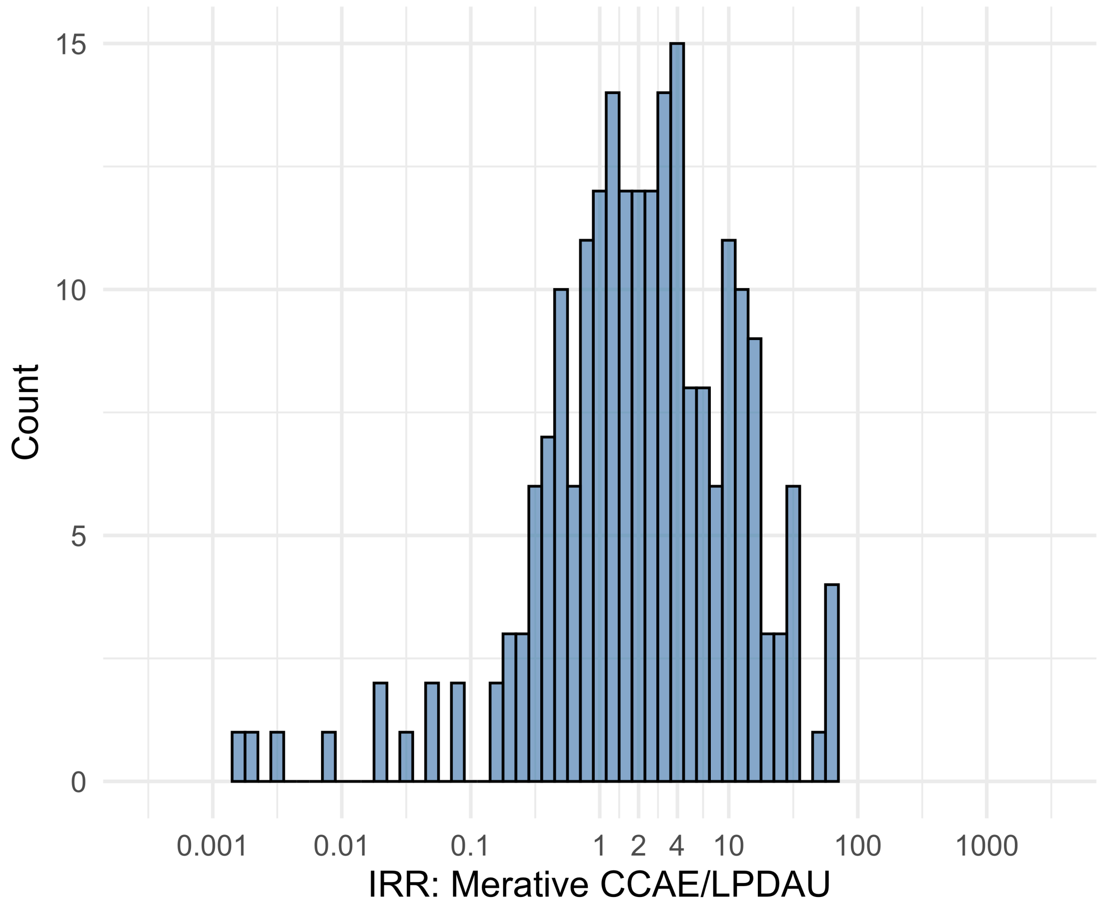


Figure S5.78. IRR distribution comparing Merative CCAE and LPDAU.

*Table S5.78. Most extreme IRRs for Merative CCAE vs LPDAU (rounded to 0.001).*

| source1 | source2 | Drug and Indication | Outcome | IRR |
|---|---|---|---|---|
| Merative CCAE | LPDAU | New users of Beta blockers nested in essential hypertension | Hospitalization with heart failure events | 65.4 |
| Merative CCAE | LPDAU | New users of Beta blockers nested in essential hypertension | Acute Kidney Injury AKI | 62.8 |
| Merative CCAE | LPDAU | New users of Beta blockers nested in essential hypertension | 4-point MACE | 61.0 |
| Merative CCAE | LPDAU | New users of Trimethoprim systemetic nested in Urinary Tract Infection | Polymorphic Ventricular Tachycardia or Torsades de Pointes | 0.0 |
| Merative CCAE | LPDAU | New users of Beta blockers nested in essential hypertension | Polymorphic Ventricular Tachycardia or Torsades de Pointes | 0.0 |
| Merative CCAE | LPDAU | New users of Cephalosporin systemetic nested in Urinary Tract Infection | Polymorphic Ventricular Tachycardia or Torsades de Pointes | 0.0 |

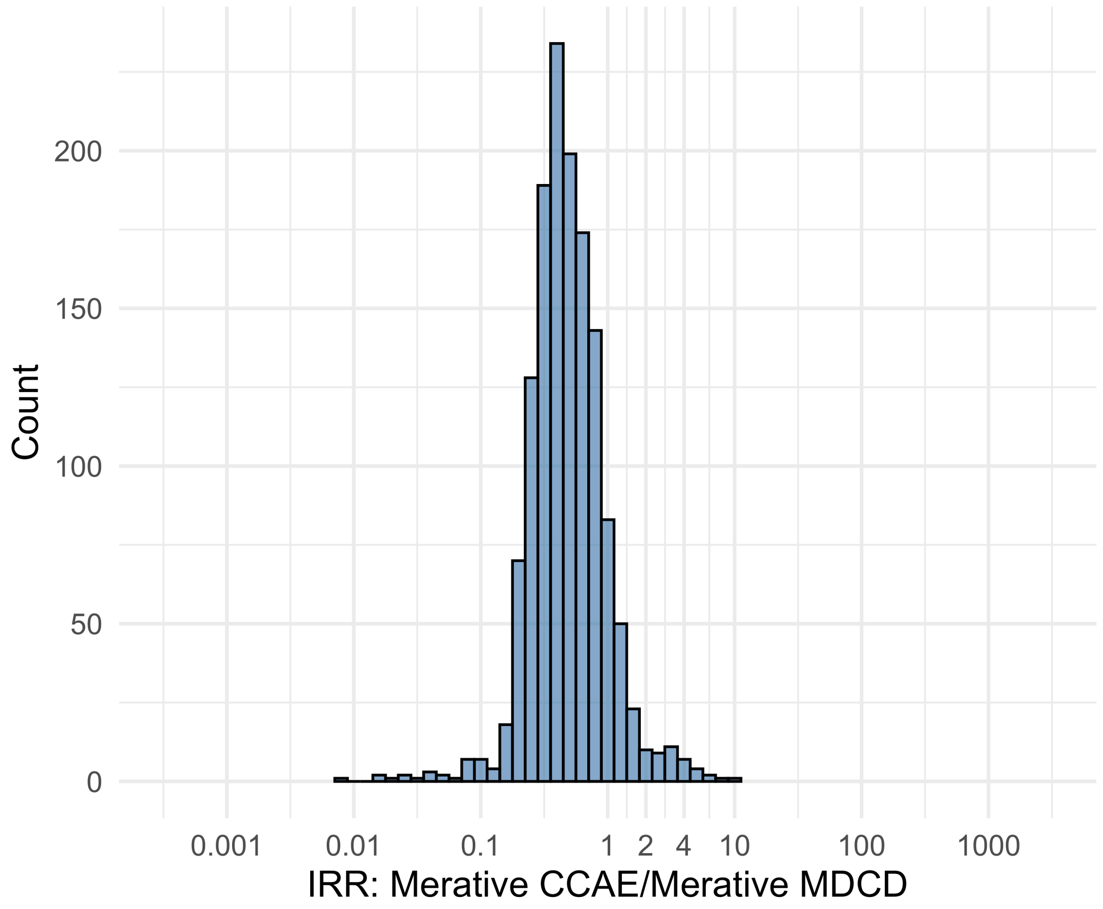


Figure S5.79. IRR distribution comparing Merative CCAE and Merative MDCD.

*Table S5.79. Most extreme IRRs for Merative CCAE vs Merative MDCD (rounded to 0.001).*

| source1 | source2 | Drug and Indication | Outcome | IRR |
|---|---|---|---|---|
| Merative CCAE | Merative MDCD | New users of Tumor Necrosis Factor alpha (TNFa) inhibitors nested in Ulcerative colitis | Persons with gout | 8.9 |
| Merative CCAE | Merative MDCD | New users of Trimethoprim systemetic nested in Acute Typical Pneumonia | All events of Autoimmune hepatitis, with a washout period of 365 days | 7.3 |
| Merative CCAE | Merative MDCD | New users of SGLT2 inhibitor nested in Left Heart Failure | All events of Autoimmune hepatitis, with a washout period of 365 days | 6.0 |
| Merative CCAE | Merative MDCD | New users of JAK inhibitors nested in Rheumatoid arthritis | Cardiovascular-related mortality | 0.0 |
| Merative CCAE | Merative MDCD | New users of Tumor Necrosis Factor alpha (TNFa) inhibitors nested in Psoriatic Arthritis | Lower extremity amputation | 0.0 |
| Merative CCAE | Merative MDCD | New users of Tumor Necrosis Factor alpha (TNFa) inhibitors nested in Psoriatic Arthritis | Cardiovascular-related mortality | 0.0 |

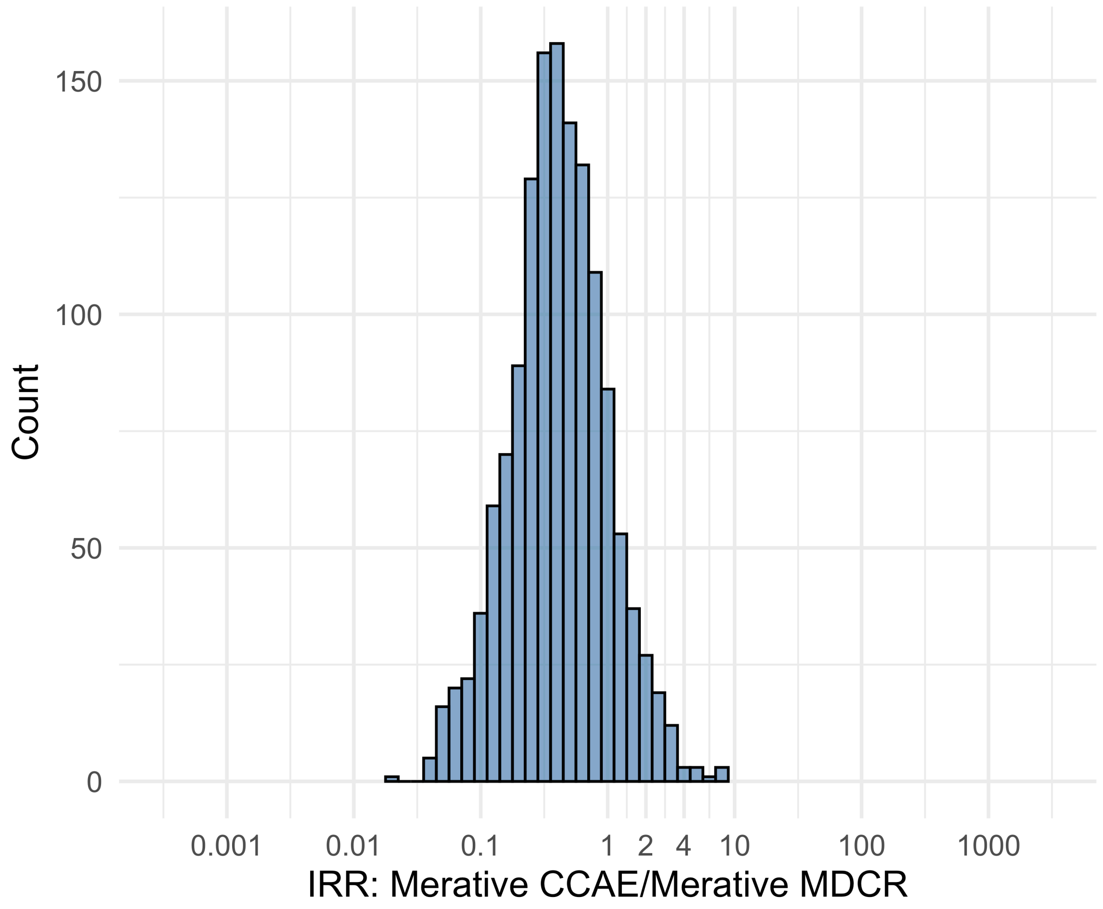


Figure S5.80. IRR distribution comparing Merative CCAE and Merative MDCR.

*Table S5.80. Most extreme IRRs for Merative CCAE vs Merative MDCR (rounded to 0.001).*

| source1 | source2 | Drug and Indication | Outcome | IRR |
|---|---|---|---|---|
| Merative CCAE | Merative MDCR | New users of Tumor Necrosis Factor alpha (TNFa) inhibitors nested in Rheumatoid arthritis | Anaphylaxis | 8.8 |
| Merative CCAE | Merative MDCR | New users of Tumor Necrosis Factor alpha (TNFa) inhibitors nested in Rheumatoid arthritis | Bells Palsy | 8.2 |
| Merative CCAE | Merative MDCR | New users of Tumor Necrosis Factor alpha (TNFa) inhibitors nested in Rheumatoid arthritis | Myocarditis Pericarditis | 8.1 |
| Merative CCAE | Merative MDCR | New users of JAK inhibitors nested in Rheumatoid arthritis | Cardiovascular-related mortality | 0.0 |
| Merative CCAE | Merative MDCR | New users of Trimethoprim systemetic nested in Urinary Tract Infection | Hospitalization with heart failure events | 0.0 |
| Merative CCAE | Merative MDCR | New users of Trimethoprim systemetic nested in Urinary Tract Infection | Cardiovascular-related mortality | 0.0 |

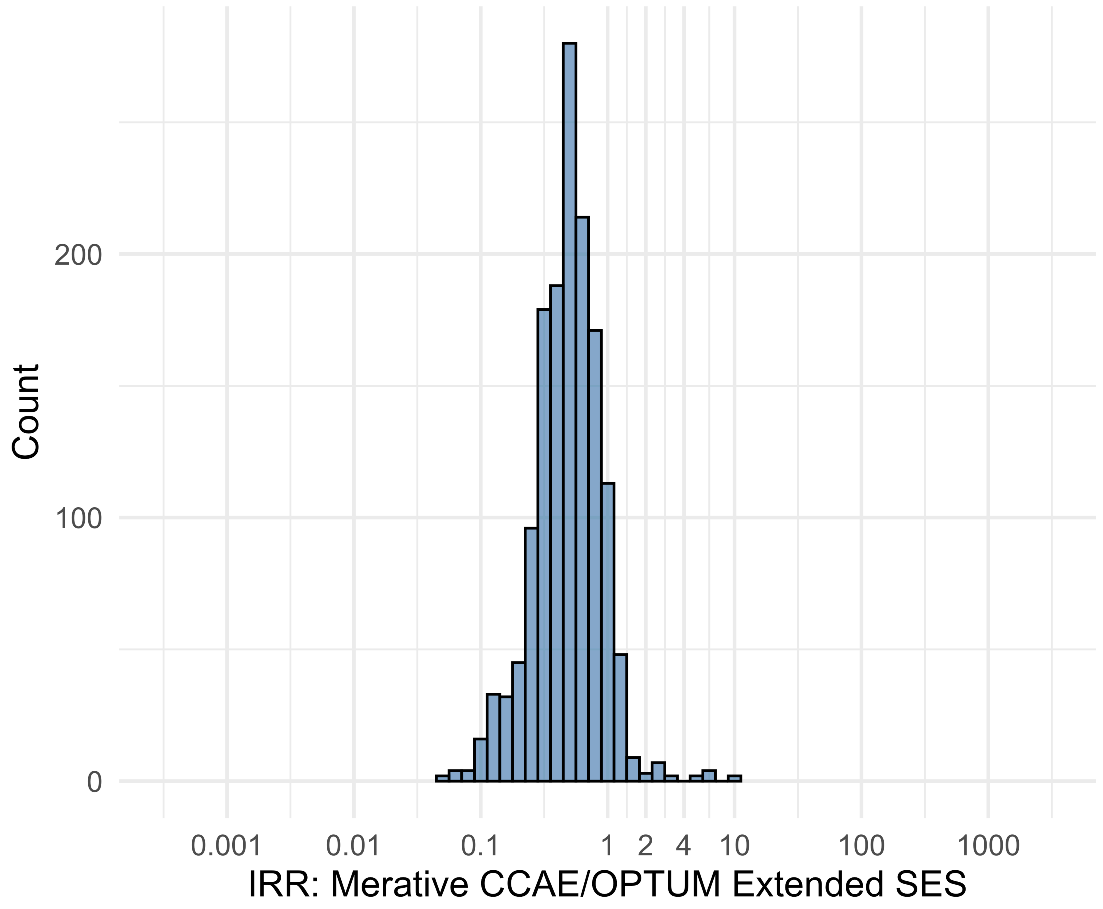


Figure S5.81. IRR distribution comparing Merative CCAE and OPTUM Extended SES.

*Table S5.81. Most extreme IRRs for Merative CCAE vs OPTUM Extended SES (rounded to 0.001).*

| source1 | source2 | Drug and Indication | Outcome | IRR |
|---|---|---|---|---|
| Merative CCAE | OPTUM Extended SES | New users of Trimethoprim systemetic nested in Acute Typical Pneumonia | Progressive multifocal leukoencephalopathy | 10.8 |
| Merative CCAE | OPTUM Extended SES | New users of Tumor Necrosis Factor alpha (TNFa) inhibitors nested in Rheumatoid arthritis | Cardiovascular-related mortality | 10.1 |
| Merative CCAE | OPTUM Extended SES | New users of Tumor Necrosis Factor alpha (TNFa) inhibitors nested in Psoriatic Arthritis | Isolated Immune Thrombocytopenia (ITP), with a washout period of 365 days | 6.9 |
| Merative CCAE | OPTUM Extended SES | New users of Beta blockers nested in essential hypertension | Polymorphic Ventricular Tachycardia or Torsades de Pointes | 0.0 |
| Merative CCAE | OPTUM Extended SES | New users of SGLT2 inhibitor nested in Type 2 diabetes mellitus | Polymorphic Ventricular Tachycardia or Torsades de Pointes | 0.1 |
| Merative CCAE | OPTUM Extended SES | New users of Tumor Necrosis Factor alpha (TNFa) inhibitors nested in Psoriatic Arthritis | Lower extremity amputation | 0.1 |

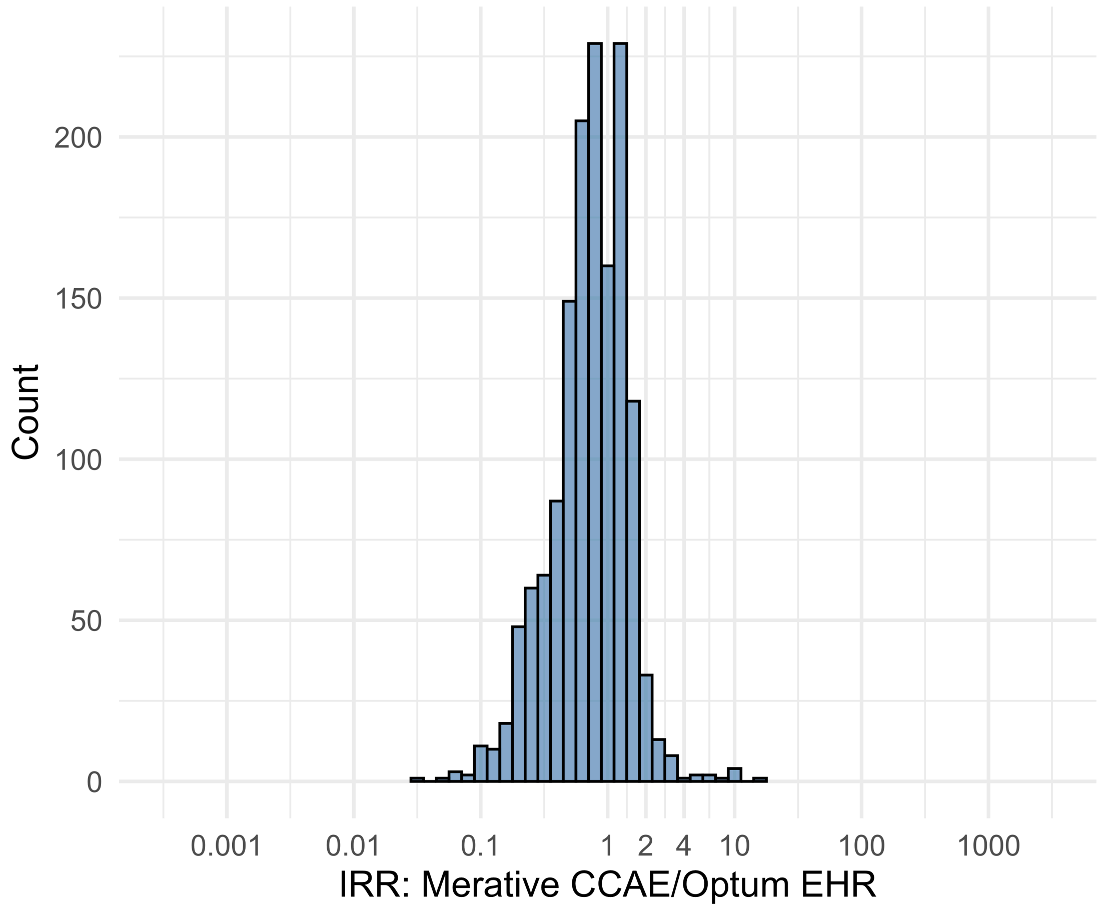


Figure S5.82. IRR distribution comparing Merative CCAE and Optum EHR.

*Table S5.82. Most extreme IRRs for Merative CCAE vs Optum EHR (rounded to 0.001).*

| source1 | source2 | Drug and Indication | Outcome | IRR |
|---|---|---|---|---|
| Merative CCAE | Optum EHR | New users of Tumor Necrosis Factor alpha (TNFa) inhibitors nested in Rheumatoid arthritis | Earliest event of Thrombotic microangiopathy (TMA) or Microangiopathic hemolytic anemia (MAHA) | 17.5 |
| Merative CCAE | Optum EHR | New users of Tumor Necrosis Factor alpha (TNFa) inhibitors nested in Rheumatoid arthritis | Stevens-Johnson syndrome, toxic epidermal necrolysis spectrum | 9.7 |
| Merative CCAE | Optum EHR | New users of Tumor Necrosis Factor alpha (TNFa) inhibitors nested in Ulcerative colitis | Earliest event of Thrombotic microangiopathy (TMA) or Microangiopathic hemolytic anemia (MAHA) | 9.2 |
| Merative CCAE | Optum EHR | New users of Beta blockers nested in Acute Myocardial Infarction | Thrombosis with Thrombocytopenia (TWT) | 0.0 |
| Merative CCAE | Optum EHR | New users of Cephalosporin systemetic nested in Urinary Tract Infection | Thrombosis with Thrombocytopenia (TWT) | 0.1 |
| Merative CCAE | Optum EHR | New users of Fluoroquinolone systemic nested in Urinary Tract Infection | Thrombosis with Thrombocytopenia (TWT) | 0.1 |

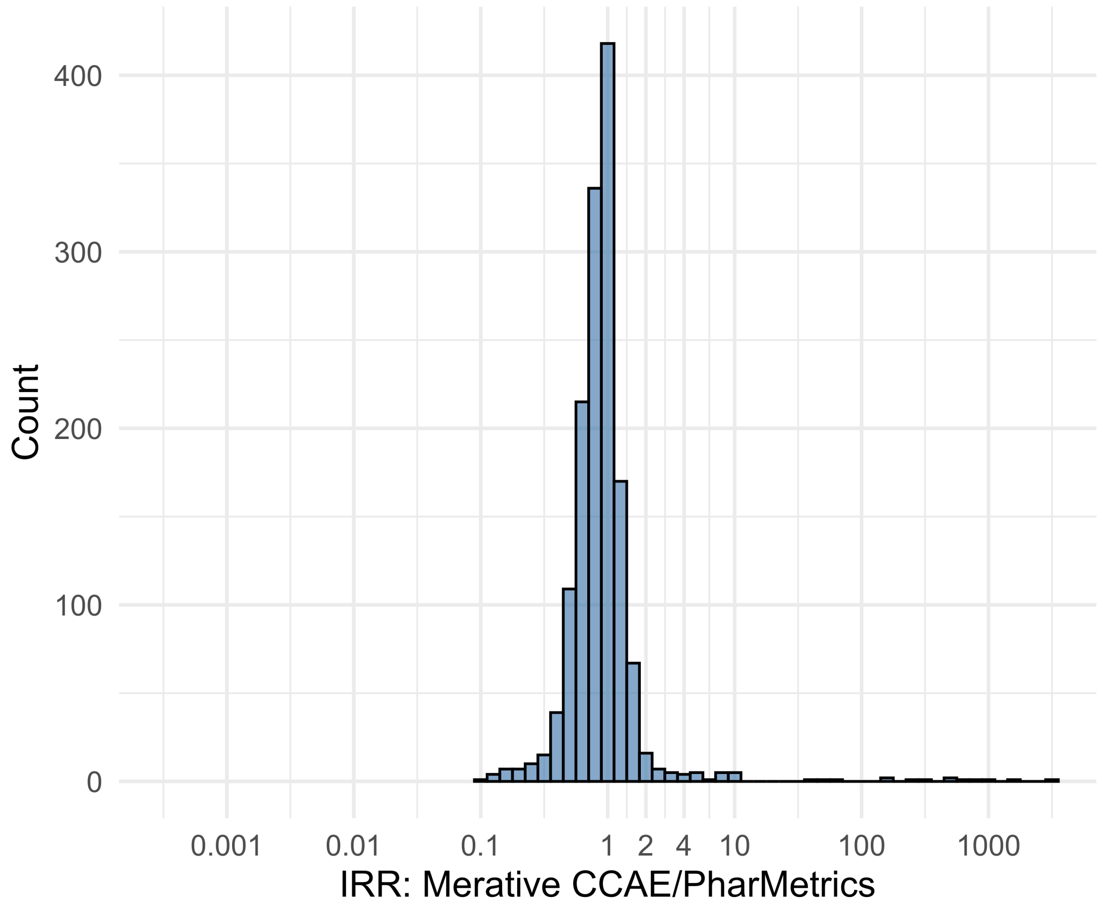


Figure S5.83. IRR distribution comparing Merative CCAE and PharMetrics.

*Table S5.83. Most extreme IRRs for Merative CCAE vs PharMetrics (rounded to 0.001).*

| source1 | source2 | Drug and Indication | Outcome | IRR |
|---|---|---|---|---|
| Merative CCAE | PharMetrics | New users of Beta blockers nested in essential hypertension | Cardiovascular-related mortality | 2842.4 |
| Merative CCAE | PharMetrics | New users of Beta blockers nested in Left Heart Failure | Cardiovascular-related mortality | 1679.2 |
| Merative CCAE | PharMetrics | New users of Fluoroquinolone systemic nested in Acute Typical Pneumonia | Cardiovascular-related mortality | 1014.0 |
| Merative CCAE | PharMetrics | New users of Tumor Necrosis Factor alpha (TNFa) inhibitors nested in Psoriatic Arthritis | Renal Cancer | 0.1 |
| Merative CCAE | PharMetrics | New users of Tumor Necrosis Factor alpha (TNFa) inhibitors nested in Psoriatic Arthritis | Lower extremity amputation | 0.1 |
| Merative CCAE | PharMetrics | New users of Cephalosporin systemetic nested in Urinary Tract Infection | Progressive multifocal leukoencephalopathy | 0.1 |

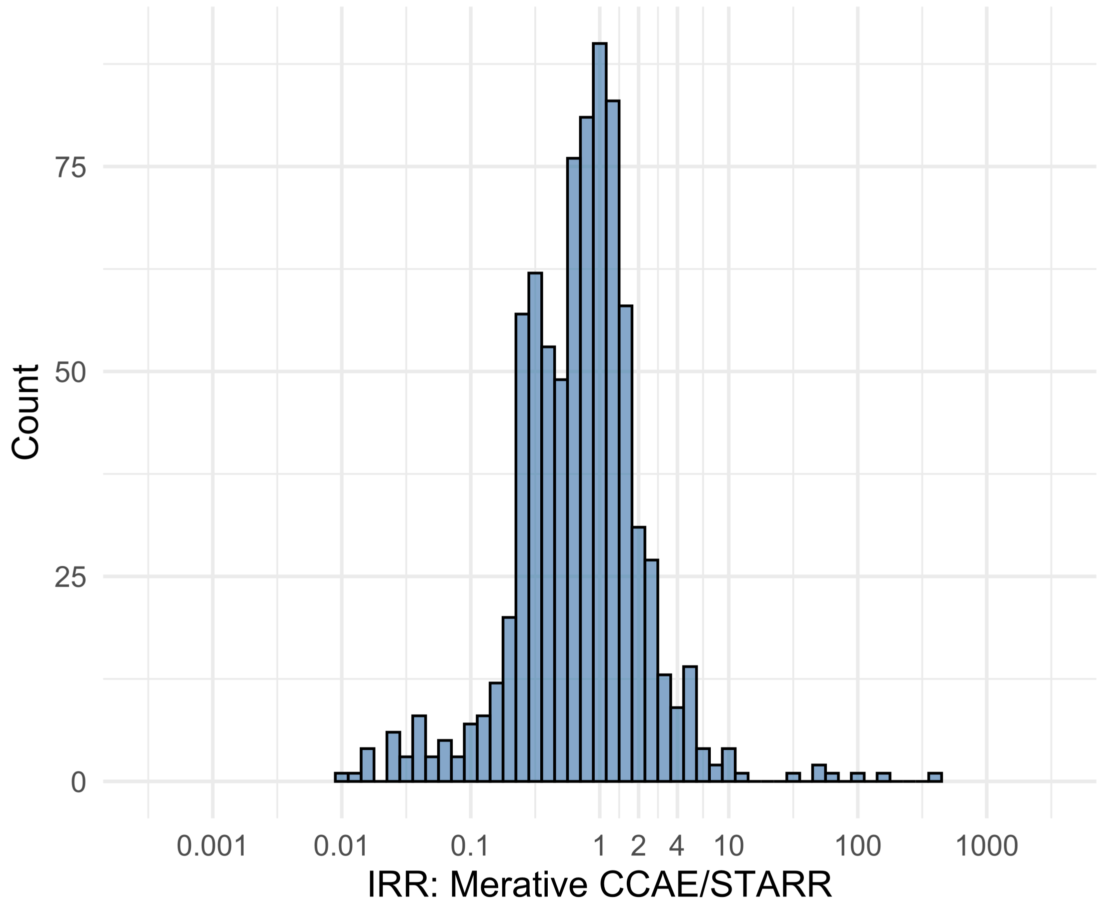


Figure S5.84. IRR distribution comparing Merative CCAE and STARR.

*Table S5.84. Most extreme IRRs for Merative CCAE vs STARR (rounded to 0.001).*

| source1 | source2 | Drug and Indication | Outcome | IRR |
|---|---|---|---|---|
| Merative CCAE | STARR | New users of Beta blockers nested in essential hypertension | Febrile Neutropenia or Neutropenic Fever | 365.2 |
| Merative CCAE | STARR | New users of Beta blockers nested in Left Heart Failure | Febrile Neutropenia or Neutropenic Fever | 167.0 |
| Merative CCAE | STARR | New users of Fluoroquinolone systemic nested in Acute Typical Pneumonia | Febrile Neutropenia or Neutropenic Fever | 90.7 |
| Merative CCAE | STARR | New users of Trimethoprim systemetic nested in Urinary Tract Infection | Polymorphic Ventricular Tachycardia or Torsades de Pointes | 0.0 |
| Merative CCAE | STARR | New users of Beta blockers nested in Acute Myocardial Infarction | Thrombosis with Thrombocytopenia (TWT) | 0.0 |
| Merative CCAE | STARR | New users of Fluoroquinolone systemic nested in Urinary Tract Infection | Polymorphic Ventricular Tachycardia or Torsades de Pointes | 0.0 |

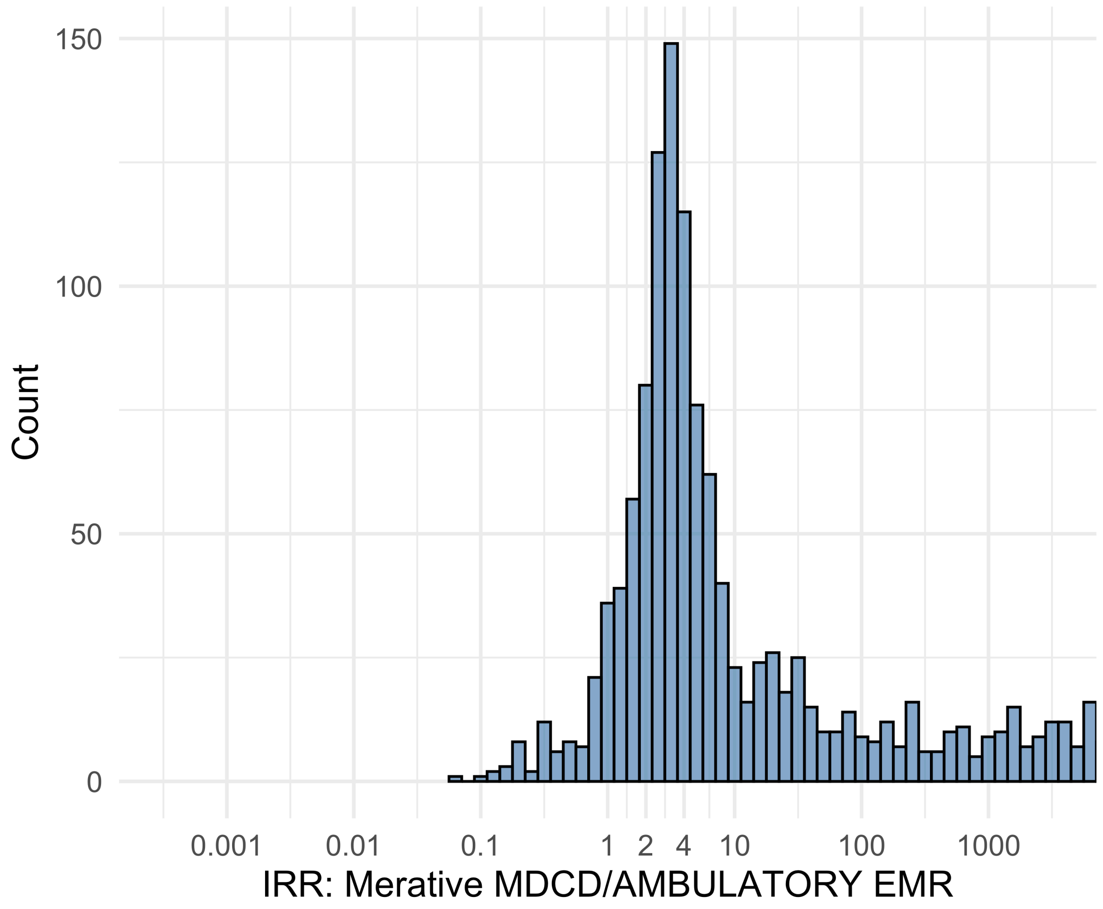


Figure S5.85. IRR distribution comparing Merative MDCD and AMBULATORY EMR.

*Table S5.85. Most extreme IRRs for Merative MDCD vs AMBULATORY EMR (rounded to 0.001).*

| source1 | source2 | Drug and Indication | Outcome | IRR |
|---|---|---|---|---|
| Merative MDCD | AMBULATORY EMR | New users of Beta blockers nested in essential hypertension | Hospitalization with heart failure events | 605067.2 |
| Merative MDCD | AMBULATORY EMR | New users of Beta blockers nested in essential hypertension | 4-point MACE | 383496.7 |
| Merative MDCD | AMBULATORY EMR | New users of Beta blockers nested in essential hypertension | Total cardiovascular disease events (ischemic stroke, hemorrhagic stroke, heart failure, acute myocardial infarction or sudden cardiac death) | 378136.5 |
| Merative MDCD | AMBULATORY EMR | New users of JAK inhibitors nested in Rheumatoid arthritis | Anaphylaxis | 0.1 |
| Merative MDCD | AMBULATORY EMR | New users of Beta blockers nested in Left Heart Failure | Polymorphic Ventricular Tachycardia or Torsades de Pointes | 0.1 |
| Merative MDCD | AMBULATORY EMR | New users of Trimethoprim systemetic nested in Acute Typical Pneumonia | Anaphylaxis | 0.1 |

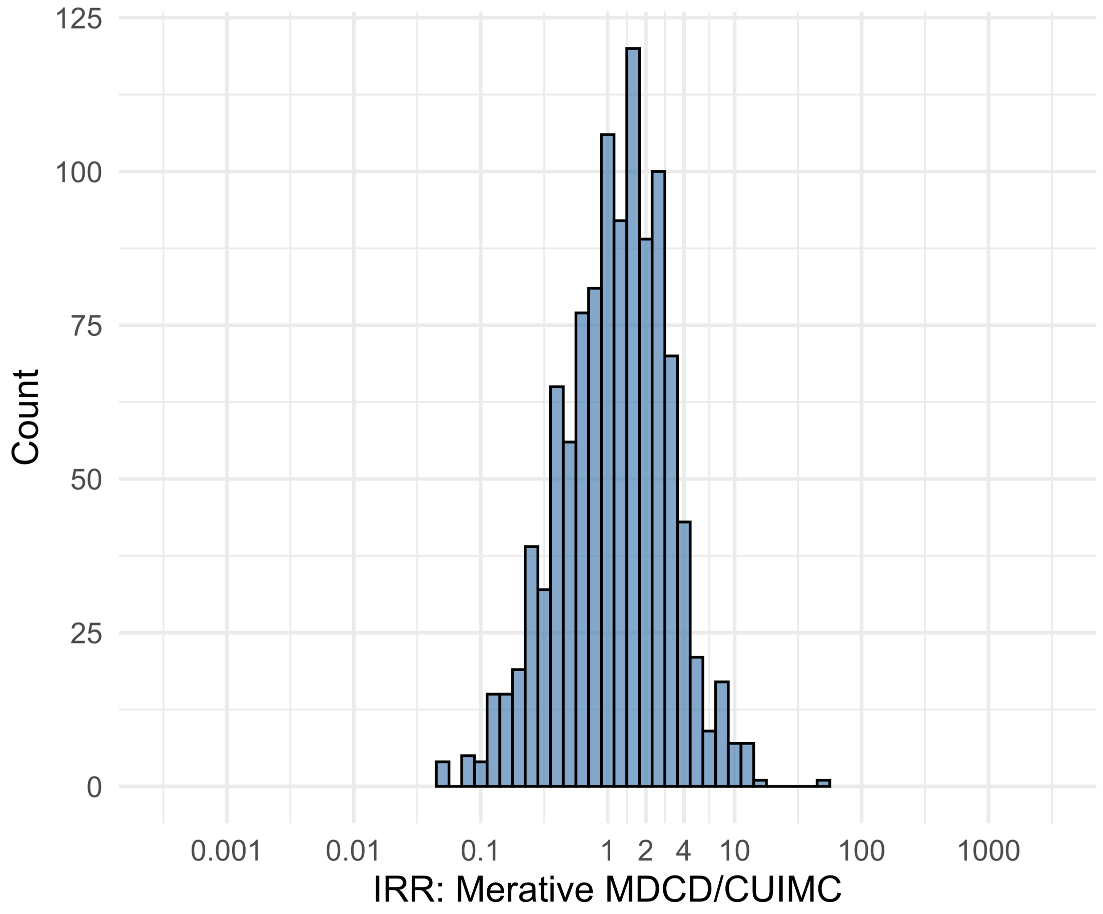


Figure S5.86. IRR distribution comparing Merative MDCD and CUIMC.

*Table S5.86. Most extreme IRRs for Merative MDCD vs CUIMC (rounded to 0.001).*

| source1 | source2 | Drug and Indication | Outcome | IRR |
|---|---|---|---|---|
| Merative MDCD | CUIMC | New users of Beta blockers nested in Acute Myocardial Infarction | Abnormal weight gain events | 45.3 |
| Merative MDCD | CUIMC | New users of Tumor Necrosis Factor alpha (TNFa) inhibitors nested in Rheumatoid arthritis | Acute Myocardial Infarction including its complications | 14.4 |
| Merative MDCD | CUIMC | New users of Tumor Necrosis Factor alpha (TNFa) inhibitors nested in Rheumatoid arthritis | Coronary Vessel Revascularization | 13.9 |
| Merative MDCD | CUIMC | New users of Fluoroquinolone systemic nested in Urinary Tract Infection | Polymorphic Ventricular Tachycardia or Torsades de Pointes | 0.0 |
| Merative MDCD | CUIMC | New users of SGLT2 inhibitor nested in Left Heart Failure | All events of Autoimmune hepatitis, with a washout period of 365 days | 0.1 |
| Merative MDCD | CUIMC | New users of Trimethoprim systemetic nested in Urinary Tract Infection | Progressive multifocal leukoencephalopathy | 0.1 |

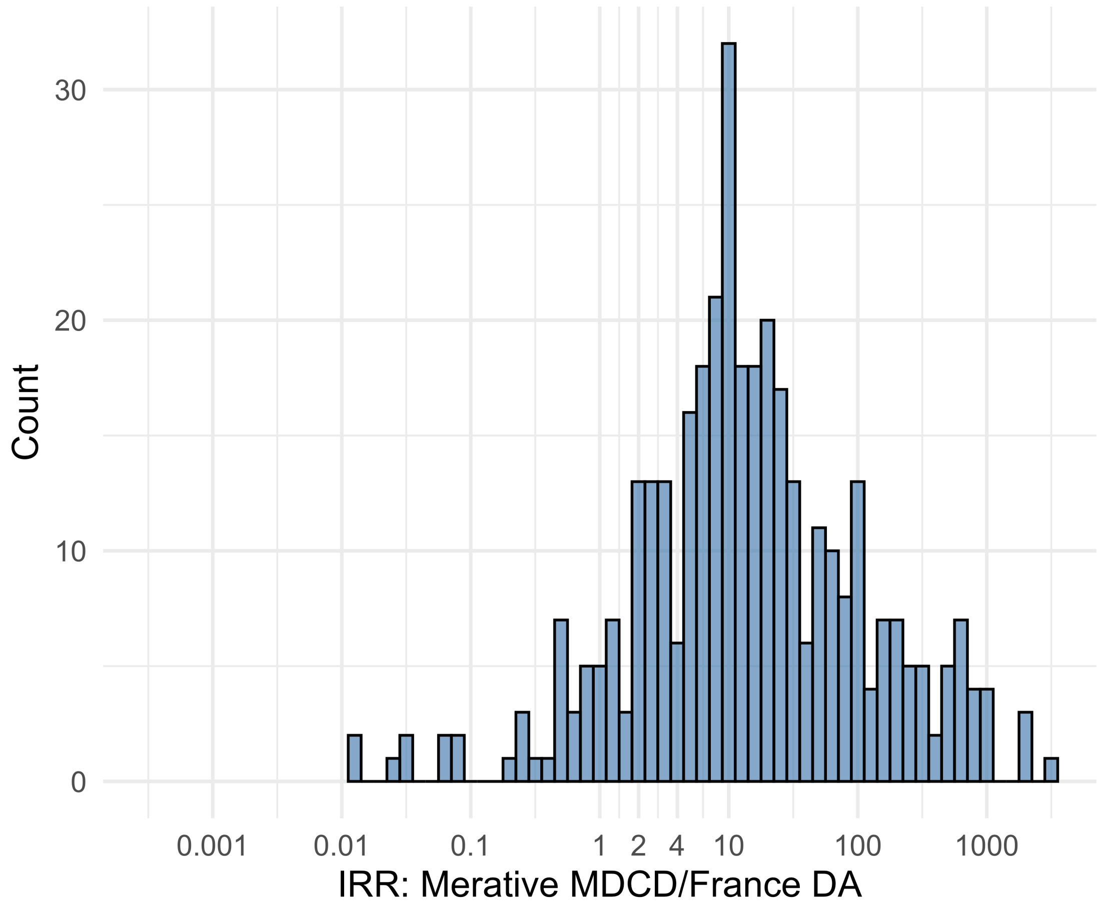


Figure S5.87. IRR distribution comparing Merative MDCD and France DA.

*Table S5.87. Most extreme IRRs for Merative MDCD vs France DA (rounded to 0.001).*

| source1 | source2 | Drug and Indication | Outcome | IRR |
|---|---|---|---|---|
| Merative MDCD | France DA | New users of Beta blockers nested in essential hypertension | Hospitalization with heart failure events | 2972.4 |
| Merative MDCD | France DA | New users of Beta blockers nested in essential hypertension | Acute Kidney Injury AKI | 2016.7 |
| Merative MDCD | France DA | New users of Beta blockers nested in essential hypertension | 4-point MACE | 1883.9 |
| Merative MDCD | France DA | New users of Trimethoprim systemetic nested in Urinary Tract Infection | Polymorphic Ventricular Tachycardia or Torsades de Pointes | 0.0 |
| Merative MDCD | France DA | New users of Trimethoprim systemetic nested in Urinary Tract Infection | Progressive multifocal leukoencephalopathy | 0.0 |
| Merative MDCD | France DA | New users of Fluoroquinolone systemic nested in Urinary Tract Infection | Polymorphic Ventricular Tachycardia or Torsades de Pointes | 0.0 |

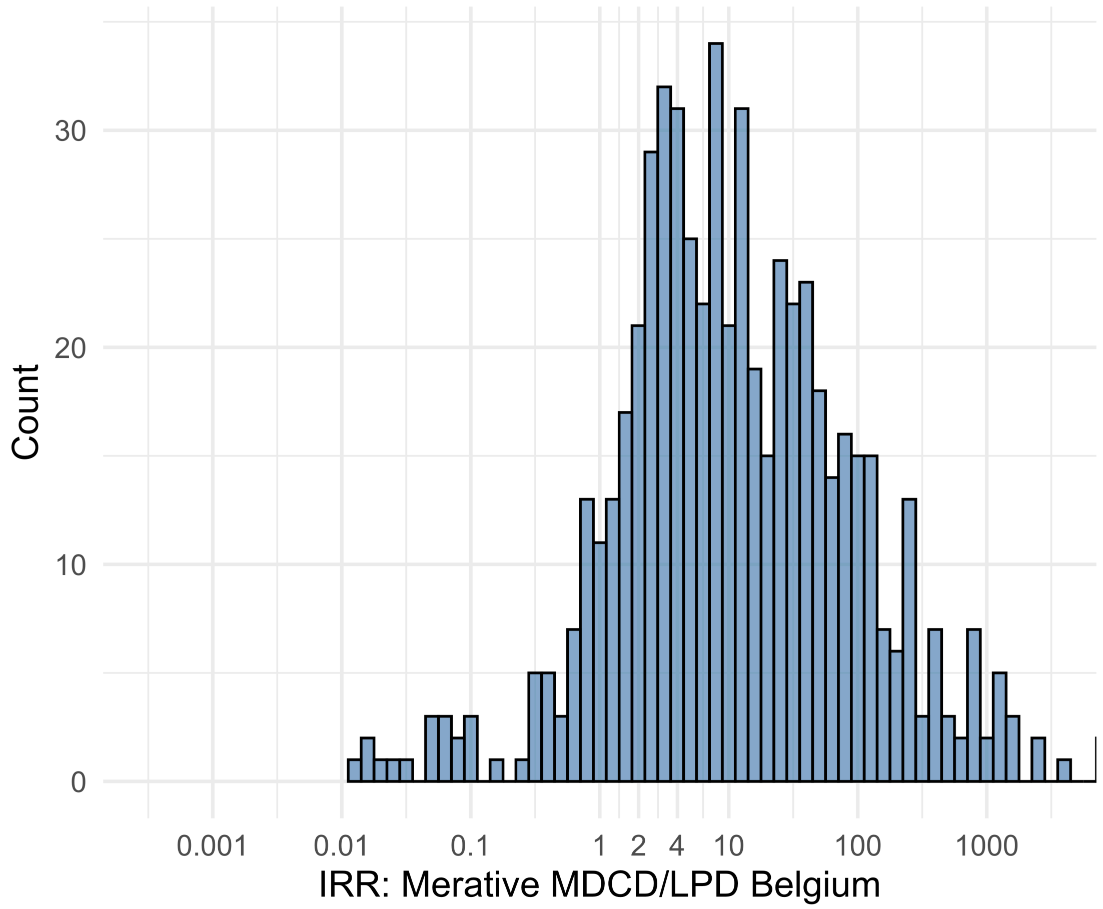


Figure S5.88. IRR distribution comparing Merative MDCD and LPD Belgium.

*Table S5.88. Most extreme IRRs for Merative MDCD vs LPD Belgium (rounded to 0.001).*

| source1 | source2 | Drug and Indication | Outcome | IRR |
|---|---|---|---|---|
| Merative MDCD | LPD Belgium | New users of Beta blockers nested in essential hypertension | Hospitalization with heart failure events | 11567.8 |
| Merative MDCD | LPD Belgium | New users of Beta blockers nested in essential hypertension | 4-point MACE | 7331.7 |
| Merative MDCD | LPD Belgium | New users of Beta blockers nested in essential hypertension | Total cardiovascular disease events (ischemic stroke, hemorrhagic stroke, heart failure, acute myocardial infarction or sudden cardiac death) | 7229.3 |
| Merative MDCD | LPD Belgium | New users of Cephalosporin systemetic nested in Urinary Tract Infection | Polymorphic Ventricular Tachycardia or Torsades de Pointes | 0.0 |
| Merative MDCD | LPD Belgium | New users of Trimethoprim systemetic nested in Urinary Tract Infection | Polymorphic Ventricular Tachycardia or Torsades de Pointes | 0.0 |
| Merative MDCD | LPD Belgium | New users of Trimethoprim systemetic nested in Urinary Tract Infection | Progressive multifocal leukoencephalopathy | 0.0 |

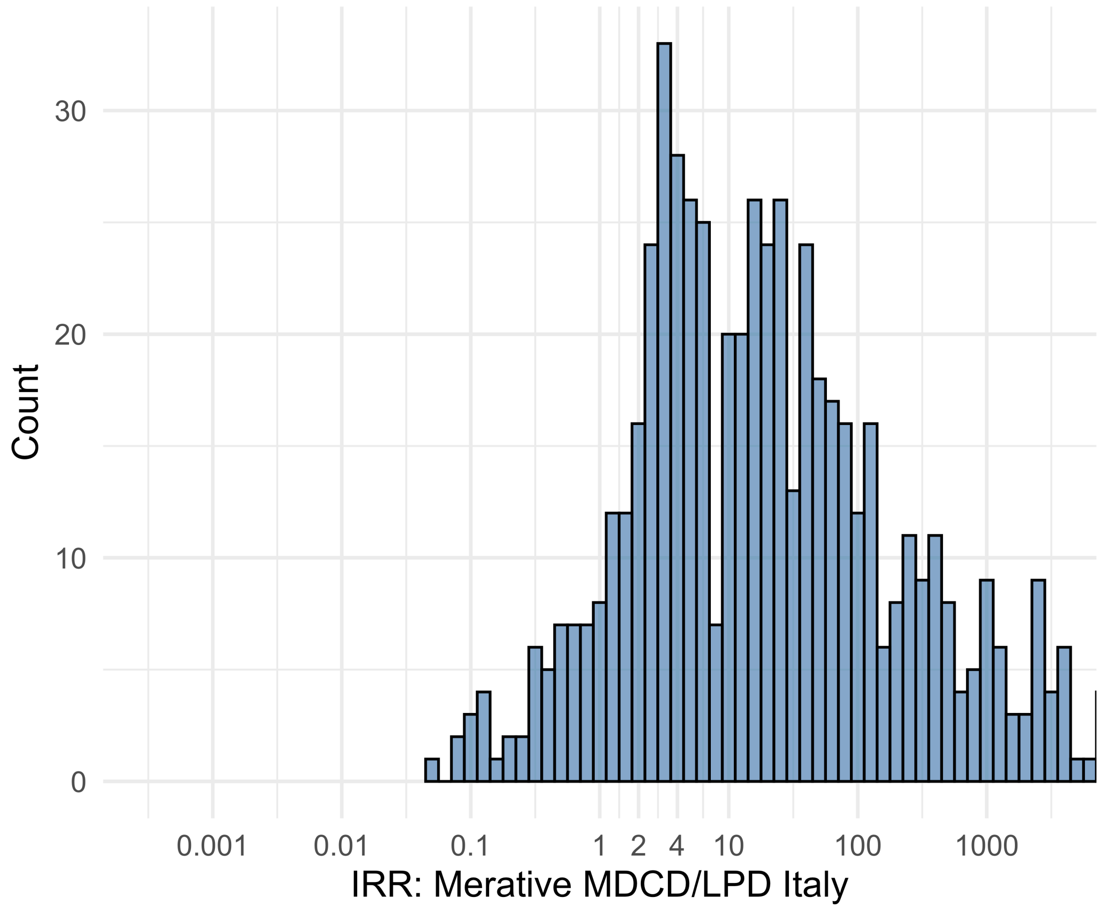


Figure S5.89. IRR distribution comparing Merative MDCD and LPD Italy.

*Table S5.89. Most extreme IRRs for Merative MDCD vs LPD Italy (rounded to 0.001).*

| source1 | source2 | Drug and Indication | Outcome | IRR |
|---|---|---|---|---|
| Merative MDCD | LPD Italy | New users of Beta blockers nested in essential hypertension | Hospitalization with heart failure events | 37610.0 |
| Merative MDCD | LPD Italy | New users of Beta blockers nested in essential hypertension | 4-point MACE | 23837.6 |
| Merative MDCD | LPD Italy | New users of Beta blockers nested in essential hypertension | Total cardiovascular disease events (ischemic stroke, hemorrhagic stroke, heart failure, acute myocardial infarction or sudden cardiac death) | 23504.4 |
| Merative MDCD | LPD Italy | New users of GLP-1 receptor antagonists nested in obesity | Polymorphic Ventricular Tachycardia or Torsades de Pointes | 0.1 |
| Merative MDCD | LPD Italy | New users of Cephalosporin systemetic nested in Acute Typical Pneumonia | Progressive multifocal leukoencephalopathy | 0.1 |
| Merative MDCD | LPD Italy | New users of SGLT2 inhibitor nested in Type 2 diabetes mellitus | Progressive multifocal leukoencephalopathy | 0.1 |

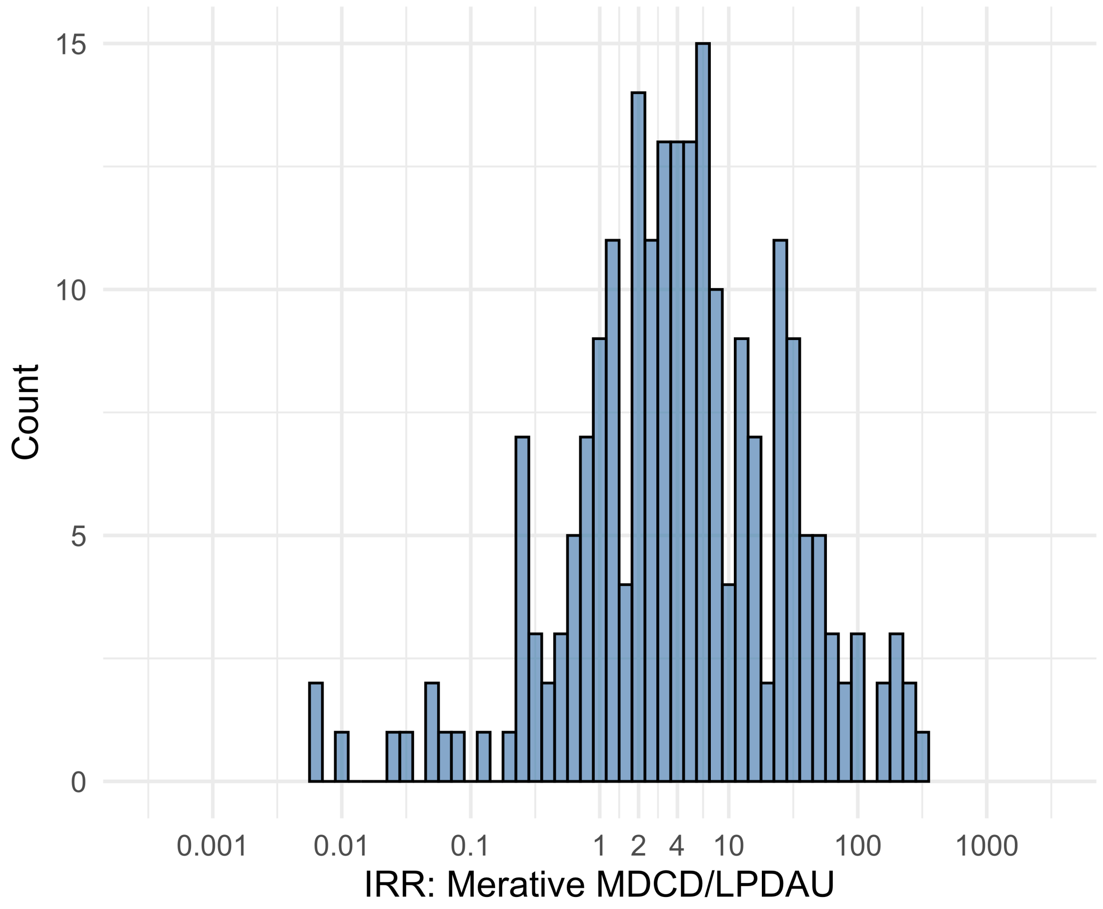


Figure S5.90. IRR distribution comparing Merative MDCD and LPDAU.

*Table S5.90. Most extreme IRRs for Merative MDCD vs LPDAU (rounded to 0.001).*

| source1 | source2 | Drug and Indication | Outcome | IRR |
|---|---|---|---|---|
| Merative MDCD | LPDAU | New users of Beta blockers nested in essential hypertension | Hospitalization with heart failure events | 325.5 |
| Merative MDCD | LPDAU | New users of Cephalosporin systemetic nested in Urinary Tract Infection | Hospitalization with heart failure events | 247.5 |
| Merative MDCD | LPDAU | New users of Cephalosporin systemetic nested in Urinary Tract Infection | Acute Kidney Injury AKI | 231.0 |
| Merative MDCD | LPDAU | New users of Trimethoprim systemetic nested in Urinary Tract Infection | Polymorphic Ventricular Tachycardia or Torsades de Pointes | 0.0 |
| Merative MDCD | LPDAU | New users of Trimethoprim systemetic nested in Urinary Tract Infection | Progressive multifocal leukoencephalopathy | 0.0 |
| Merative MDCD | LPDAU | New users of Cephalosporin systemetic nested in Urinary Tract Infection | Polymorphic Ventricular Tachycardia or Torsades de Pointes | 0.0 |

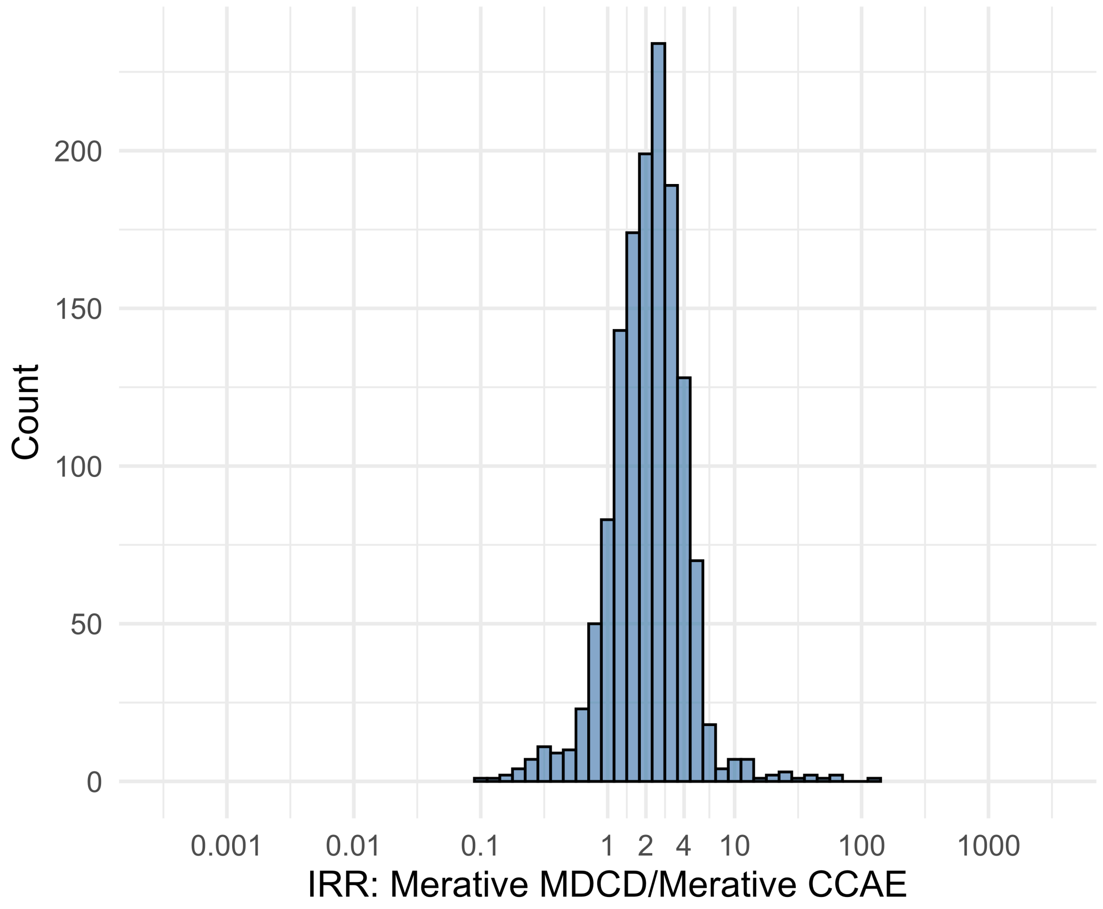


Figure S5.91. IRR distribution comparing Merative MDCD and Merative CCAE.

*Table S5.91. Most extreme IRRs for Merative MDCD vs Merative CCAE (rounded to 0.001).*

| source1 | source2 | Drug and Indication | Outcome | IRR |
|---|---|---|---|---|
| Merative MDCD | Merative CCAE | New users of JAK inhibitors nested in Rheumatoid arthritis | Cardiovascular-related mortality | 128.8 |
| Merative MDCD | Merative CCAE | New users of Tumor Necrosis Factor alpha (TNFa) inhibitors nested in Psoriatic Arthritis | Lower extremity amputation | 65.5 |
| Merative MDCD | Merative CCAE | New users of Tumor Necrosis Factor alpha (TNFa) inhibitors nested in Psoriatic Arthritis | Cardiovascular-related mortality | 65.1 |
| Merative MDCD | Merative CCAE | New users of Tumor Necrosis Factor alpha (TNFa) inhibitors nested in Ulcerative colitis | Persons with gout | 0.1 |
| Merative MDCD | Merative CCAE | New users of Trimethoprim systemetic nested in Acute Typical Pneumonia | All events of Autoimmune hepatitis, with a washout period of 365 days | 0.1 |
| Merative MDCD | Merative CCAE | New users of SGLT2 inhibitor nested in Left Heart Failure | All events of Autoimmune hepatitis, with a washout period of 365 days | 0.2 |

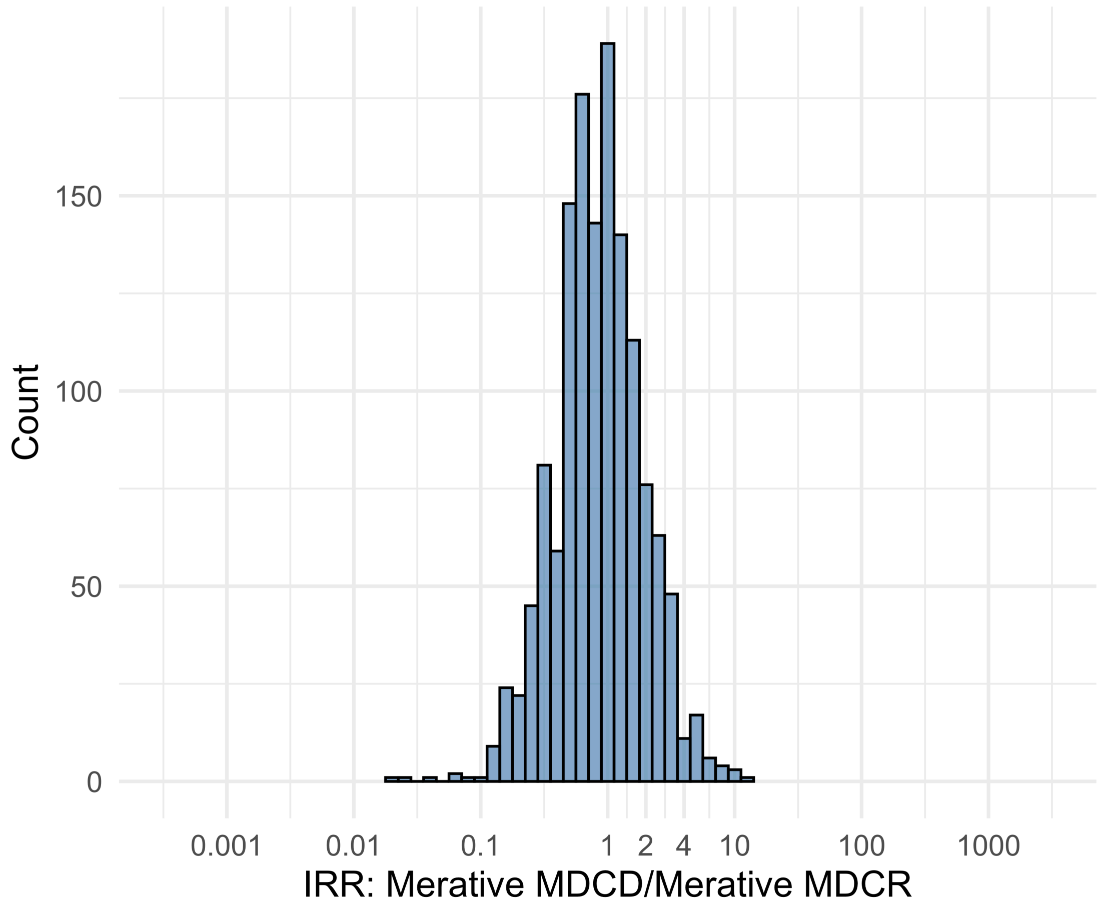


Figure S5.92. IRR distribution comparing Merative MDCD and Merative MDCR.

*Table S5.92. Most extreme IRRs for Merative MDCD vs Merative MDCR (rounded to 0.001).*

| source1 | source2 | Drug and Indication | Outcome | IRR |
|---|---|---|---|---|
| Merative MDCD | Merative MDCR | New users of SGLT2 inhibitor nested in Left Heart Failure | Cardiovascular-related mortality | 13.6 |
| Merative MDCD | Merative MDCR | New users of Tumor Necrosis Factor alpha (TNFa) inhibitors nested in Rheumatoid arthritis | Bells Palsy | 10.0 |
| Merative MDCD | Merative MDCR | New users of Tumor Necrosis Factor alpha (TNFa) inhibitors nested in Rheumatoid arthritis | Narcolepsy | 9.0 |
| Merative MDCD | Merative MDCR | New users of Tumor Necrosis Factor alpha (TNFa) inhibitors nested in Ulcerative colitis | Persons with gout | 0.0 |
| Merative MDCD | Merative MDCR | New users of Tumor Necrosis Factor alpha (TNFa) inhibitors nested in Ulcerative colitis | Coronary Vessel Revascularization | 0.0 |
| Merative MDCD | Merative MDCR | New users of Tumor Necrosis Factor alpha (TNFa) inhibitors nested in Rheumatoid arthritis | Bladder Cancer | 0.0 |

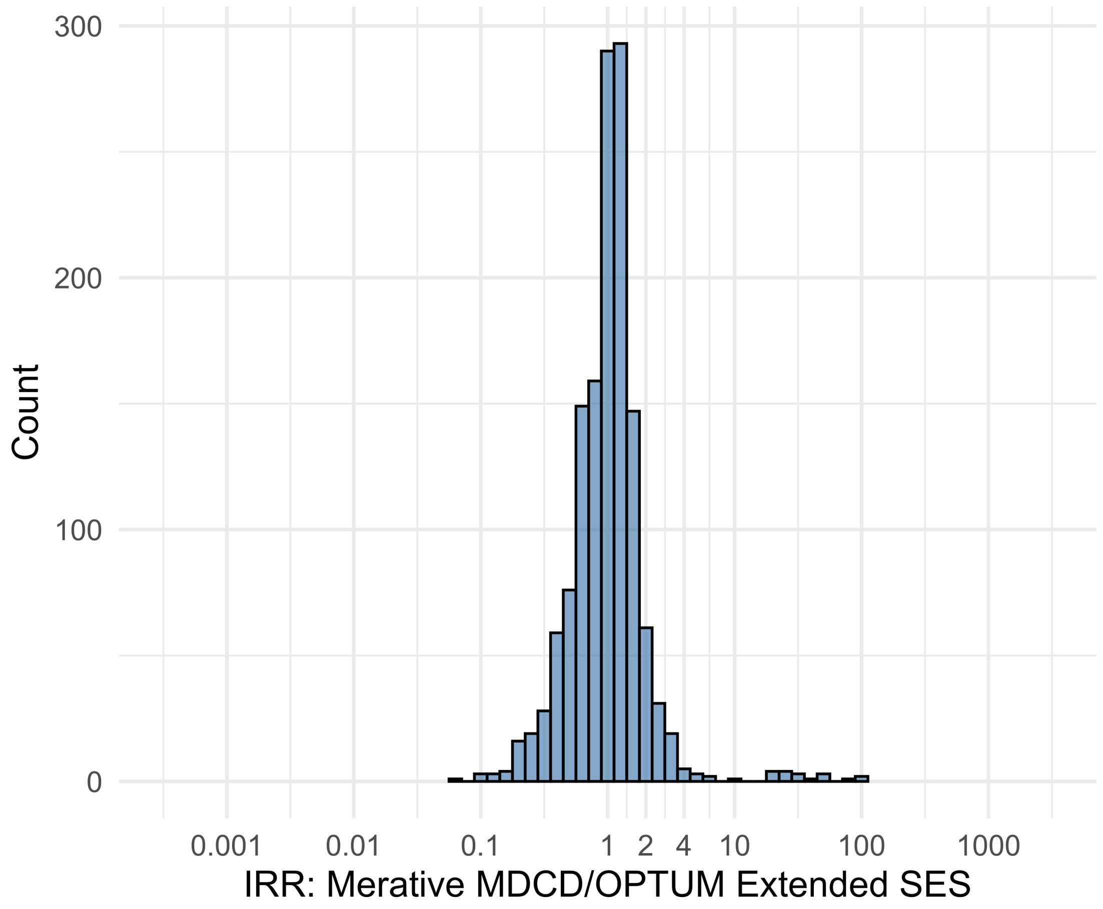


Figure S5.93. IRR distribution comparing Merative MDCD and OPTUM Extended SES.

*Table S5.93. Most extreme IRRs for Merative MDCD vs OPTUM Extended SES (rounded to 0.001).*

| source1 | source2 | Drug and Indication | Outcome | IRR |
|---|---|---|---|---|
| Merative MDCD | OPTUM Extended SES | New users of JAK inhibitors nested in Rheumatoid arthritis | Cardiovascular-related mortality | 95.5 |
| Merative MDCD | OPTUM Extended SES | New users of Tumor Necrosis Factor alpha (TNFa) inhibitors nested in Rheumatoid arthritis | Cardiovascular-related mortality | 89.8 |
| Merative MDCD | OPTUM Extended SES | New users of Tumor Necrosis Factor alpha (TNFa) inhibitors nested in Psoriatic Arthritis | Cardiovascular-related mortality | 74.4 |
| Merative MDCD | OPTUM Extended SES | New users of Tumor Necrosis Factor alpha (TNFa) inhibitors nested in Ulcerative colitis | Persons with gout | 0.1 |
| Merative MDCD | OPTUM Extended SES | New users of Trimethoprim systemetic nested in Acute Typical Pneumonia | All events of Autoimmune hepatitis, with a washout period of 365 days | 0.1 |
| Merative MDCD | OPTUM Extended SES | New users of Tumor Necrosis Factor alpha (TNFa) inhibitors nested in Rheumatoid arthritis | Autoimmune hemolytic anemia | 0.1 |

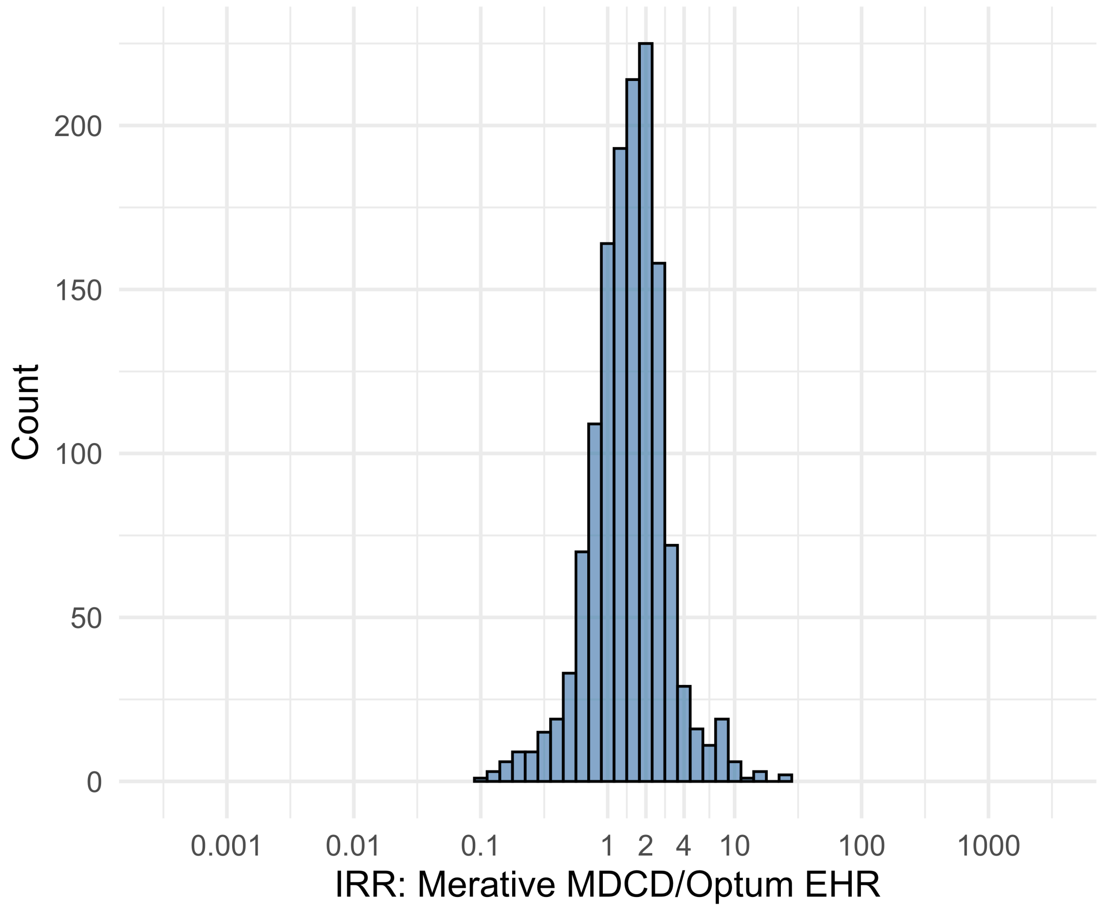


Figure S5.94. IRR distribution comparing Merative MDCD and Optum EHR.

*Table S5.94. Most extreme IRRs for Merative MDCD vs Optum EHR (rounded to 0.001).*

| source1 | source2 | Drug and Indication | Outcome | IRR |
|---|---|---|---|---|
| Merative MDCD | Optum EHR | New users of JAK inhibitors nested in Rheumatoid arthritis | Guillain Barre Syndrome | 23.6 |
| Merative MDCD | Optum EHR | New users of JAK inhibitors nested in Rheumatoid arthritis | Stevens-Johnson syndrome, toxic epidermal necrolysis spectrum | 23.6 |
| Merative MDCD | Optum EHR | New users of Tumor Necrosis Factor alpha (TNFa) inhibitors nested in Rheumatoid arthritis | All events Drug Rash with Eosinophilia and Systemic Symptoms (DRESS) | 16.2 |
| Merative MDCD | Optum EHR | New users of Tumor Necrosis Factor alpha (TNFa) inhibitors nested in Ulcerative colitis | Persons with gout | 0.1 |
| Merative MDCD | Optum EHR | New users of Trimethoprim systemetic nested in Urinary Tract Infection | Thrombosis with Thrombocytopenia (TWT) | 0.1 |
| Merative MDCD | Optum EHR | New users of Trimethoprim systemetic nested in Acute Typical Pneumonia | All events of Autoimmune hepatitis, with a washout period of 365 days | 0.1 |

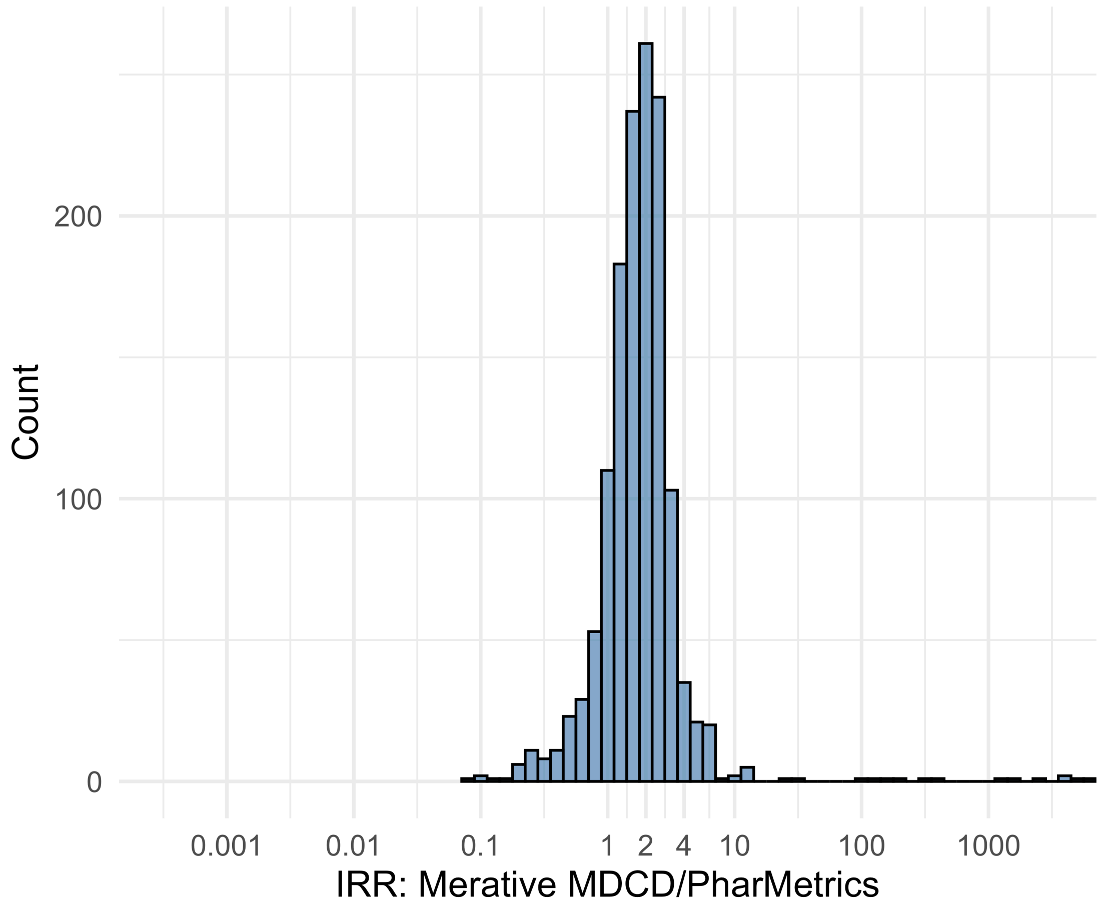


Figure S5.95. IRR distribution comparing Merative MDCD and PharMetrics.

*Table S5.95. Most extreme IRRs for Merative MDCD vs PharMetrics (rounded to 0.001).*

| source1 | source2 | Drug and Indication | Outcome | IRR |
|---|---|---|---|---|
| Merative MDCD | PharMetrics | New users of Beta blockers nested in essential hypertension | Cardiovascular-related mortality | 35942.3 |
| Merative MDCD | PharMetrics | New users of Fluoroquinolone systemic nested in Acute Typical Pneumonia | Cardiovascular-related mortality | 13572.7 |
| Merative MDCD | PharMetrics | New users of Cephalosporin systemetic nested in Urinary Tract Infection | Cardiovascular-related mortality | 13213.9 |
| Merative MDCD | PharMetrics | New users of Tumor Necrosis Factor alpha (TNFa) inhibitors nested in Ulcerative colitis | Persons with gout | 0.1 |
| Merative MDCD | PharMetrics | New users of SGLT2 inhibitor nested in Left Heart Failure | All events of Autoimmune hepatitis, with a washout period of 365 days | 0.1 |
| Merative MDCD | PharMetrics | New users of Trimethoprim systemetic nested in Acute Typical Pneumonia | All events of Autoimmune hepatitis, with a washout period of 365 days | 0.1 |

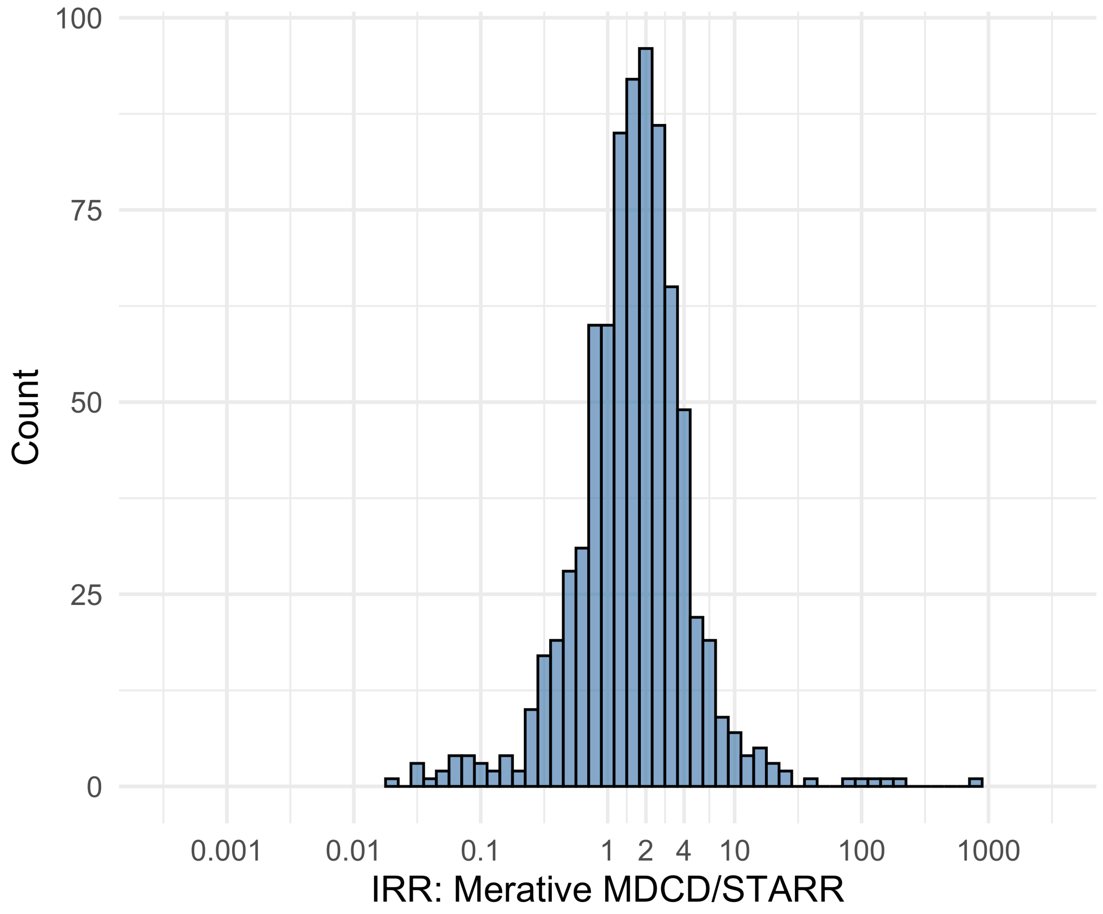


Figure S5.96. IRR distribution comparing Merative MDCD and STARR.

*Table S5.96. Most extreme IRRs for Merative MDCD vs STARR (rounded to 0.001).*

| source1 | source2 | Drug and Indication | Outcome | IRR |
|---|---|---|---|---|
| Merative MDCD | STARR | New users of Beta blockers nested in essential hypertension | Febrile Neutropenia or Neutropenic Fever | 727.7 |
| Merative MDCD | STARR | New users of Beta blockers nested in Left Heart Failure | Febrile Neutropenia or Neutropenic Fever | 205.4 |
| Merative MDCD | STARR | New users of Fluoroquinolone systemic nested in Acute Typical Pneumonia | Febrile Neutropenia or Neutropenic Fever | 156.9 |
| Merative MDCD | STARR | New users of Trimethoprim systemetic nested in Urinary Tract Infection | Progressive multifocal leukoencephalopathy | 0.0 |
| Merative MDCD | STARR | New users of Cephalosporin systemetic nested in Acute Typical Pneumonia | Progressive multifocal leukoencephalopathy | 0.0 |
| Merative MDCD | STARR | New users of SGLT2 inhibitor nested in Type 2 diabetes mellitus | Progressive multifocal leukoencephalopathy | 0.0 |

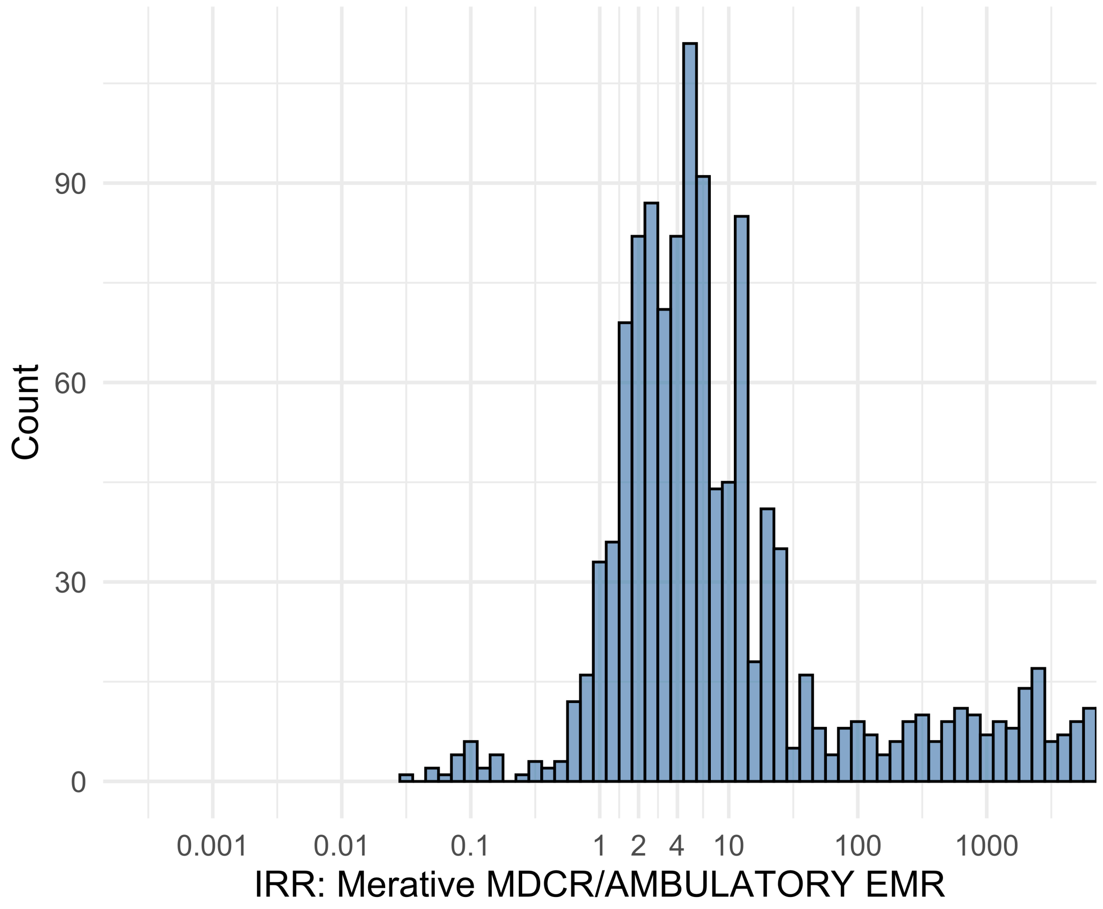


Figure S5.97. IRR distribution comparing Merative MDCR and AMBULATORY EMR.

*Table S5.97. Most extreme IRRs for Merative MDCR vs AMBULATORY EMR (rounded to 0.001).*

| source1 | source2 | Drug and Indication | Outcome | IRR |
|---|---|---|---|---|
| Merative MDCR | AMBULATORY EMR | New users of Beta blockers nested in essential hypertension | Hospitalization with heart failure events | 581913.0 |
| Merative MDCR | AMBULATORY EMR | New users of Beta blockers nested in essential hypertension | 4-point MACE | 468389.9 |
| Merative MDCR | AMBULATORY EMR | New users of Beta blockers nested in essential hypertension | Total cardiovascular disease events (ischemic stroke, hemorrhagic stroke, heart failure, acute myocardial infarction or sudden cardiac death) | 464793.0 |
| Merative MDCR | AMBULATORY EMR | New users of Tumor Necrosis Factor alpha (TNFa) inhibitors nested in Rheumatoid arthritis | Anaphylaxis | 0.0 |
| Merative MDCR | AMBULATORY EMR | New users of Beta blockers nested in Left Heart Failure | Polymorphic Ventricular Tachycardia or Torsades de Pointes | 0.0 |
| Merative MDCR | AMBULATORY EMR | New users of Trimethoprim systemetic nested in Acute Typical Pneumonia | Anaphylaxis | 0.1 |

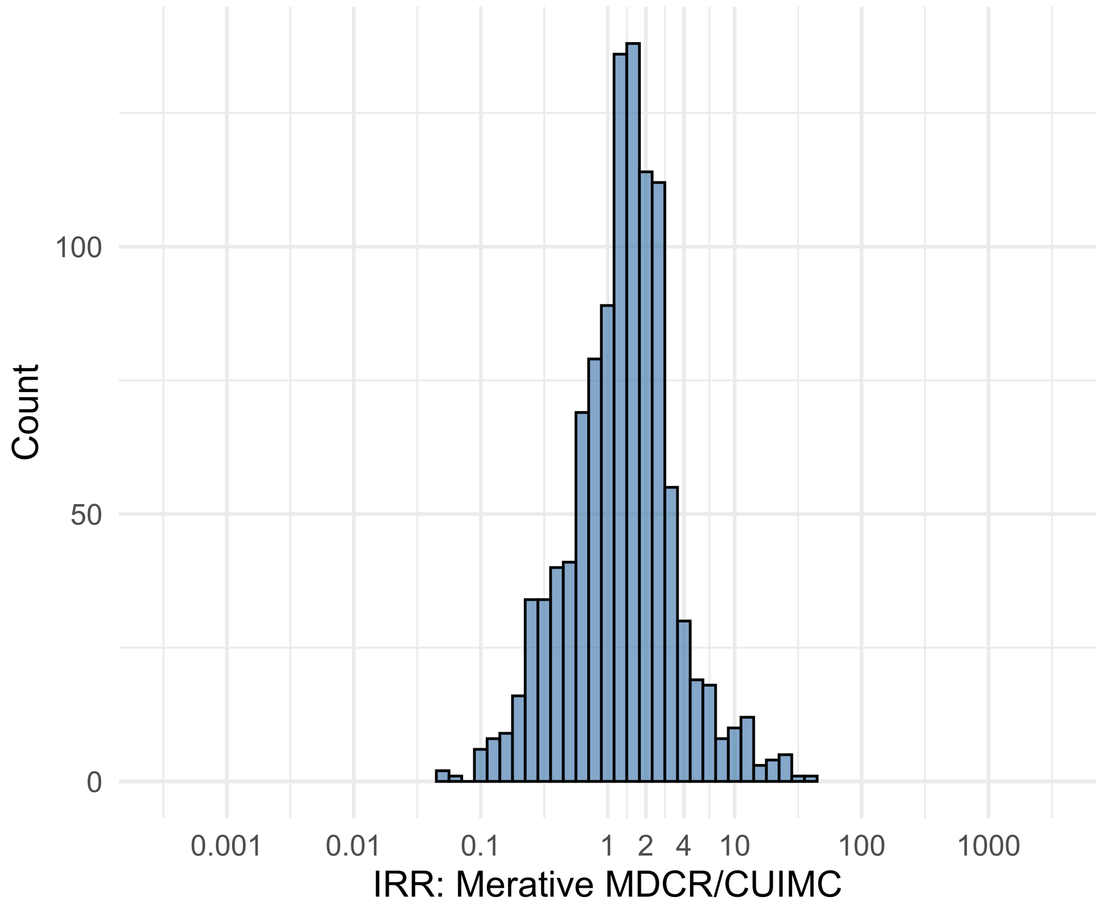


Figure S5.98. IRR distribution comparing Merative MDCR and CUIMC.

*Table S5.98. Most extreme IRRs for Merative MDCR vs CUIMC (rounded to 0.001).*

| source1 | source2 | Drug and Indication | Outcome | IRR |
|---|---|---|---|---|
| Merative MDCR | CUIMC | New users of Tumor Necrosis Factor alpha (TNFa) inhibitors nested in Crohns disease | Stroke (ischemic or hemorrhagic) events | 39.1 |
| Merative MDCR | CUIMC | New users of Tumor Necrosis Factor alpha (TNFa) inhibitors nested in Rheumatoid arthritis | Coronary Vessel Revascularization | 31.0 |
| Merative MDCR | CUIMC | New users of Tumor Necrosis Factor alpha (TNFa) inhibitors nested in Rheumatoid arthritis | Acute Myocardial Infarction including its complications | 27.9 |
| Merative MDCR | CUIMC | New users of Tumor Necrosis Factor alpha (TNFa) inhibitors nested in Rheumatoid arthritis | Myocarditis Pericarditis | 0.0 |
| Merative MDCR | CUIMC | New users of Trimethoprim systemetic nested in Acute Typical Pneumonia | Earliest event of Thrombotic microangiopathy (TMA) or Microangiopathic hemolytic anemia (MAHA) | 0.1 |
| Merative MDCR | CUIMC | New users of Beta blockers nested in essential hypertension | Progressive multifocal leukoencephalopathy | 0.1 |

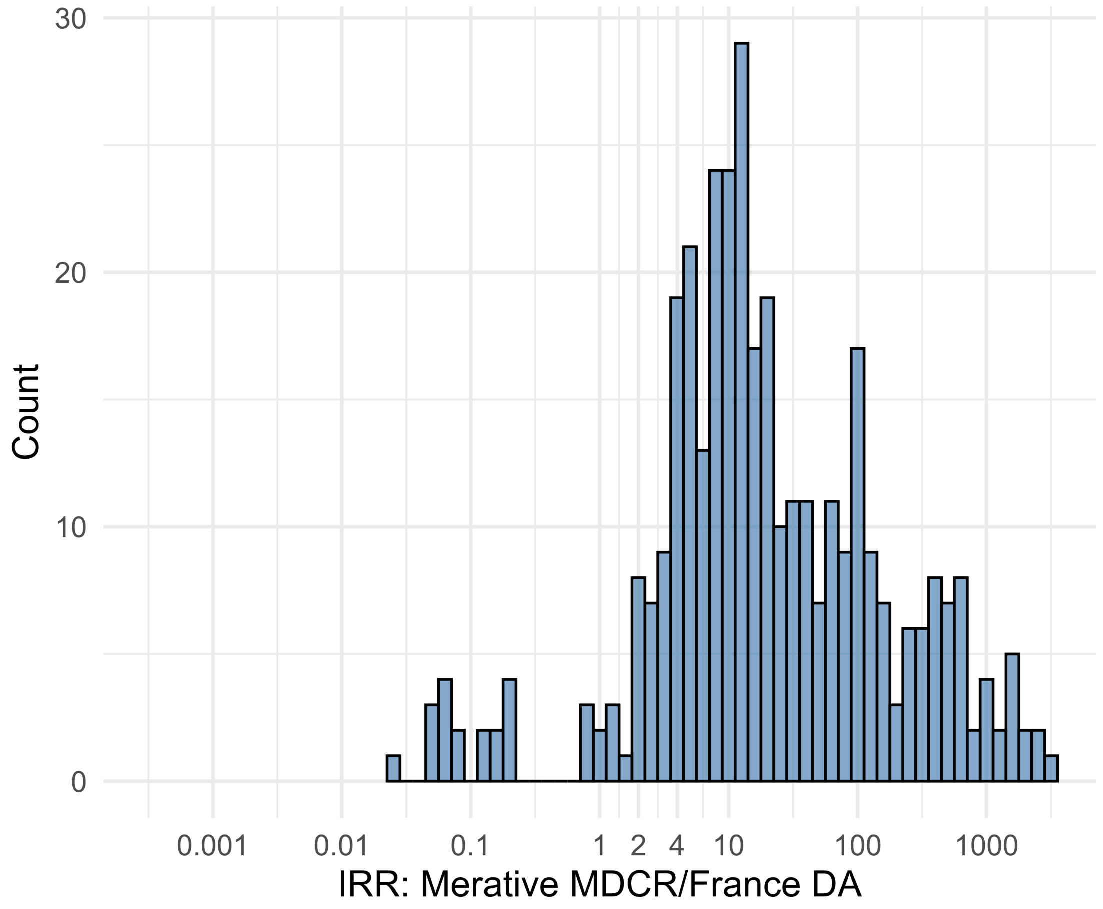


Figure S5.99. IRR distribution comparing Merative MDCR and France DA.

*Table S5.99. Most extreme IRRs for Merative MDCR vs France DA (rounded to 0.001).*

| source1 | source2 | Drug and Indication | Outcome | IRR |
|---|---|---|---|---|
| Merative MDCR | France DA | New users of Beta blockers nested in essential hypertension | Hospitalization with heart failure events | 2858.6 |
| Merative MDCR | France DA | New users of Beta blockers nested in essential hypertension | 4-point MACE | 2300.9 |
| Merative MDCR | France DA | New users of Beta blockers nested in essential hypertension | Total cardiovascular disease events (ischemic stroke, hemorrhagic stroke, heart failure, acute myocardial infarction or sudden cardiac death) | 2283.3 |
| Merative MDCR | France DA | New users of Trimethoprim systemetic nested in Urinary Tract Infection | Polymorphic Ventricular Tachycardia or Torsades de Pointes | 0.0 |
| Merative MDCR | France DA | New users of Trimethoprim systemetic nested in Urinary Tract Infection | Progressive multifocal leukoencephalopathy | 0.1 |
| Merative MDCR | France DA | New users of Trimethoprim systemetic nested in Urinary Tract Infection | All events Drug Rash with Eosinophilia and Systemic Symptoms (DRESS) | 0.1 |

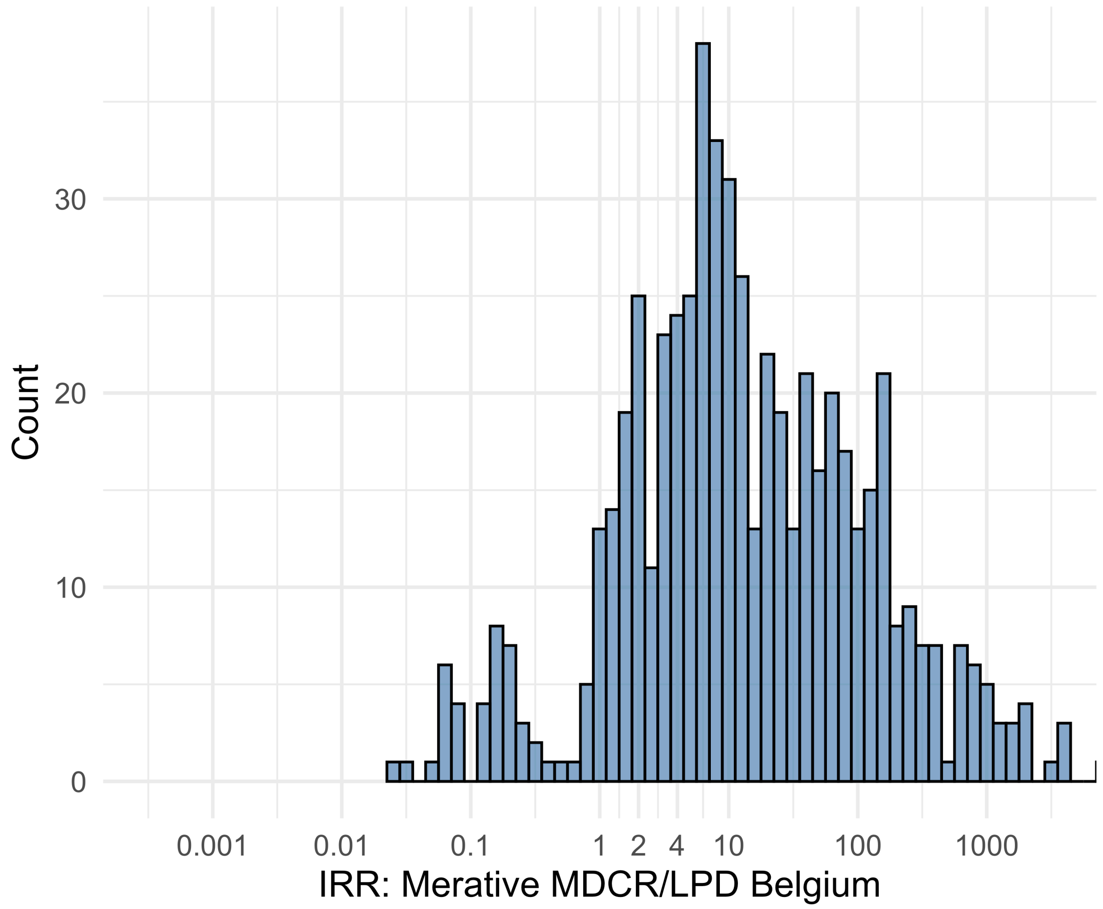


Figure S5.100. IRR distribution comparing Merative MDCR and LPD Belgium.

*Table S5.100. Most extreme IRRs for Merative MDCR vs LPD Belgium (rounded to 0.001).*

| source1 | source2 | Drug and Indication | Outcome | IRR |
|---|---|---|---|---|
| Merative MDCR | LPD Belgium | New users of Beta blockers nested in essential hypertension | Hospitalization with heart failure events | 11125.1 |
| Merative MDCR | LPD Belgium | New users of Beta blockers nested in essential hypertension | 4-point MACE | 8954.7 |
| Merative MDCR | LPD Belgium | New users of Beta blockers nested in essential hypertension | Total cardiovascular disease events (ischemic stroke, hemorrhagic stroke, heart failure, acute myocardial infarction or sudden cardiac death) | 8886.0 |
| Merative MDCR | LPD Belgium | New users of Cephalosporin systemetic nested in Urinary Tract Infection | Polymorphic Ventricular Tachycardia or Torsades de Pointes | 0.0 |
| Merative MDCR | LPD Belgium | New users of Trimethoprim systemetic nested in Urinary Tract Infection | Polymorphic Ventricular Tachycardia or Torsades de Pointes | 0.0 |
| Merative MDCR | LPD Belgium | New users of Fluoroquinolone systemic nested in Acute Typical Pneumonia | Polymorphic Ventricular Tachycardia or Torsades de Pointes | 0.1 |

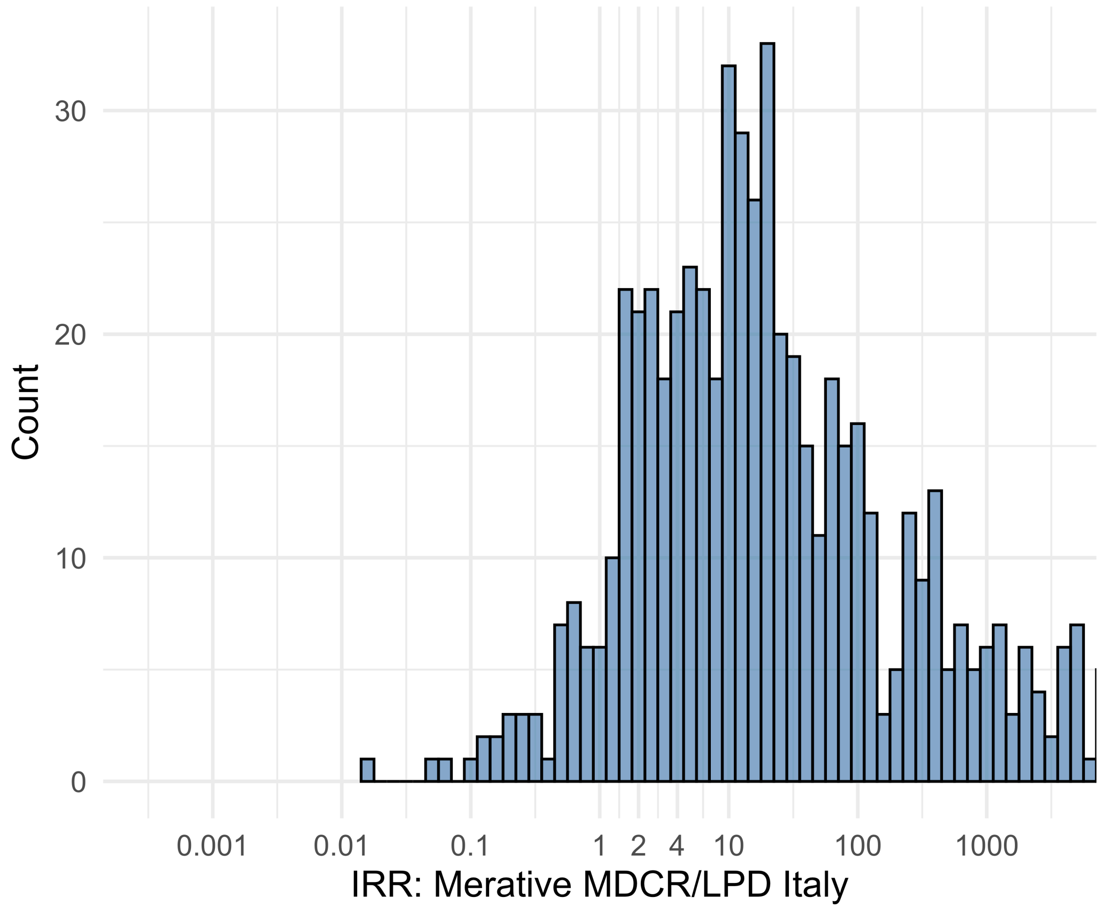


Figure S5.101. IRR distribution comparing Merative MDCR and LPD Italy.

*Table S5.101. Most extreme IRRs for Merative MDCR vs LPD Italy (rounded to 0.001).*

| source1 | source2 | Drug and Indication | Outcome | IRR |
|---|---|---|---|---|
| Merative MDCR | LPD Italy | New users of Beta blockers nested in essential hypertension | Hospitalization with heart failure events | 36170.8 |
| Merative MDCR | LPD Italy | New users of Beta blockers nested in essential hypertension | 4-point MACE | 29114.4 |
| Merative MDCR | LPD Italy | New users of Beta blockers nested in essential hypertension | Total cardiovascular disease events (ischemic stroke, hemorrhagic stroke, heart failure, acute myocardial infarction or sudden cardiac death) | 28890.8 |
| Merative MDCR | LPD Italy | New users of Beta blockers nested in essential hypertension | Progressive multifocal leukoencephalopathy | 0.0 |
| Merative MDCR | LPD Italy | New users of Cephalosporin systemetic nested in Urinary Tract Infection | Progressive multifocal leukoencephalopathy | 0.1 |
| Merative MDCR | LPD Italy | New users of Fluoroquinolone systemic nested in Urinary Tract Infection | Progressive multifocal leukoencephalopathy | 0.1 |

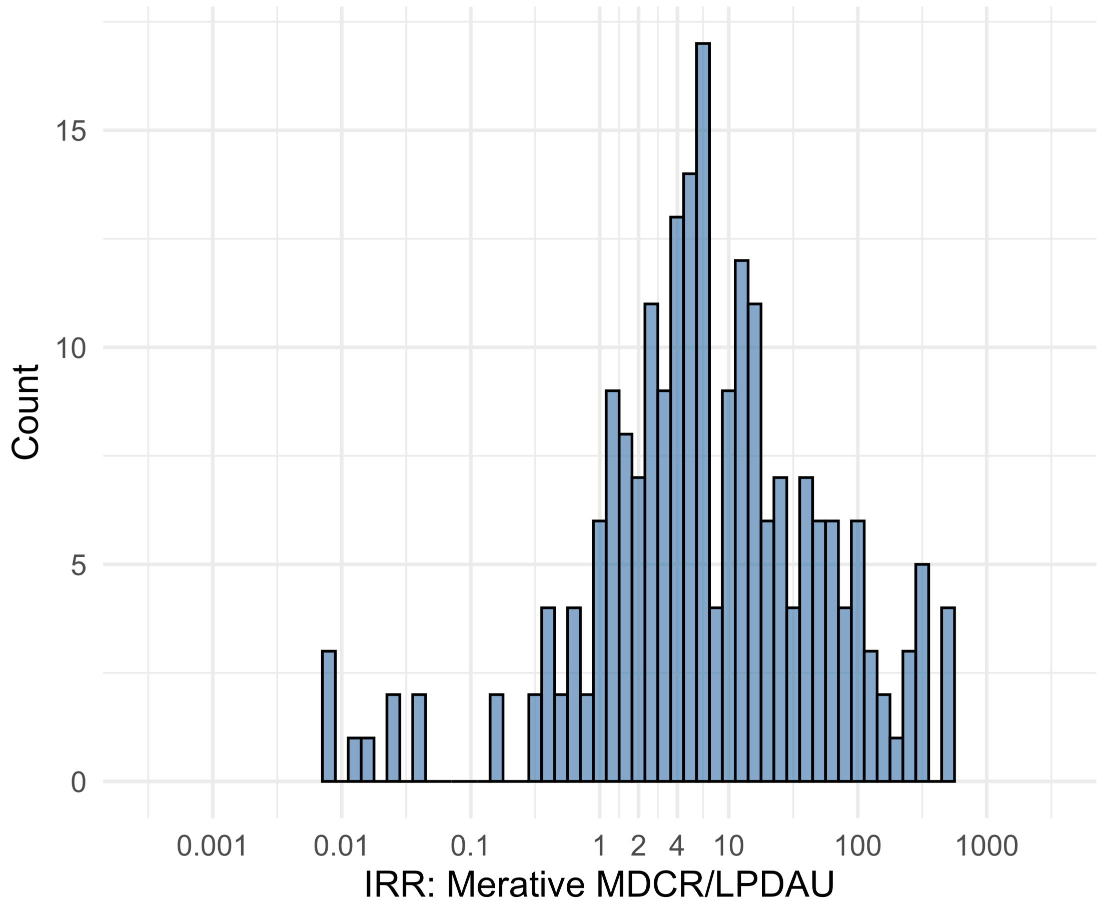


Figure S5.102. IRR distribution comparing Merative MDCR and LPDAU.

*Table S5.102. Most extreme IRRs for Merative MDCR vs LPDAU (rounded to 0.001).*

| source1 | source2 | Drug and Indication | Outcome | IRR |
|---|---|---|---|---|
| Merative MDCR | LPDAU | New users of Cephalosporin systemetic nested in Urinary Tract Infection | Hospitalization with heart failure events | 516.3 |
| Merative MDCR | LPDAU | New users of Cephalosporin systemetic nested in Urinary Tract Infection | Acute Kidney Injury AKI | 468.3 |
| Merative MDCR | LPDAU | New users of Cephalosporin systemetic nested in Urinary Tract Infection | 4-point MACE | 464.1 |
| Merative MDCR | LPDAU | New users of Beta blockers nested in essential hypertension | Polymorphic Ventricular Tachycardia or Torsades de Pointes | 0.0 |
| Merative MDCR | LPDAU | New users of Beta blockers nested in essential hypertension | All events Drug Rash with Eosinophilia and Systemic Symptoms (DRESS) | 0.0 |
| Merative MDCR | LPDAU | New users of Beta blockers nested in essential hypertension | Progressive multifocal leukoencephalopathy | 0.0 |

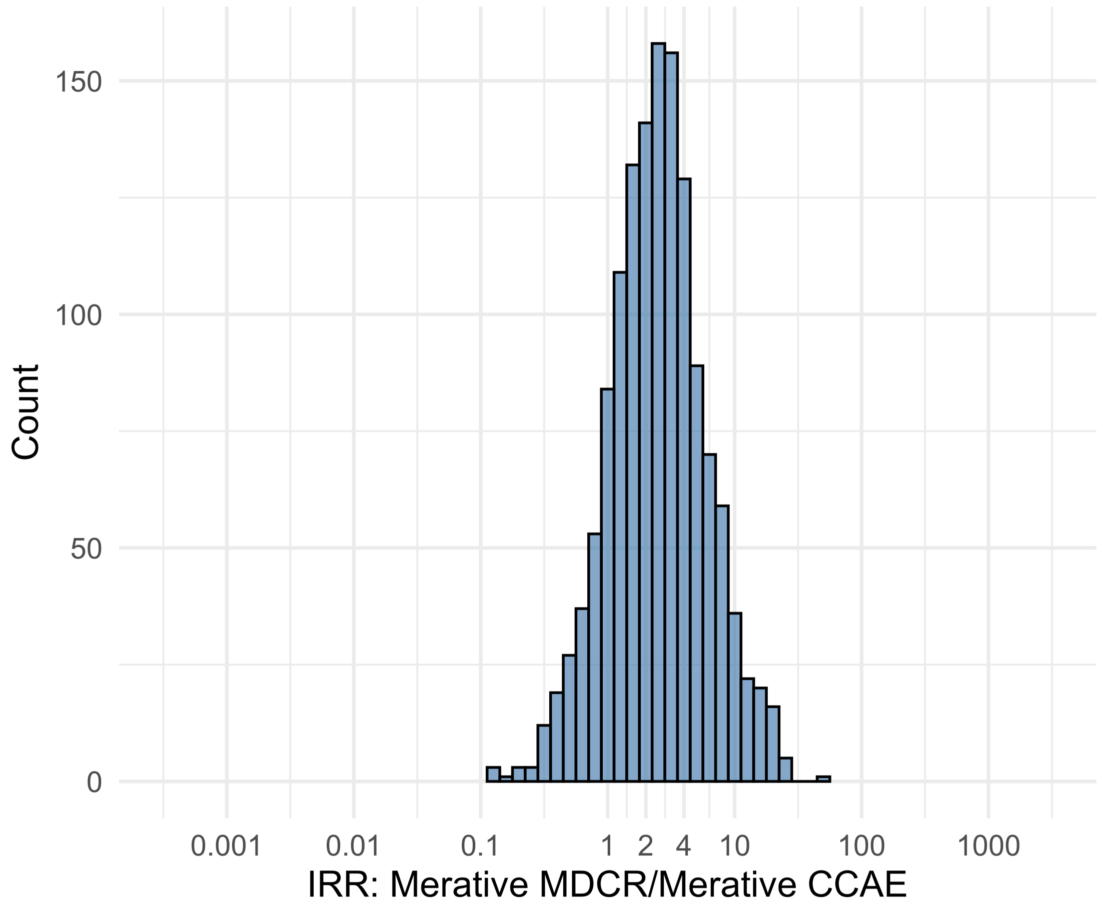


Figure S5.103. IRR distribution comparing Merative MDCR and Merative CCAE.

*Table S5.103. Most extreme IRRs for Merative MDCR vs Merative CCAE (rounded to 0.001).*

| source1 | source2 | Drug and Indication | Outcome | IRR |
|---|---|---|---|---|
| Merative MDCR | Merative CCAE | New users of JAK inhibitors nested in Rheumatoid arthritis | Cardiovascular-related mortality | 48.2 |
| Merative MDCR | Merative CCAE | New users of Trimethoprim systemetic nested in Urinary Tract Infection | Hospitalization with heart failure events | 26.1 |
| Merative MDCR | Merative CCAE | New users of Trimethoprim systemetic nested in Urinary Tract Infection | Cardiovascular-related mortality | 23.9 |
| Merative MDCR | Merative CCAE | New users of Tumor Necrosis Factor alpha (TNFa) inhibitors nested in Rheumatoid arthritis | Anaphylaxis | 0.1 |
| Merative MDCR | Merative CCAE | New users of Tumor Necrosis Factor alpha (TNFa) inhibitors nested in Rheumatoid arthritis | Bells Palsy | 0.1 |
| Merative MDCR | Merative CCAE | New users of Tumor Necrosis Factor alpha (TNFa) inhibitors nested in Rheumatoid arthritis | Myocarditis Pericarditis | 0.1 |

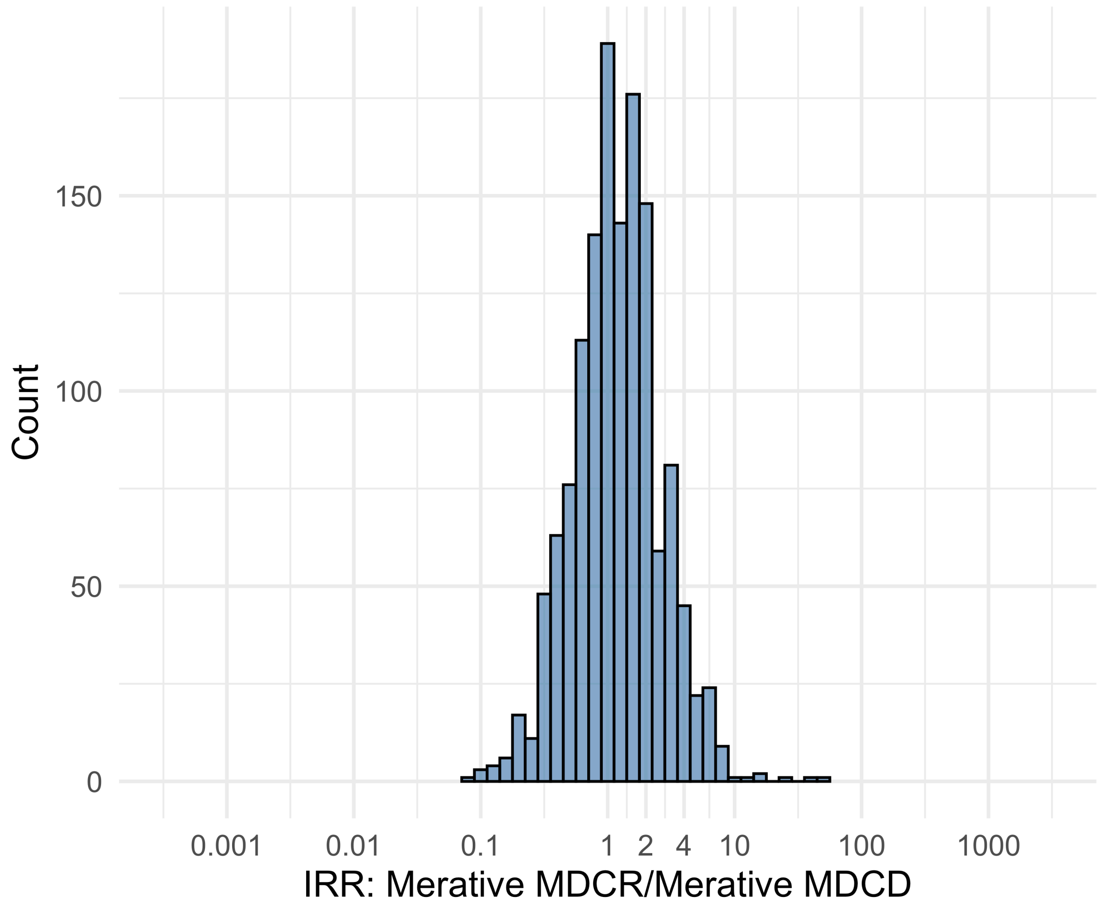


Figure S5.104. IRR distribution comparing Merative MDCR and Merative MDCD.

*Table S5.104. Most extreme IRRs for Merative MDCR vs Merative MDCD (rounded to 0.001).*

| source1 | source2 | Drug and Indication | Outcome | IRR |
|---|---|---|---|---|
| Merative MDCR | Merative MDCD | New users of Tumor Necrosis Factor alpha (TNFa) inhibitors nested in Ulcerative colitis | Persons with gout | 53.0 |
| Merative MDCR | Merative MDCD | New users of Tumor Necrosis Factor alpha (TNFa) inhibitors nested in Ulcerative colitis | Coronary Vessel Revascularization | 42.4 |
| Merative MDCR | Merative MDCD | New users of Tumor Necrosis Factor alpha (TNFa) inhibitors nested in Rheumatoid arthritis | Bladder Cancer | 23.4 |
| Merative MDCR | Merative MDCD | New users of SGLT2 inhibitor nested in Left Heart Failure | Cardiovascular-related mortality | 0.1 |
| Merative MDCR | Merative MDCD | New users of Tumor Necrosis Factor alpha (TNFa) inhibitors nested in Rheumatoid arthritis | Bells Palsy | 0.1 |
| Merative MDCR | Merative MDCD | New users of Tumor Necrosis Factor alpha (TNFa) inhibitors nested in Rheumatoid arthritis | Narcolepsy | 0.1 |

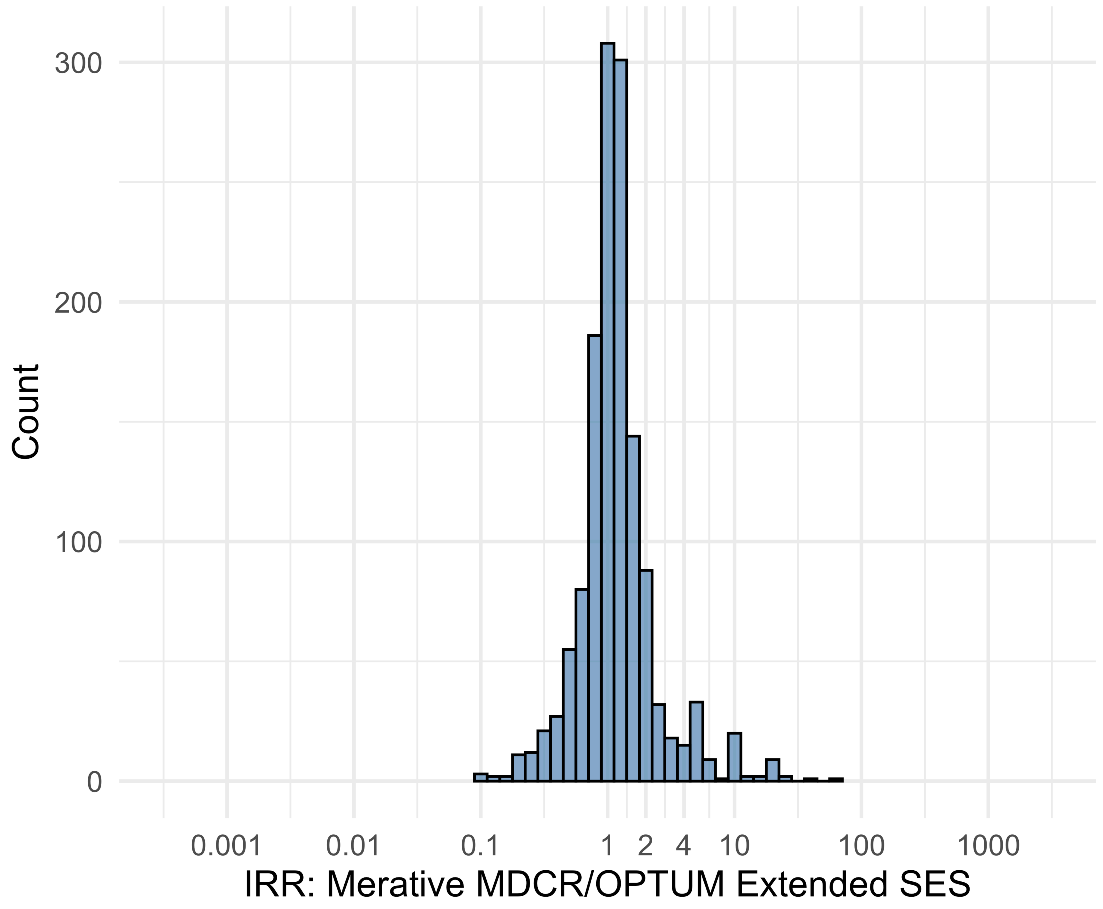


Figure S5.105. IRR distribution comparing Merative MDCR and OPTUM Extended SES.

*Table S5.105. Most extreme IRRs for Merative MDCR vs OPTUM Extended SES (rounded to 0.001).*

| source1 | source2 | Drug and Indication | Outcome | IRR |
|---|---|---|---|---|
| Merative MDCR | OPTUM Extended SES | New users of Tumor Necrosis Factor alpha (TNFa) inhibitors nested in Rheumatoid arthritis | Cardiovascular-related mortality | 59.2 |
| Merative MDCR | OPTUM Extended SES | New users of JAK inhibitors nested in Rheumatoid arthritis | Cardiovascular-related mortality | 35.7 |
| Merative MDCR | OPTUM Extended SES | New users of Trimethoprim systemetic nested in Urinary Tract Infection | Cardiovascular-related mortality | 24.6 |
| Merative MDCR | OPTUM Extended SES | New users of Tumor Necrosis Factor alpha (TNFa) inhibitors nested in Rheumatoid arthritis | Myocarditis Pericarditis | 0.1 |
| Merative MDCR | OPTUM Extended SES | New users of Beta blockers nested in Left Heart Failure | Polymorphic Ventricular Tachycardia or Torsades de Pointes | 0.1 |
| Merative MDCR | OPTUM Extended SES | New users of Tumor Necrosis Factor alpha (TNFa) inhibitors nested in Rheumatoid arthritis | Bells Palsy | 0.1 |

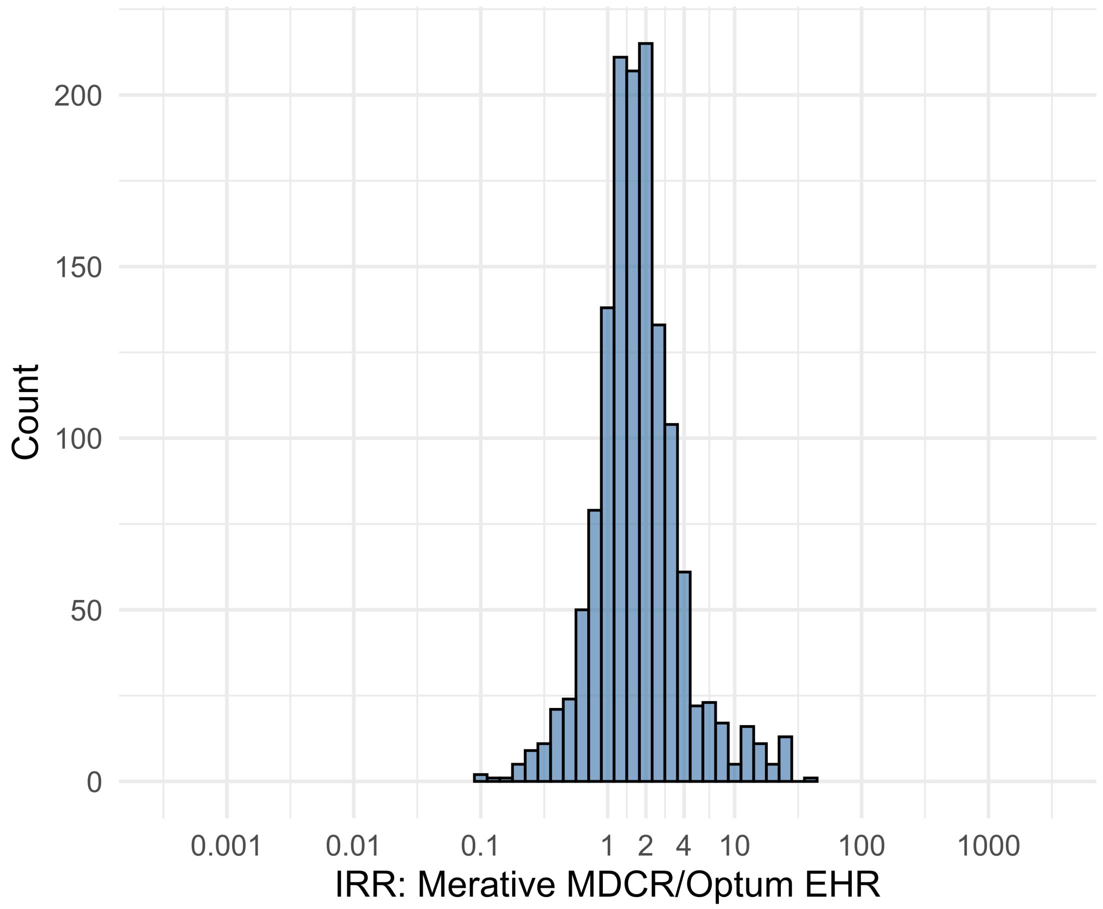


Figure S5.106. IRR distribution comparing Merative MDCR and Optum EHR.

*Table S5.106. Most extreme IRRs for Merative MDCR vs Optum EHR (rounded to 0.001).*

| source1 | source2 | Drug and Indication | Outcome | IRR |
|---|---|---|---|---|
| Merative MDCR | Optum EHR | New users of Tumor Necrosis Factor alpha (TNFa) inhibitors nested in Crohns disease | Polymorphic Ventricular Tachycardia or Torsades de Pointes | 40.9 |
| Merative MDCR | Optum EHR | New users of Tumor Necrosis Factor alpha (TNFa) inhibitors nested in Ulcerative colitis | Guillain Barre Syndrome | 25.0 |
| Merative MDCR | Optum EHR | New users of Tumor Necrosis Factor alpha (TNFa) inhibitors nested in Ulcerative colitis | Progressive multifocal leukoencephalopathy | 24.9 |
| Merative MDCR | Optum EHR | New users of Beta blockers nested in Acute Myocardial Infarction | Thrombosis with Thrombocytopenia (TWT) | 0.1 |
| Merative MDCR | Optum EHR | New users of Beta blockers nested in Left Heart Failure | Polymorphic Ventricular Tachycardia or Torsades de Pointes | 0.1 |
| Merative MDCR | Optum EHR | New users of Tumor Necrosis Factor alpha (TNFa) inhibitors nested in Rheumatoid arthritis | Myocarditis Pericarditis | 0.1 |

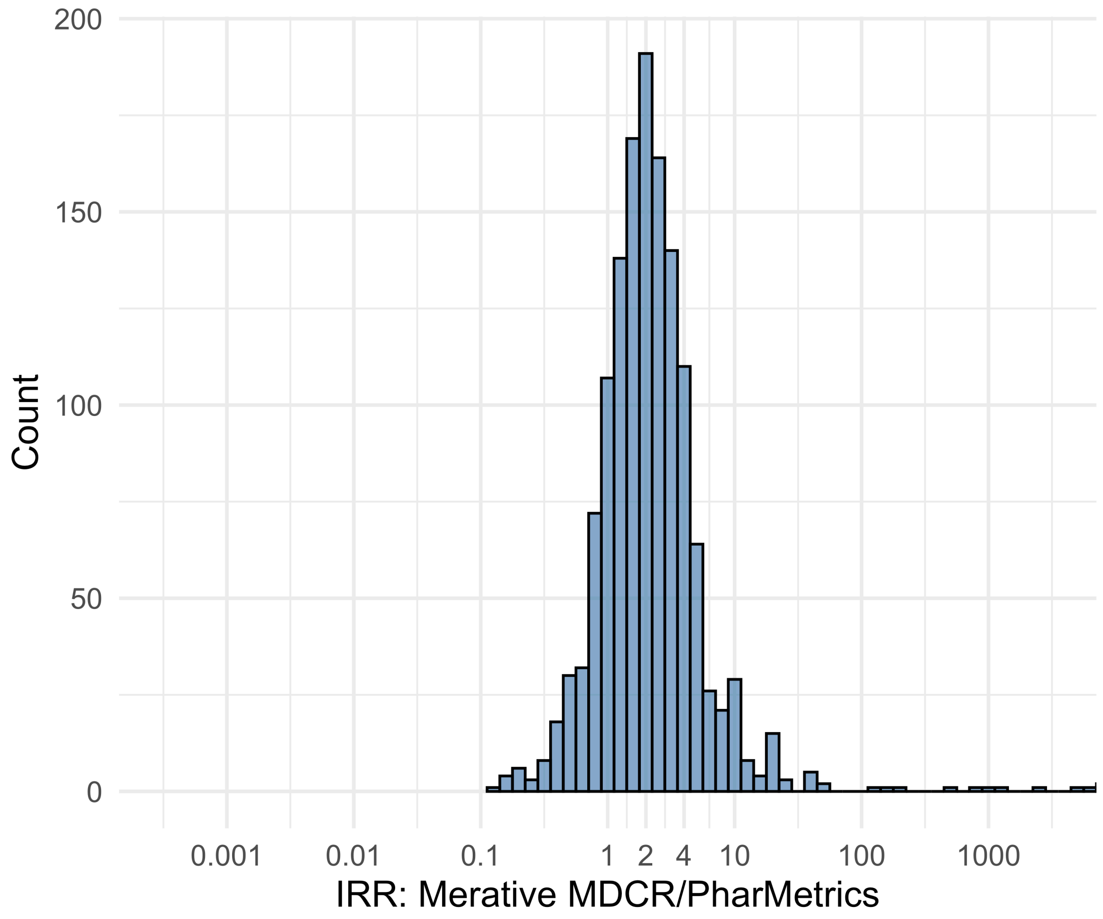


Figure S5.107. IRR distribution comparing Merative MDCR and PharMetrics.

*Table S5.107. Most extreme IRRs for Merative MDCR vs PharMetrics (rounded to 0.001).*

| source1 | source2 | Drug and Indication | Outcome | IRR |
|---|---|---|---|---|
| Merative MDCR | PharMetrics | New users of Beta blockers nested in essential hypertension | Cardiovascular-related mortality | 18312.5 |
| Merative MDCR | PharMetrics | New users of Cephalosporin systemetic nested in Urinary Tract Infection | Cardiovascular-related mortality | 11980.7 |
| Merative MDCR | PharMetrics | New users of Fluoroquinolone systemic nested in Urinary Tract Infection | Cardiovascular-related mortality | 9814.3 |
| Merative MDCR | PharMetrics | New users of Trimethoprim systemetic nested in Acute Typical Pneumonia | Earliest event of Thrombotic microangiopathy (TMA) or Microangiopathic hemolytic anemia (MAHA) | 0.1 |
| Merative MDCR | PharMetrics | New users of Tumor Necrosis Factor alpha (TNFa) inhibitors nested in Rheumatoid arthritis | Myocarditis Pericarditis | 0.2 |
| Merative MDCR | PharMetrics | New users of Tumor Necrosis Factor alpha (TNFa) inhibitors nested in Rheumatoid arthritis | Bells Palsy | 0.2 |

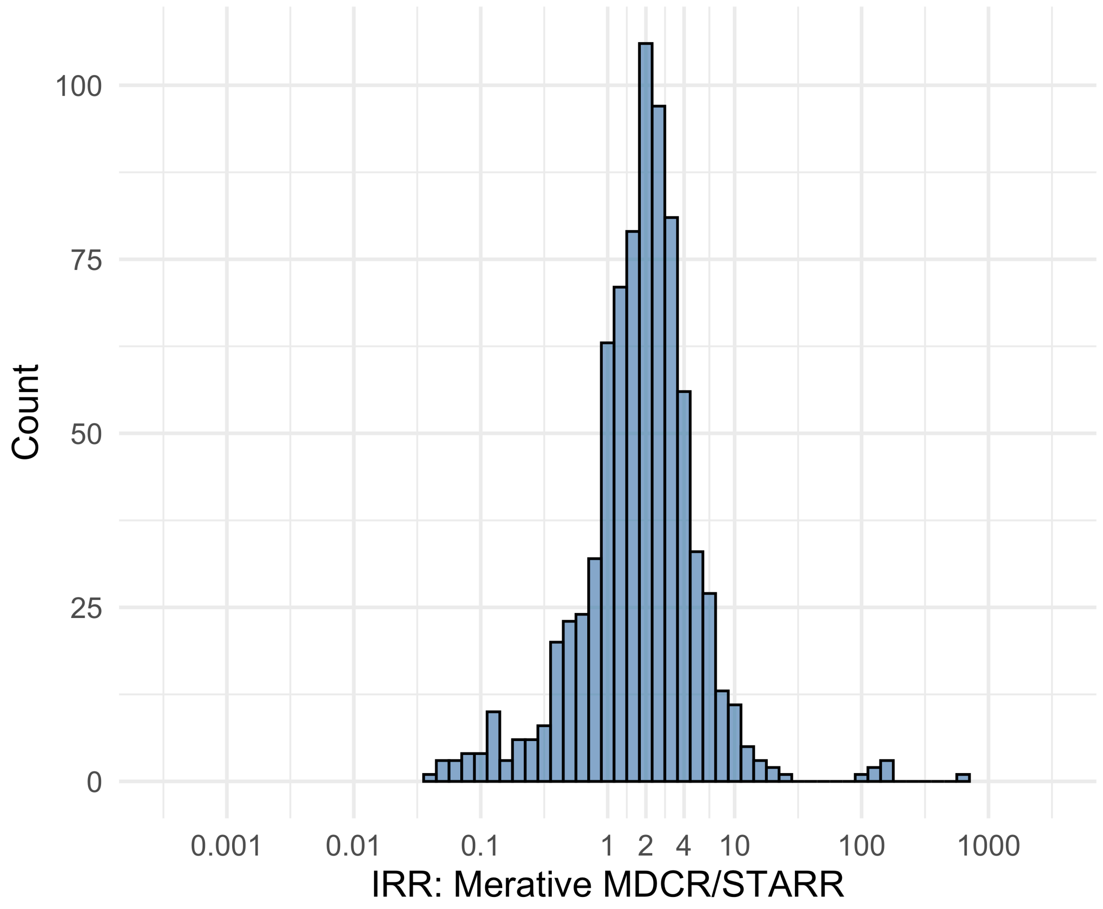


Figure S5.108. IRR distribution comparing Merative MDCR and STARR.

*Table S5.108. Most extreme IRRs for Merative MDCR vs STARR (rounded to 0.001).*

| source1 | source2 | Drug and Indication | Outcome | IRR |
|---|---|---|---|---|
| Merative MDCR | STARR | New users of Beta blockers nested in essential hypertension | Febrile Neutropenia or Neutropenic Fever | 631.4 |
| Merative MDCR | STARR | New users of Fluoroquinolone systemic nested in Urinary Tract Infection | Febrile Neutropenia or Neutropenic Fever | 163.8 |
| Merative MDCR | STARR | New users of Cephalosporin systemetic nested in Urinary Tract Infection | Febrile Neutropenia or Neutropenic Fever | 161.4 |
| Merative MDCR | STARR | New users of Beta blockers nested in Acute Myocardial Infarction | Thrombosis with Thrombocytopenia (TWT) | 0.0 |
| Merative MDCR | STARR | New users of Cephalosporin systemetic nested in Acute Typical Pneumonia | Progressive multifocal leukoencephalopathy | 0.0 |
| Merative MDCR | STARR | New users of Beta blockers nested in Acute Myocardial Infarction | Polymorphic Ventricular Tachycardia or Torsades de Pointes | 0.1 |

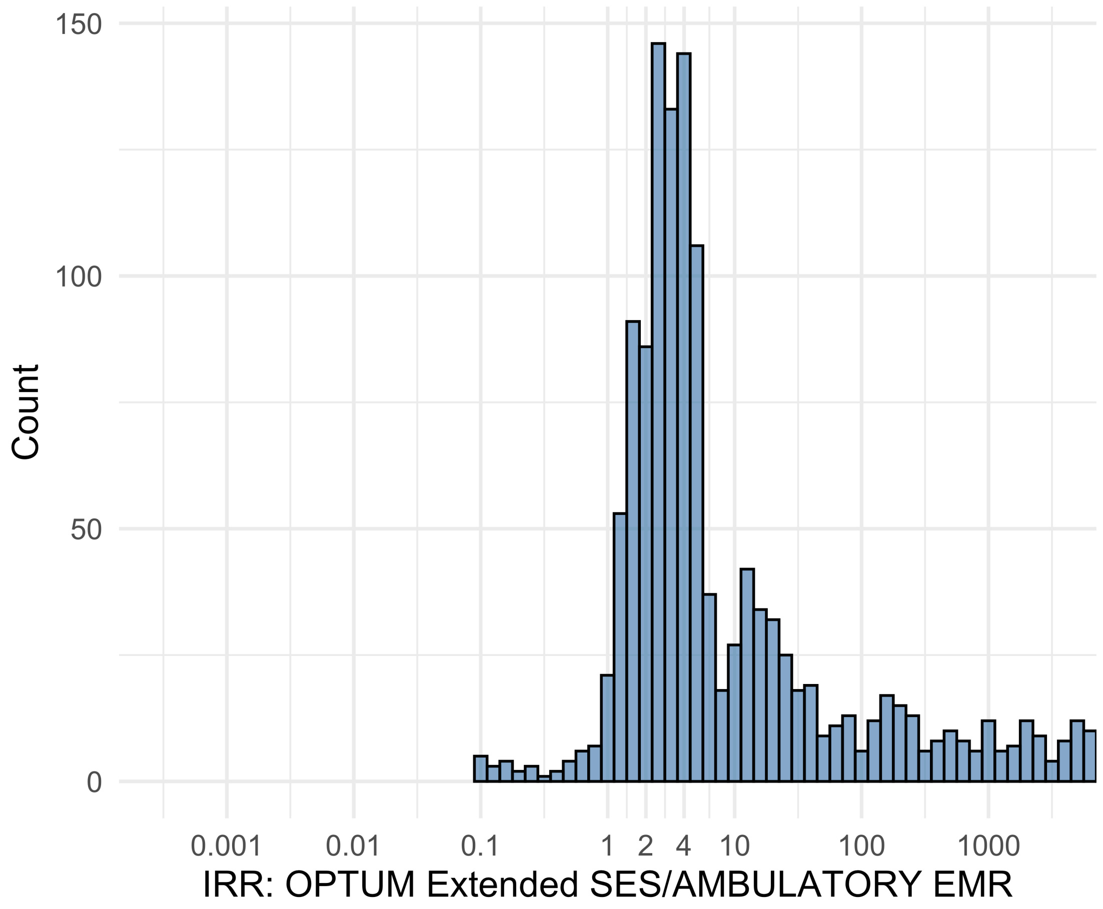


Figure S5.109. IRR distribution comparing OPTUM Extended SES and AMBULATORY EMR.

*Table S5.109. Most extreme IRRs for OPTUM Extended SES vs AMBULATORY EMR (rounded to 0.001).*

| source1 | source2 | Drug and Indication | Outcome | IRR |
|---|---|---|---|---|
| OPTUM Extended SES | AMBULATORY EMR | New users of Beta blockers nested in essential hypertension | Hospitalization with heart failure events | 457206.0 |
| OPTUM Extended SES | AMBULATORY EMR | New users of Beta blockers nested in essential hypertension | 4-point MACE | 347309.0 |
| OPTUM Extended SES | AMBULATORY EMR | New users of Beta blockers nested in essential hypertension | Total cardiovascular disease events (ischemic stroke, hemorrhagic stroke, heart failure, acute myocardial infarction or sudden cardiac death) | 343624.4 |
| OPTUM Extended SES | AMBULATORY EMR | New users of SGLT2 inhibitor nested in Left Heart Failure | Anaphylaxis | 0.1 |
| OPTUM Extended SES | AMBULATORY EMR | New users of Trimethoprim systemetic nested in Acute Typical Pneumonia | Anaphylaxis | 0.1 |

| source1 | source2 | Drug and Indication | Outcome | IRR |
|---|---|---|---|---|
| OPTUM Extended SES | AMBULATORY EMR | New users of SGLT2 inhibitor nested in Type 2 diabetes mellitus | Anaphylaxis | 0.1 |

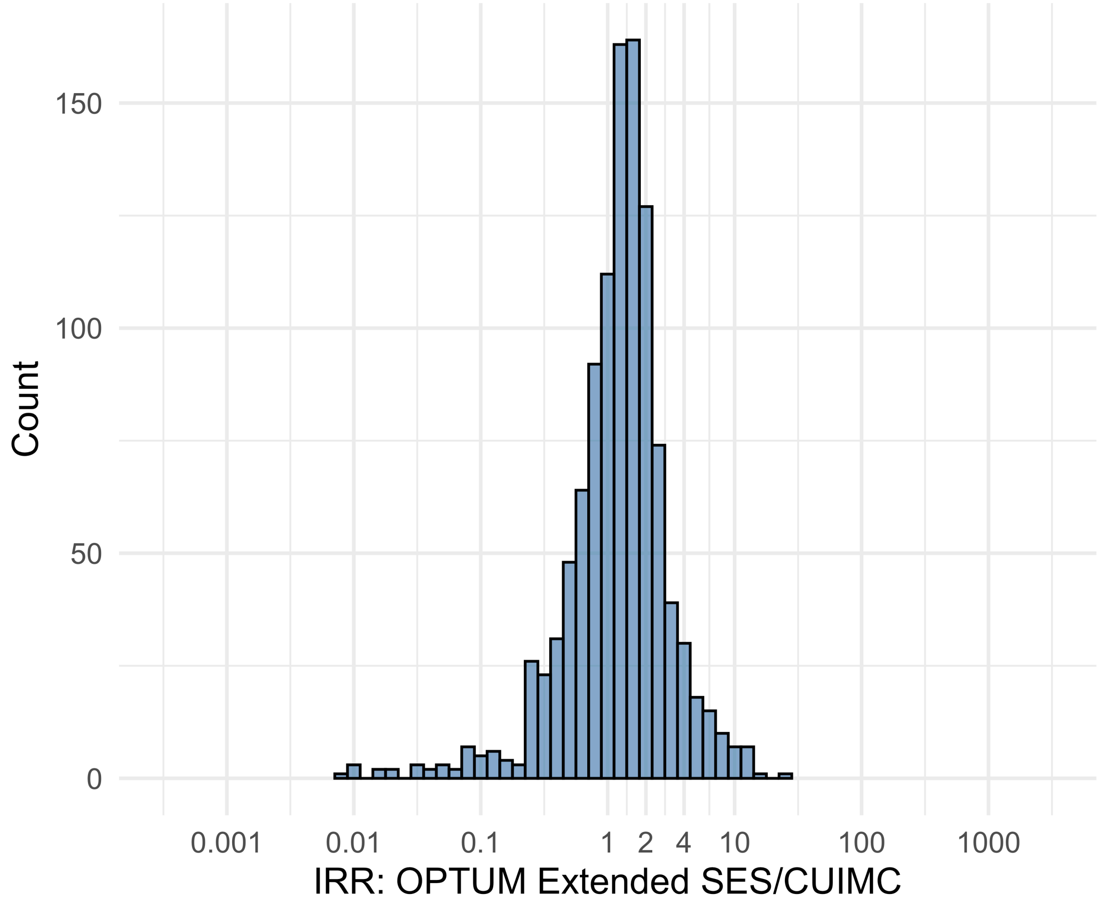


Figure S5.110. IRR distribution comparing OPTUM Extended SES and CUIMC.

*Table S5.110. Most extreme IRRs for OPTUM Extended SES vs CUIMC (rounded to 0.001).*

| source1 | source2 | Drug and Indication | Outcome | IRR |
|---|---|---|---|---|
| OPTUM Extended SES | CUIMC | New users of Beta blockers nested in Acute Myocardial Infarction | Abnormal weight gain events | 27.6 |
| OPTUM Extended SES | CUIMC | New users of SGLT2 inhibitor nested in Left Heart Failure | Rhabdomyolysis | 15.7 |
| OPTUM Extended SES | CUIMC | New users of Tumor Necrosis Factor alpha (TNFa) inhibitors nested in Rheumatoid arthritis | All events of Acute Liver Injury | 13.1 |
| OPTUM Extended SES | CUIMC | New users of Tumor Necrosis Factor alpha (TNFa) inhibitors nested in Rheumatoid arthritis | Cardiovascular-related mortality | 0.0 |
| OPTUM Extended SES | CUIMC | New users of Fluoroquinolone systemic nested in Acute Typical Pneumonia | Cardiovascular-related mortality | 0.0 |
| OPTUM Extended SES | CUIMC | New users of Cephalosporin systemetic nested in Acute Typical Pneumonia | Cardiovascular-related mortality | 0.0 |

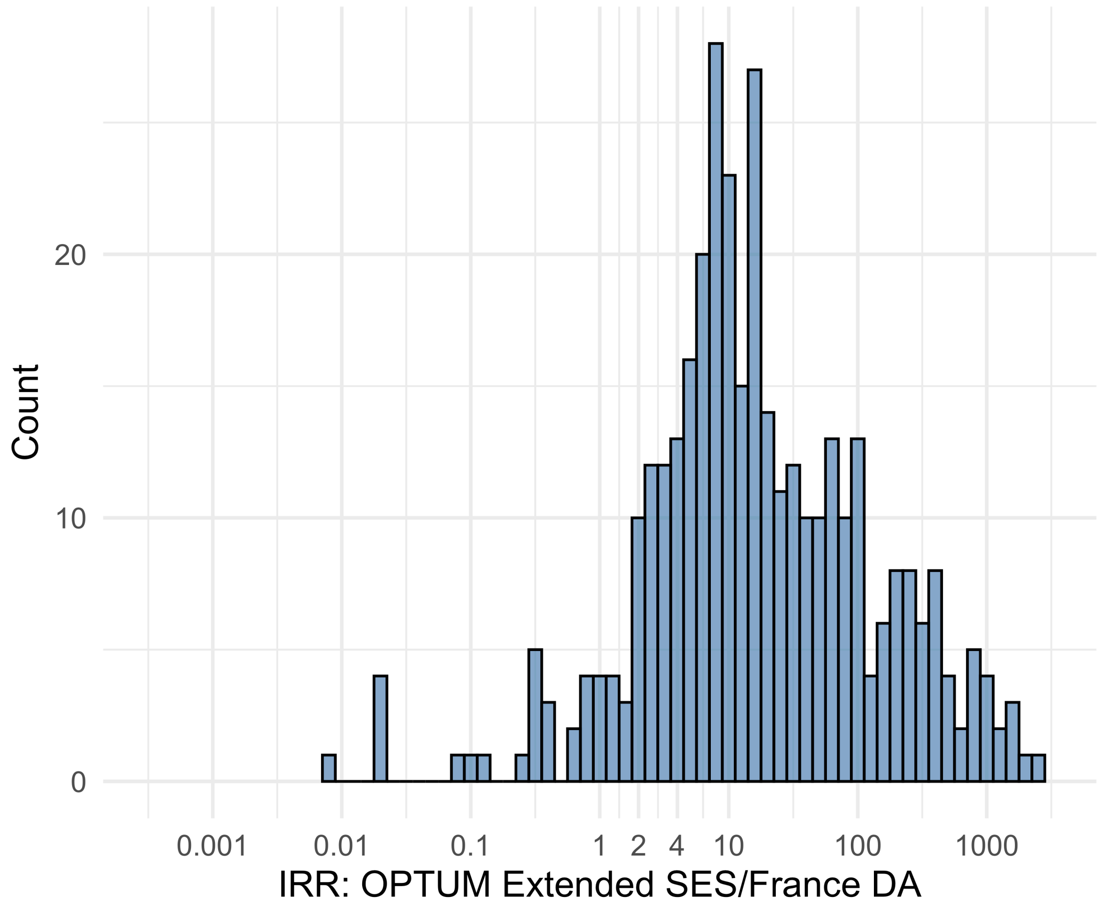


Figure S5.111. IRR distribution comparing OPTUM Extended SES and France DA.

*Table S5.111. Most extreme IRRs for OPTUM Extended SES vs France DA (rounded to 0.001).*

| source1 | source2 | Drug and Indication | Outcome | IRR |
|---|---|---|---|---|
| OPTUM Extended SES | France DA | New users of Beta blockers nested in essential hypertension | Hospitalization with heart failure events | 2246.0 |
| OPTUM Extended SES | France DA | New users of Beta blockers nested in essential hypertension | Acute Kidney Injury AKI | 1810.3 |
| OPTUM Extended SES | France DA | New users of Beta blockers nested in essential hypertension | 4-point MACE | 1706.1 |
| OPTUM Extended SES | France DA | New users of Trimethoprim systemetic nested in Urinary Tract Infection | Polymorphic Ventricular Tachycardia or Torsades de Pointes | 0.0 |
| OPTUM Extended SES | France DA | New users of Fluoroquinolone systemic nested in Urinary Tract Infection | Polymorphic Ventricular Tachycardia or Torsades de Pointes | 0.0 |
| OPTUM Extended SES | France DA | New users of GLP-1 receptor antagonists nested in Type 2 diabetes mellitus | Polymorphic Ventricular Tachycardia or Torsades de Pointes | 0.0 |

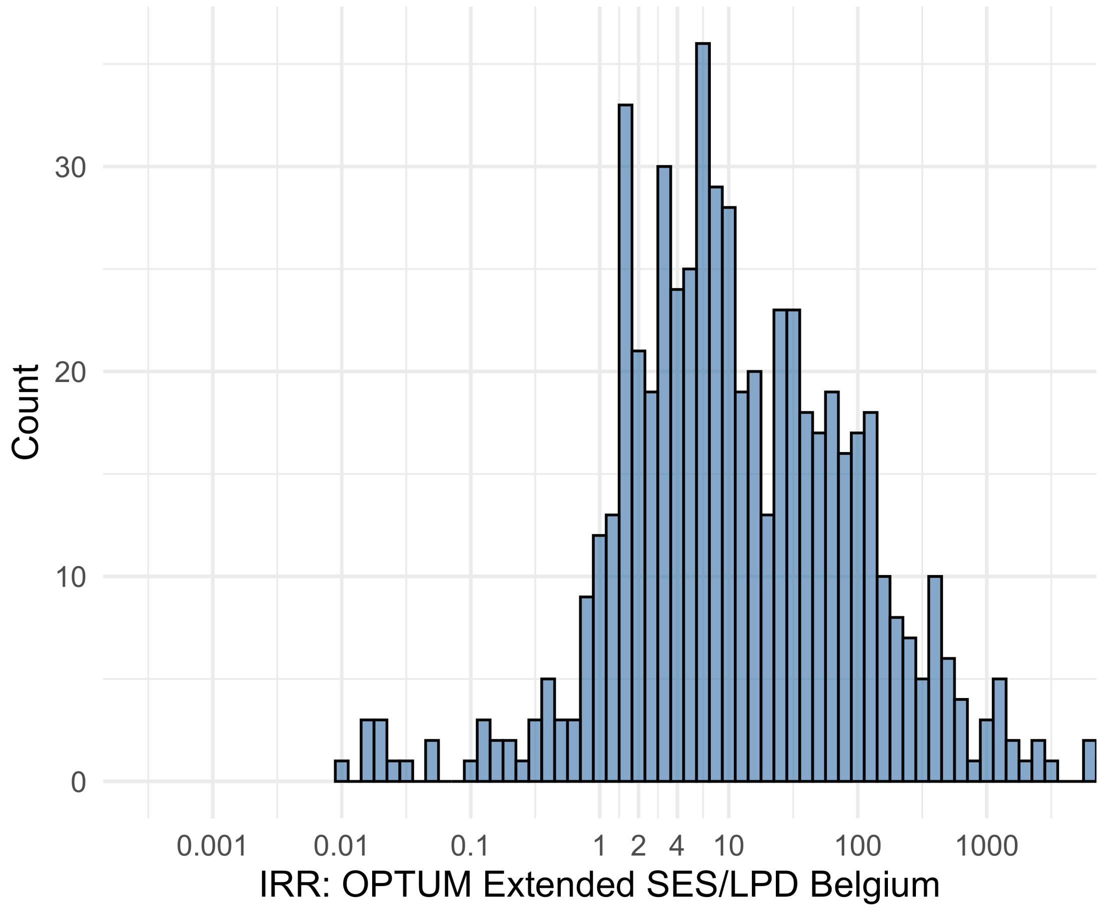


Figure S5.112. IRR distribution comparing OPTUM Extended SES and LPD Belgium.

*Table S5.112. Most extreme IRRs for OPTUM Extended SES vs LPD Belgium (rounded to 0.001).*

| source1 | source2 | Drug and Indication | Outcome | IRR |
|---|---|---|---|---|
| OPTUM Extended SES | LPD Belgium | New users of Beta blockers nested in essential hypertension | Hospitalization with heart failure events | 8740.9 |
| OPTUM Extended SES | LPD Belgium | New users of Beta blockers nested in essential hypertension | 4-point MACE | 6639.9 |
| OPTUM Extended SES | LPD Belgium | New users of Beta blockers nested in essential hypertension | Total cardiovascular disease events (ischemic stroke, hemorrhagic stroke, heart failure, acute myocardial infarction or sudden cardiac death) | 6569.5 |
| OPTUM Extended SES | LPD Belgium | New users of Trimethoprim systemetic nested in Urinary Tract Infection | Polymorphic Ventricular Tachycardia or Torsades de Pointes | 0.0 |
| OPTUM Extended SES | LPD Belgium | New users of GLP-1 receptor antagonists nested in obesity | Polymorphic Ventricular Tachycardia or Torsades de Pointes | 0.0 |
| OPTUM Extended SES | LPD Belgium | New users of GLP-1 receptor antagonists nested in obesity | All events Drug Rash with Eosinophilia and Systemic Symptoms (DRESS) | 0.0 |

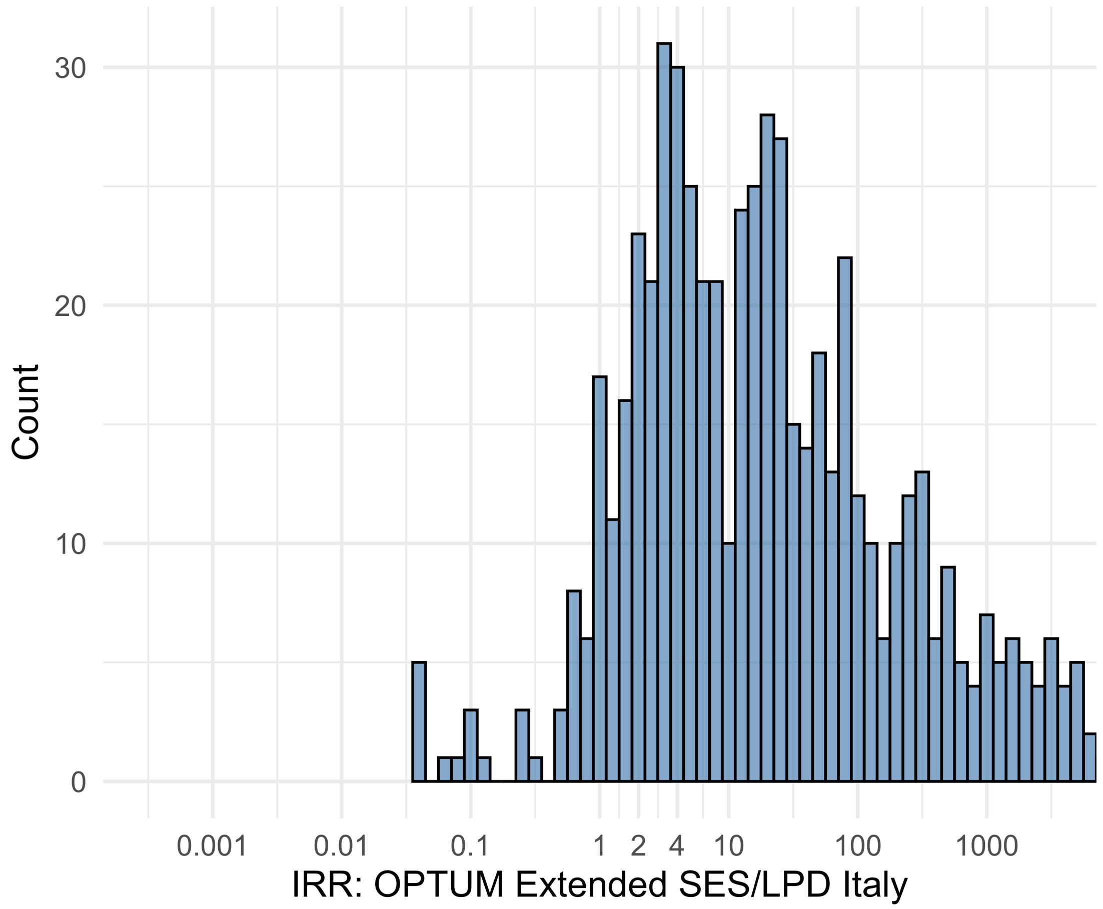


Figure S5.113. IRR distribution comparing OPTUM Extended SES and LPD Italy.

*Table S5.113. Most extreme IRRs for OPTUM Extended SES vs LPD Italy (rounded to 0.001).*

| source1 | source2 | Drug and Indication | Outcome | IRR |
|---|---|---|---|---|
| OPTUM Extended SES | LPD Italy | New users of Beta blockers nested in essential hypertension | Hospitalization with heart failure events | 28419.2 |
| OPTUM Extended SES | LPD Italy | New users of Beta blockers nested in essential hypertension | 4-point MACE | 21588.2 |
| OPTUM Extended SES | LPD Italy | New users of Beta blockers nested in essential hypertension | Total cardiovascular disease events (ischemic stroke, hemorrhagic stroke, heart failure, acute myocardial infarction or sudden cardiac death) | 21359.2 |
| OPTUM Extended SES | LPD Italy | New users of GLP-1 receptor antagonists nested in obesity | Progressive multifocal leukoencephalopathy | 0.0 |
| OPTUM Extended SES | LPD Italy | New users of GLP-1 receptor antagonists nested in obesity | Polymorphic Ventricular Tachycardia or Torsades de Pointes | 0.0 |
| OPTUM Extended SES | LPD Italy | New users of GLP-1 receptor antagonists nested in obesity | All events Drug Rash with Eosinophilia and Systemic Symptoms (DRESS) | 0.0 |

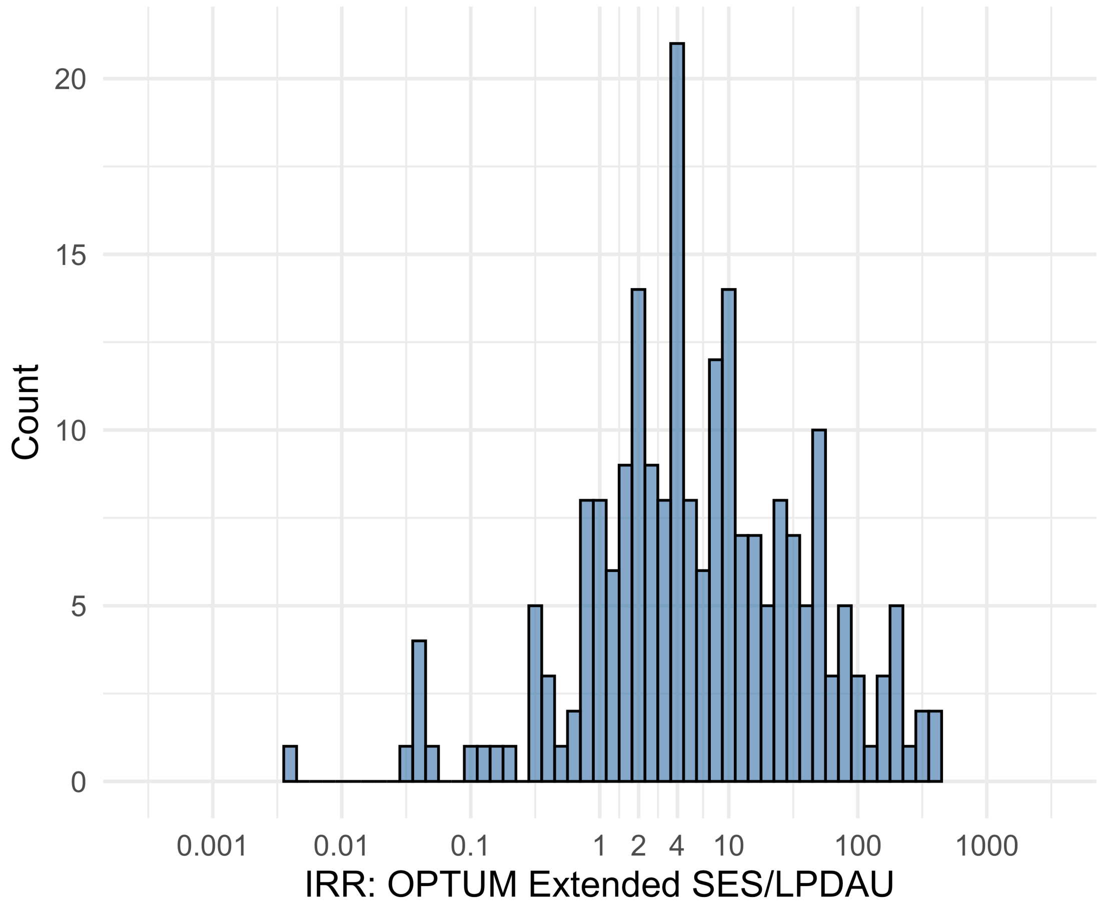


Figure S5.114. IRR distribution comparing OPTUM Extended SES and LPDAU.

*Table S5.114. Most extreme IRRs for OPTUM Extended SES vs LPDAU (rounded to 0.001).*

| source1 | source2 | Drug and Indication | Outcome | IRR |
|---|---|---|---|---|
| OPTUM Extended SES | LPDAU | New users of Cephalosporin systemetic nested in Urinary Tract Infection | Acute Kidney Injury AKI | 407.3 |
| OPTUM Extended SES | LPDAU | New users of Cephalosporin systemetic nested in Urinary Tract Infection | Hospitalization with heart failure events | 378.1 |
| OPTUM Extended SES | LPDAU | New users of Cephalosporin systemetic nested in Urinary Tract Infection | 4-point MACE | 308.8 |
| OPTUM Extended SES | LPDAU | New users of Trimethoprim systemetic nested in Urinary Tract Infection | Polymorphic Ventricular Tachycardia or Torsades de Pointes | 0.0 |
| OPTUM Extended SES | LPDAU | New users of Beta blockers nested in essential hypertension | Progressive multifocal leukoencephalopathy | 0.0 |
| OPTUM Extended SES | LPDAU | New users of Cephalosporin systemetic nested in Urinary Tract Infection | Polymorphic Ventricular Tachycardia or Torsades de Pointes | 0.0 |

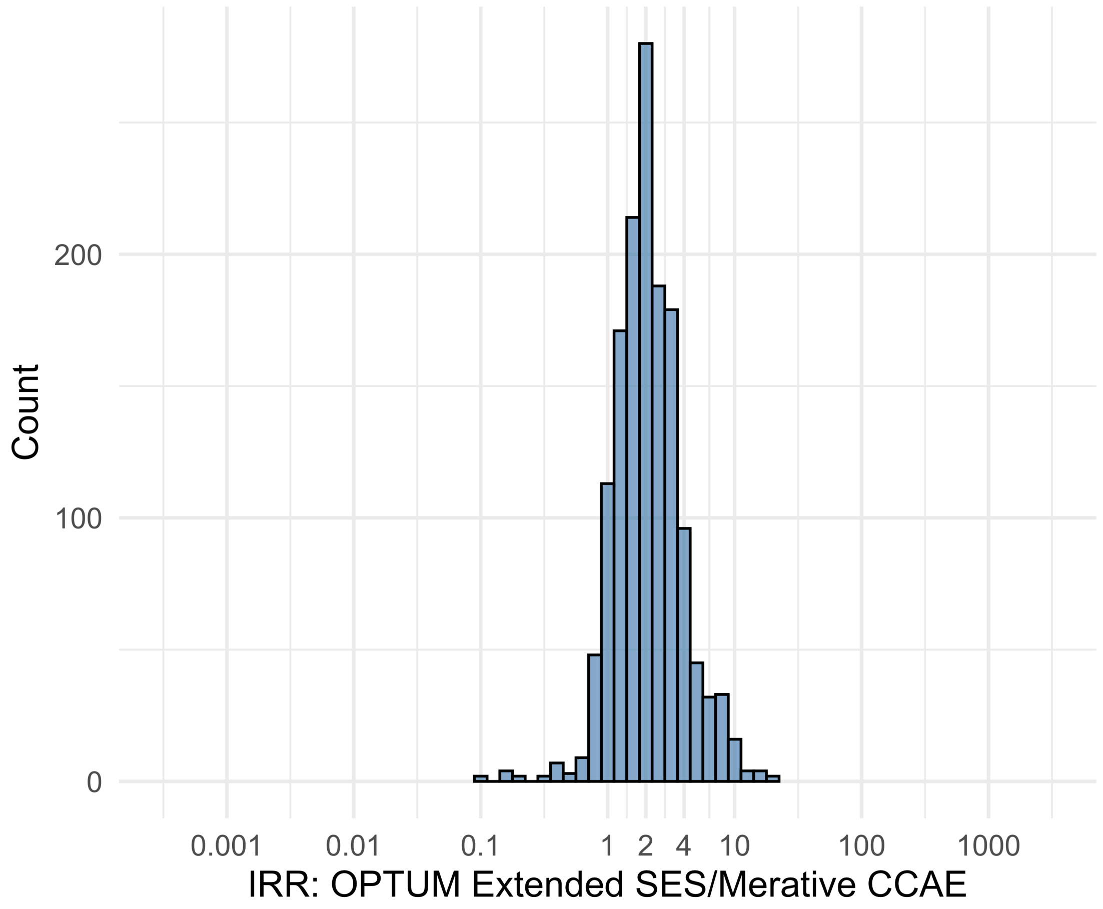


Figure S5.115. IRR distribution comparing OPTUM Extended SES and Merative CCAE.

*Table S5.115. Most extreme IRRs for OPTUM Extended SES vs Merative CCAE (rounded to 0.001).*

| source1 | source2 | Drug and Indication | Outcome | IRR |
|---|---|---|---|---|
| OPTUM Extended SES | Merative CCAE | New users of Beta blockers nested in essential hypertension | Polymorphic Ventricular Tachycardia or Torsades de Pointes | 20.7 |
| OPTUM Extended SES | Merative CCAE | New users of SGLT2 inhibitor nested in Type 2 diabetes mellitus | Polymorphic Ventricular Tachycardia or Torsades de Pointes | 18.2 |
| OPTUM Extended SES | Merative CCAE | New users of Tumor Necrosis Factor alpha (TNFa) inhibitors nested in Psoriatic Arthritis | Lower extremity amputation | 15.8 |
| OPTUM Extended SES | Merative CCAE | New users of Trimethoprim systemetic nested in Acute Typical Pneumonia | Progressive multifocal leukoencephalopathy | 0.1 |
| OPTUM Extended SES | Merative CCAE | New users of Tumor Necrosis Factor alpha (TNFa) inhibitors nested in Rheumatoid arthritis | Cardiovascular-related mortality | 0.1 |
| OPTUM Extended SES | Merative CCAE | New users of Tumor Necrosis Factor alpha (TNFa) inhibitors nested in Psoriatic Arthritis | Isolated Immune Thrombocytopenia (ITP), with a washout period of 365 days | 0.1 |

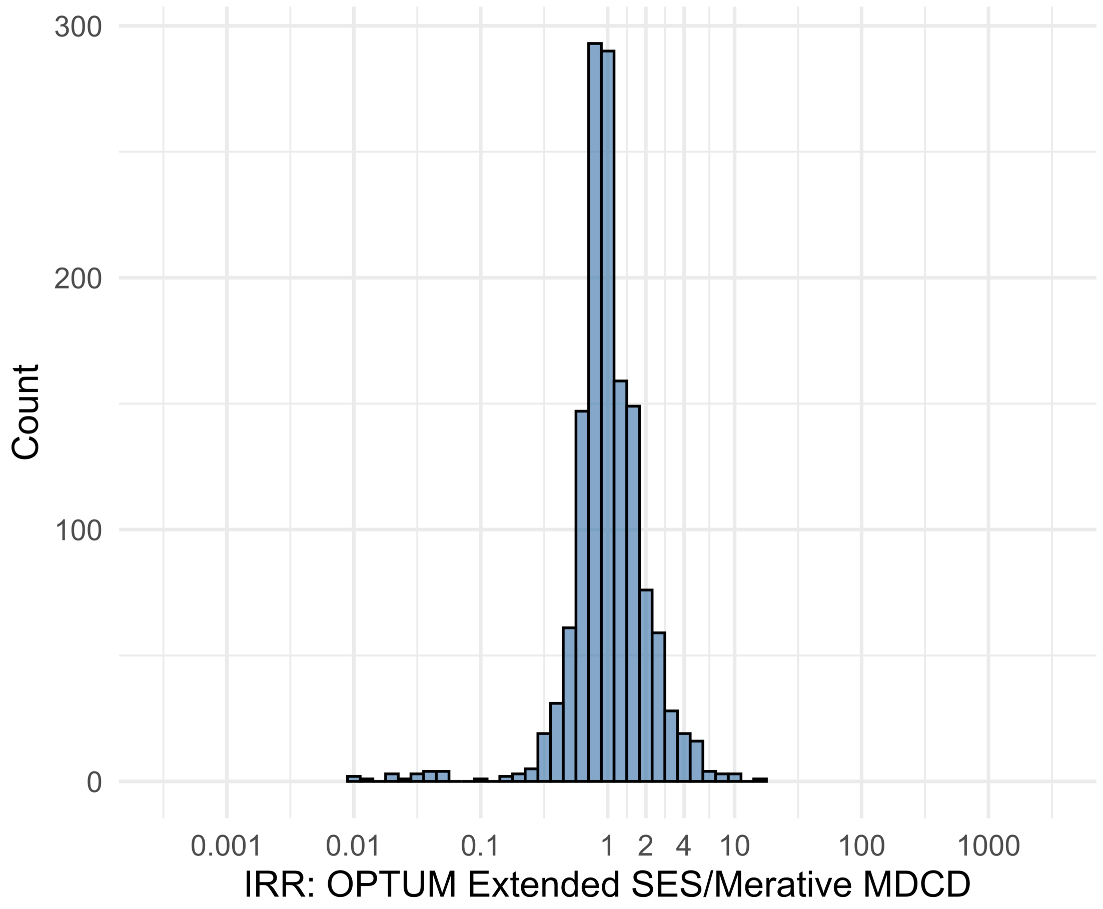


Figure S5.116. IRR distribution comparing OPTUM Extended SES and Merative MDCD.

*Table S5.116. Most extreme IRRs for OPTUM Extended SES vs Merative MDCD (rounded to 0.001).*

| source1 | source2 | Drug and Indication | Outcome | IRR |
|---|---|---|---|---|
| OPTUM Extended SES | Merative MDCD | New users of Tumor Necrosis Factor alpha (TNFa) inhibitors nested in Ulcerative colitis | Persons with gout | 16.3 |
| OPTUM Extended SES | Merative MDCD | New users of Trimethoprim systemetic nested in Acute Typical Pneumonia | All events of Autoimmune hepatitis, with a washout period of 365 days | 10.5 |
| OPTUM Extended SES | Merative MDCD | New users of Tumor Necrosis Factor alpha (TNFa) inhibitors nested in Rheumatoid arthritis | Autoimmune hemolytic anemia | 9.6 |
| OPTUM Extended SES | Merative MDCD | New users of JAK inhibitors nested in Rheumatoid arthritis | Cardiovascular-related mortality | 0.0 |
| OPTUM Extended SES | Merative MDCD | New users of Tumor Necrosis Factor alpha (TNFa) inhibitors nested in Rheumatoid arthritis | Cardiovascular-related mortality | 0.0 |
| OPTUM Extended SES | Merative MDCD | New users of Tumor Necrosis Factor alpha (TNFa) inhibitors nested in Psoriatic Arthritis | Cardiovascular-related mortality | 0.0 |

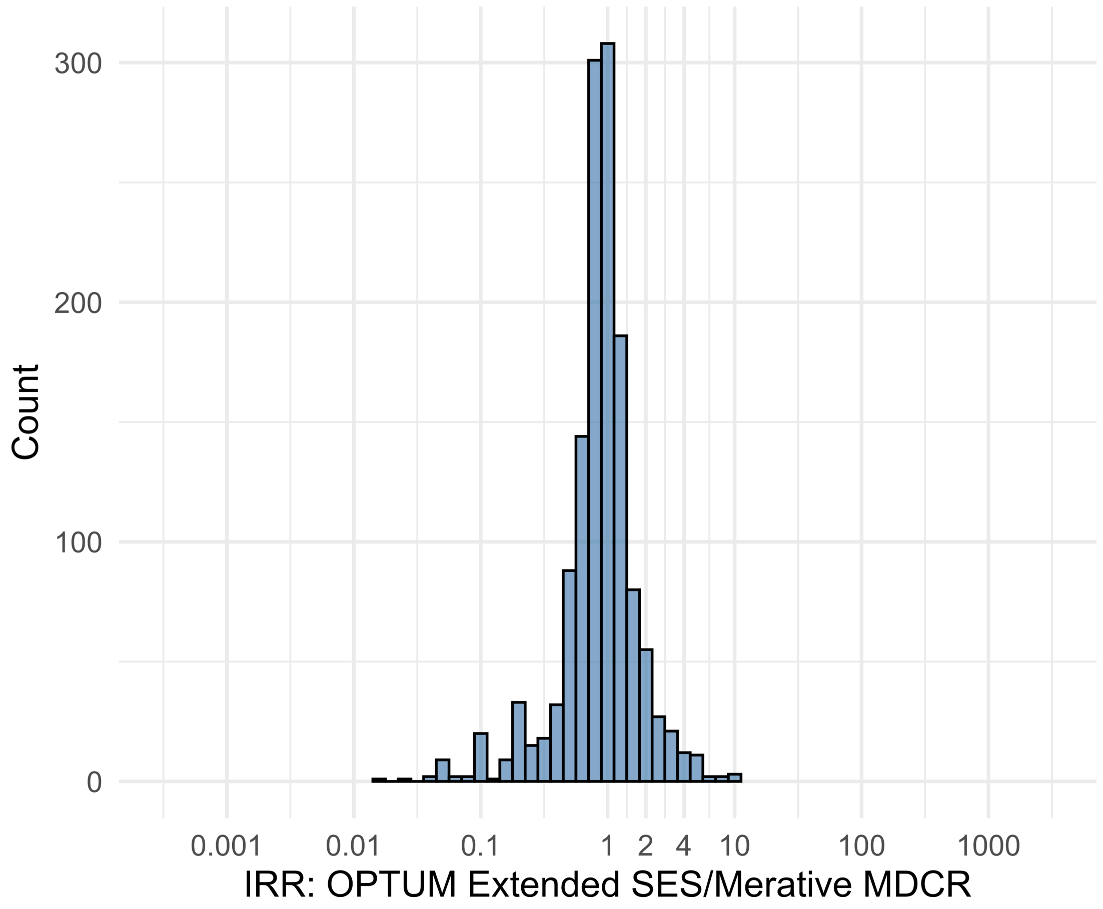


Figure S5.117. IRR distribution comparing OPTUM Extended SES and Merative MDCR.

*Table S5.117. Most extreme IRRs for OPTUM Extended SES vs Merative MDCR (rounded to 0.001).*

| source1 | source2 | Drug and Indication | Outcome | IRR |
|---|---|---|---|---|
| OPTUM Extended SES | Merative MDCR | New users of Tumor Necrosis Factor alpha (TNFa) inhibitors nested in Rheumatoid arthritis | Myocarditis Pericarditis | 10.7 |
| OPTUM Extended SES | Merative MDCR | New users of Beta blockers nested in Left Heart Failure | Polymorphic Ventricular Tachycardia or Torsades de Pointes | 10.6 |
| OPTUM Extended SES | Merative MDCR | New users of Tumor Necrosis Factor alpha (TNFa) inhibitors nested in Rheumatoid arthritis | Bells Palsy | 10.0 |
| OPTUM Extended SES | Merative MDCR | New users of Tumor Necrosis Factor alpha (TNFa) inhibitors nested in Rheumatoid arthritis | Cardiovascular-related mortality | 0.0 |
| OPTUM Extended SES | Merative MDCR | New users of JAK inhibitors nested in Rheumatoid arthritis | Cardiovascular-related mortality | 0.0 |
| OPTUM Extended SES | Merative MDCR | New users of Trimethoprim systemetic nested in Urinary Tract Infection | Cardiovascular-related mortality | 0.0 |

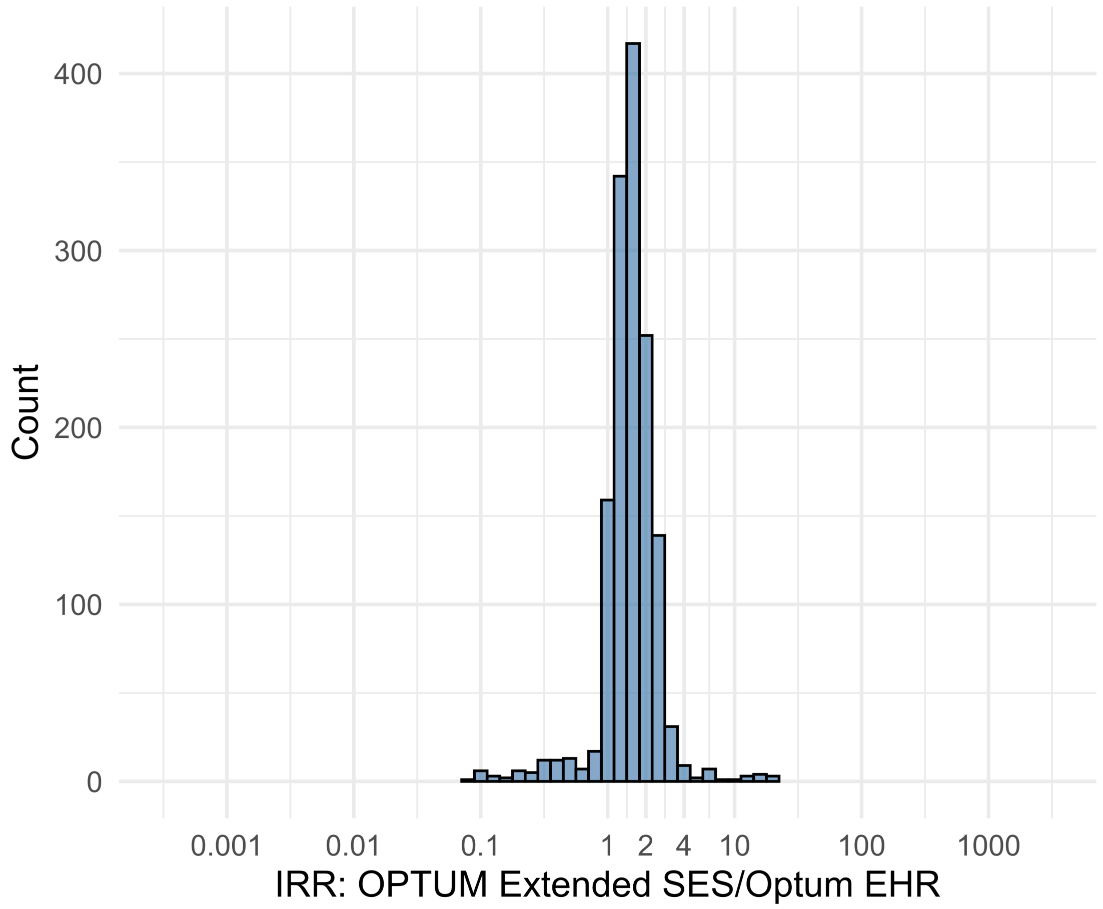


Figure S5.118. IRR distribution comparing OPTUM Extended SES and Optum EHR.

*Table S5.118. Most extreme IRRs for OPTUM Extended SES vs Optum EHR (rounded to 0.001).*

| source1 | source2 | Drug and Indication | Outcome | IRR |
|---|---|---|---|---|
| OPTUM Extended SES | Optum EHR | New users of Tumor Necrosis Factor alpha (TNFa) inhibitors nested in Rheumatoid arthritis | Stevens-Johnson syndrome, toxic epidermal necrolysis spectrum | 20.2 |
| OPTUM Extended SES | Optum EHR | New users of Tumor Necrosis Factor alpha (TNFa) inhibitors nested in Crohns disease | Acquired Pure Red Cell Aplasia | 18.8 |
| OPTUM Extended SES | Optum EHR | New users of Tumor Necrosis Factor alpha (TNFa) inhibitors nested in Psoriatic Arthritis | Rhabdomyolysis | 18.5 |
| OPTUM Extended SES | Optum EHR | New users of Tumor Necrosis Factor alpha (TNFa) inhibitors nested in Rheumatoid arthritis | Cardiovascular-related mortality | 0.1 |
| OPTUM Extended SES | Optum EHR | New users of Fluoroquinolone systemic nested in Acute Typical Pneumonia | Cardiovascular-related mortality | 0.1 |
| OPTUM Extended SES | Optum EHR | New users of Cephalosporin systemetic nested in Acute Typical Pneumonia | Cardiovascular-related mortality | 0.1 |

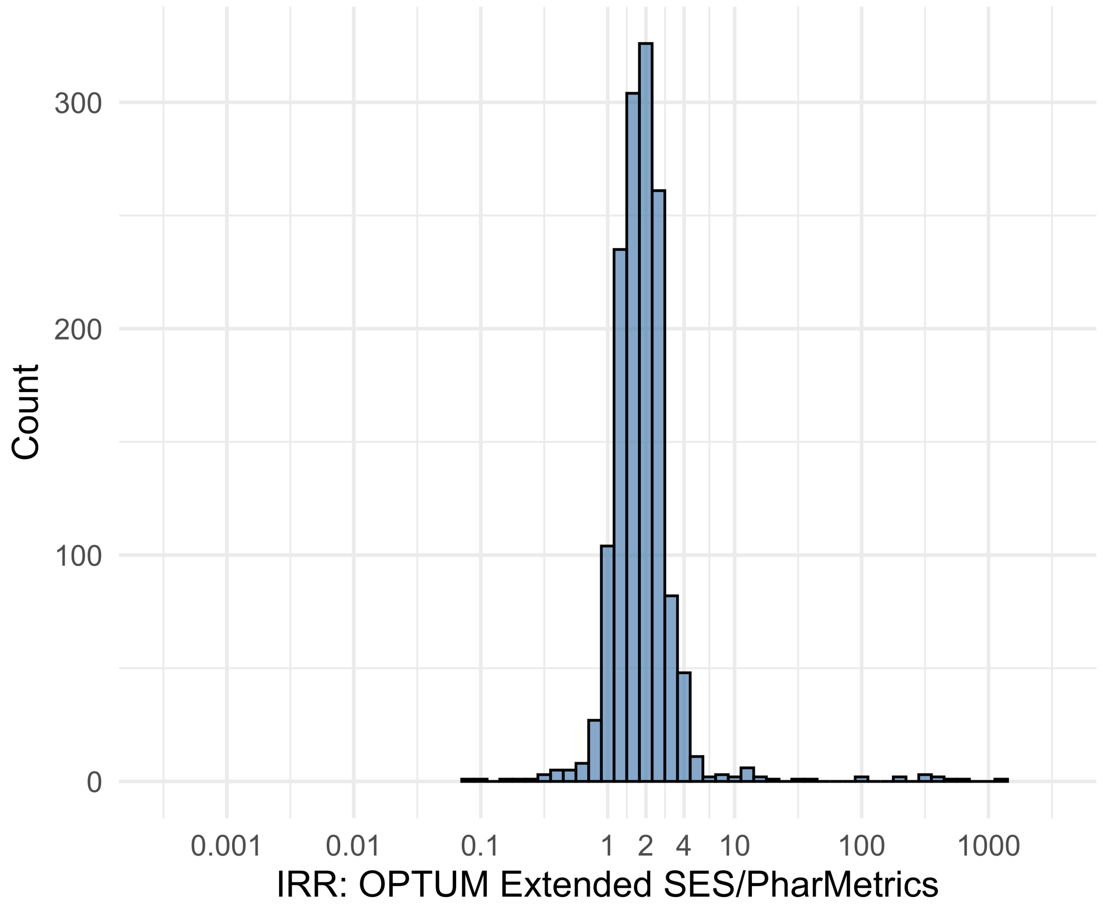


Figure S5.119. IRR distribution comparing OPTUM Extended SES and PharMetrics.

*Table S5.119. Most extreme IRRs for OPTUM Extended SES vs PharMetrics (rounded to 0.001).*

| source1 | source2 | Drug and Indication | Outcome | IRR |
|---|---|---|---|---|
| OPTUM Extended SES | PharMetrics | New users of Beta blockers nested in essential hypertension | Cardiovascular-related mortality | 1203.5 |
| OPTUM Extended SES | PharMetrics | New users of Cephalosporin systemetic nested in Urinary Tract Infection | Cardiovascular-related mortality | 623.2 |
| OPTUM Extended SES | PharMetrics | New users of Cephalosporin systemetic nested in Acute Typical Pneumonia | Cardiovascular-related mortality | 462.8 |
| OPTUM Extended SES | PharMetrics | New users of Tumor Necrosis Factor alpha (TNFa) inhibitors nested in Ulcerative colitis | Autoimmune hemolytic anemia | 0.1 |
| OPTUM Extended SES | PharMetrics | New users of Tumor Necrosis Factor alpha (TNFa) inhibitors nested in Psoriatic Arthritis | Isolated Immune Thrombocytopenia (ITP), with a washout period of 365 days | 0.1 |
| OPTUM Extended SES | PharMetrics | New users of Tumor Necrosis Factor alpha (TNFa) inhibitors nested in Ulcerative colitis | Guillain Barre Syndrome | 0.2 |

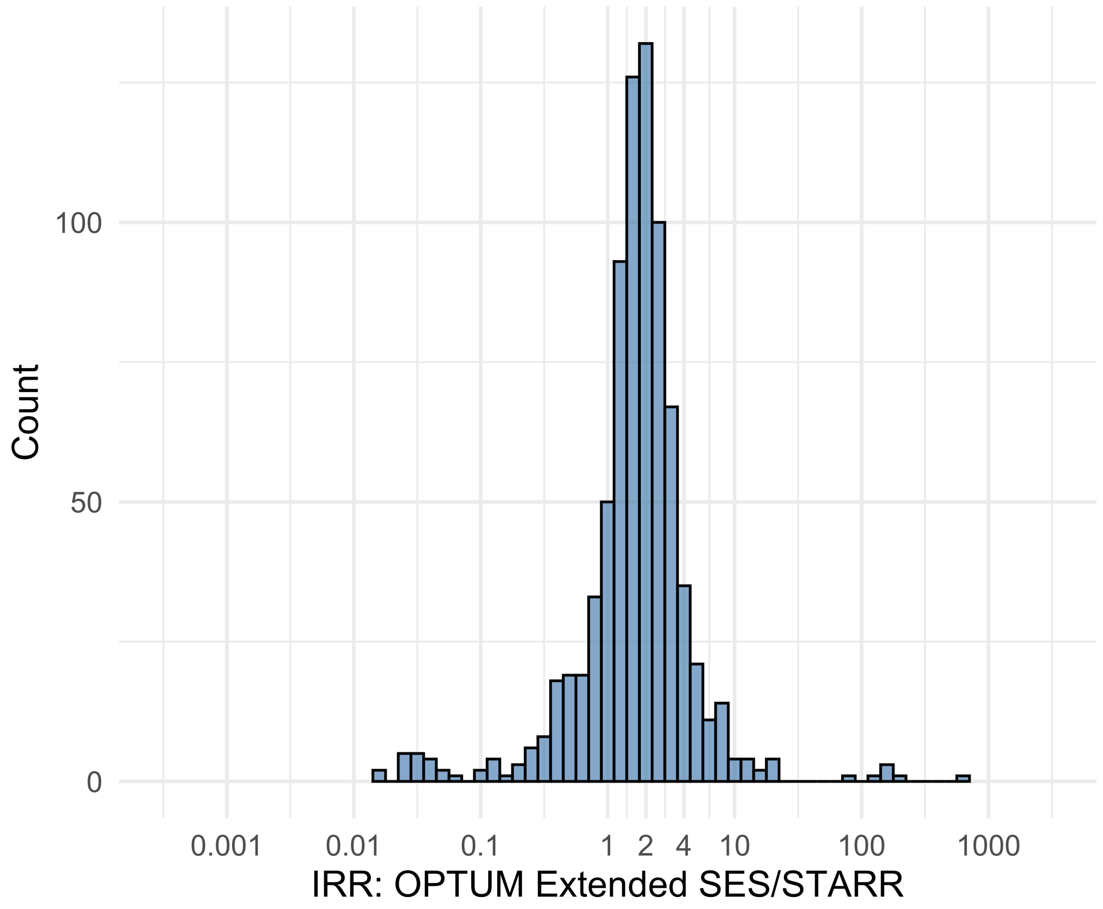


Figure S5.120. IRR distribution comparing OPTUM Extended SES and STARR.

*Table S5.120. Most extreme IRRs for OPTUM Extended SES vs STARR (rounded to 0.001).*

| source1 | source2 | Drug and Indication | Outcome | IRR |
|---|---|---|---|---|
| OPTUM Extended SES | STARR | New users of Beta blockers nested in essential hypertension | Febrile Neutropenia or Neutropenic Fever | 686.2 |
| OPTUM Extended SES | STARR | New users of Beta blockers nested in Left Heart Failure | Febrile Neutropenia or Neutropenic Fever | 199.1 |
| OPTUM Extended SES | STARR | New users of Cephalosporin systemetic nested in Urinary Tract Infection | Febrile Neutropenia or Neutropenic Fever | 164.4 |
| OPTUM Extended SES | STARR | New users of SGLT2 inhibitor nested in Type 2 diabetes mellitus | Progressive multifocal leukoencephalopathy | 0.0 |
| OPTUM Extended SES | STARR | New users of Fluoroquinolone systemic nested in Acute Typical Pneumonia | Cardiovascular-related mortality | 0.0 |
| OPTUM Extended SES | STARR | New users of Cephalosporin systemetic nested in Acute Typical Pneumonia | Cardiovascular-related mortality | 0.0 |

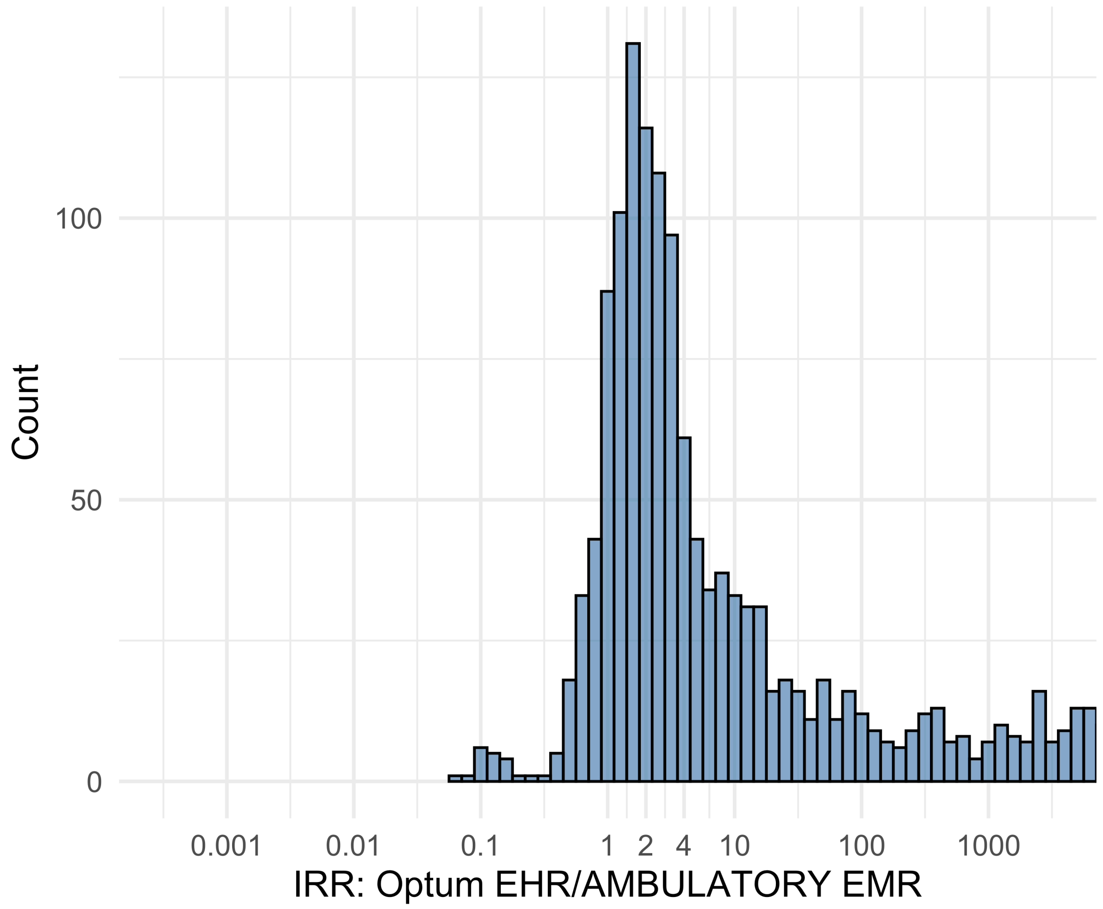


Figure S5.121. IRR distribution comparing Optum EHR and AMBULATORY EMR.

*Table S5.121. Most extreme IRRs for Optum EHR vs AMBULATORY EMR (rounded to 0.001).*

| source1 | source2 | Drug and Indication | Outcome | IRR |
|---|---|---|---|---|
| Optum EHR | AMBULATORY EMR | New users of Beta blockers nested in essential hypertension | Hospitalization with heart failure events | 256048.1 |
| Optum EHR | AMBULATORY EMR | New users of Beta blockers nested in essential hypertension | 4-point MACE | 218330.0 |
| Optum EHR | AMBULATORY EMR | New users of Beta blockers nested in essential hypertension | Total cardiovascular disease events (ischemic stroke, hemorrhagic stroke, heart failure, acute myocardial infarction or sudden cardiac death) | 216660.9 |
| Optum EHR | AMBULATORY EMR | New users of Tumor Necrosis Factor alpha (TNFa) inhibitors nested in Psoriatic Arthritis | Anaphylaxis | 0.1 |
| Optum EHR | AMBULATORY EMR | New users of Trimethoprim systemetic nested in Acute Typical Pneumonia | Anaphylaxis | 0.1 |
| Optum EHR | AMBULATORY EMR | New users of Cephalosporin systemetic nested in Urinary Tract Infection | Anaphylaxis | 0.1 |

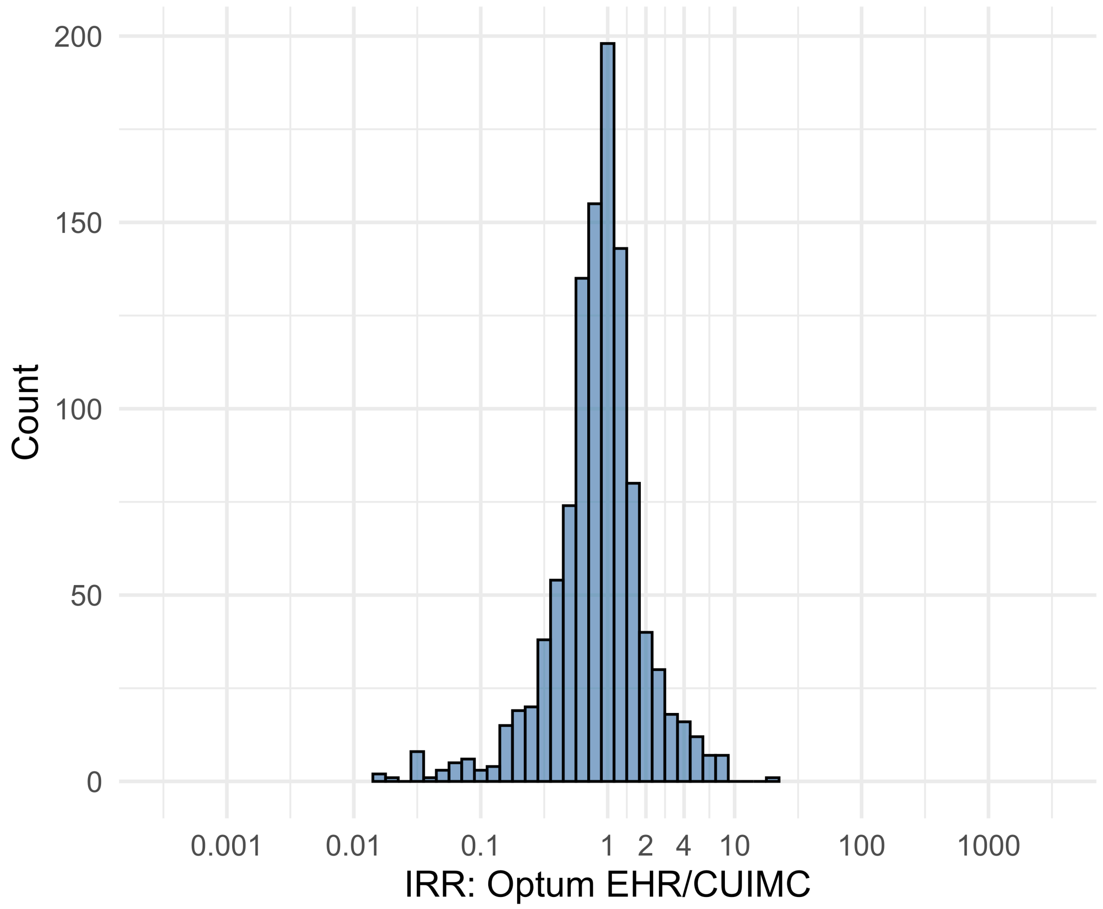


Figure S5.122. IRR distribution comparing Optum EHR and CUIMC.

*Table S5.122. Most extreme IRRs for Optum EHR vs CUIMC (rounded to 0.001).*

| source1 | source2 | Drug and Indication | Outcome | IRR |
|---|---|---|---|---|
| Optum EHR | CUIMC | New users of Beta blockers nested in Acute Myocardial Infarction | Abnormal weight gain events | 18.5 |
| Optum EHR | CUIMC | New users of Tumor Necrosis Factor alpha (TNFa) inhibitors nested in Rheumatoid arthritis | Coronary Vessel Revascularization | 8.4 |
| Optum EHR | CUIMC | New users of SGLT2 inhibitor nested in Left Heart Failure | Rhabdomyolysis | 8.2 |
| Optum EHR | CUIMC | New users of Tumor Necrosis Factor alpha (TNFa) inhibitors nested in Rheumatoid arthritis | All events Drug Rash with Eosinophilia and Systemic Symptoms (DRESS) | 0.0 |
| Optum EHR | CUIMC | New users of Tumor Necrosis Factor alpha (TNFa) inhibitors nested in Rheumatoid arthritis | Polymorphic Ventricular Tachycardia or Torsades de Pointes | 0.0 |
| Optum EHR | CUIMC | New users of Trimethoprim systemetic nested in Urinary Tract Infection | Polymorphic Ventricular Tachycardia or Torsades de Pointes | 0.0 |

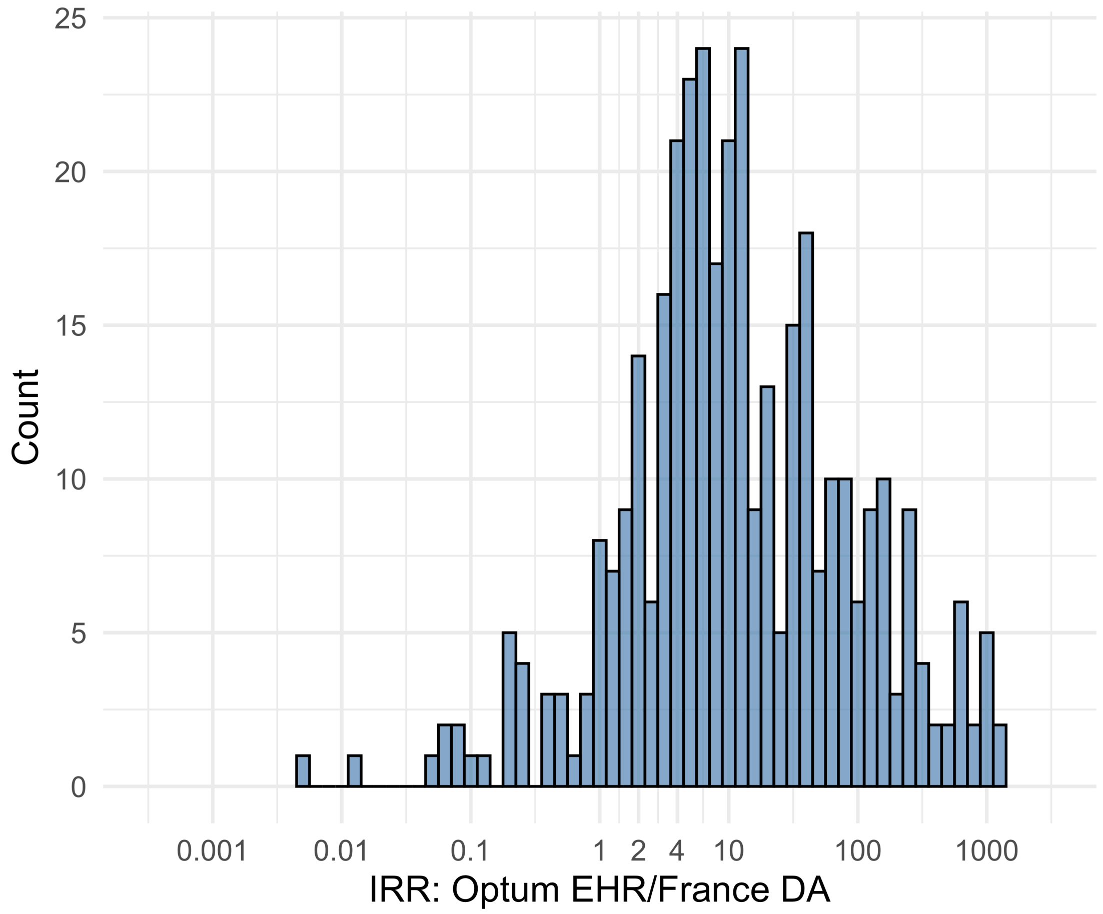


Figure S5.123. IRR distribution comparing Optum EHR and France DA.

*Table S5.123. Most extreme IRRs for Optum EHR vs France DA (rounded to 0.001).*

| source1 | source2 | Drug and Indication | Outcome | IRR |
|---|---|---|---|---|
| Optum EHR | France DA | New users of Beta blockers nested in essential hypertension | Hospitalization with heart failure events | 1257.8 |
| Optum EHR | France DA | New users of Beta blockers nested in essential hypertension | Acute Kidney Injury AKI | 1134.0 |
| Optum EHR | France DA | New users of Beta blockers nested in essential hypertension | 4-point MACE | 1072.5 |
| Optum EHR | France DA | New users of Trimethoprim systemetic nested in Urinary Tract Infection | Polymorphic Ventricular Tachycardia or Torsades de Pointes | 0.0 |
| Optum EHR | France DA | New users of GLP-1 receptor antagonists nested in Type 2 diabetes mellitus | Progressive multifocal leukoencephalopathy | 0.0 |
| Optum EHR | France DA | New users of Trimethoprim systemetic nested in Urinary Tract Infection | All events Drug Rash with Eosinophilia and Systemic Symptoms (DRESS) | 0.1 |

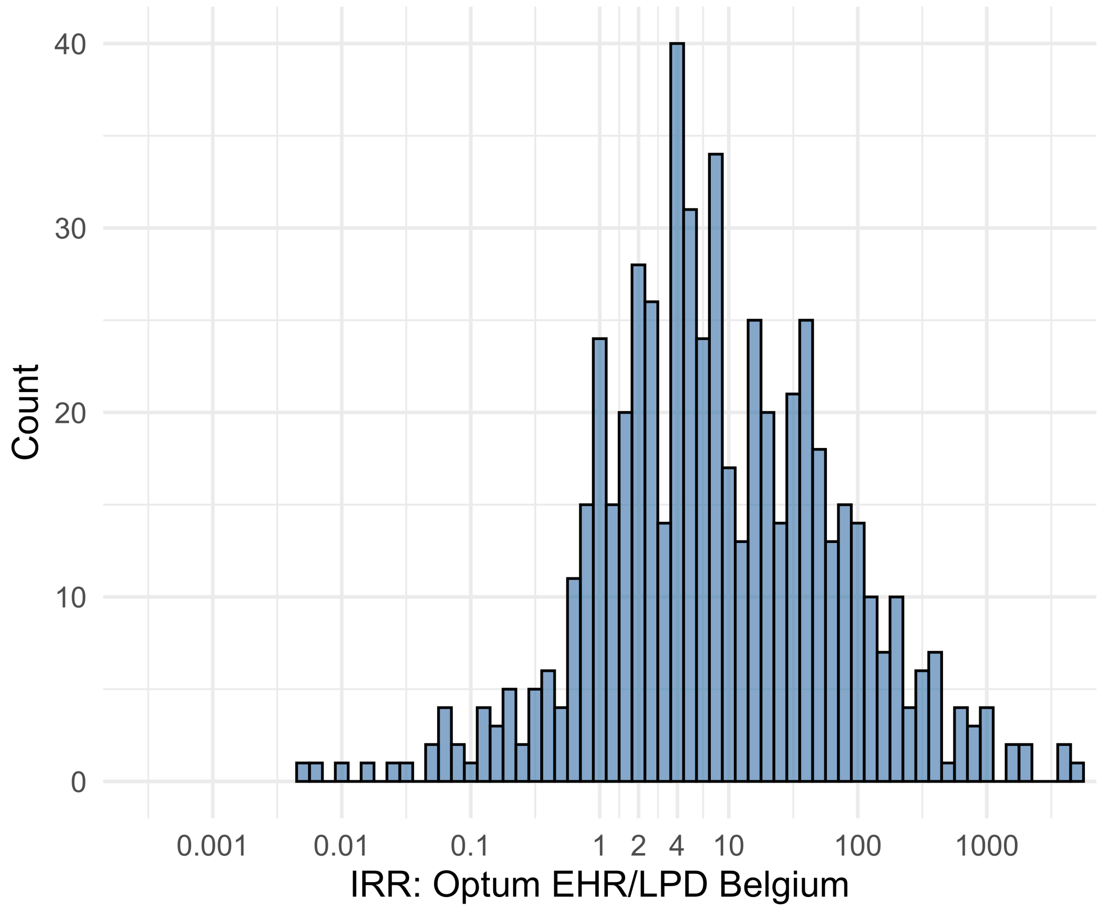


Figure S5.124. IRR distribution comparing Optum EHR and LPD Belgium.

*Table S5.124. Most extreme IRRs for Optum EHR vs LPD Belgium (rounded to 0.001).*

| source1 | source2 | Drug and Indication | Outcome | IRR |
|---|---|---|---|---|
| Optum EHR | LPD Belgium | New users of Beta blockers nested in essential hypertension | Hospitalization with heart failure events | 4895.2 |
| Optum EHR | LPD Belgium | New users of Beta blockers nested in essential hypertension | 4-point MACE | 4174.1 |
| Optum EHR | LPD Belgium | New users of Beta blockers nested in essential hypertension | Total cardiovascular disease events (ischemic stroke, hemorrhagic stroke, heart failure, acute myocardial infarction or sudden cardiac death) | 4142.2 |
| Optum EHR | LPD Belgium | New users of GLP-1 receptor antagonists nested in obesity | Progressive multifocal leukoencephalopathy | 0.0 |
| Optum EHR | LPD Belgium | New users of Trimethoprim systemetic nested in Urinary Tract Infection | Polymorphic Ventricular Tachycardia or Torsades de Pointes | 0.0 |
| Optum EHR | LPD Belgium | New users of GLP-1 receptor antagonists nested in Type 2 diabetes mellitus | Progressive multifocal leukoencephalopathy | 0.0 |

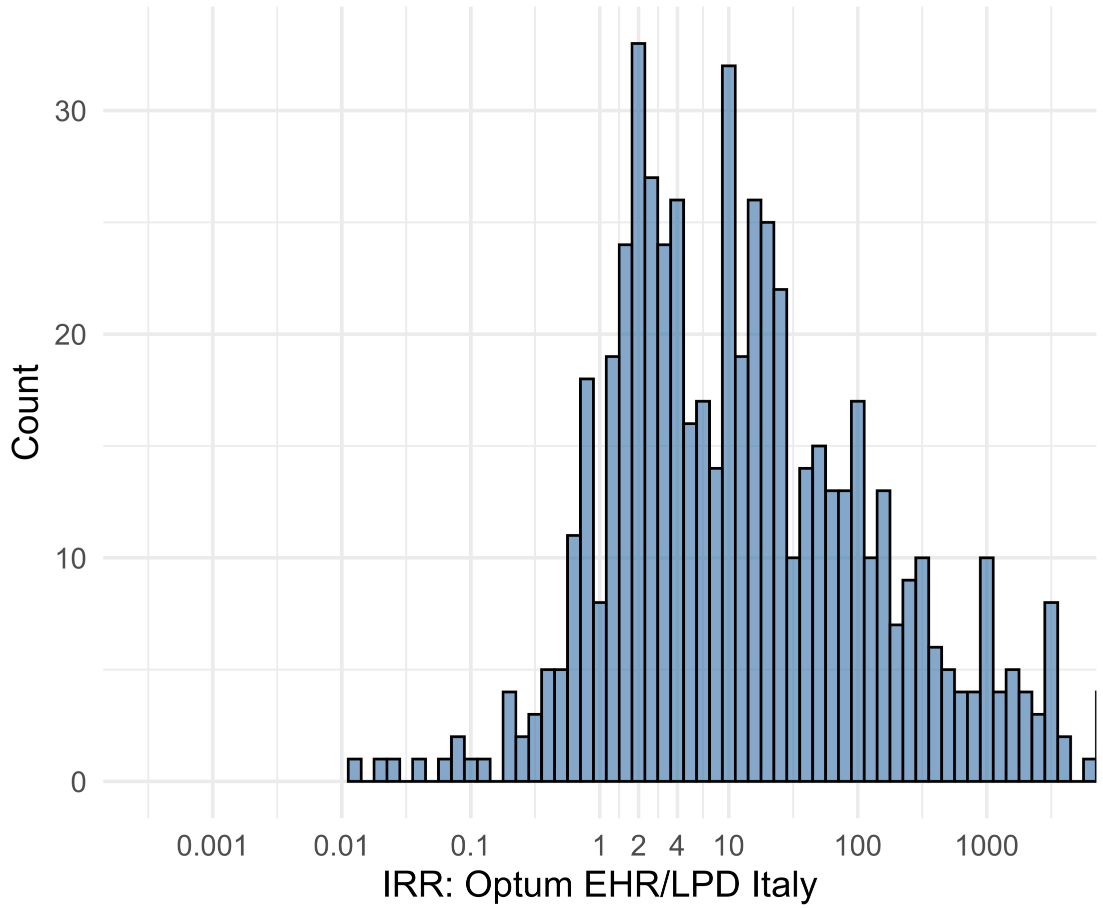


Figure S5.125. IRR distribution comparing Optum EHR and LPD Italy.

*Table S5.125. Most extreme IRRs for Optum EHR vs LPD Italy (rounded to 0.001).*

| source1 | source2 | Drug and Indication | Outcome | IRR |
|---|---|---|---|---|
| Optum EHR | LPD Italy | New users of Beta blockers nested in essential hypertension | Hospitalization with heart failure events | 15915.5 |
| Optum EHR | LPD Italy | New users of Beta blockers nested in essential hypertension | 4-point MACE | 13571.0 |
| Optum EHR | LPD Italy | New users of Beta blockers nested in essential hypertension | Total cardiovascular disease events (ischemic stroke, hemorrhagic stroke, heart failure, acute myocardial infarction or sudden cardiac death) | 13467.3 |
| Optum EHR | LPD Italy | New users of GLP-1 receptor antagonists nested in obesity | Progressive multifocal leukoencephalopathy | 0.0 |
| Optum EHR | LPD Italy | New users of GLP-1 receptor antagonists nested in Type 2 diabetes mellitus | Progressive multifocal leukoencephalopathy | 0.0 |
| Optum EHR | LPD Italy | New users of SGLT2 inhibitor nested in Type 2 diabetes mellitus | Progressive multifocal leukoencephalopathy | 0.0 |

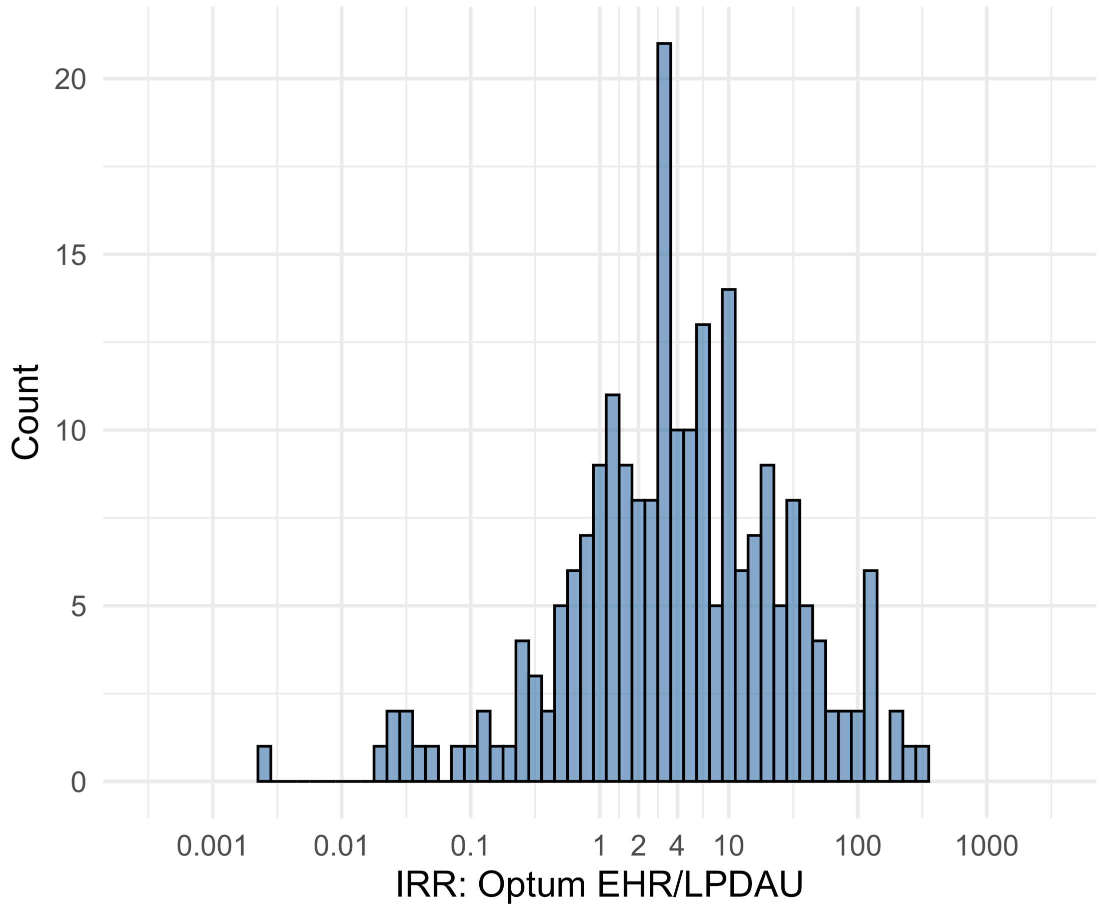


Figure S5.126. IRR distribution comparing Optum EHR and LPDAU.

*Table S5.126. Most extreme IRRs for Optum EHR vs LPDAU (rounded to 0.001).*

| source1 | source2 | Drug and Indication | Outcome | IRR |
|---|---|---|---|---|
| Optum EHR | LPDAU | New users of Cephalosporin systemetic nested in Urinary Tract Infection | Acute Kidney Injury AKI | 287.0 |
| Optum EHR | LPDAU | New users of Cephalosporin systemetic nested in Urinary Tract Infection | Hospitalization with heart failure events | 258.9 |
| Optum EHR | LPDAU | New users of Cephalosporin systemetic nested in Urinary Tract Infection | 4-point MACE | 186.8 |
| Optum EHR | LPDAU | New users of Trimethoprim systemetic nested in Urinary Tract Infection | Polymorphic Ventricular Tachycardia or Torsades de Pointes | 0.0 |
| Optum EHR | LPDAU | New users of Beta blockers nested in essential hypertension | Progressive multifocal leukoencephalopathy | 0.0 |
| Optum EHR | LPDAU | New users of Beta blockers nested in essential hypertension | All events Drug Rash with Eosinophilia and Systemic Symptoms (DRESS) | 0.0 |

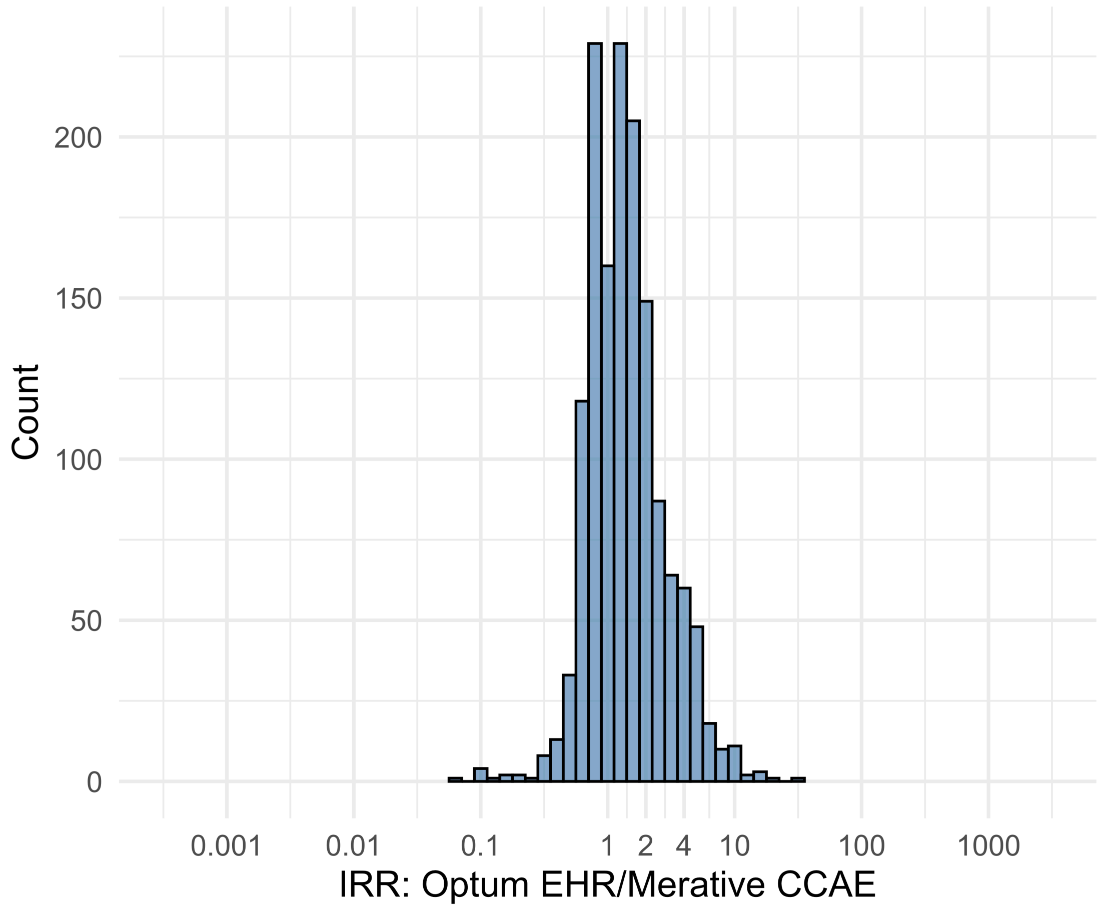


Figure S5.127. IRR distribution comparing Optum EHR and Merative CCAE.

*Table S5.127. Most extreme IRRs for Optum EHR vs Merative CCAE (rounded to 0.001).*

| source1 | source2 | Drug and Indication | Outcome | IRR |
|---|---|---|---|---|
| Optum EHR | Merative CCAE | New users of Beta blockers nested in Acute Myocardial Infarction | Thrombosis with Thrombocytopenia (TWT) | 29.6 |
| Optum EHR | Merative CCAE | New users of Cephalosporin systemetic nested in Urinary Tract Infection | Thrombosis with Thrombocytopenia (TWT) | 20.0 |
| Optum EHR | Merative CCAE | New users of Fluoroquinolone systemic nested in Urinary Tract Infection | Thrombosis with Thrombocytopenia (TWT) | 17.0 |
| Optum EHR | Merative CCAE | New users of Tumor Necrosis Factor alpha (TNFa) inhibitors nested in Rheumatoid arthritis | Earliest event of Thrombotic microangiopathy (TMA) or Microangiopathic hemolytic anemia (MAHA) | 0.1 |
| Optum EHR | Merative CCAE | New users of Tumor Necrosis Factor alpha (TNFa) inhibitors nested in Rheumatoid arthritis | Stevens-Johnson syndrome, toxic epidermal necrolysis spectrum | 0.1 |
| Optum EHR | Merative CCAE | New users of Tumor Necrosis Factor alpha (TNFa) inhibitors nested in Ulcerative colitis | Earliest event of Thrombotic microangiopathy (TMA) or Microangiopathic hemolytic anemia (MAHA) | 0.1 |

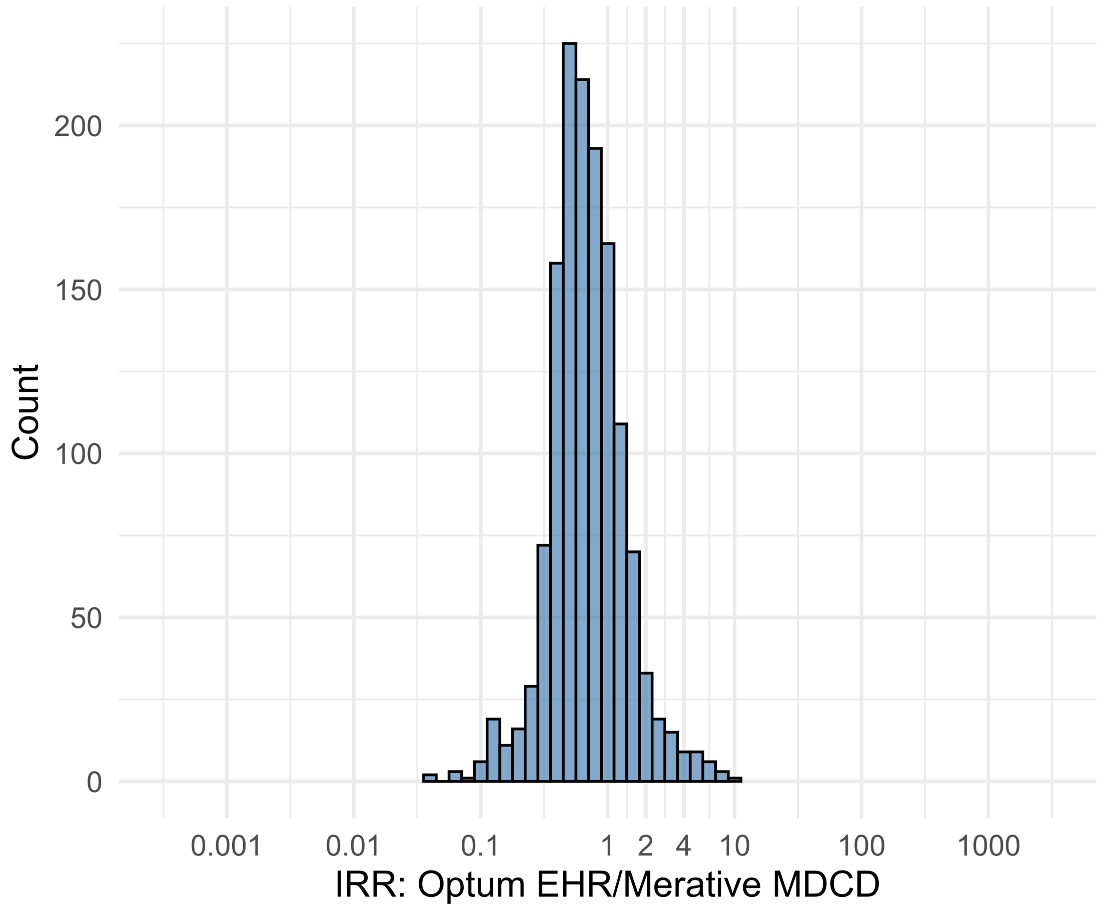


Figure S5.128. IRR distribution comparing Optum EHR and Merative MDCD.

*Table S5.128. Most extreme IRRs for Optum EHR vs Merative MDCD (rounded to 0.001).*

| source1 | source2 | Drug and Indication | Outcome | IRR |
|---|---|---|---|---|
| Optum EHR | Merative MDCD | New users of Tumor Necrosis Factor alpha (TNFa) inhibitors nested in Ulcerative colitis | Persons with gout | 10.4 |
| Optum EHR | Merative MDCD | New users of Trimethoprim systemetic nested in Urinary Tract Infection | Thrombosis with Thrombocytopenia (TWT) | 7.8 |
| Optum EHR | Merative MDCD | New users of Trimethoprim systemetic nested in Acute Typical Pneumonia | All events of Autoimmune hepatitis, with a washout period of 365 days | 7.7 |
| Optum EHR | Merative MDCD | New users of JAK inhibitors nested in Rheumatoid arthritis | Guillain Barre Syndrome | 0.0 |
| Optum EHR | Merative MDCD | New users of JAK inhibitors nested in Rheumatoid arthritis | Stevens-Johnson syndrome, toxic epidermal necrolysis spectrum | 0.0 |
| Optum EHR | Merative MDCD | New users of Tumor Necrosis Factor alpha (TNFa) inhibitors nested in Rheumatoid arthritis | All events Drug Rash with Eosinophilia and Systemic Symptoms (DRESS) | 0.1 |

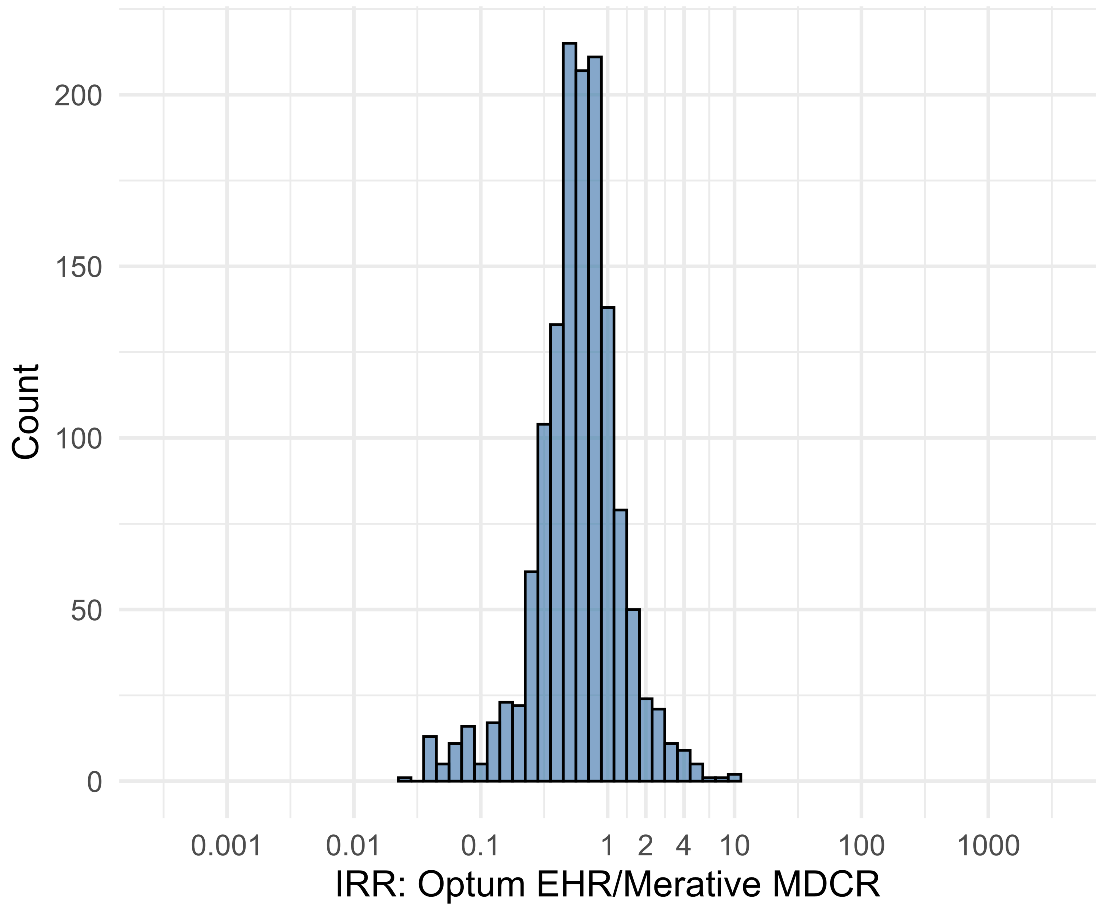


Figure S5.129. IRR distribution comparing Optum EHR and Merative MDCR.

*Table S5.129. Most extreme IRRs for Optum EHR vs Merative MDCR (rounded to 0.001).*

| source1 | source2 | Drug and Indication | Outcome | IRR |
|---|---|---|---|---|
| Optum EHR | Merative MDCR | New users of Beta blockers nested in Acute Myocardial Infarction | Thrombosis with Thrombocytopenia (TWT) | 10.1 |
| Optum EHR | Merative MDCR | New users of Beta blockers nested in Left Heart Failure | Polymorphic Ventricular Tachycardia or Torsades de Pointes | 10.0 |
| Optum EHR | Merative MDCR | New users of Tumor Necrosis Factor alpha (TNFa) inhibitors nested in Rheumatoid arthritis | Myocarditis Pericarditis | 7.3 |
| Optum EHR | Merative MDCR | New users of Tumor Necrosis Factor alpha (TNFa) inhibitors nested in Crohns disease | Polymorphic Ventricular Tachycardia or Torsades de Pointes | 0.0 |
| Optum EHR | Merative MDCR | New users of Tumor Necrosis Factor alpha (TNFa) inhibitors nested in Ulcerative colitis | Guillain Barre Syndrome | 0.0 |
| Optum EHR | Merative MDCR | New users of Tumor Necrosis Factor alpha (TNFa) inhibitors nested in Ulcerative colitis | Progressive multifocal leukoencephalopathy | 0.0 |

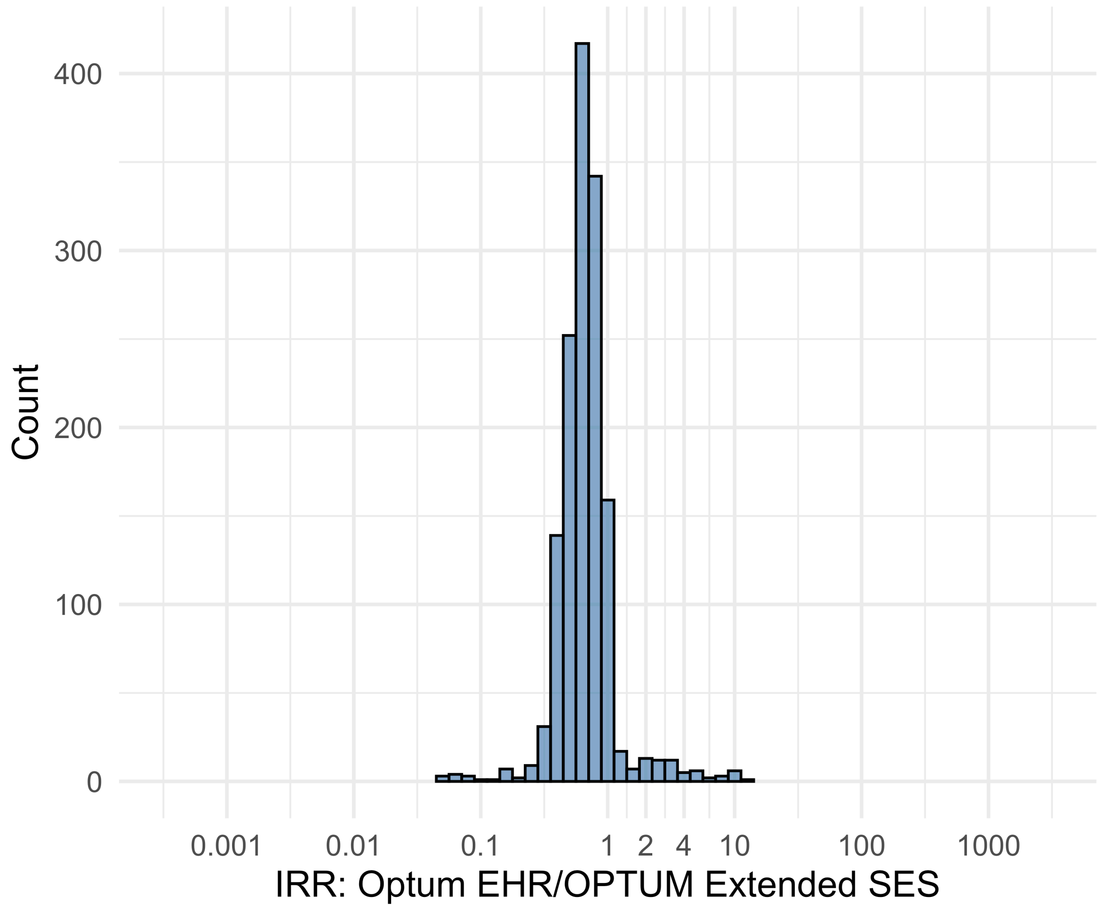


Figure S5.130. IRR distribution comparing Optum EHR and OPTUM Extended SES.

*Table S5.130. Most extreme IRRs for Optum EHR vs OPTUM Extended SES (rounded to 0.001).*

| source1 | source2 | Drug and Indication | Outcome | IRR |
|---|---|---|---|---|
| Optum EHR | OPTUM Extended SES | New users of Tumor Necrosis Factor alpha (TNFa) inhibitors nested in Rheumatoid arthritis | Cardiovascular-related mortality | 13.9 |
| Optum EHR | OPTUM Extended SES | New users of Fluoroquinolone systemic nested in Acute Typical Pneumonia | Cardiovascular-related mortality | 11.0 |
| Optum EHR | OPTUM Extended SES | New users of Cephalosporin systemetic nested in Acute Typical Pneumonia | Cardiovascular-related mortality | 11.0 |
| Optum EHR | OPTUM Extended SES | New users of Tumor Necrosis Factor alpha (TNFa) inhibitors nested in Rheumatoid arthritis | Stevens-Johnson syndrome, toxic epidermal necrolysis spectrum | 0.0 |
| Optum EHR | OPTUM Extended SES | New users of Tumor Necrosis Factor alpha (TNFa) inhibitors nested in Crohns disease | Acquired Pure Red Cell Aplasia | 0.1 |
| Optum EHR | OPTUM Extended SES | New users of Tumor Necrosis Factor alpha (TNFa) inhibitors nested in Psoriatic Arthritis | Rhabdomyolysis | 0.1 |

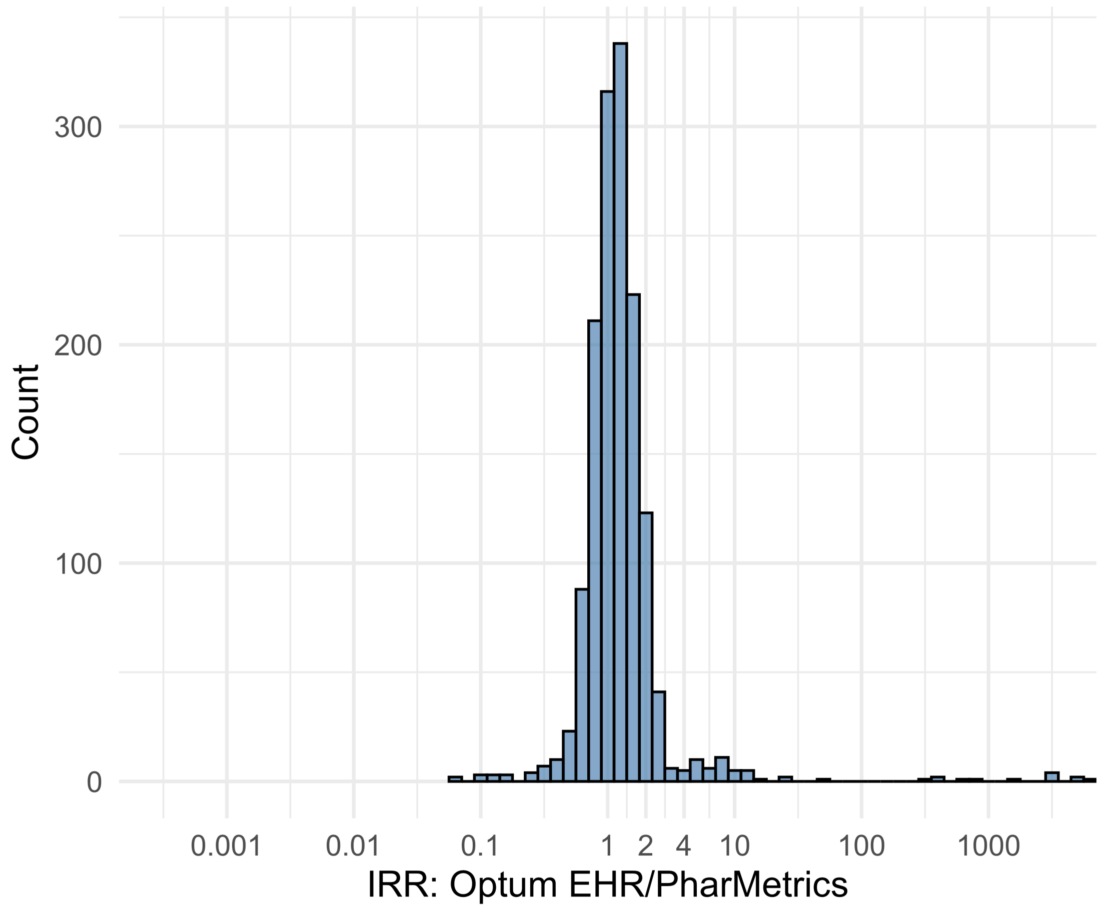


Figure S5.131. IRR distribution comparing Optum EHR and PharMetrics.

*Table S5.131. Most extreme IRRs for Optum EHR vs PharMetrics (rounded to 0.001).*

| source1 | source2 | Drug and Indication | Outcome | IRR |
|---|---|---|---|---|
| Optum EHR | PharMetrics | New users of Beta blockers nested in essential hypertension | Cardiovascular-related mortality | 6562.6 |
| Optum EHR | PharMetrics | New users of Cephalosporin systemetic nested in Acute Typical Pneumonia | Cardiovascular-related mortality | 5094.5 |
| Optum EHR | PharMetrics | New users of Cephalosporin systemetic nested in Urinary Tract Infection | Cardiovascular-related mortality | 4977.7 |
| Optum EHR | PharMetrics | New users of JAK inhibitors nested in Rheumatoid arthritis | Guillain Barre Syndrome | 0.1 |
| Optum EHR | PharMetrics | New users of Beta blockers nested in Left Heart Failure | Progressive multifocal leukoencephalopathy | 0.1 |
| Optum EHR | PharMetrics | New users of Tumor Necrosis Factor alpha (TNFa) inhibitors nested in Crohns disease | Stevens-Johnson syndrome, toxic epidermal necrolysis spectrum | 0.1 |

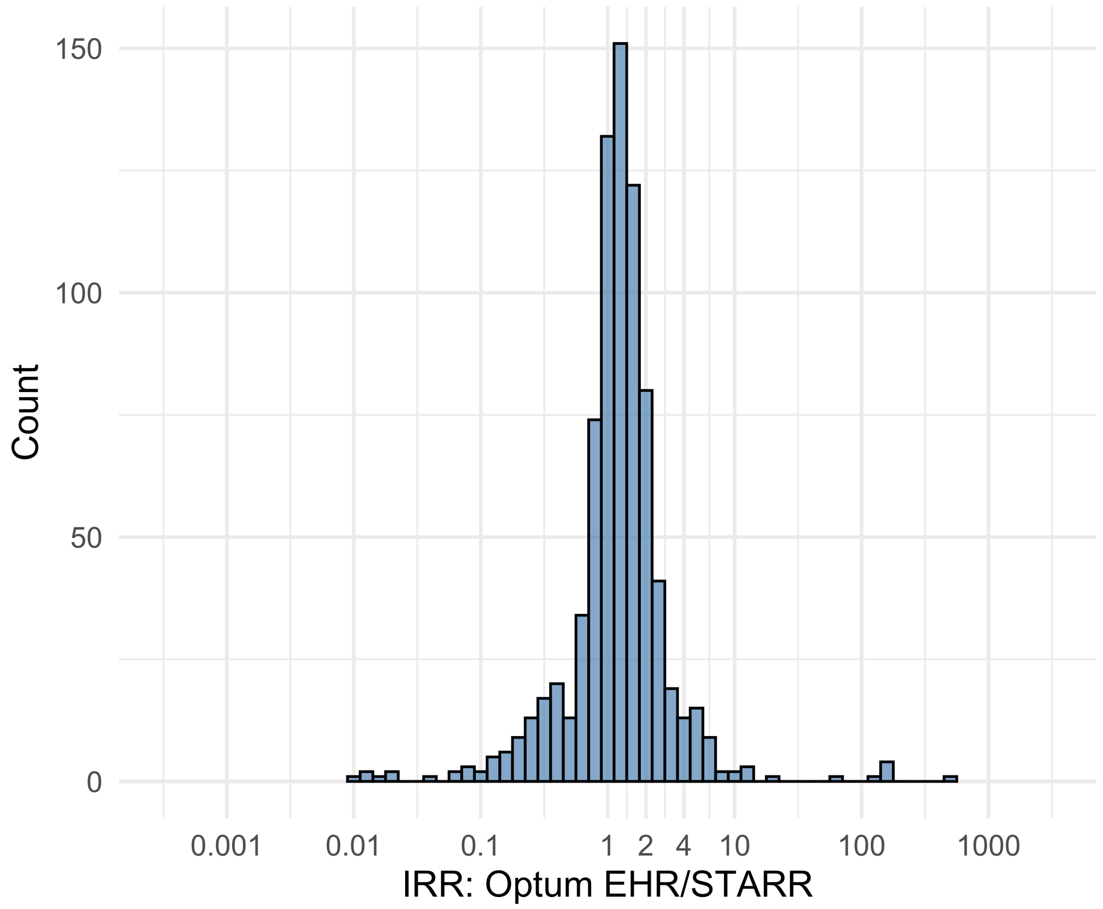


Figure S5.132. IRR distribution comparing Optum EHR and STARR.

*Table S5.132. Most extreme IRRs for Optum EHR vs STARR (rounded to 0.001).*

| source1 | source2 | Drug and Indication | Outcome | IRR |
|---|---|---|---|---|
| Optum EHR | STARR | New users of Beta blockers nested in essential hypertension | Febrile Neutropenia or Neutropenic Fever | 538.0 |
| Optum EHR | STARR | New users of Beta blockers nested in Left Heart Failure | Febrile Neutropenia or Neutropenic Fever | 163.1 |
| Optum EHR | STARR | New users of Fluoroquinolone systemic nested in Urinary Tract Infection | Febrile Neutropenia or Neutropenic Fever | 151.2 |
| Optum EHR | STARR | New users of SGLT2 inhibitor nested in Type 2 diabetes mellitus | Progressive multifocal leukoencephalopathy | 0.0 |
| Optum EHR | STARR | New users of GLP-1 receptor antagonists nested in obesity | Progressive multifocal leukoencephalopathy | 0.0 |
| Optum EHR | STARR | New users of GLP-1 receptor antagonists nested in Type 2 diabetes mellitus | Progressive multifocal leukoencephalopathy | 0.0 |

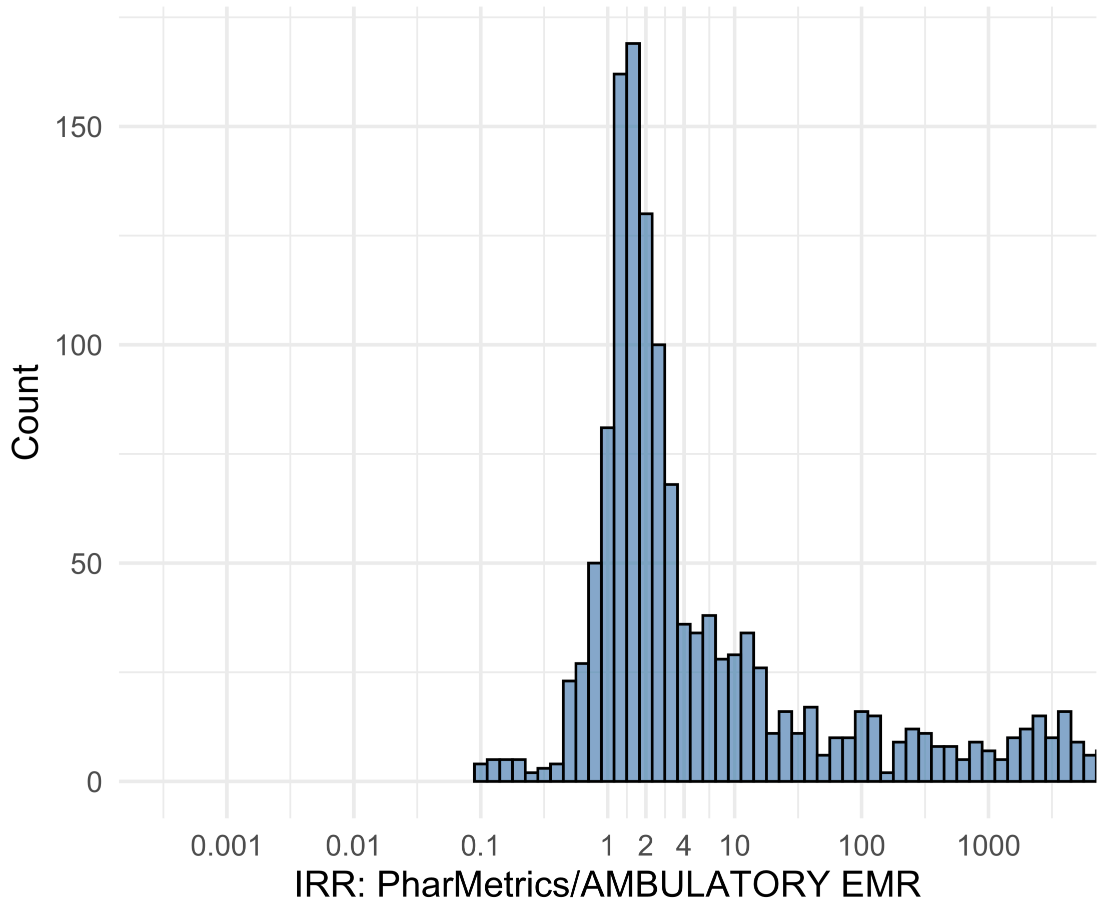


Figure S5.133. IRR distribution comparing PharMetrics and AMBULATORY EMR.

*Table S5.133. Most extreme IRRs for PharMetrics vs AMBULATORY EMR (rounded to 0.001).*

| source1 | source2 | Drug and Indication | Outcome | IRR |
|---|---|---|---|---|
| PharMetrics | AMBULATORY EMR | New users of Beta blockers nested in essential hypertension | Hospitalization with heart failure events | 206293.3 |
| PharMetrics | AMBULATORY EMR | New users of Beta blockers nested in essential hypertension | 4-point MACE | 163601.3 |
| PharMetrics | AMBULATORY EMR | New users of Beta blockers nested in essential hypertension | Total cardiovascular disease events (ischemic stroke, hemorrhagic stroke, heart failure, acute myocardial infarction or sudden cardiac death) | 161143.8 |
| PharMetrics | AMBULATORY EMR | New users of Trimethoprim systemetic nested in Acute Typical Pneumonia | Anaphylaxis | 0.1 |
| PharMetrics | AMBULATORY EMR | New users of SGLT2 inhibitor nested in Type 2 diabetes mellitus | Anaphylaxis | 0.1 |

| source1 | source2 | Drug and Indication | Outcome | IRR |
|---|---|---|---|---|
| PharMetrics | AMBULATORY EMR | New users of SGLT2 inhibitor nested in Left Heart Failure | Anaphylaxis | 0.1 |

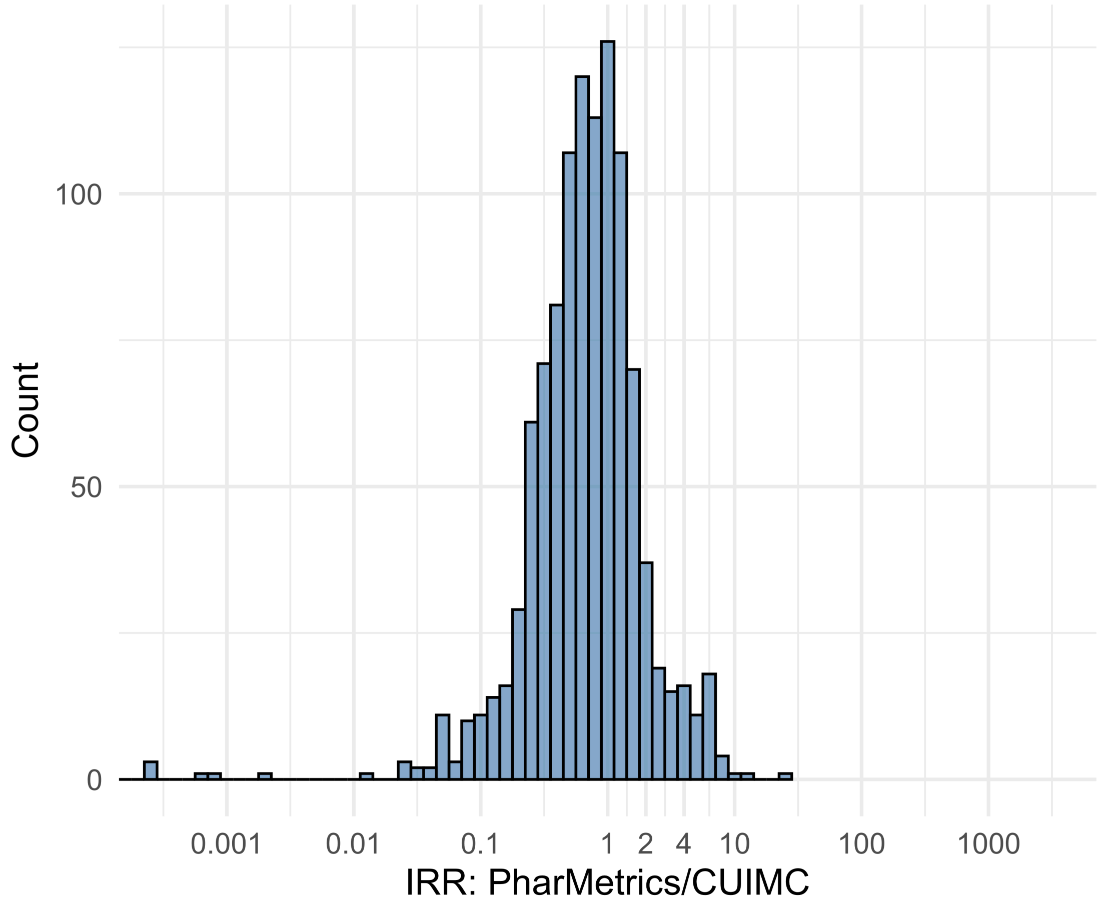


Figure S5.134. IRR distribution comparing PharMetrics and CUIMC.

*Table S5.134. Most extreme IRRs for PharMetrics vs CUIMC (rounded to 0.001).*

| source1 | source2 | Drug and Indication | Outcome | IRR |
|---|---|---|---|---|
| PharMetrics | CUIMC | New users of Beta blockers nested in Acute Myocardial Infarction | Abnormal weight gain events | 23.2 |
| PharMetrics | CUIMC | New users of SGLT2 inhibitor nested in Left Heart Failure | Rhabdomyolysis | 11.6 |
| PharMetrics | CUIMC | New users of Cephalosporin systemetic nested in Acute Typical Pneumonia | Abnormal weight gain events | 10.8 |
| PharMetrics | CUIMC | New users of Cephalosporin systemetic nested in Acute Typical Pneumonia | Cardiovascular-related mortality | <0.001 |
| PharMetrics | CUIMC | New users of Beta blockers nested in essential hypertension | Cardiovascular-related mortality | <0.001 |
| PharMetrics | CUIMC | New users of Fluoroquinolone systemic nested in Acute Typical Pneumonia | Cardiovascular-related mortality | <0.001 |

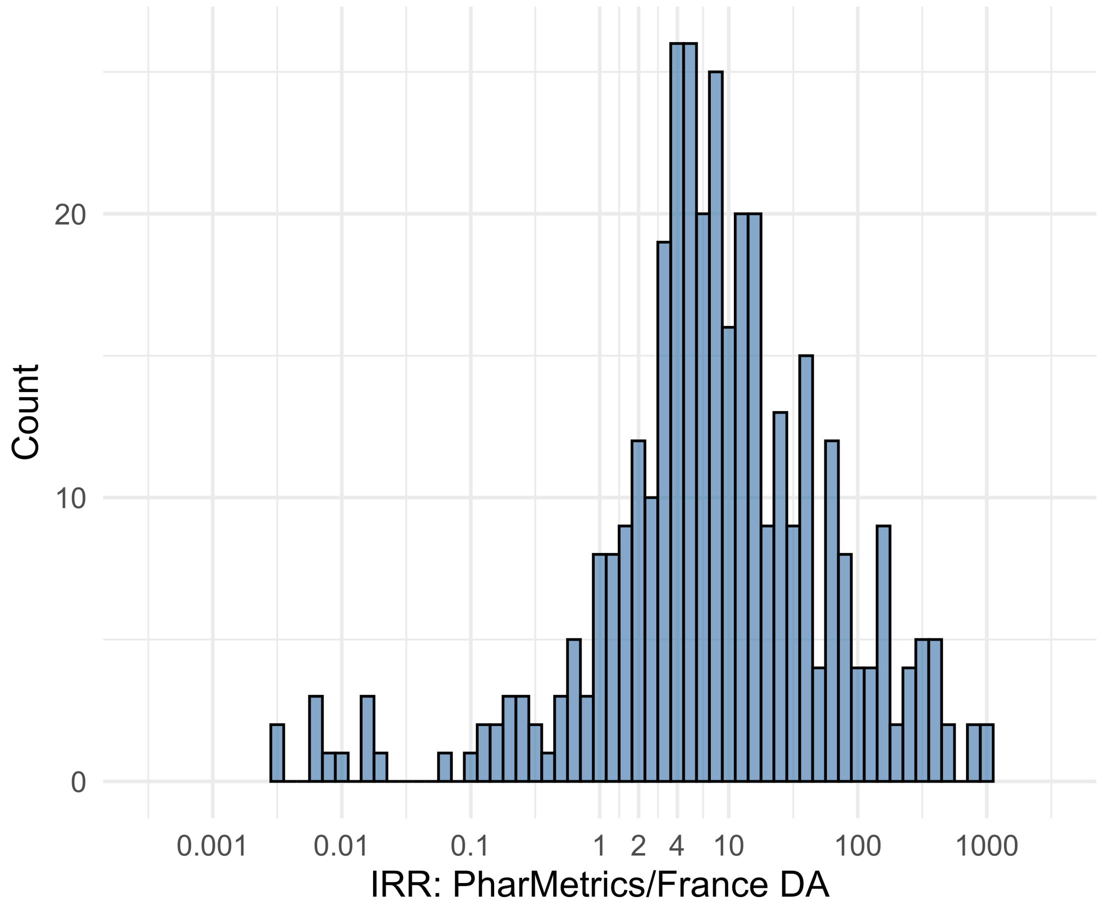


Figure S5.135. IRR distribution comparing PharMetrics and France DA.

*Table S5.135. Most extreme IRRs for PharMetrics vs France DA (rounded to 0.001).*

| source1 | source2 | Drug and Indication | Outcome | IRR |
|---|---|---|---|---|
| PharMetrics | France DA | New users of Beta blockers nested in essential hypertension | Hospitalization with heart failure events | 1013.4 |
| PharMetrics | France DA | New users of Beta blockers nested in essential hypertension | Acute Kidney Injury AKI | 930.4 |
| PharMetrics | France DA | New users of Beta blockers nested in essential hypertension | 4-point MACE | 803.7 |
| PharMetrics | France DA | New users of Trimethoprim systemetic nested in Urinary Tract Infection | Cardiovascular-related mortality | 0.0 |
| PharMetrics | France DA | New users of Trimethoprim systemetic nested in Urinary Tract Infection | Polymorphic Ventricular Tachycardia or Torsades de Pointes | 0.0 |
| PharMetrics | France DA | New users of Beta blockers nested in essential hypertension | Cardiovascular-related mortality | 0.0 |

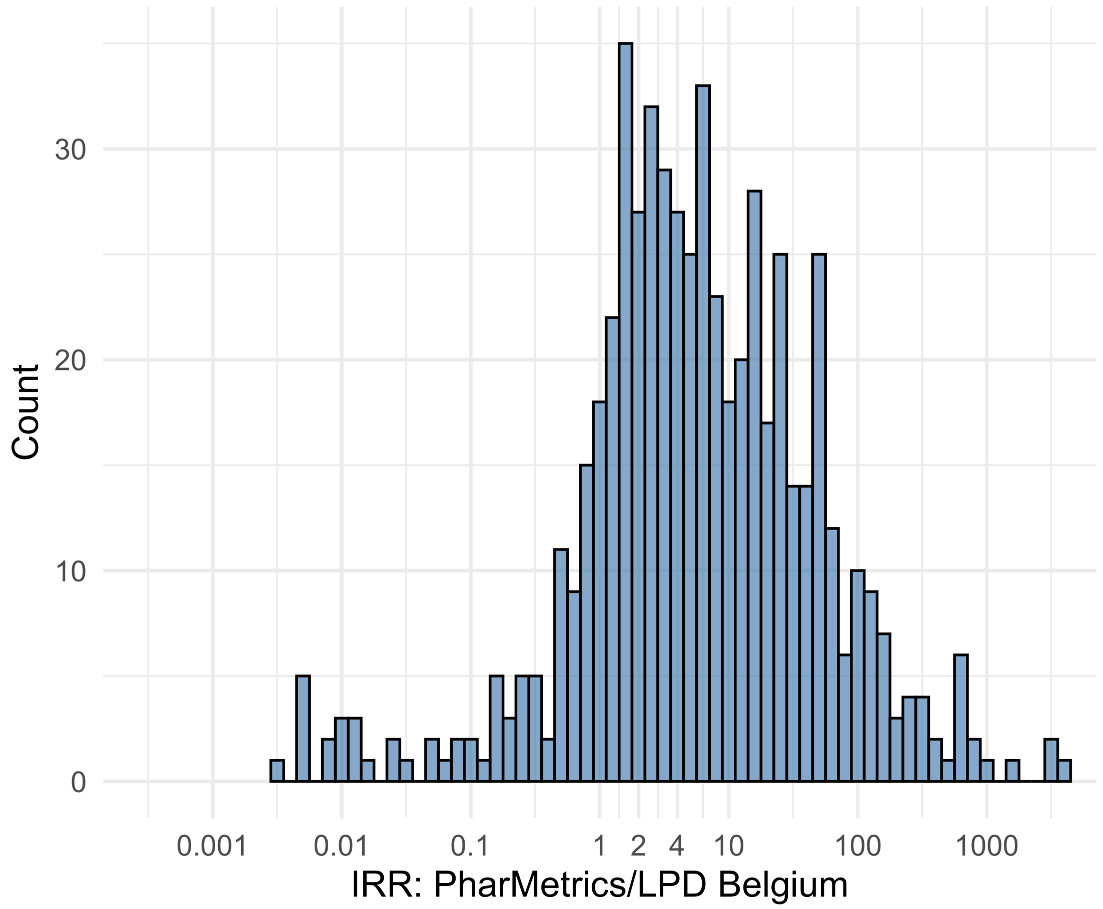


Figure S5.136. IRR distribution comparing PharMetrics and LPD Belgium.

*Table S5.136. Most extreme IRRs for PharMetrics vs LPD Belgium (rounded to 0.001).*

| source1 | source2 | Drug and Indication | Outcome | IRR |
|---|---|---|---|---|
| PharMetrics | LPD Belgium | New users of Beta blockers nested in essential hypertension | Hospitalization with heart failure events | 3943.9 |
| PharMetrics | LPD Belgium | New users of Beta blockers nested in essential hypertension | 4-point MACE | 3127.8 |
| PharMetrics | LPD Belgium | New users of Beta blockers nested in essential hypertension | Total cardiovascular disease events (ischemic stroke, hemorrhagic stroke, heart failure, acute myocardial infarction or sudden cardiac death) | 3080.8 |
| PharMetrics | LPD Belgium | New users of Cephalosporin systemetic nested in Urinary Tract Infection | Cardiovascular-related mortality | 0.0 |
| PharMetrics | LPD Belgium | New users of GLP-1 receptor antagonists nested in obesity | Cardiovascular-related mortality | 0.0 |
| PharMetrics | LPD Belgium | New users of GLP-1 receptor antagonists nested in obesity | Progressive multifocal leukoencephalopathy | 0.0 |

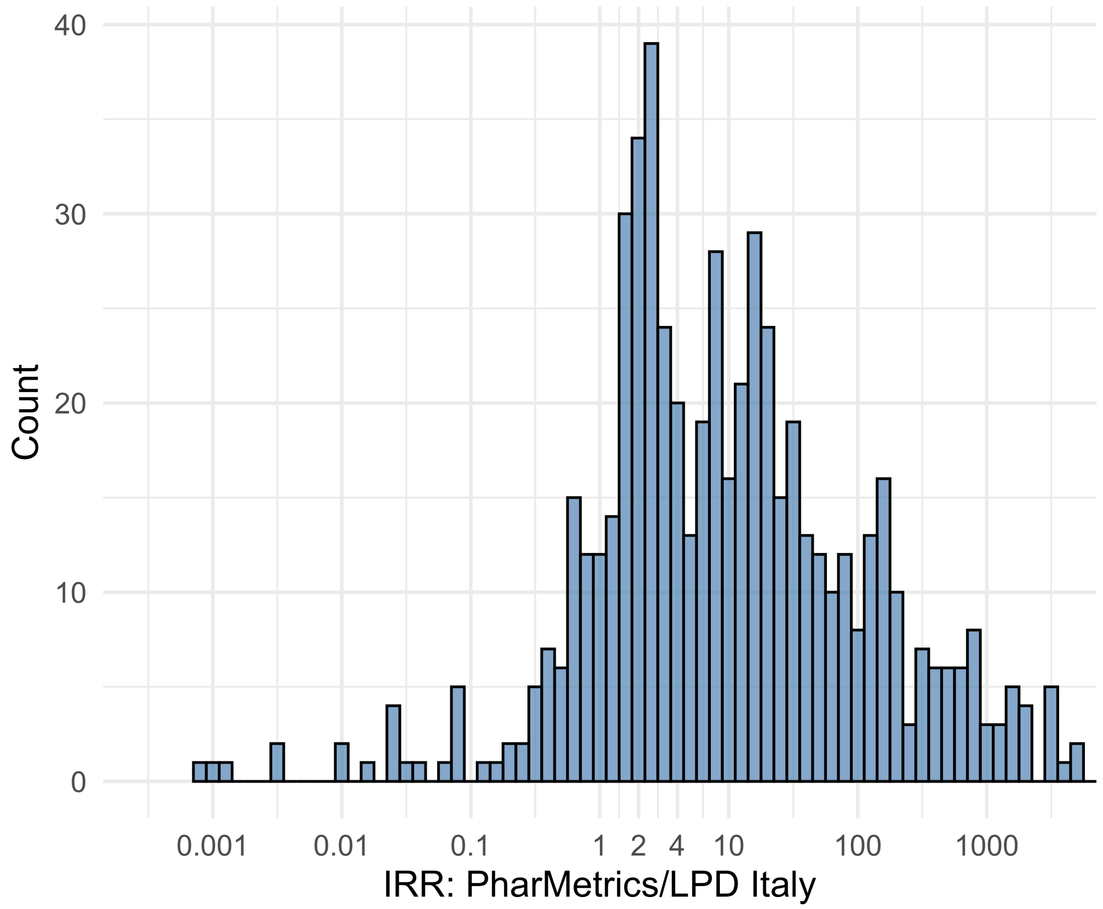


Figure S5.137. IRR distribution comparing PharMetrics and LPD Italy.

*Table S5.137. Most extreme IRRs for PharMetrics vs LPD Italy (rounded to 0.001).*

| source1 | source2 | Drug and Indication | Outcome | IRR |
|---|---|---|---|---|
| PharMetrics | LPD Italy | New users of Beta blockers nested in essential hypertension | Hospitalization with heart failure events | 12822.9 |
| PharMetrics | LPD Italy | New users of Beta blockers nested in essential hypertension | 4-point MACE | 10169.2 |
| PharMetrics | LPD Italy | New users of Beta blockers nested in essential hypertension | Total cardiovascular disease events (ischemic stroke, hemorrhagic stroke, heart failure, acute myocardial infarction or sudden cardiac death) | 10016.4 |
| PharMetrics | LPD Italy | New users of Beta blockers nested in essential hypertension | Cardiovascular-related mortality | 0.0 |
| PharMetrics | LPD Italy | New users of Cephalosporin systemetic nested in Urinary Tract Infection | Cardiovascular-related mortality | 0.0 |
| PharMetrics | LPD Italy | New users of Cephalosporin systemetic nested in Acute Typical Pneumonia | Cardiovascular-related mortality | 0.0 |

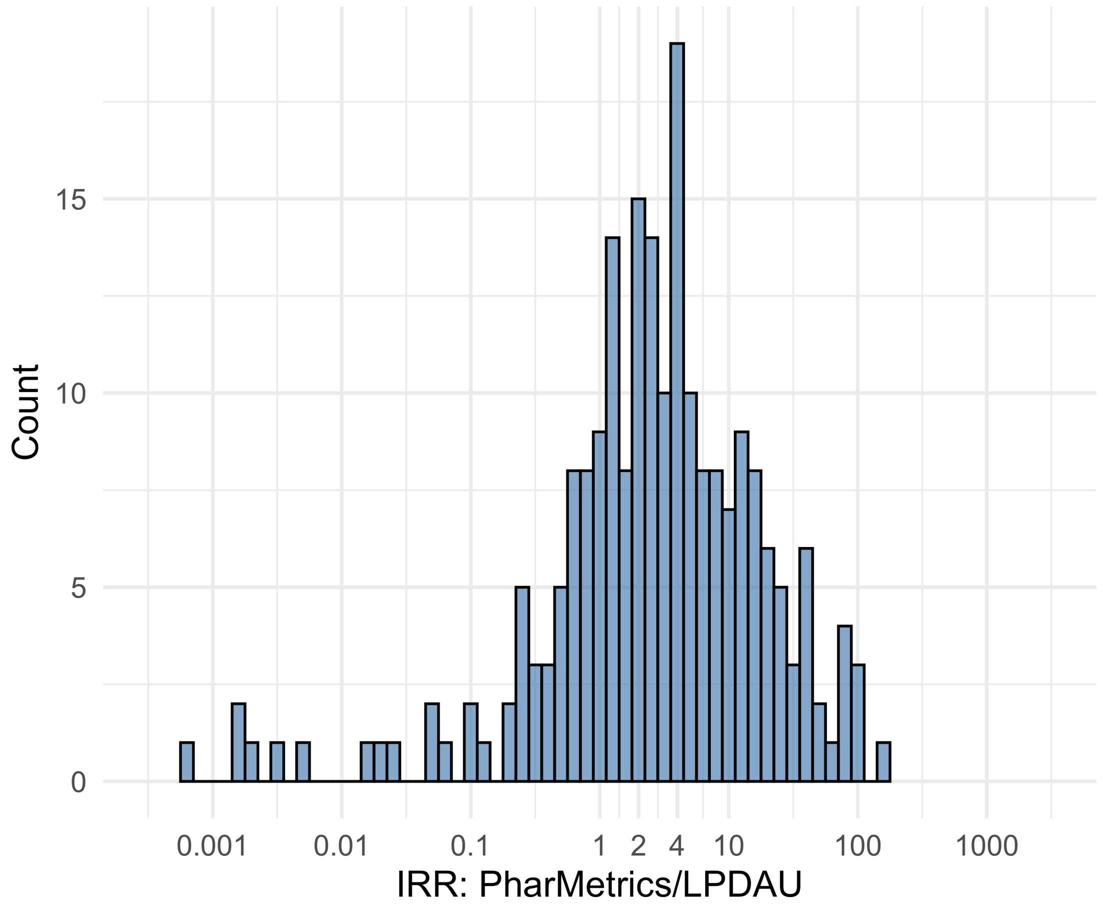


Figure S5.138. IRR distribution comparing PharMetrics and LPDAU.

*Table S5.138. Most extreme IRRs for PharMetrics vs LPDAU (rounded to 0.001).*

| source1 | source2 | Drug and Indication | Outcome | IRR |
|---|---|---|---|---|
| PharMetrics | LPDAU | New users of Cephalosporin systemetic nested in Urinary Tract Infection | Acute Kidney Injury AKI | 151.5 |
| PharMetrics | LPDAU | New users of Beta blockers nested in essential hypertension | Hospitalization with heart failure events | 111.0 |
| PharMetrics | LPDAU | New users of Cephalosporin systemetic nested in Urinary Tract Infection | Hospitalization with heart failure events | 107.8 |
| PharMetrics | LPDAU | New users of Beta blockers nested in essential hypertension | Cardiovascular-related mortality | 0.0 |
| PharMetrics | LPDAU | New users of Trimethoprim systemetic nested in Urinary Tract Infection | Cardiovascular-related mortality | 0.0 |
| PharMetrics | LPDAU | New users of Trimethoprim systemetic nested in Urinary Tract Infection | Polymorphic Ventricular Tachycardia or Torsades de Pointes | 0.0 |

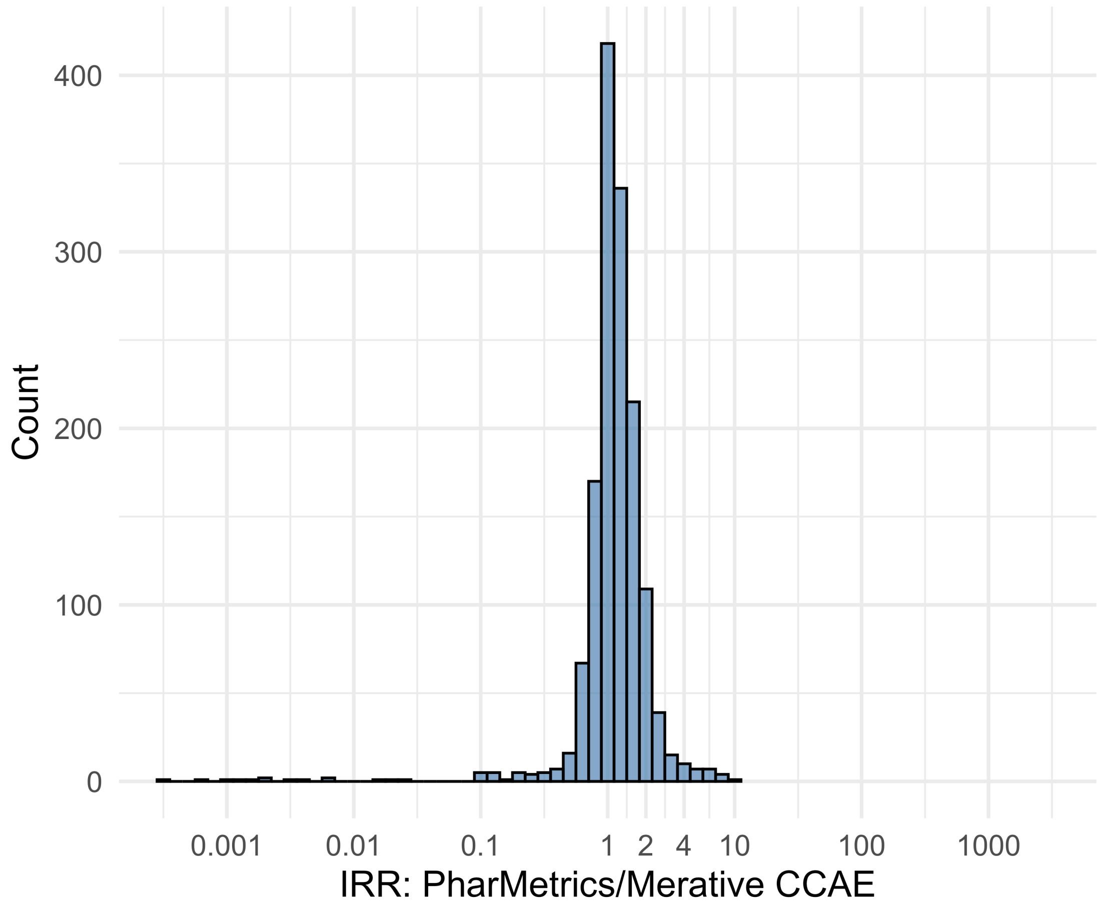


Figure S5.139. IRR distribution comparing PharMetrics and Merative CCAE.

*Table S5.139. Most extreme IRRs for PharMetrics vs Merative CCAE (rounded to 0.001).*

| source1 | source2 | Drug and Indication | Outcome | IRR |
|---|---|---|---|---|
| PharMetrics | Merative CCAE | New users of Tumor Necrosis Factor alpha (TNFa) inhibitors nested in Psoriatic Arthritis | Renal Cancer | 10.9 |
| PharMetrics | Merative CCAE | New users of Tumor Necrosis Factor alpha (TNFa) inhibitors nested in Psoriatic Arthritis | Lower extremity amputation | 8.6 |
| PharMetrics | Merative CCAE | New users of Cephalosporin systemetic nested in Urinary Tract Infection | Progressive multifocal leukoencephalopathy | 8.4 |
| PharMetrics | Merative CCAE | New users of Beta blockers nested in essential hypertension | Cardiovascular-related mortality | <0.001 |
| PharMetrics | Merative CCAE | New users of Beta blockers nested in Left Heart Failure | Cardiovascular-related mortality | 0.0 |
| PharMetrics | Merative CCAE | New users of Fluoroquinolone systemic nested in Acute Typical Pneumonia | Cardiovascular-related mortality | 0.0 |

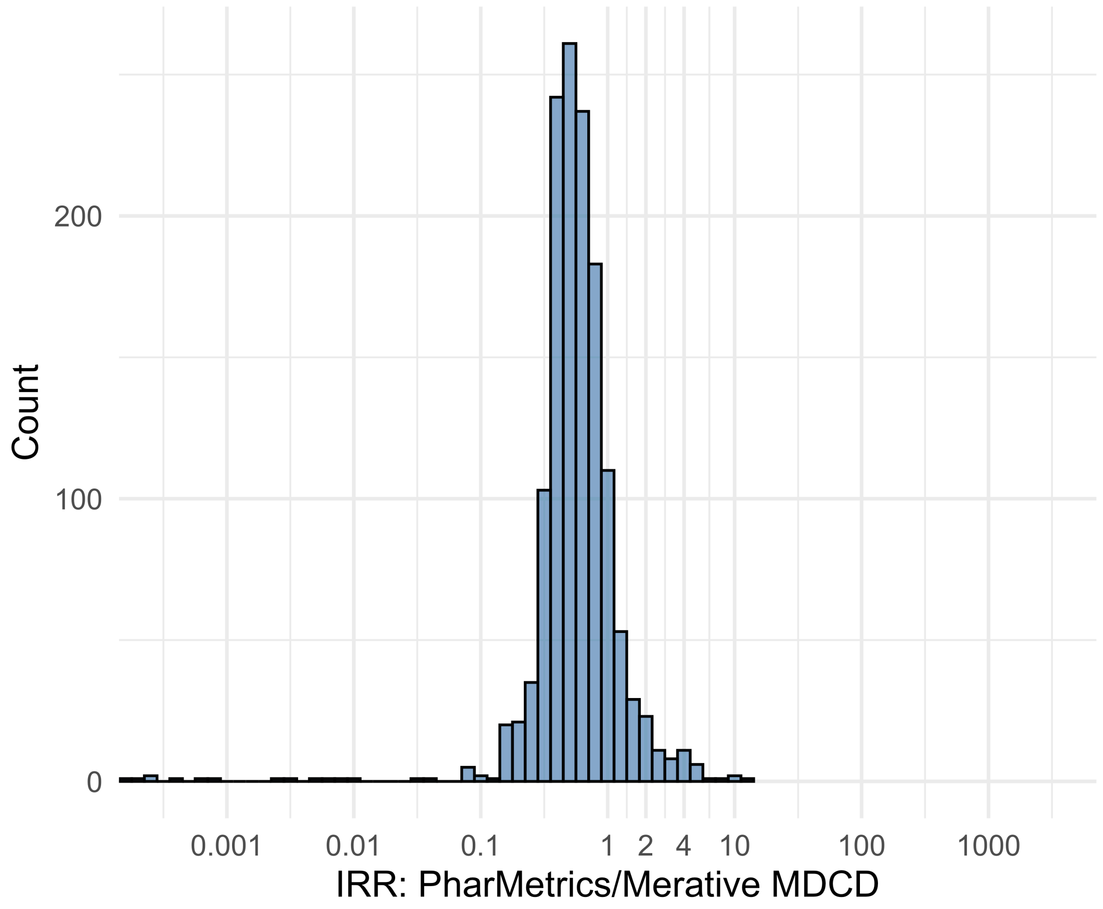


Figure S5.140. IRR distribution comparing PharMetrics and Merative MDCD.

*Table S5.140. Most extreme IRRs for PharMetrics vs Merative MDCD (rounded to 0.001).*

| source1 | source2 | Drug and Indication | Outcome | IRR |
|---|---|---|---|---|
| PharMetrics | Merative MDCD | New users of Tumor Necrosis Factor alpha (TNFa) inhibitors nested in Ulcerative colitis | Persons with gout | 12.4 |
| PharMetrics | Merative MDCD | New users of SGLT2 inhibitor nested in Left Heart Failure | All events of Autoimmune hepatitis, with a washout period of 365 days | 10.7 |
| PharMetrics | Merative MDCD | New users of Trimethoprim systemetic nested in Acute Typical Pneumonia | All events of Autoimmune hepatitis, with a washout period of 365 days | 10.6 |
| PharMetrics | Merative MDCD | New users of Beta blockers nested in essential hypertension | Cardiovascular-related mortality | <0.001 |
| PharMetrics | Merative MDCD | New users of Fluoroquinolone systemic nested in Acute Typical Pneumonia | Cardiovascular-related mortality | <0.001 |
| PharMetrics | Merative MDCD | New users of Cephalosporin systemetic nested in Urinary Tract Infection | Cardiovascular-related mortality | <0.001 |

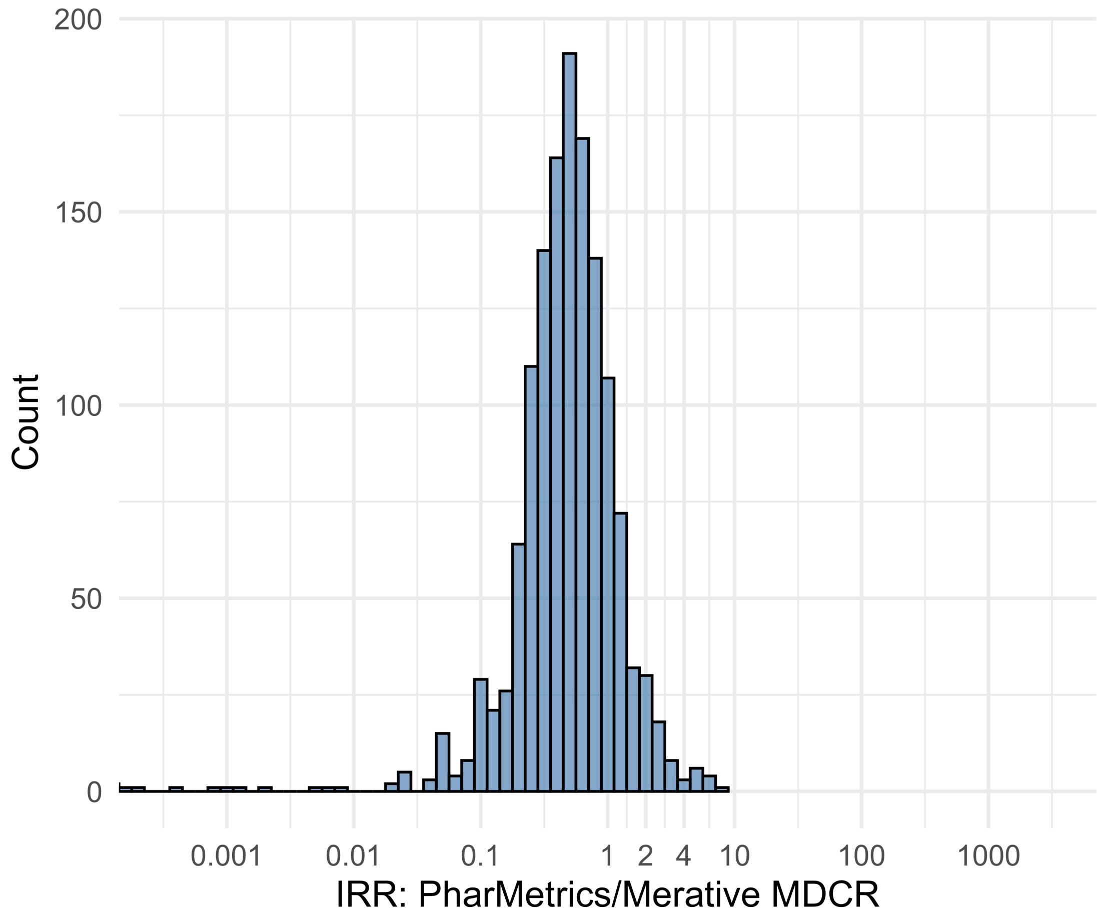


Figure S5.141. IRR distribution comparing PharMetrics and Merative MDCR.

*Table S5.141. Most extreme IRRs for PharMetrics vs Merative MDCR (rounded to 0.001).*

| source1 | source2 | Drug and Indication | Outcome | IRR |
|---|---|---|---|---|
| PharMetrics | Merative MDCR | New users of Trimethoprim systemetic nested in Acute Typical Pneumonia | Earliest event of Thrombotic microangiopathy (TMA) or Microangiopathic hemolytic anemia (MAHA) | 7.3 |
| PharMetrics | Merative MDCR | New users of Tumor Necrosis Factor alpha (TNFa) inhibitors nested in Rheumatoid arthritis | Myocarditis Pericarditis | 6.2 |
| PharMetrics | Merative MDCR | New users of Tumor Necrosis Factor alpha (TNFa) inhibitors nested in Rheumatoid arthritis | Bells Palsy | 6.2 |
| PharMetrics | Merative MDCR | New users of Beta blockers nested in essential hypertension | Cardiovascular-related mortality | <0.001 |
| PharMetrics | Merative MDCR | New users of Cephalosporin systemetic nested in Urinary Tract Infection | Cardiovascular-related mortality | <0.001 |
| PharMetrics | Merative MDCR | New users of Fluoroquinolone systemic nested in Urinary Tract Infection | Cardiovascular-related mortality | <0.001 |

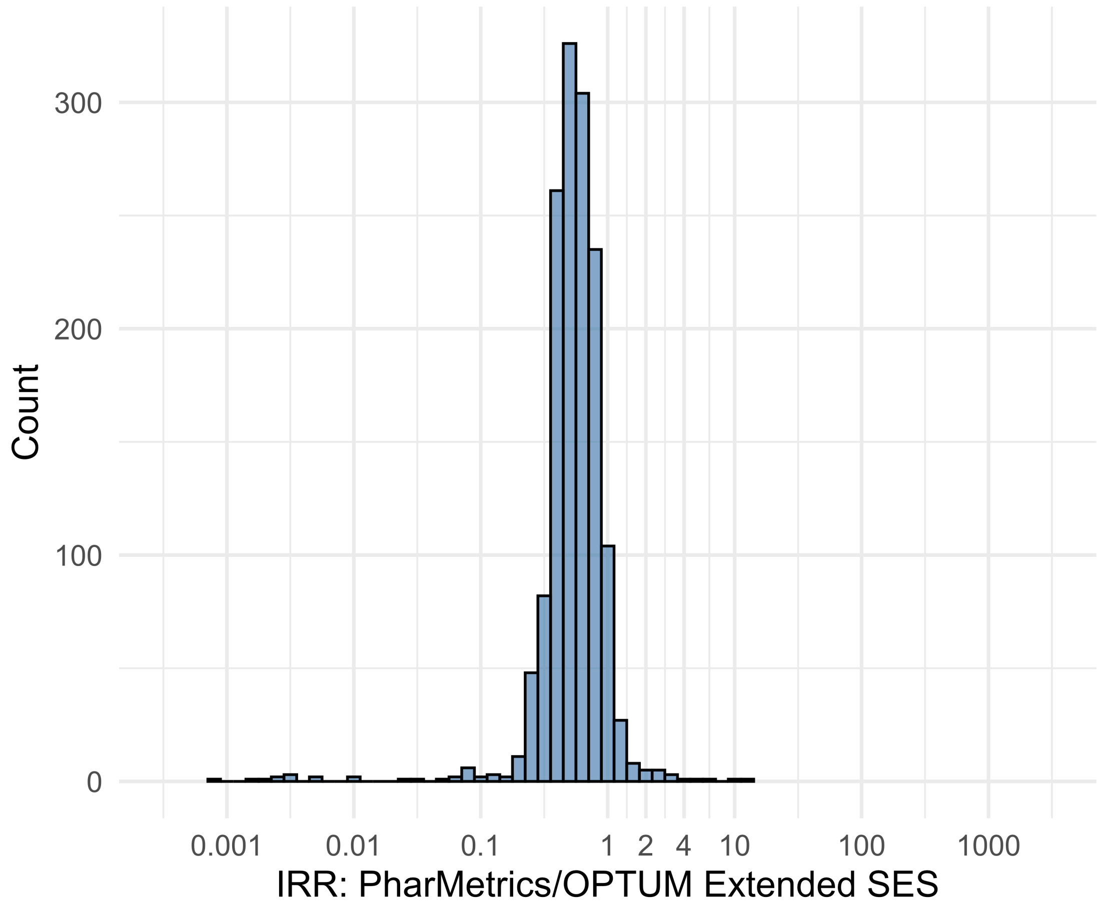


Figure S5.142. IRR distribution comparing PharMetrics and OPTUM Extended SES.

*Table S5.142. Most extreme IRRs for PharMetrics vs OPTUM Extended SES (rounded to 0.001).*

| source1 | source2 | Drug and Indication | Outcome | IRR |
|---|---|---|---|---|
| PharMetrics | OPTUM Extended SES | New users of Tumor Necrosis Factor alpha (TNFa) inhibitors nested in Ulcerative colitis | Autoimmune hemolytic anemia | 11.8 |
| PharMetrics | OPTUM Extended SES | New users of Tumor Necrosis Factor alpha (TNFa) inhibitors nested in Psoriatic Arthritis | Isolated Immune Thrombocytopenia (ITP), with a washout period of 365 days | 9.8 |
| PharMetrics | OPTUM Extended SES | New users of Tumor Necrosis Factor alpha (TNFa) inhibitors nested in Ulcerative colitis | Guillain Barre Syndrome | 6.1 |
| PharMetrics | OPTUM Extended SES | New users of Beta blockers nested in essential hypertension | Cardiovascular-related mortality | 0.0 |
| PharMetrics | OPTUM Extended SES | New users of Cephalosporin systemetic nested in Urinary Tract Infection | Cardiovascular-related mortality | 0.0 |
| PharMetrics | OPTUM Extended SES | New users of Cephalosporin systemetic nested in Acute Typical Pneumonia | Cardiovascular-related mortality | 0.0 |

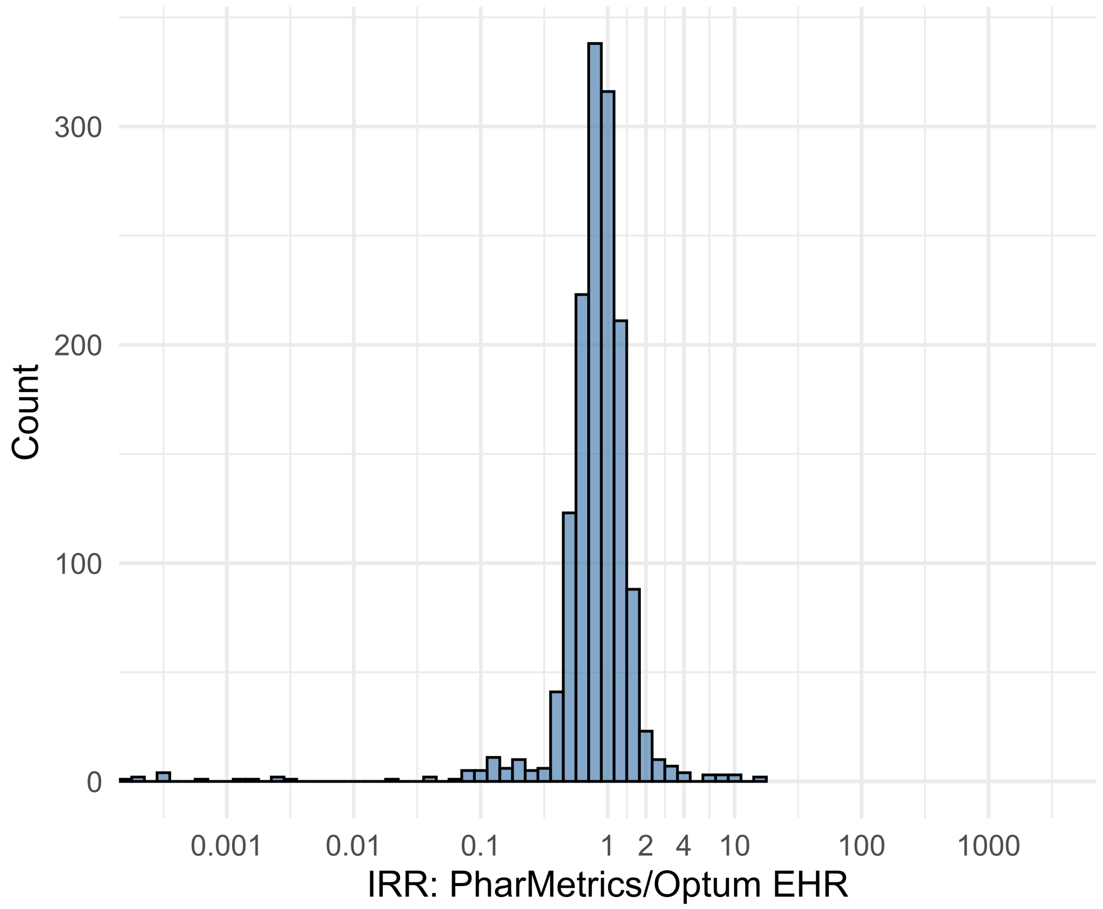


Figure S5.143. IRR distribution comparing PharMetrics and Optum EHR.

*Table S5.143. Most extreme IRRs for PharMetrics vs Optum EHR (rounded to 0.001).*

| source1 | source2 | Drug and Indication | Outcome | IRR |
|---|---|---|---|---|
| PharMetrics | Optum EHR | New users of JAK inhibitors nested in Rheumatoid arthritis | Guillain Barre Syndrome | 16.4 |
| PharMetrics | Optum EHR | New users of Beta blockers nested in Left Heart Failure | Progressive multifocal leukoencephalopathy | 15.6 |
| PharMetrics | Optum EHR | New users of Tumor Necrosis Factor alpha (TNFa) inhibitors nested in Crohns disease | Stevens-Johnson syndrome, toxic epidermal necrolysis spectrum | 11.0 |
| PharMetrics | Optum EHR | New users of Beta blockers nested in essential hypertension | Cardiovascular-related mortality | <0.001 |
| PharMetrics | Optum EHR | New users of Cephalosporin systemetic nested in Acute Typical Pneumonia | Cardiovascular-related mortality | <0.001 |
| PharMetrics | Optum EHR | New users of Cephalosporin systemetic nested in Urinary Tract Infection | Cardiovascular-related mortality | <0.001 |

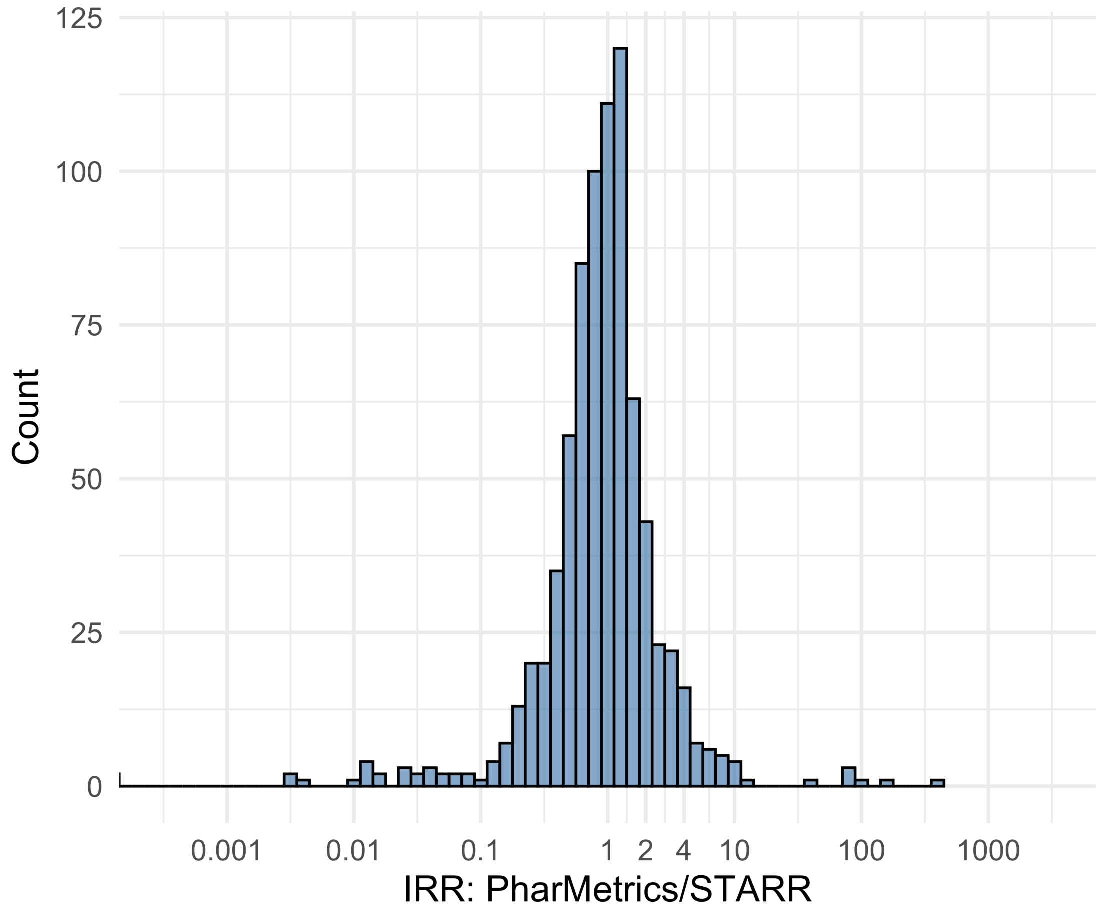


Figure S5.144. IRR distribution comparing PharMetrics and STARR.

*Table S5.144. Most extreme IRRs for PharMetrics vs STARR (rounded to 0.001).*

| source1 | source2 | Drug and Indication | Outcome | IRR |
|---|---|---|---|---|
| PharMetrics | STARR | New users of Beta blockers nested in essential hypertension | Febrile Neutropenia or Neutropenic Fever | 378.7 |
| PharMetrics | STARR | New users of Beta blockers nested in Left Heart Failure | Febrile Neutropenia or Neutropenic Fever | 143.3 |
| PharMetrics | STARR | New users of Fluoroquinolone systemic nested in Acute Typical Pneumonia | Febrile Neutropenia or Neutropenic Fever | 103.4 |
| PharMetrics | STARR | New users of Cephalosporin systemetic nested in Acute Typical Pneumonia | Cardiovascular-related mortality | <0.001 |
| PharMetrics | STARR | New users of Beta blockers nested in essential hypertension | Cardiovascular-related mortality | <0.001 |
| PharMetrics | STARR | New users of Fluoroquinolone systemic nested in Acute Typical Pneumonia | Cardiovascular-related mortality | <0.001 |

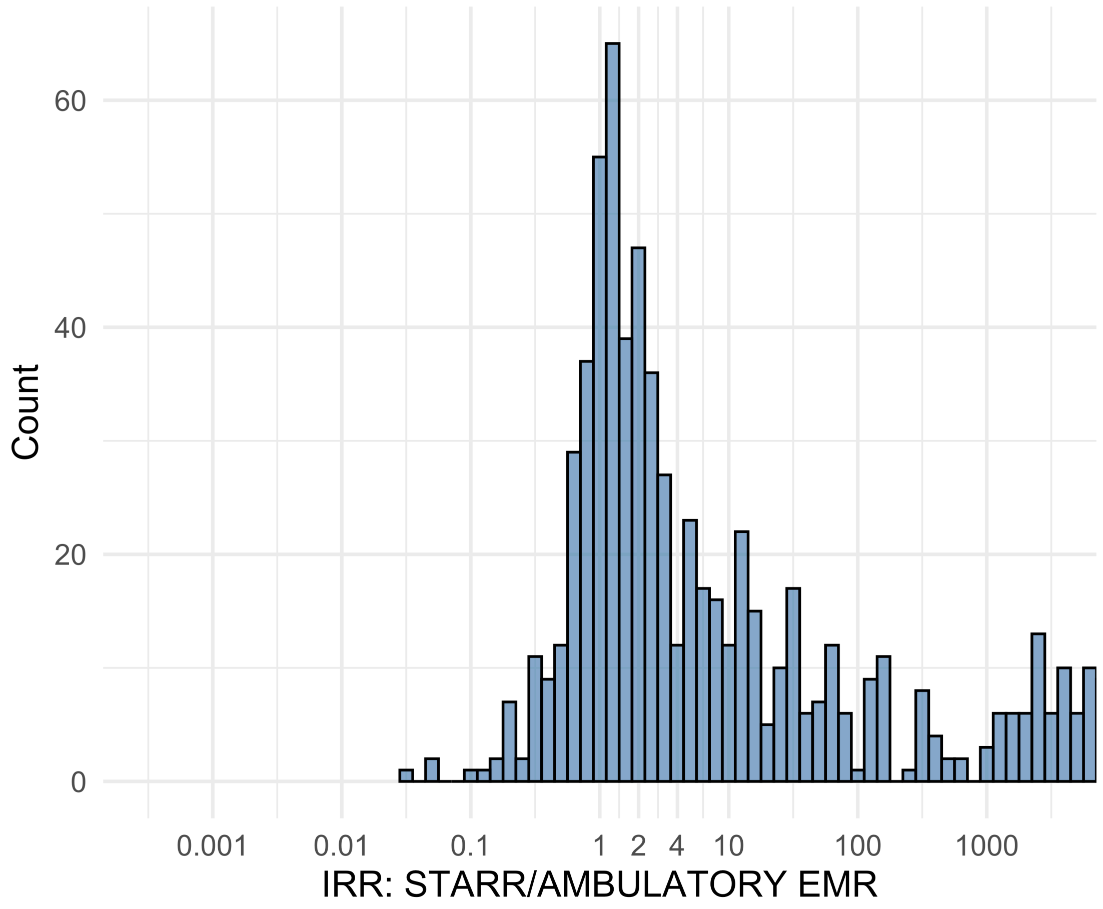


Figure S5.145. IRR distribution comparing STARR and AMBULATORY EMR.

*Table S5.145. Most extreme IRRs for STARR vs AMBULATORY EMR (rounded to 0.001).*

| source1 | source2 | Drug and Indication | Outcome | IRR |
|---|---|---|---|---|
| STARR | AMBULATORY EMR | New users of Beta blockers nested in essential hypertension | Hospitalization with heart failure events | 209719.6 |
| STARR | AMBULATORY EMR | New users of Beta blockers nested in essential hypertension | 4-point MACE | 139856.2 |
| STARR | AMBULATORY EMR | New users of Beta blockers nested in essential hypertension | Total cardiovascular disease events (ischemic stroke, hemorrhagic stroke, heart failure, acute myocardial infarction or sudden cardiac death) | 137115.1 |
| STARR | AMBULATORY EMR | New users of SGLT2 inhibitor nested in Type 2 diabetes mellitus | Anaphylaxis | 0.0 |
| STARR | AMBULATORY EMR | New users of GLP-1 receptor antagonists nested in obesity | Anaphylaxis | 0.1 |
| STARR | AMBULATORY EMR | New users of GLP-1 receptor antagonists nested in Type 2 diabetes mellitus | Anaphylaxis | 0.1 |

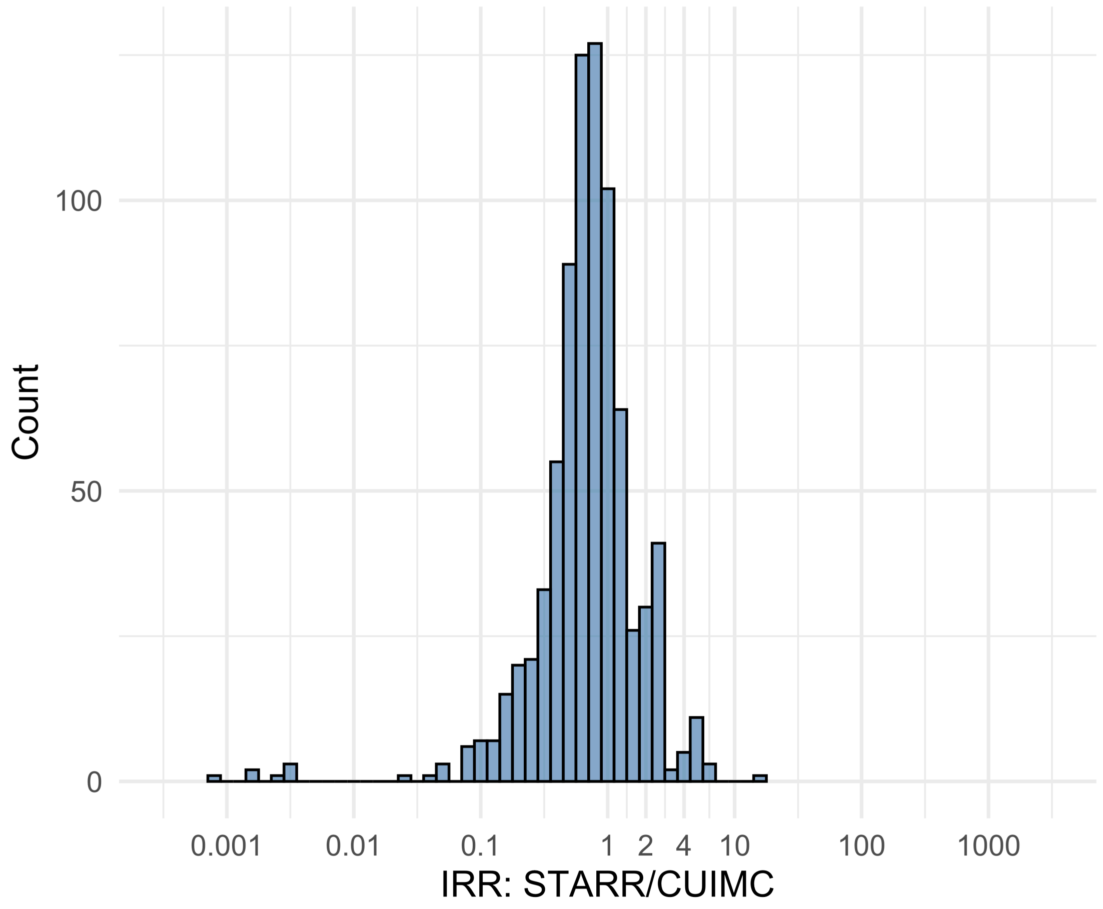


Figure S5.146. IRR distribution comparing STARR and CUIMC.

*Table S5.146. Most extreme IRRs for STARR vs CUIMC (rounded to 0.001).*

| source1 | source2 | Drug and Indication | Outcome | IRR |
|---|---|---|---|---|
| STARR | CUIMC | New users of GLP-1 receptor antagonists nested in obesity | Abnormal weight loss events | 15.5 |
| STARR | CUIMC | New users of Fluoroquinolone systemic nested in Acute Typical Pneumonia | Earliest event of Thrombotic microangiopathy (TMA) or Microangiopathic hemolytic anemia (MAHA) | 6.8 |
| STARR | CUIMC | New users of Beta blockers nested in Acute Myocardial Infarction | Acquired Pure Red Cell Aplasia | 5.9 |
| STARR | CUIMC | New users of Beta blockers nested in essential hypertension | Febrile Neutropenia or Neutropenic Fever | 0.0 |
| STARR | CUIMC | New users of Fluoroquinolone systemic nested in Acute Typical Pneumonia | Febrile Neutropenia or Neutropenic Fever | 0.0 |
| STARR | CUIMC | New users of Fluoroquinolone systemic nested in Urinary Tract Infection | Febrile Neutropenia or Neutropenic Fever | 0.0 |

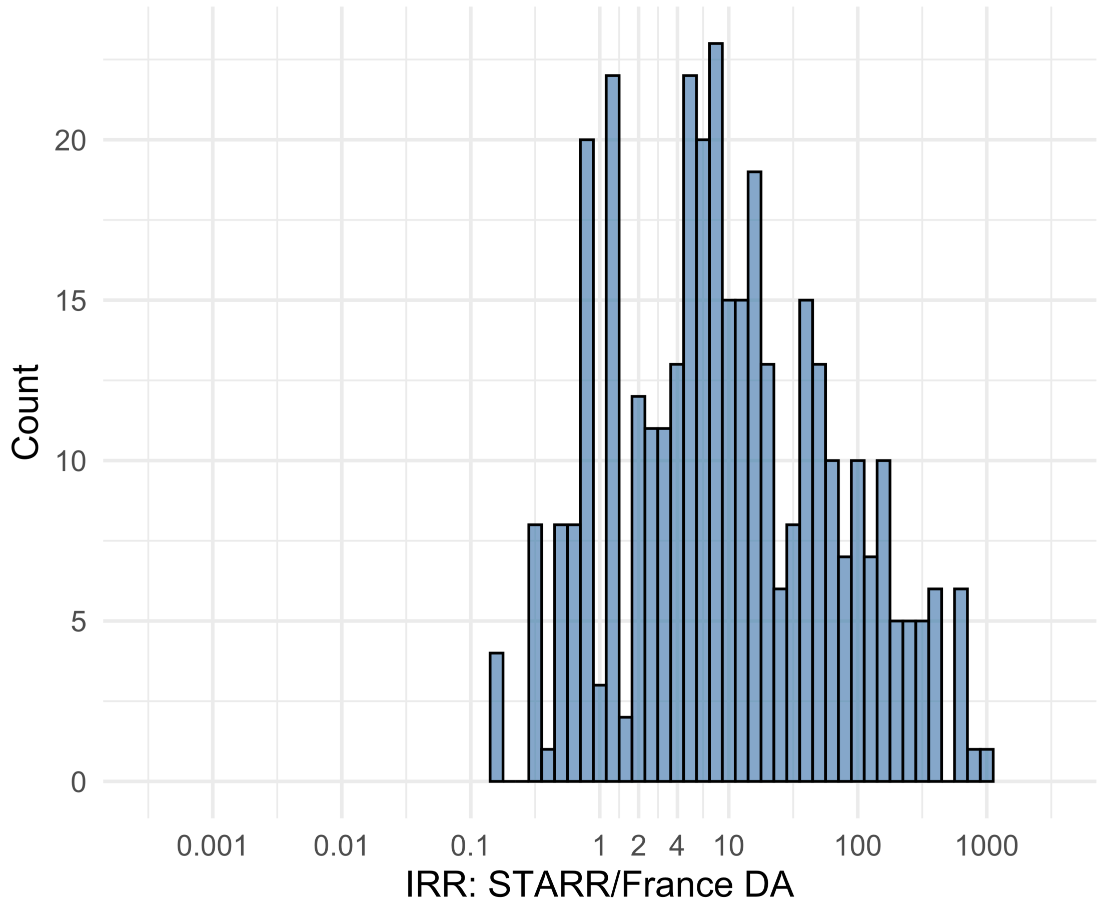


Figure S5.147. IRR distribution comparing STARR and France DA.

*Table S5.147. Most extreme IRRs for STARR vs France DA (rounded to 0.001).*

| source1 | source2 | Drug and Indication | Outcome | IRR |
|---|---|---|---|---|
| STARR | France DA | New users of Beta blockers nested in essential hypertension | Hospitalization with heart failure events | 1030.2 |
| STARR | France DA | New users of Cephalosporin systemetic nested in Urinary Tract Infection | Hospitalization with heart failure events | 743.0 |
| STARR | France DA | New users of Beta blockers nested in essential hypertension | 4-point MACE | 687.0 |
| STARR | France DA | New users of Beta blockers nested in essential hypertension | Febrile Neutropenia or Neutropenic Fever | 0.1 |
| STARR | France DA | New users of Beta blockers nested in essential hypertension | Polymorphic Ventricular Tachycardia or Torsades de Pointes | 0.1 |
| STARR | France DA | New users of Beta blockers nested in essential hypertension | All events Drug Rash with Eosinophilia and Systemic Symptoms (DRESS) | 0.1 |

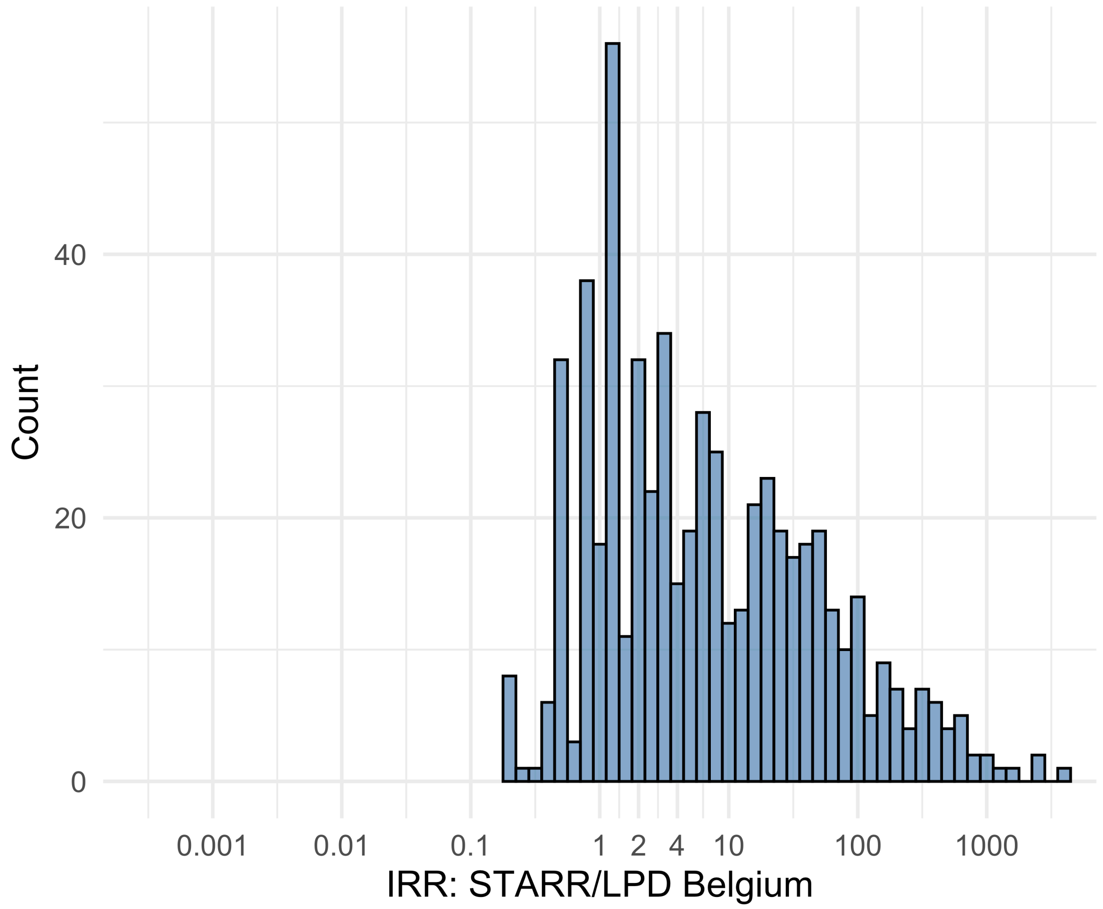


Figure S5.148. IRR distribution comparing STARR and LPD Belgium.

*Table S5.148. Most extreme IRRs for STARR vs LPD Belgium (rounded to 0.001).*

| source1 | source2 | Drug and Indication | Outcome | IRR |
|---|---|---|---|---|
| STARR | LPD Belgium | New users of Beta blockers nested in essential hypertension | Hospitalization with heart failure events | 4009.5 |
| STARR | LPD Belgium | New users of Beta blockers nested in essential hypertension | 4-point MACE | 2673.8 |
| STARR | LPD Belgium | New users of Beta blockers nested in essential hypertension | Total cardiovascular disease events (ischemic stroke, hemorrhagic stroke, heart failure, acute myocardial infarction or sudden cardiac death) | 2621.4 |
| STARR | LPD Belgium | New users of GLP-1 receptor antagonists nested in obesity | Narcolepsy | 0.2 |
| STARR | LPD Belgium | New users of Cephalosporin systemetic nested in Urinary Tract Infection | Febrile Neutropenia or Neutropenic Fever | 0.2 |
| STARR | LPD Belgium | New users of Cephalosporin systemetic nested in Urinary Tract Infection | Polymorphic Ventricular Tachycardia or Torsades de Pointes | 0.2 |

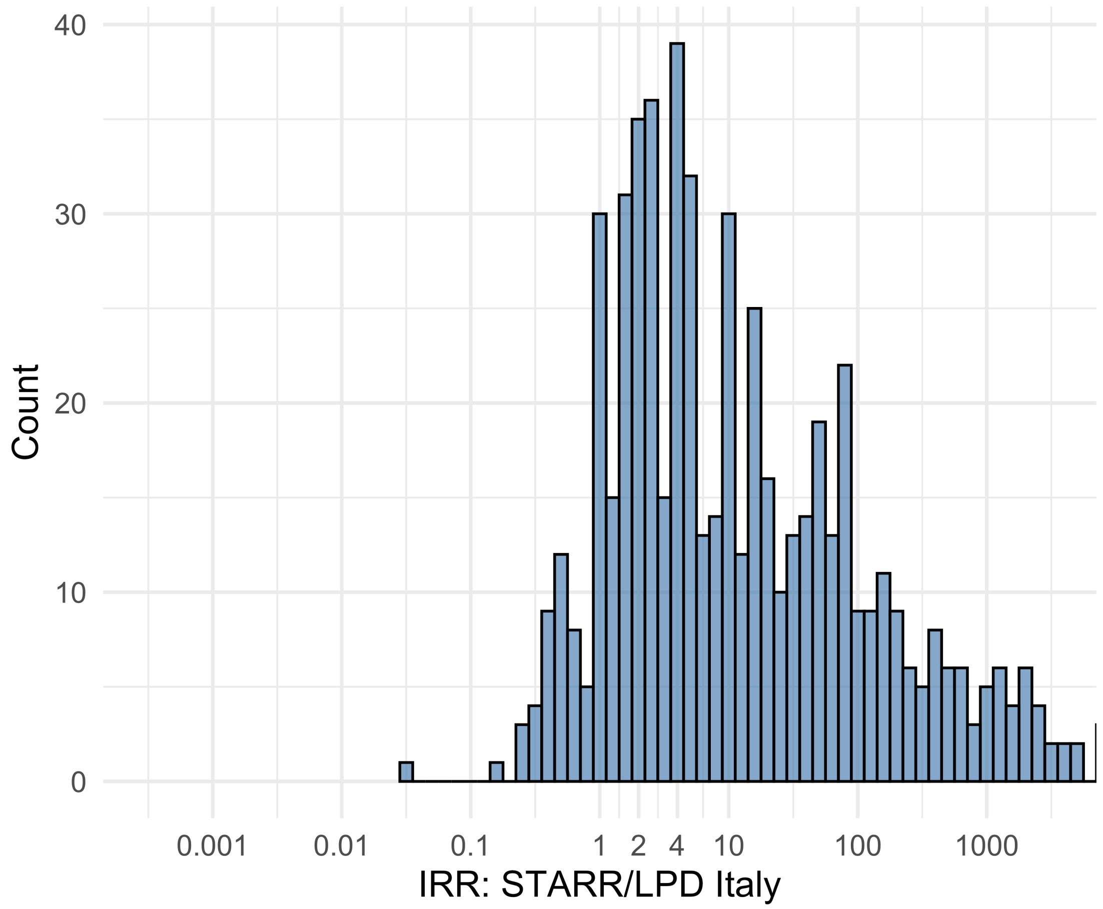


Figure S5.149. IRR distribution comparing STARR and LPD Italy.

*Table S5.149. Most extreme IRRs for STARR vs LPD Italy (rounded to 0.001).*

| source1 | source2 | Drug and Indication | Outcome | IRR |
|---|---|---|---|---|
| STARR | LPD Italy | New users of Beta blockers nested in essential hypertension | Hospitalization with heart failure events | 13035.8 |
| STARR | LPD Italy | New users of Beta blockers nested in essential hypertension | 4-point MACE | 8693.2 |
| STARR | LPD Italy | New users of Beta blockers nested in essential hypertension | Total cardiovascular disease events (ischemic stroke, hemorrhagic stroke, heart failure, acute myocardial infarction or sudden cardiac death) | 8522.9 |
| STARR | LPD Italy | New users of Beta blockers nested in essential hypertension | Progressive multifocal leukoencephalopathy | 0.0 |
| STARR | LPD Italy | New users of Cephalosporin systemetic nested in Urinary Tract Infection | Progressive multifocal leukoencephalopathy | 0.2 |
| STARR | LPD Italy | New users of Cephalosporin systemetic nested in Acute Typical Pneumonia | Renal Cancer | 0.2 |

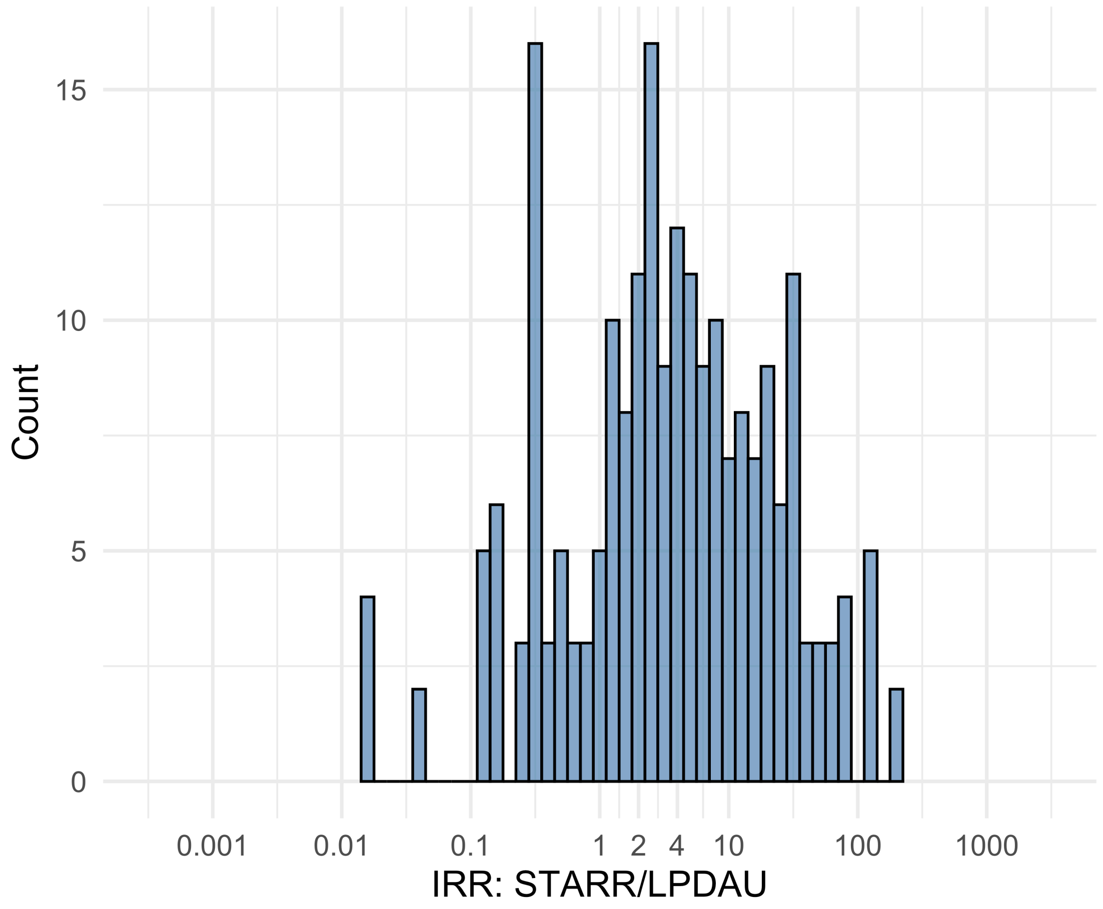


Figure S5.150. IRR distribution comparing STARR and LPDAU.

*Table S5.150. Most extreme IRRs for STARR vs LPDAU (rounded to 0.001).*

| source1 | source2 | Drug and Indication | Outcome | IRR |
|---|---|---|---|---|
| STARR | LPDAU | New users of Cephalosporin systemetic nested in Urinary Tract Infection | Hospitalization with heart failure events | 214.2 |
| STARR | LPDAU | New users of Cephalosporin systemetic nested in Urinary Tract Infection | Acute Kidney Injury AKI | 187.9 |
| STARR | LPDAU | New users of Trimethoprim systemetic nested in Urinary Tract Infection | Hospitalization with heart failure events | 126.2 |
| STARR | LPDAU | New users of Beta blockers nested in essential hypertension | Febrile Neutropenia or Neutropenic Fever | 0.0 |
| STARR | LPDAU | New users of Beta blockers nested in essential hypertension | Polymorphic Ventricular Tachycardia or Torsades de Pointes | 0.0 |
| STARR | LPDAU | New users of Beta blockers nested in essential hypertension | All events Drug Rash with Eosinophilia and Systemic Symptoms (DRESS) | 0.0 |

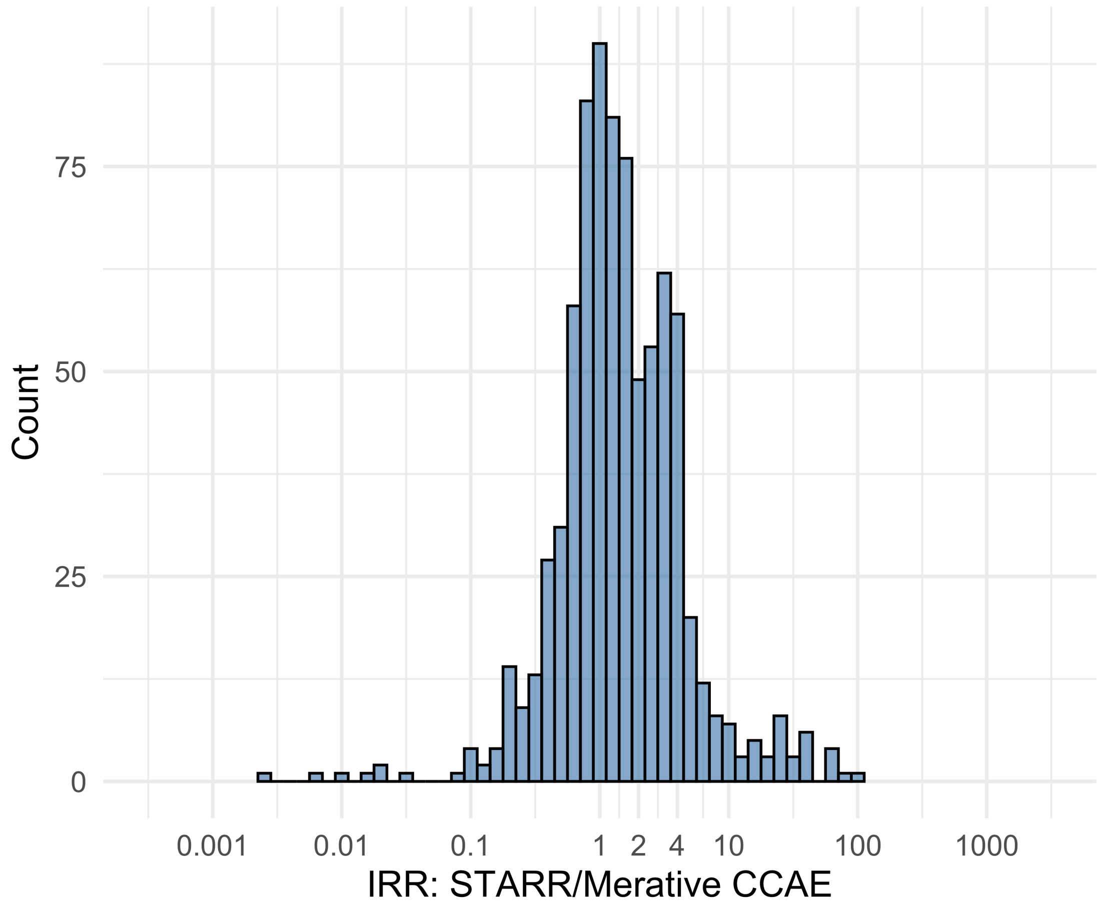


Figure S5.151. IRR distribution comparing STARR and Merative CCAE.

*Table S5.151. Most extreme IRRs for STARR vs Merative CCAE (rounded to 0.001).*

| source1 | source2 | Drug and Indication | Outcome | IRR |
|---|---|---|---|---|
| STARR | Merative CCAE | New users of Trimethoprim systemetic nested in Urinary Tract Infection | Polymorphic Ventricular Tachycardia or Torsades de Pointes | 95.8 |
| STARR | Merative CCAE | New users of Beta blockers nested in Acute Myocardial Infarction | Thrombosis with Thrombocytopenia (TWT) | 78.5 |
| STARR | Merative CCAE | New users of Fluoroquinolone systemic nested in Urinary Tract Infection | Polymorphic Ventricular Tachycardia or Torsades de Pointes | 63.3 |
| STARR | Merative CCAE | New users of Beta blockers nested in essential hypertension | Febrile Neutropenia or Neutropenic Fever | 0.0 |
| STARR | Merative CCAE | New users of Beta blockers nested in Left Heart Failure | Febrile Neutropenia or Neutropenic Fever | 0.0 |
| STARR | Merative CCAE | New users of Fluoroquinolone systemic nested in Acute Typical Pneumonia | Febrile Neutropenia or Neutropenic Fever | 0.0 |

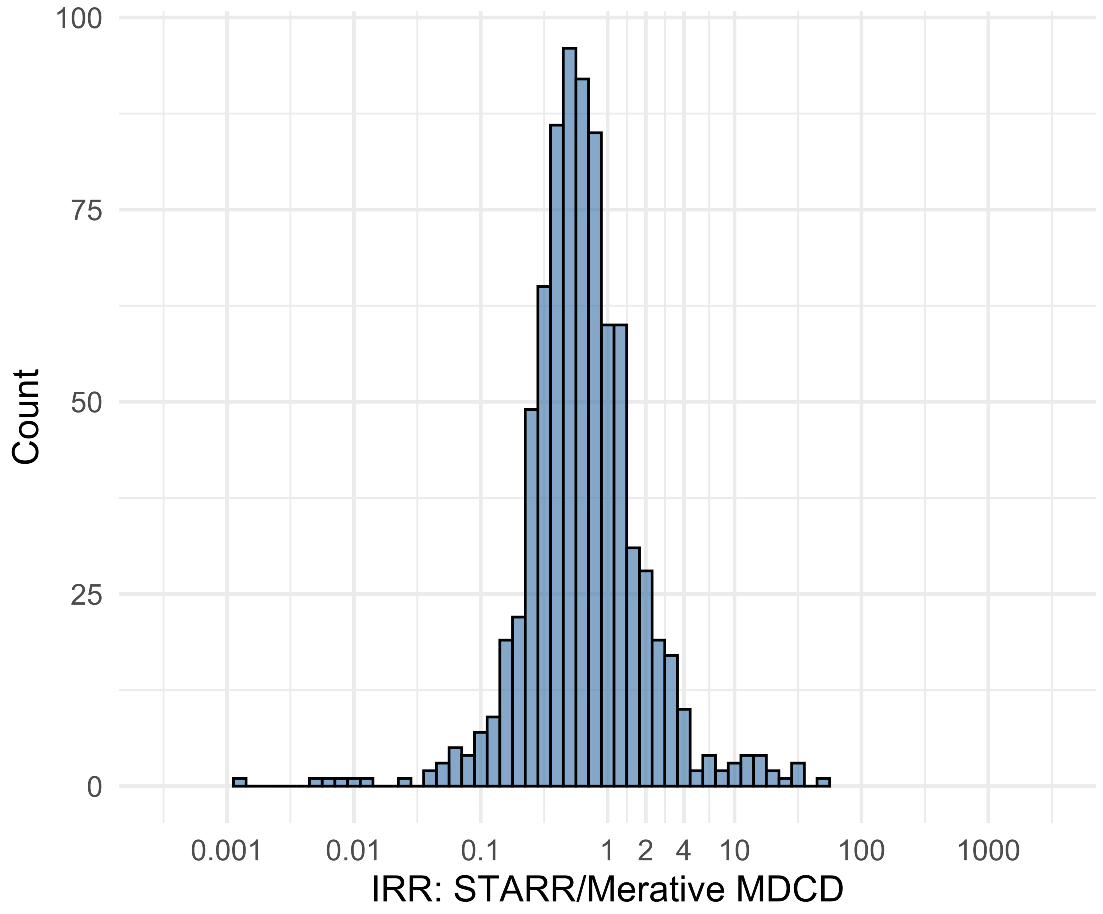


Figure S5.152. IRR distribution comparing STARR and Merative MDCD.

*Table S5.152. Most extreme IRRs for STARR vs Merative MDCD (rounded to 0.001).*

| source1 | source2 | Drug and Indication | Outcome | IRR |
|---|---|---|---|---|
| STARR | Merative MDCD | New users of Trimethoprim systemetic nested in Urinary Tract Infection | Progressive multifocal leukoencephalopathy | 45.7 |
| STARR | Merative MDCD | New users of Cephalosporin systemetic nested in Acute Typical Pneumonia | Progressive multifocal leukoencephalopathy | 29.4 |
| STARR | Merative MDCD | New users of SGLT2 inhibitor nested in Type 2 diabetes mellitus | Progressive multifocal leukoencephalopathy | 29.1 |
| STARR | Merative MDCD | New users of Beta blockers nested in essential hypertension | Febrile Neutropenia or Neutropenic Fever | 0.0 |
| STARR | Merative MDCD | New users of Beta blockers nested in Left Heart Failure | Febrile Neutropenia or Neutropenic Fever | 0.0 |
| STARR | Merative MDCD | New users of Fluoroquinolone systemic nested in Acute Typical Pneumonia | Febrile Neutropenia or Neutropenic Fever | 0.0 |

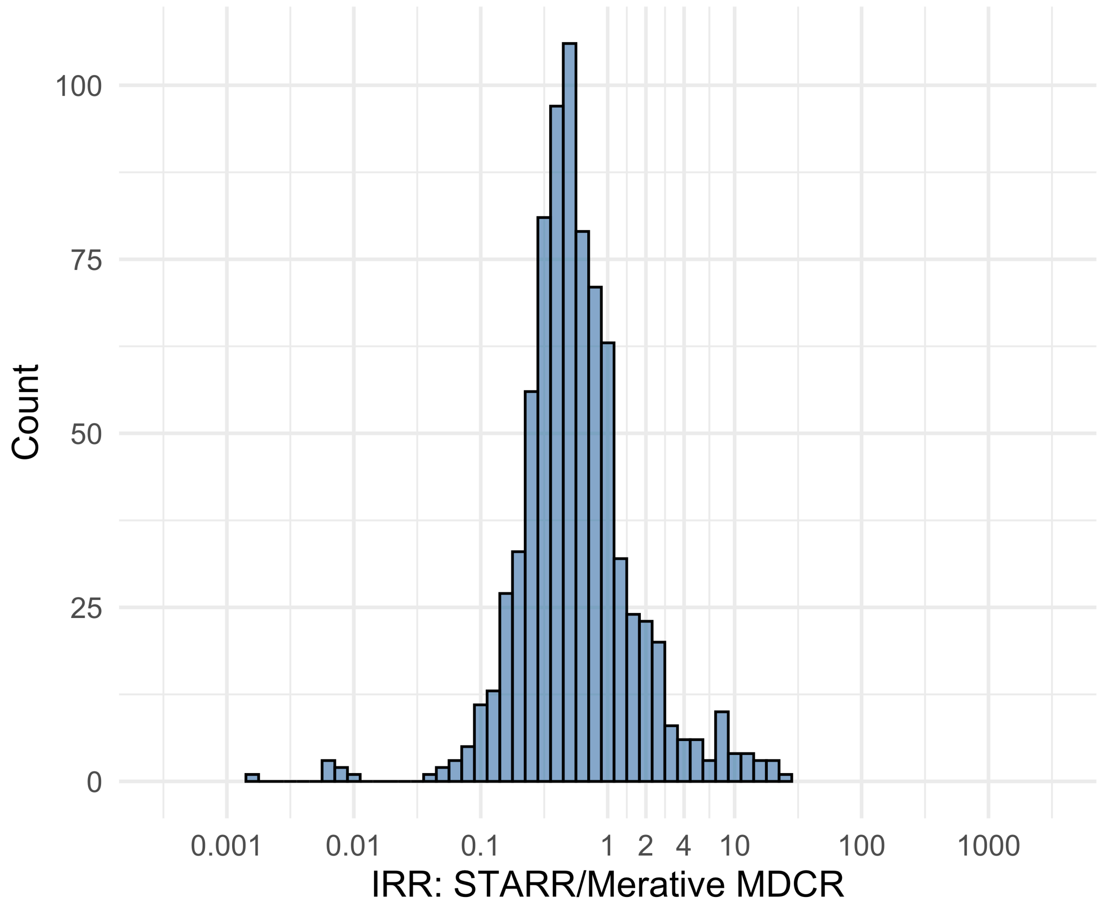


Figure S5.153. IRR distribution comparing STARR and Merative MDCR.

*Table S5.153. Most extreme IRRs for STARR vs Merative MDCR (rounded to 0.001).*

| source1 | source2 | Drug and Indication | Outcome | IRR |
|---|---|---|---|---|
| STARR | Merative MDCR | New users of Beta blockers nested in Acute Myocardial Infarction | Thrombosis with Thrombocytopenia (TWT) | 26.7 |
| STARR | Merative MDCR | New users of Cephalosporin systemetic nested in Acute Typical Pneumonia | Progressive multifocal leukoencephalopathy | 20.7 |
| STARR | Merative MDCR | New users of Beta blockers nested in Acute Myocardial Infarction | Polymorphic Ventricular Tachycardia or Torsades de Pointes | 19.7 |
| STARR | Merative MDCR | New users of Beta blockers nested in essential hypertension | Febrile Neutropenia or Neutropenic Fever | 0.0 |
| STARR | Merative MDCR | New users of Fluoroquinolone systemic nested in Urinary Tract Infection | Febrile Neutropenia or Neutropenic Fever | 0.0 |
| STARR | Merative MDCR | New users of Cephalosporin systemetic nested in Urinary Tract Infection | Febrile Neutropenia or Neutropenic Fever | 0.0 |

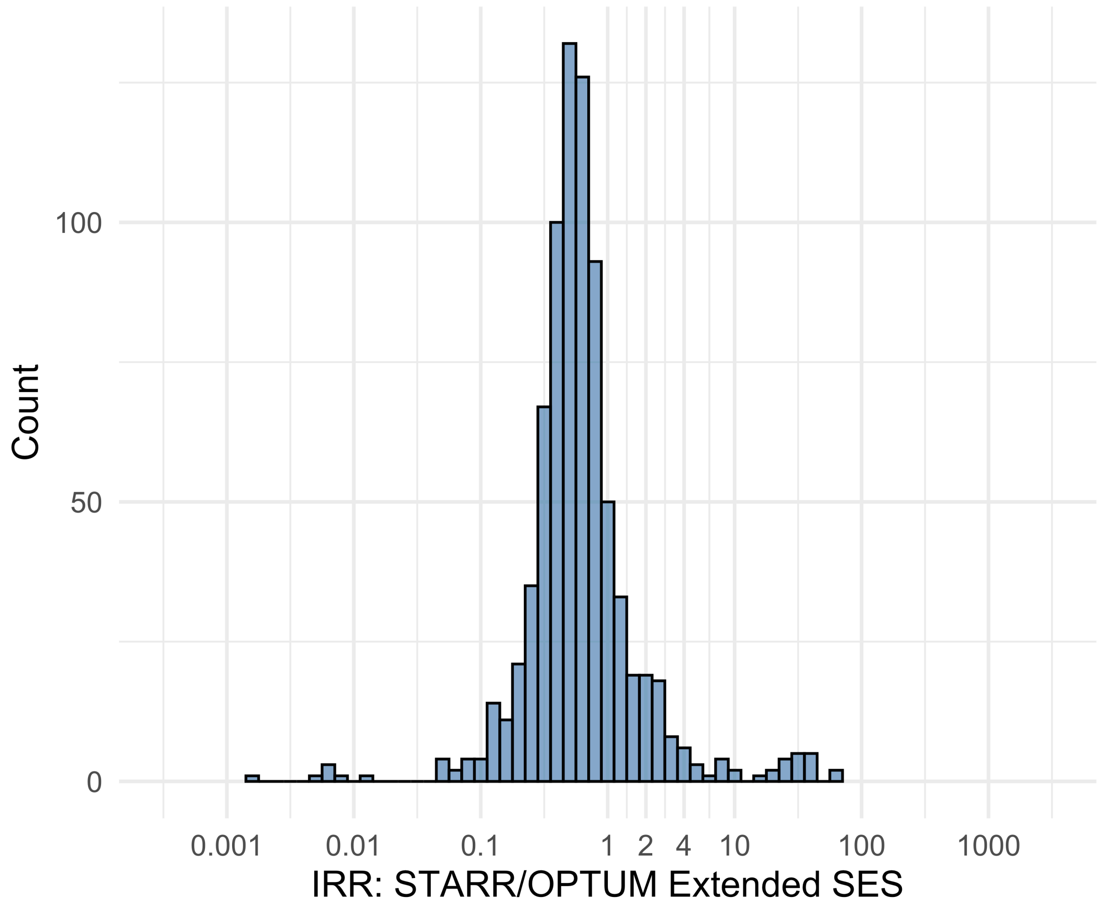


Figure S5.154. IRR distribution comparing STARR and OPTUM Extended SES.

*Table S5.154. Most extreme IRRs for STARR vs OPTUM Extended SES (rounded to 0.001).*

| source1 | source2 | Drug and Indication | Outcome | IRR |
|---|---|---|---|---|
| STARR | OPTUM Extended SES | New users of SGLT2 inhibitor nested in Type 2 diabetes mellitus | Progressive multifocal leukoencephalopathy | 57.9 |
| STARR | OPTUM Extended SES | New users of Fluoroquinolone systemic nested in Acute Typical Pneumonia | Cardiovascular-related mortality | 56.7 |
| STARR | OPTUM Extended SES | New users of Cephalosporin systemetic nested in Acute Typical Pneumonia | Cardiovascular-related mortality | 40.9 |
| STARR | OPTUM Extended SES | New users of Beta blockers nested in essential hypertension | Febrile Neutropenia or Neutropenic Fever | 0.0 |
| STARR | OPTUM Extended SES | New users of Beta blockers nested in Left Heart Failure | Febrile Neutropenia or Neutropenic Fever | 0.0 |
| STARR | OPTUM Extended SES | New users of Cephalosporin systemetic nested in Urinary Tract Infection | Febrile Neutropenia or Neutropenic Fever | 0.0 |

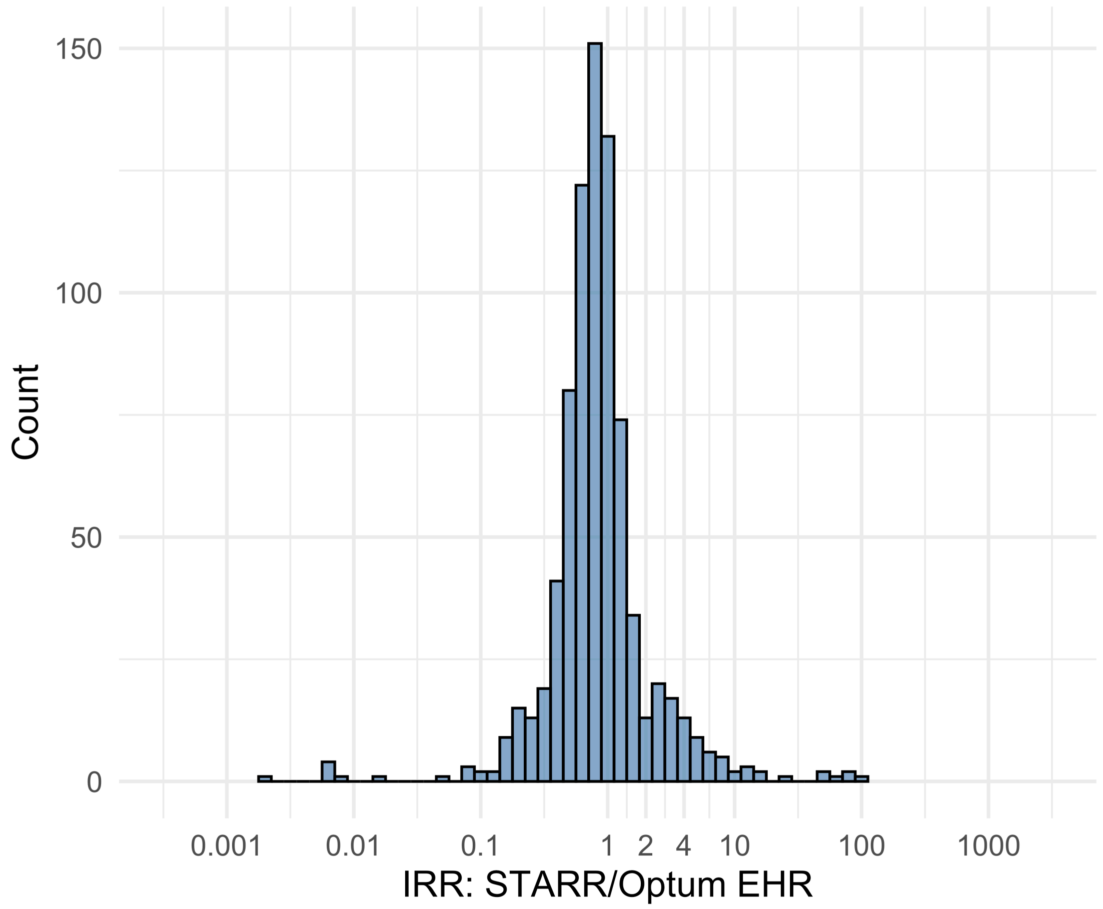


Figure S5.155. IRR distribution comparing STARR and Optum EHR.

*Table S5.155. Most extreme IRRs for STARR vs Optum EHR (rounded to 0.001).*

| source1 | source2 | Drug and Indication | Outcome | IRR |
|---|---|---|---|---|
| STARR | Optum EHR | New users of SGLT2 inhibitor nested in Type 2 diabetes mellitus | Progressive multifocal leukoencephalopathy | 94.9 |
| STARR | Optum EHR | New users of GLP-1 receptor antagonists nested in obesity | Progressive multifocal leukoencephalopathy | 85.3 |
| STARR | Optum EHR | New users of GLP-1 receptor antagonists nested in Type 2 diabetes mellitus | Progressive multifocal leukoencephalopathy | 72.1 |
| STARR | Optum EHR | New users of Beta blockers nested in essential hypertension | Febrile Neutropenia or Neutropenic Fever | 0.0 |
| STARR | Optum EHR | New users of Beta blockers nested in Left Heart Failure | Febrile Neutropenia or Neutropenic Fever | 0.0 |
| STARR | Optum EHR | New users of Fluoroquinolone systemic nested in Urinary Tract Infection | Febrile Neutropenia or Neutropenic Fever | 0.0 |

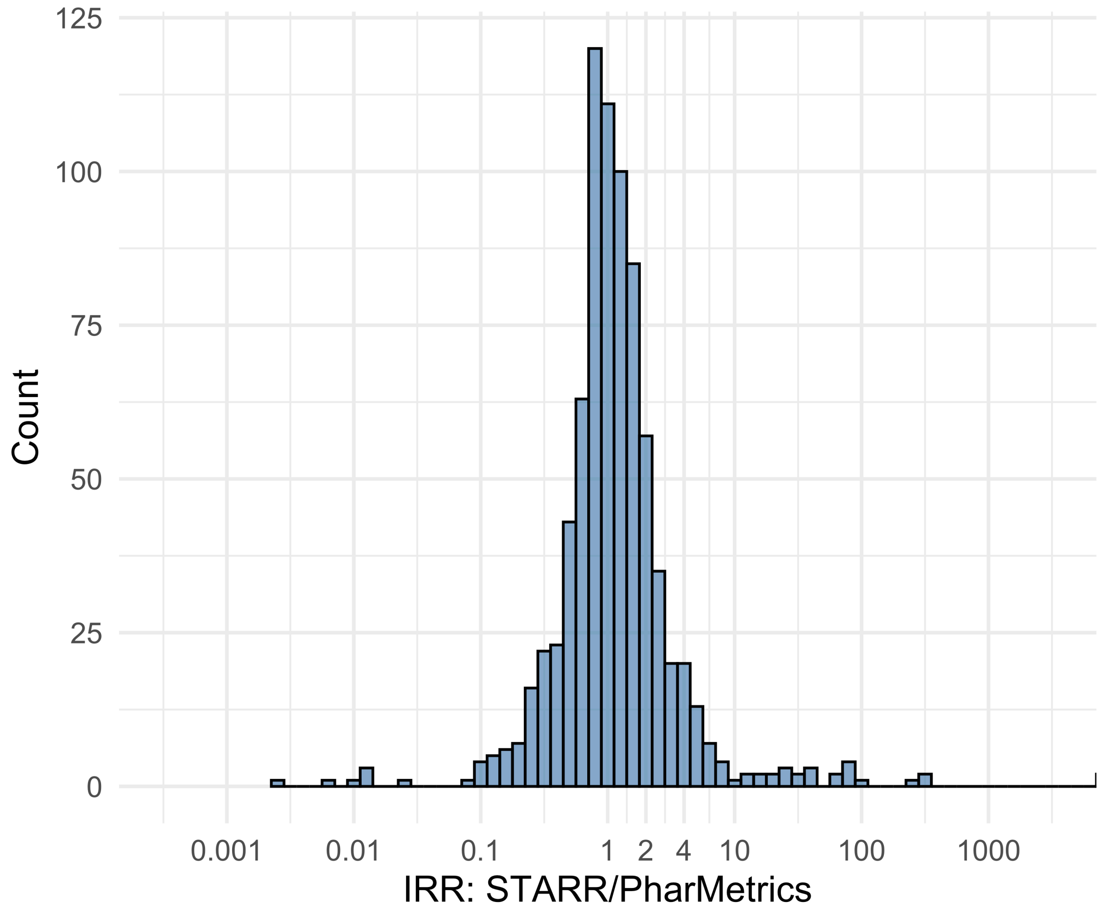


Figure S5.156. IRR distribution comparing STARR and PharMetrics.

*Table S5.156. Most extreme IRRs for STARR vs PharMetrics (rounded to 0.001).*

| source1 | source2 | Drug and Indication | Outcome | IRR |
|---|---|---|---|---|
| STARR | PharMetrics | New users of Cephalosporin systemetic nested in Acute Typical Pneumonia | Cardiovascular-related mortality | 18914.3 |
| STARR | PharMetrics | New users of Beta blockers nested in essential hypertension | Cardiovascular-related mortality | 18660.0 |
| STARR | PharMetrics | New users of Fluoroquinolone systemic nested in Acute Typical Pneumonia | Cardiovascular-related mortality | 16616.1 |
| STARR | PharMetrics | New users of Beta blockers nested in essential hypertension | Febrile Neutropenia or Neutropenic Fever | 0.0 |
| STARR | PharMetrics | New users of Beta blockers nested in Left Heart Failure | Febrile Neutropenia or Neutropenic Fever | 0.0 |
| STARR | PharMetrics | New users of Fluoroquinolone systemic nested in Acute Typical Pneumonia | Febrile Neutropenia or Neutropenic Fever | 0.0 |
